# Association between Bisphosphonate use and COVID-19 related outcomes: a retrospective cohort study

**DOI:** 10.1101/2022.06.14.22276397

**Authors:** Jeffrey Thompson, Yidi Wang, Tobias Dreischulte, Olga Barreiro, Rodrigo J. Gonzalez, Pavel Hanč, Colette Matysiak, Harold R. Neely, Marietta Rottenkolber, Tom Haskell, Stefan Endres, Ulrich H. von Andrian

**Author notes:** Send correspondence to: Ulrich H. von Andrian, M.D. Dept. of Immunology Harvard Medical School Boston, MA 02115, USA. Equal contribution (Jeffrey Thompson, Yidi Wang).

## Abstract

**Background:** Although there are several efficacious vaccines against COVID-19, vaccination rates in many regions around the world remain insufficient to prevent continued high disease burden and emergence of viral variants. Repurposing of existing therapeutics that prevent or mitigate severe COVID-19 could help to address these challenges. The objective of this study was to determine whether prior use of bisphosphonates is associated with reduced incidence and/or severity of COVID-19.

**Methods:** A retrospective cohort study utilizing payer-complete health insurance claims data from 8,239,790 patients with continuous medical and prescription insurance from 1-1-2019 to 6-30-2020 was performed. The primary exposure of interest was use of any bisphosphonate from 1-1-2019 to 2-29-2020. Outcomes of interest included: (a) testing for SARS-CoV-2 infection; (b) COVID-19 diagnosis; and (c) hospitalization with COVID-19 diagnosis between 3-1-2020 and 6-30-2020.

**Results:** 7,906,603 patients for whom continuous medical and prescription insurance information was available were selected. 450,366 bisphosphonate users were identified and 1:1 propensity score-matched to bisphosphonate non-users by age, gender, insurance type, primary-care-provider visit in 2019, and comorbidity burden. Bisphosphonate users had lower odds ratios (OR) of testing for SARS-CoV-2 infection (OR=0.22; 95%CI:0.21-0.23; p<0.001), COVID-19 diagnosis (OR=0.23; 95%CI:0.22-0.24; p<0.001), and COVID-19-related hospitalization (OR=0.26; 95%CI:0.24-0.29; p<0.001). Sensitivity analyses yielded results consistent with the primary analysis. Bisphosphonate-use was also associated with decreased odds of acute bronchitis (OR=0.23; 95%CI:0.22-0.23; p<0.001) or pneumonia (OR=0.32; 95%CI:0.31-0.34; p<0.001) in 2019, suggesting that bisphosphonates may protect against respiratory infections by a variety of pathogens, including but not limited to SARS-CoV-2.

**Conclusions:** Prior bisphosphonate-use was associated with dramatically reduced odds of SARS-CoV-2 testing, COVID-19 diagnosis, and COVID-19-related hospitalizations. Prospective clinical trials will be required to establish a causal role for bisphosphonate-use in COVID-19-related outcomes.

## INTRODUCTION

Throughout the COVID-19 pandemic, massive global efforts to repurpose existing drugs as potential therapeutic options for COVID-19 have been undertaken. Drug repurposing, whereby a drug already proven to be safe and effective in humans for another approved clinical indication is evaluated for novel clinical use, may allow for faster identification and deployment of therapeutic agents compared to traditional drug discovery pipelines. Using *in silico* and *in vitro* analyses, a growing list of drugs have been suggested to be potentially efficacious in treating COVID-19 by either direct or indirect antiviral actions^1^. Another potentially beneficial class of drugs may be agents that boost or modulate anti-viral immune responses to SARS-CoV-2 infection to reduce clinical symptoms and/or mitigate disease progression. Regardless of the mechanism of action, ultimately, randomized prospective clinical studies are needed to test the safety and efficacy of each candidate in treating or preventing COVID-19. Observational studies can help prioritize candidates for prospective clinical testing, by examining associations between the use of a candidate drug and the incidence or severity of disease in users compared to a matched group of non-users. Drugs with strong observational evidence for potential effectiveness against COVID-19 may then be considered for prospective trials^1^.

Here, we have investigated bisphosphonates (BPs), a class of small-molecule drugs that inhibit bone resorption by osteoclasts^2^. BPs are widely prescribed as either oral or intravenous formulations to treat osteoporosis, Paget disease, and malignancy-induced hypercalcemia. Additionally, BPs are used as adjuvant therapy for breast cancer^3^. BPs are subdivided into two classes, nitrogen-containing (amino-BPs) and nitrogen-free BPs (non-amino-BPs)^4^. Both accumulate in bone but have distinct molecular mechanisms by which they kill osteoclasts to prevent bone resorption^2^.

Aside from depleting osteoclasts, clinical and experimental studies indicate that BPs exert a plethora of immunomodulatory effects, providing a rationale for exploring BPs as potential repurposed drug candidates for COVID-19 (ref. ^5^). Indeed, amino-BPs regulate the activation, expansion, and/or function of a major subset of human γδT cells^6–8^ as well as neutrophils^9^, monocytes^10^, and macrophages^11, 12^; they can modulate the antigen-presentation capacity of dendritic cells^13^; and in animal studies, both amino-BPs and non-amino-BPs exerted potent adjuvant-like activity to boost antibody and T cells responses to viral antigens^14^. Furthermore, observational studies have reported decreased in-hospital mortality for patients in the ICU^15^, and reduced incidence of pneumoniae and pneumonia-related mortality in patients treated with amino-BPs versus controls^16^. These immunological and clinical effects of BPs combine with several other characteristics that make BPs well-suited as repurposed drug candidates in the context of a pandemic: they are globally accessible as generics, affordable, straightforward to administer, and have known safety profiles in adult^17^ and paediatric populations^18, 19^.

In light of these considerations, we have analysed a database of health insurance claims in the U.S. to determine if prior BP-use is associated with a differential incidence and/or severity of COVID-19-related outcomes. Specifically, we assessed the relationship between use of BPs and COVID-19-related hospitalizations and COVID-19 diagnosis, as well as testing for SARS-CoV-2 infection (as a proxy for severe COVID-19 symptoms given the restricted access to testing during the initial surge). Outcomes were measured from March 1, 2020 to June 30, 2020, a period that roughly coincided with the first wave of COVID-19 in the U.S. and predated the advent of potential outcome modifiers, such as vaccines or other effective treatment options.

## METHODS

### Study Design

A retrospective cohort study was performed using health insurance claims data from January 1, 2019 to June 30, 2020 (study period) in order to assess the relationship between use of BPs and three COVID-19-related outcomes: (a) testing for SARS-CoV-2 infection; (b) COVID-19 diagnosis; and (c) hospitalization with a COVID-19 diagnosis, whereby COVID-19-related hospitalization was deemed the primary endpoint and COVID-19 diagnosis and testing were secondary endpoints. Primary and secondary endpoints were assessed during the observation period of March 1, 2020 to June 30, 2020, roughly corresponding to the first nation-wide surge of COVID-19 in the U.S. (Fig. 1A). In the primary analysis, the risk of COVID-19-related outcomes was assessed among BP users compared to a matched sample of BP non-users with similar demographic and clinical characteristics.

**Figure 1:**
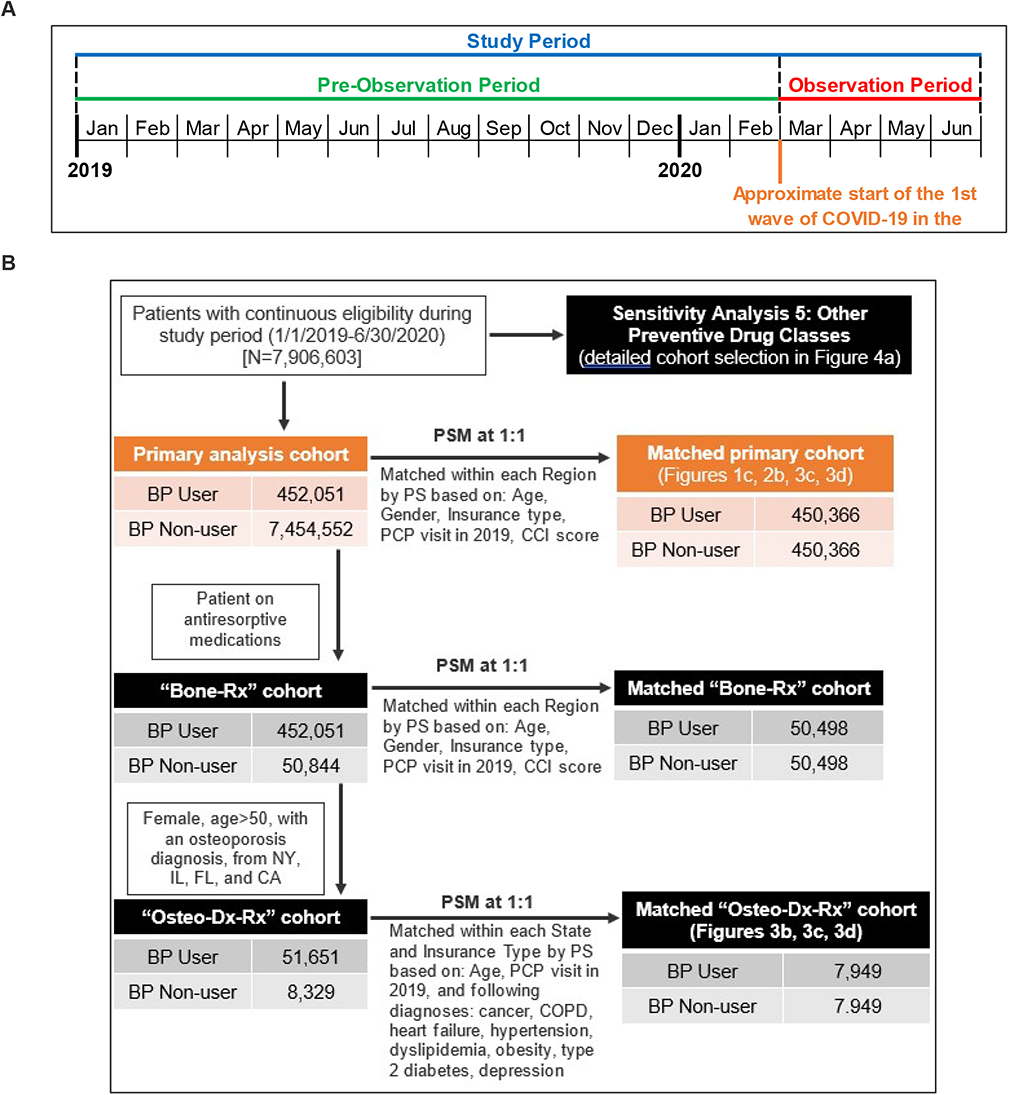
Study Periods, Cohort Selection, and Analyses of BP use on COVID-19-Related Outcomes. **A.** Schematic overview of the study timeline. **B.** Schematic flow diagram illustrating the identification of the study population and matched control populations for primary analysis and sensitivity analyses cohorts. *BP: bisphosphonate; CA: California; CCI: Charlson comorbidity index; CI: confidence interval; COPD: chronic obstructive pulmonary disease; FL: Florida; IL: Illinois; NY: New York; OR: odds ratio; PCP: primary care physician; PS: propensity score; PSM: propensity score match*

### Data Source

Data used for this study included closed medical (inpatient and outpatient) and outpatient-pharmacy-dispensed claims between January 1, 2019 and June 30, 2020, from the Komodo Health payer-complete dataset (https://www.komodohealth.com). This dataset is derived from over 150 private insurers in the U.S. and includes patients with commercial, individual, state exchange-purchased, Medicare Advantage, and Medicaid managed-care insurance coverage. The dataset also provides information on insurance eligibility periods. Closed claims within this dataset represent those that had undergone insurance adjudication. In total, the Komodo Health payer-complete dataset includes health insurance claims data from over 140 million individuals in the U.S. from 2015 to 2020.

### Cohort Definition

All patients were required to have continuous medical and prescription insurance eligibility during the entire study period. Patients with missing information for age, gender, insurance type, or state/region were excluded.

### Exposures of Interest

The primary exposure of interest was the use of any amino- or non-amino BP medication. Exposure to BPs and all other medications of interest were assessed over a 14-month pre-observation period preceding the COVID-19 pandemic in the U.S. This long duration was chosen because of the extended bioavailability of BPs, which accumulate in bone where they are retained and slowly released for up to several years^20^. Patients were classified as BP users if they had any claim at any time during the pre-observation period for one of the following: alendronate, alendronic acid, etidronate, ibandronate, ibandronic acid, pamidronate, risedronate, and zoledronic acid (full details in Appendix 1).

### Timing of BP Dose

The effect of timing and formulation of BPs on COVID-19-related outcomes was more closely examined by varying the window between BP exposure and outcome measurement. The primary analysis BP user cohort, along with their propensity-score matched (see below for cohort matching) BP non-user cohort, were stratified as follows: two cohorts were used as the reference comparator with known BP-exposure during all or most of the pre-observation and the entire observation period, specifically (i) BP users who took oral alendronic acid (dosed daily or weekly) throughout the pre-observation period (i.e. at least one claim or drug-on-hand in each quarter in 2019 and in Jan/Feb. 2020) that also had a days-supply extending past June 30, 2020, and (ii) users of infusion zoledronic acid (dosed annually) with a claim in Q3 or Q4 2019; two cohorts with BP-exposure only during the pre-observation period, namely (iii) users of alendronic acid occurring during the first six months of 2019 with days-covered ending prior to June 30, 2019 and no other BP claims thereafter, and (iv) users of zoledronic acid in January or February 2019 with no other BP claims during the remainder of the study period; and, two cohorts with short-term BP exposure, specifically new users of (v) alendronic acid or (vi) zoledronic acid in February 2020, with no prior BP claims during the pre-observation period.

### Covariates

As covariates, we considered factors that may influence either the use of BPs or potential modulators of primary or secondary study endpoints. These included: age; gender; insurance type (commercial, dual, Medicaid, Medicare); having had any primary care physician (PCP) visit in 2019; and comorbidity burden. The variable ‘PCP visit in 2019’ was used to control for prior healthcare-use behaviour and was assigned based on any physician office claim from January 1, 2019 to December 31, 2019 with one of the following provider types: family practice, general practice, geriatric medicine, internal medicine, and preventive medicine. Comorbidity score assignment was calculated following the Charlson Comorbidity Index (CCI) methodology^21^, and was based on diagnosis codes present on any medical claim (inpatient or outpatient) occurring during the pre-observation period. The assigned CCI score was used as the comorbidity covariate for the primary cohort propensity score matching, but to better control for differences in comorbidity burden when assessing outcomes, all regression analyses involving the primary analysis cohort included the following individual comorbidity covariates in lieu of the aggregate CCI score: osteoporosis, cancer, chronic obstructive pulmonary disease (COPD), depression, dyslipidaemia, hypertension, obesity, type 2 diabetes, cardiovascular disease overall, sickle cell anemia, stroke, dementia, HIV/AIDS, chronic kidney disease/end-stage renal disease (CKD/ESRD), and liver disease (Appendix 1).

### Cohort Matching

For the primary analysis, BP users were propensity-score (PS) matched to BP non-users via a PS calculated using multiple variables, including age, gender, insurance type, CCI, and any PCP visit in 2019, to yield comparable populations by demographics and clinical characteristics (Fig. 1B). To account for the differential geographic spread of COVID-19 across the U.S. during the observation period, matching was performed within each geographic region separately (Northeast, Midwest, South, West) and then combined. In addition to this within-region stratified match, a cohort build was also performed after restricting to patients from New York (NY) state only, since this state was the site of the largest outbreak in the initial COVID-19 surge in the U.S. All matching algorithms used a greedy-match propensity score technique^22^ to match BP users to non-users with a maximum permitted propensity-score difference of 0.015.

### Definition of Endpoints

Primary and secondary endpoints were assigned using inpatient and outpatient medical claims that occurred during the four-month observation period. The primary endpoint, COVID-19-related hospitalization, was assigned based on the presence of an International Classification of Diseases, Tenth Revision (ICD-10) code on any inpatient medical service claim indicating test-confirmed 2019 Novel Coronavirus (2019-nCoV) acute respiratory disease, specifically U07.1. The first secondary endpoint, SARS-CoV-2 testing, was assigned using Current Procedural Terminology (CPT) codes indicating a test for active infection, specifically 87635, 87636, and 87637. The second secondary endpoint, COVID-19-related diagnosis, was assigned based on any medical service claim with the ICD-10 diagnosis code U07.1.

### Statistical Analysis

To analyze data, mySQL, Excel, and SAS 9.4 was used on a secure hosted environment provided by Komodo Health. All data used in this study was subject to data restrictions per working agreements with Komodo Health, which dictated that only aggregate results can be exported (no patient-level data), so no deidentified data can be shared.

Unadjusted analyses assessing the association between BP-use and COVID-19-related outcomes were performed for the primary analysis cohort using chi-square tests for categorical variables and calculation of the crude unadjusted odds ratio (OR) in the matched cohort groups overall, when stratified by region and in NY state alone, and when further stratified by age group and gender. Chi-square tests for categorical variables and t-tests for continuous variables were also performed to assess differences in demographic and clinical characteristics of BP users compared to BP non-users both pre-match and post-match to assess the success of the propensity-score match.

Multivariate logistic regression analyses, modelled separately to determine the adjusted OR for each COVID-19-related primary and secondary outcome while adjusting for demographic and clinical characteristics, were performed on the matched primary analysis cohort with all regions combined, when stratified by region, and in NY state alone. The primary exposure of interest was BP-use (yes/no) during the pre-observation period. Additional demographic/clinical characteristics also included as regression model covariates were: age group, gender, region (for all regions-combined analyses), insurance type, PCP visit in 2019, and the following comorbid conditions: osteoporosis, cancer, COPD, depression, dyslipidaemia, hypertension, obesity, type 2 diabetes, cardiovascular disease overall, sickle cell anaemia, stroke, dementia, HIV/AIDS, CKD/ESRD, and liver disease.

All tests were two-tailed, and *p*-values of less than 0.05 were considered significant. All analyses were performed using SAS 9.4 (Cary, NC).

### Sensitivity Analyses

Multiple sensitivity analyses were performed to assess the reliability of the primary analysis results and/or to address potential unmeasured confounding (full details in Appendix 1).

1. The first sensitivity analysis addressed potential confounding by indication (i.e. the possibility of the indication for BP use rather than BP use itself being responsible for differences in outcomes among BP users and non-users) by restricting the control group to an active comparator cohort of patients who had used non-BP anti-resorptive bone medications during the pre-observation period. Users of non-BP anti-resorptive bone medications, the smaller patient population, were then 1:1 matched to BP users, providing a sample where all patients had used bone health medications during the pre-observation period (“*Bone-Rx*” cohort) (Fig. 1B). Cohort matching and regression modelling were performed following the same methodology employed for the primary analysis.
2. The second sensitivity analysis further addressed potential baseline differences between users of BPs and users of non-BP anti-resorptive bone medications in terms of indication for treatment and risk of SARS-CoV-2 exposure. To homogenise indication for treatment, we restricted the “Bone-Rx” cohort to females aged older than 50 years with an osteoporosis diagnosis (ICD-10: M80.x, M81.x, M82.x), which is the main (but not the only) indication for use of anti-resorptive bone medications. In order to homogenise risk of COVID-19 exposure, we additionally (a) restricted both groups to residents of New York, Illinois, Florida, and California (four states with a high incidence of COVID-19 cases during the observation period, with each representing a geographic region)^23^, and (b) matched within each state by insurance-type strata (i.e. BP non-users matched to BP users with Medicaid coverage residing in New York) to control for differences in socioeconomic characteristics. Non-BP anti-resorptive bone medication users were then matched to BP users by age, PCP visit in 2019, and the following select comorbid conditions that include those thought to impact COVID-19 severity: cancer, COPD, depression, dyslipidaemia, heart failure, hypertension, obesity, and type 2 diabetes^24^. In addition to assessing COVID-19-related outcomes, the matched cohorts that resulted from this analysis, older female patients from New York, Illinois, Florida, or California with a diagnosis of osteoporosis who were users of BP or non-BP anti-resorptive medications (“*Osteo-Dx-Rx*” cohort), were used for the third and fourth sensitivity analyses (see below).
3. The third sensitivity analysis evaluated the relationship between BP-use and exploratory negative control outcomes that were not anticipated to be impacted by the pharmacological mechanism of BPs. Separate comparisons were made in the primary cohort, the “*Bone-Rx*” cohort, and the “*Osteo-Dx-Rx*” cohort to evaluate the odds of any acute cholecystitis or acute pancreatitis-related inpatient/emergency-room (ER) service, which were chosen since both would present in an acute manner that require emergency intervention. These exploratory outcomes were assessed: (1) during the observation period among the already matched BP user/non-user populations; and (2) during a pre-pandemic period (the second half of 2019) when restricted to BP users in the first half of 2019 and their previously-assigned BP non-user matched pair to mitigate any COVID-19-related impacts on healthcare resource utilization.
4. The fourth sensitivity analysis assessed the relationship between BP-use and exploratory positive control outcomes (anticipated to be impacted by the immunomodulatory pharmacological mechanism of BPs) occurring in 2019. For this analysis, the primary, “*Bone-Rx*”, and “*Osteo-Dx-Rx*” cohorts were restricted to BP users who had any BP claim during the first half of 2019 and their previously-assigned BP non-user matched pair to assess the relationship between BP-use and medical services for other respiratory infectious diseases (acute bronchitis, pneumonia).
5. The fifth sensitivity analysis addressed potential bias due to the ‘healthy adherer’ effect, whereby users of a preventive drug may have better disease outcomes due to their healthier behaviours rather than due to drug treatment itself^25^. Two strategies were employed to validate the findings from our primary analysis while controlling for the potential impact of healthy adherer effect-associated bias. First, we tested whether effects observed with exposure to BPs were similarly observed with exposure to other preventive drugs, namely statins, antihypertensives, antidiabetics, and antidepressants. Second, we assessed whether the association between BP-use and COVID-19-related outcomes was maintained among the matched user/non-user populations of these other preventive drugs, i.e. BP users were compared to BP non-users within, for example, the statin user population and separately within the matched statin non-user population.

## RESULTS

### Study Population

A total of 8,239,790 patients met the inclusion criterion of continuous medical and prescription insurance eligibility over the full study period, of which 333,107 were excluded due to missing demographic information, resulting in a total eligible sample of 7,906,603 patients (Fig. 1B). Of this full population, 452,051 (5.7%) and 7,454,552 (94.3%) patients were classified as BP users and BP non-users, respectively. Within BP users, more than 99% were prescribed an amino-BP, with oral alendronic acid (75.4%), zoledronic acid infusion (11.5%), and oral ibandronic acid (8.4%) as the most prevalent formulations (Table S1).

Prior to propensity-score matching, there were significant differences between BP users and non-users across all demographic and clinical characteristics. BP users were older (age >60: 82.7% versus 27.7%; p<0.001), predominantly female (91.0% versus 57.2%; p<0.001), with a higher comorbidity burden (mean CCI 0.95 versus 0.60; p<0.001), with a larger proportion of patients residing in the Western U.S. (21.1% versus 15.4%; p<0.001), covered by Medicare (43.3% versus 13.7%; p<0.001), and having visited a PCP in 2019 (63.8% versus 44.7%; p<0.001). Propensity-score matching yielded 450,366 BP users and 450,366 BP non-users with no significant differences across all characteristics used in matching (Table 1). Differences did exist, however, in the distribution of individual comorbid condition indicators that were used as covariates in the regression analysis, with the BP non-user cohort having a higher proportion of patients with COPD (10.2% versus 8.5%; p<0.001), cardiovascular disease (25.1% versus 18.7%; p<0.001), dyslipidemia (36.9% versus 34.6%; p<0.001), hypertension (46.4% versus 38.8%; p<0.001), obesity (10.3% versus 6.7%; p<0.001), and type 2 diabetes (22.9% versus 18.2%; p<0.001). Over 98% of all BP user/non-user matches for the primary analysis cohort were completed with differences in matched propensity scores <0.000001 (overall mean difference of 0.000004, max difference of 0.0147).

**Table 1:**
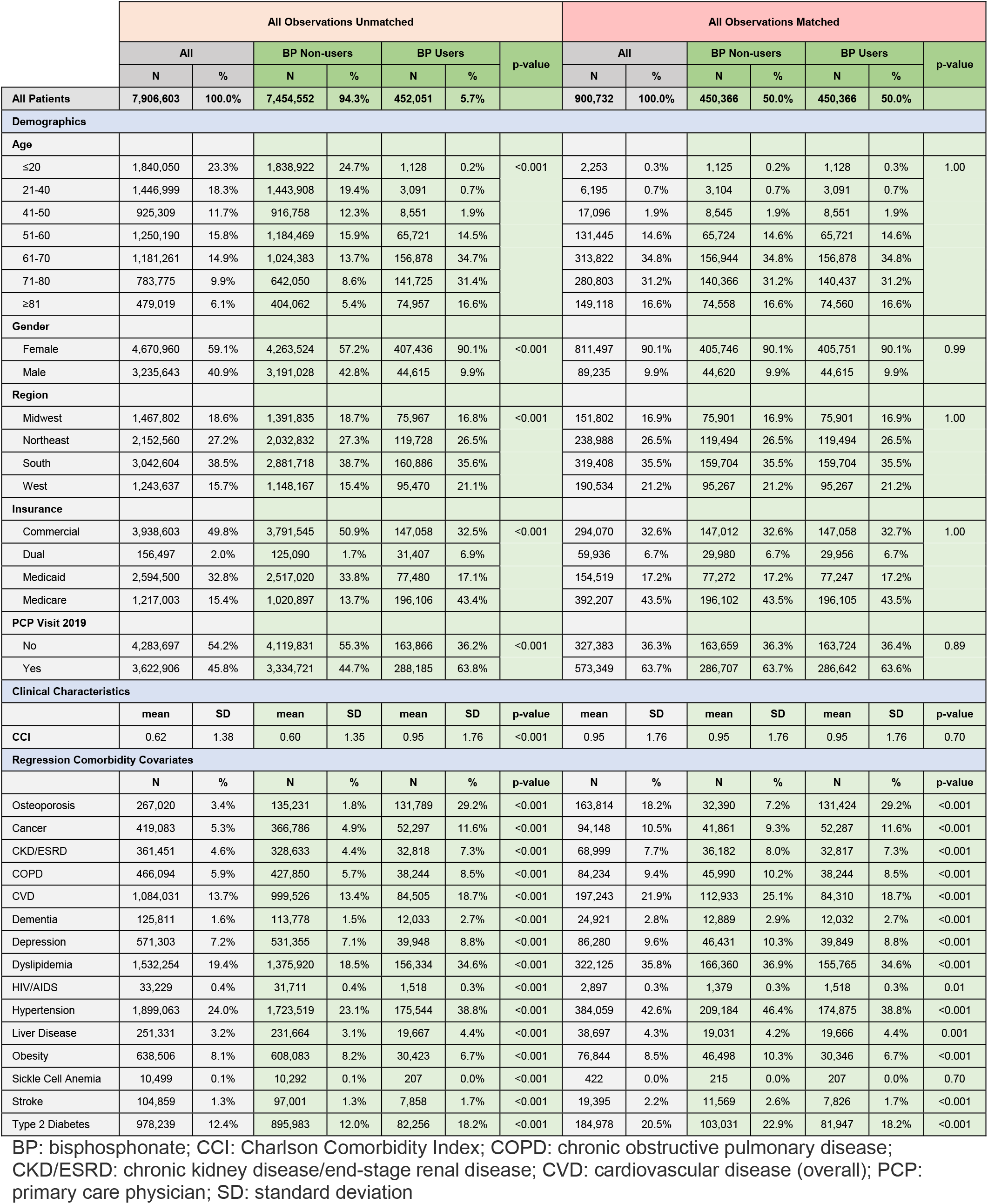
Primary Analysis Cohort (All Regions), Patient Characteristics Pre/Post Match

|  | All Observations Unmatched |  |  |  |  |  |  | All Observations Matched |  |  |  |  |  |  |
| --- | --- | --- | --- | --- | --- | --- | --- | --- | --- | --- | --- | --- | --- | --- |
|  | All |  | BP Non-users |  | BP Users |  | p-value | All |  | BP Non-users |  | BP Users |  | p-value |
|  | N | % | N | % | N | % |  | N | % | N | % | N | % |  |
| All Patients | 7,906,603 | 100.0% | 7,454,552 | 94.3% | 452,051 | 5.7% |  | 900,732 | 100.0% | 450,366 | 50.0% | 450,366 | 50.0% |  |
| Demographics |  |  |  |  |  |  |  |  |  |  |  |  |  |  |
| Age |  |  |  |  |  |  |  |  |  |  |  |  |  |  |
| ≤20 | 1,840,050 | 23.3% | 1,838,922 | 24.7% | 1,128 | 0.2% | <0.001 | 2,253 | 0.3% | 1,125 | 0.2% | 1,128 | 0.3% | 1.00 |
| 21-40 | 1,446,999 | 18.3% | 1,443,908 | 19.4% | 3,091 | 0.7% |  | 6,195 | 0.7% | 3,104 | 0.7% | 3,091 | 0.7% |  |
| 41-50 | 925,309 | 11.7% | 916,758 | 12.3% | 8,551 | 1.9% |  | 17,096 | 1.9% | 8,545 | 1.9% | 8,551 | 1.9% |  |
| 51-60 | 1,250,190 | 15.8% | 1,184,469 | 15.9% | 65,721 | 14.5% |  | 131,445 | 14.6% | 65,724 | 14.6% | 65,721 | 14.6% |  |
| 61-70 | 1,181,261 | 14.9% | 1,024,383 | 13.7% | 156,878 | 34.7% |  | 313,822 | 34.8% | 156,944 | 34.8% | 156,878 | 34.8% |  |
| 71-80 | 783,775 | 9.9% | 642,050 | 8.6% | 141,725 | 31.4% |  | 280,803 | 31.2% | 140,366 | 31.2% | 140,437 | 31.2% |  |
| ≥81 | 479,019 | 6.1% | 404,062 | 5.4% | 74,957 | 16.6% |  | 149,118 | 16.6% | 74,558 | 16.6% | 74,560 | 16.6% |  |
| Gender |  |  |  |  |  |  |  |  |  |  |  |  |  |  |
| Female | 4,670,960 | 59.1% | 4,263,524 | 57.2% | 407,436 | 90.1% | <0.001 | 811,497 | 90.1% | 405,746 | 90.1% | 405,751 | 90.1% | 0.99 |
| Male | 3,235,643 | 40.9% | 3,191,028 | 42.8% | 44,615 | 9.9% |  | 89,235 | 9.9% | 44,620 | 9.9% | 44,615 | 9.9% |  |
| Region |  |  |  |  |  |  |  |  |  |  |  |  |  |  |
| Midwest | 1,467,802 | 18.6% | 1,391,835 | 18.7% | 75,967 | 16.8% | <0.001 | 151,802 | 16.9% | 75,901 | 16.9% | 75,901 | 16.9% | 1.00 |
| Northeast | 2,152,560 | 27.2% | 2,032,832 | 27.3% | 119,728 | 26.5% |  | 238,988 | 26.5% | 119,494 | 26.5% | 119,494 | 26.5% |  |
| South | 3,042,604 | 38.5% | 2,881,718 | 38.7% | 160,886 | 35.6% |  | 319,408 | 35.5% | 159,704 | 35.5% | 159,704 | 35.5% |  |
| West | 1,243,637 | 15.7% | 1,148,167 | 15.4% | 95,470 | 21.1% |  | 190,534 | 21.2% | 95,267 | 21.2% | 95,267 | 21.2% |  |
| Insurance |  |  |  |  |  |  |  |  |  |  |  |  |  |  |
| Commercial | 3,938,603 | 49.8% | 3,791,545 | 50.9% | 147,058 | 32.5% | <0.001 | 294,070 | 32.6% | 147,012 | 32.6% | 147,058 | 32.7% | 1.00 |
| Dual | 156,497 | 2.0% | 125,090 | 1.7% | 31,407 | 6.9% |  | 59,936 | 6.7% | 29,980 | 6.7% | 29,956 | 6.7% |  |
| Medicaid | 2,594,500 | 32.8% | 2,517,020 | 33.8% | 77,480 | 17.1% |  | 154,519 | 17.2% | 77,272 | 17.2% | 77,247 | 17.2% |  |
| Medicare | 1,217,003 | 15.4% | 1,020,897 | 13.7% | 196,106 | 43.4% |  | 392,207 | 43.5% | 196,102 | 43.5% | 196,105 | 43.5% |  |
| PCP Visit 2019 |  |  |  |  |  |  |  |  |  |  |  |  |  |  |
| No | 4,283,697 | 54.2% | 4,119,831 | 55.3% | 163,866 | 36.2% | <0.001 | 327,383 | 36.3% | 163,659 | 36.3% | 163,724 | 36.4% | 0.89 |
| Yes | 3,622,906 | 45.8% | 3,334,721 | 44.7% | 288,185 | 63.8% |  | 573,349 | 63.7% | 286,707 | 63.7% | 286,642 | 63.6% |  |
| Clinical Characteristics |  |  |  |  |  |  |  |  |  |  |  |  |  |  |
|  | mean | SD | mean | SD | mean | SD | p-value | mean | SD | mean | SD | mean | SD | p-value |
| CCI | 0.62 | 1.38 | 0.60 | 1.35 | 0.95 | 1.76 | <0.001 | 0.95 | 1.76 | 0.95 | 1.76 | 0.95 | 1.76 | 0.70 |
| Regression Comorbidity Covariates |  |  |  |  |  |  |  |  |  |  |  |  |  |  |
|  | N | % | N | % | N | % | p-value | N | % | N | % | N | % | p-value |
| Osteoporosis | 267,020 | 3.4% | 135,231 | 1.8% | 131,789 | 29.2% | <0.001 | 163,814 | 18.2% | 32,390 | 7.2% | 131,424 | 29.2% | <0.001 |
| Cancer | 419,083 | 5.3% | 366,786 | 4.9% | 52,297 | 11.6% | <0.001 | 94,148 | 10.5% | 41,861 | 9.3% | 52,287 | 11.6% | <0.001 |
| CKD/ESRD | 361,451 | 4.6% | 328,633 | 4.4% | 32,818 | 7.3% | <0.001 | 68,999 | 7.7% | 36,182 | 8.0% | 32,817 | 7.3% | <0.001 |
| COPD | 466,094 | 5.9% | 427,850 | 5.7% | 38,244 | 8.5% | <0.001 | 84,234 | 9.4% | 45,990 | 10.2% | 38,244 | 8.5% | <0.001 |
| CVD | 1,084,031 | 13.7% | 999,526 | 13.4% | 84,505 | 18.7% | <0.001 | 197,243 | 21.9% | 112,933 | 25.1% | 84,310 | 18.7% | <0.001 |
| Dementia | 125,811 | 1.6% | 113,778 | 1.5% | 12,033 | 2.7% | <0.001 | 24,921 | 2.8% | 12,889 | 2.9% | 12,032 | 2.7% | <0.001 |
| Depression | 571,303 | 7.2% | 531,355 | 7.1% | 39,948 | 8.8% | <0.001 | 86,280 | 9.6% | 46,431 | 10.3% | 39,849 | 8.8% | <0.001 |
| Dyslipidemia | 1,532,254 | 19.4% | 1,375,920 | 18.5% | 156,334 | 34.6% | <0.001 | 322,125 | 35.8% | 166,360 | 36.9% | 155,765 | 34.6% | <0.001 |
| HIV/AIDS | 33,229 | 0.4% | 31,711 | 0.4% | 1,518 | 0.3% | <0.001 | 2,897 | 0.3% | 1,379 | 0.3% | 1,518 | 0.3% | 0.01 |
| Hypertension | 1,899,063 | 24.0% | 1,723,519 | 23.1% | 175,544 | 38.8% | <0.001 | 384,059 | 42.6% | 209,184 | 46.4% | 174,875 | 38.8% | <0.001 |
| Liver Disease | 251,331 | 3.2% | 231,664 | 3.1% | 19,667 | 4.4% | <0.001 | 38,697 | 4.3% | 19,031 | 4.2% | 19,666 | 4.4% | 0.001 |
| Obesity | 638,506 | 8.1% | 608,083 | 8.2% | 30,423 | 6.7% | <0.001 | 76,844 | 8.5% | 46,498 | 10.3% | 30,346 | 6.7% | <0.001 |
| Sickle Cell Anemia | 10,499 | 0.1% | 10,292 | 0.1% | 207 | 0.0% | <0.001 | 422 | 0.0% | 215 | 0.0% | 207 | 0.0% | 0.70 |
| Stroke | 104,859 | 1.3% | 97,001 | 1.3% | 7,858 | 1.7% | <0.001 | 19,395 | 2.2% | 11,569 | 2.6% | 7,826 | 1.7% | <0.001 |
| Type 2 Diabetes | 978,239 | 12.4% | 895,983 | 12.0% | 82,256 | 18.2% | <0.001 | 184,978 | 20.5% | 103,031 | 22.9% | 81,947 | 18.2% | <0.001 |

BP: bisphosphonate; CCI: Charlson Comorbidity Index; COPD: chronic obstructive pulmonary disease; CKD/ESRD: chronic kidney disease/end-stage renal disease; CVD: cardiovascular disease (overall); PCP: primary care physician; SD: standard deviation

Similar profiles in pre-match *versus* post-match characteristics were seen when patients were stratified by region or restricted to NY-state (Tables S2A-E). Demographic distributions, including differences between BP user *versus* BP non-user characteristics pre- and post-matching for all sensitivity analysis cohorts are detailed in Appendix 2.

### BP use and COVID-19-Related Outcomes

Among the full matched cohort, BP users had significantly lower rates and unadjusted (crude) odds of testing (1.2% vs. 5.1%; OR=0.22; 95%CI:0.21-0.22; p<0.001), diagnosis (0.7% vs. 2.9%; OR=0.22; 95%CI:0.21-0.23; p<0.001), and hospitalization (0.2% vs. 0.7%; OR=0.24; 95%CI:0.22-0.26; p<0.001) as compared to BP non-users (Fig. 2). Consistent findings were seen when sub-stratifying the full matched cohort by age, gender, age*gender, within grouped regions, by individual region, and in NY-state alone (Tables S3A-F).

**Figure 2:**
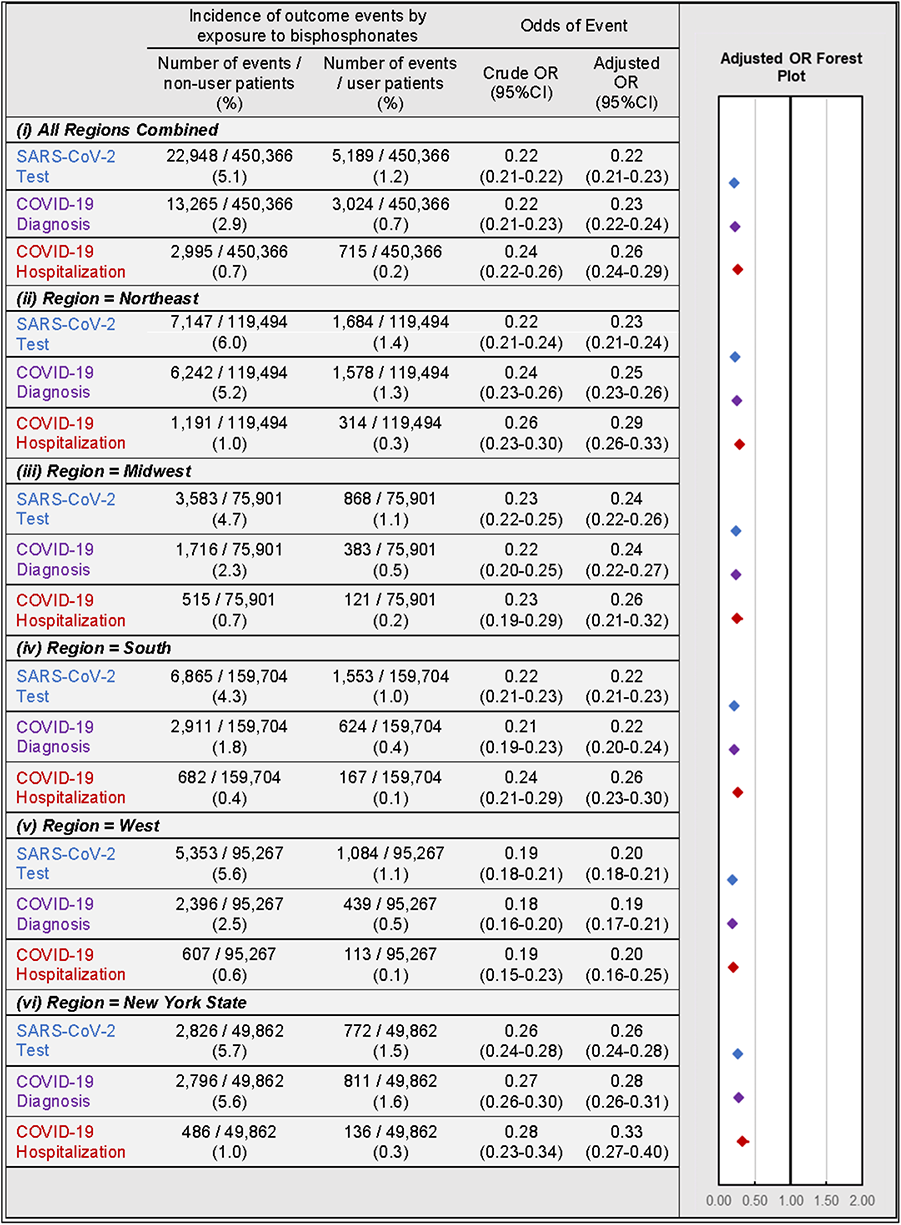
Association of BP use and COVID-19-Related Outcomes. Incidence (left) and regression-adjusted results for odds (right) of SARS-CoV-2 testing (blue), COVID-19 diagnosis (purple), and COVID-19-related hospitalizations (red) of BP users compared with BP non-users in the all-regions combined primary analysis cohort (i) and when stratified by region/state into: Northeast (ii), Midwest (iii), South (iv), West (v), and New York state (vi).

Multivariate regression analyses yielded similar results for all outcomes while additionally controlling for patient demographic and comorbidity characteristics. In the full matched cohort, BP users had lower adjusted odds of testing (OR=0.22; 95%CI:0.21-0.23; p<0.001), diagnosis (OR=0.23; 95%CI:0.22-0.24; p<0.001), and hospitalizations (OR=0.26; 95%CI:0.24-0.29; p<0.001). These findings were robust when comparing BP users with BP non-users when stratified by geographic region or NY-state alone.

### Timing of last BP exposure and COVID-19-Related Outcomes

The above results demonstrate that any BP exposure during the 14-months pre-observation period is associated with a marked reduction in each of the three COVID-19-related outcomes. To further investigate the relationship between COVID-19-related outcomes and the timing of BP exposure, we focused on the two most commonly prescribed BPs, alendronic acid (oral formulation dosed daily or weekly) and zoledronic acid (infusion dosed annually). For each BP type, COVID-19-related outcomes were assessed among users: (i-ii) with exposure or days covered (based on prescription frequency) during the pre-observation period and throughout the observation period; (iii-iv) with exposure or days covered ending prior to the observation period; and (v-vi) newly initiating therapy prior to the observation period (Fig. 3A). Strikingly, all subgroups of BP users had decreased odds of COVID-19-related outcomes (Fig. 3B) except for the odds of hospitalization among zoledronic acid users who were last dosed in January/February of 2019 (OR=0.52; 95%CI:0.20-1.40; p=0.20) or newly initiated in February of 2020 (OR=0.49; 95%CI:0.13-1.88; p=0.30).

**Figure 3:**
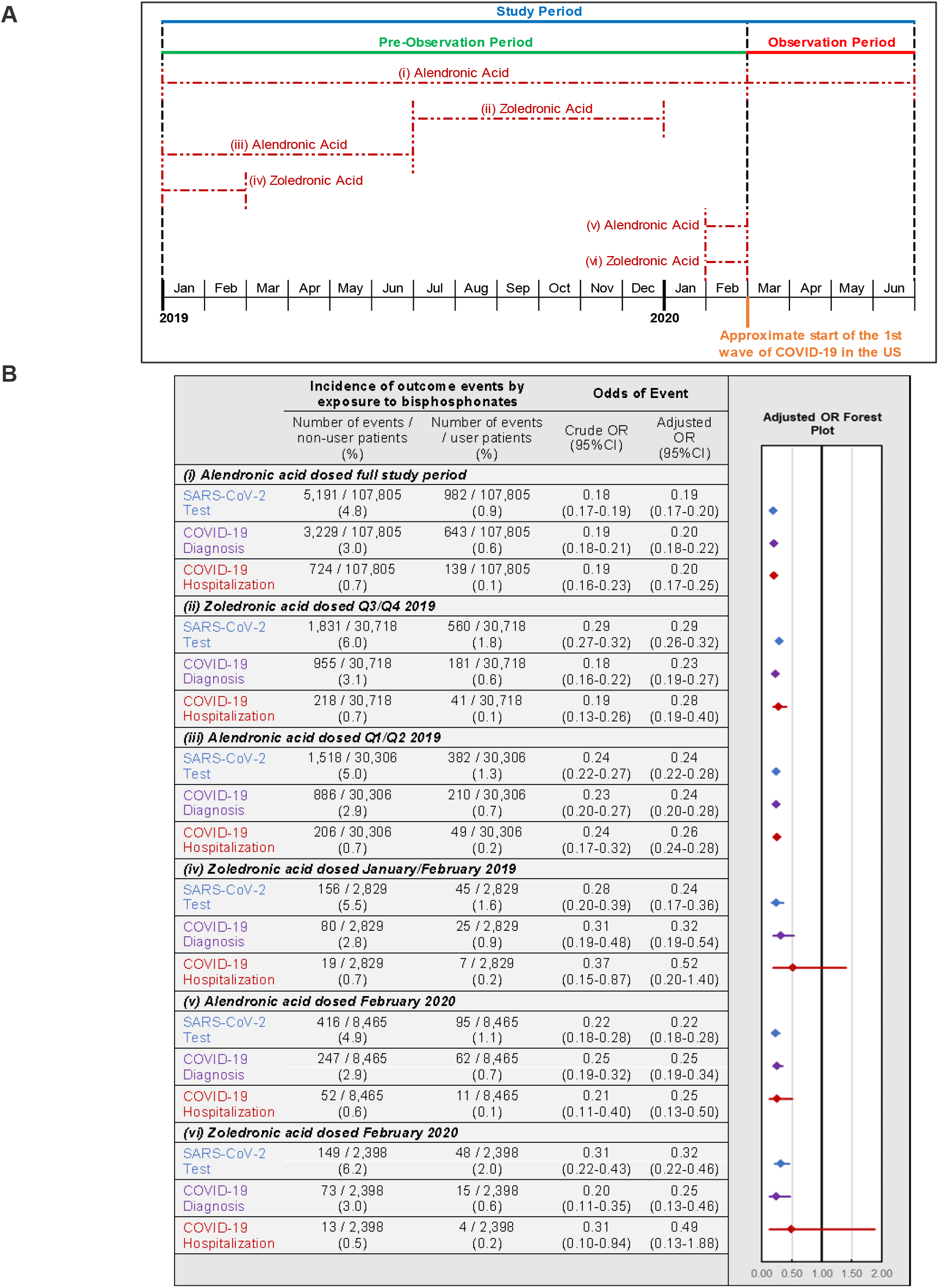
Timing of BP use and COVID-19-Related Outcomes. **A.** Schematic of BP user sub-stratification by timing of exposure to alendronic acid or zoledronic acid prior to outcome assessment. Broken lines represent periods of active BP dosing. For zoledronic acid users, days covered was considered to extend 1 year past the dosing period based on dosing guidelines. **B.** Incidence (left) and regression-adjusted results (right) for odds of SARS-CoV-2 testing, COVID-19 diagnosis, and COVID-19-related hospitalizations of BP users compared with BP non-users in pre-specified subgroups. *CI: confidence interval; OR: odds ratio*.

### Sensitivity Analysis 1: COVID-19-Related Outcomes Among All Users of Anti-Resorptive Medications (“Bone-Rx” Cohort)

The first sensitivity analysis was performed to address potential confounding by indication. To validate our primary findings in more comparable cohorts, analysis was restricted to comparing BP users to patients using non-BP anti-resorptive bone medications during the pre-observation period. Compared to non-BP users of anti-resorptive medications, BP users had decreased odds of testing (OR=0.31; 95%CI:0.28-0.33; p<0.001), diagnosis (OR=0.35; 95%CI:0.31-0.38; p<0.001), and hospitalization (OR=0.45; 95%CI:0.36-0.56; p<0.001) (Fig. 4A). Furthermore, these findings were robust when assessed separately across every geographic region as well as NY state for all outcomes except hospitalizations when restricted to the Western U.S. (p=0.08; Table S4).

**Figure 4:**
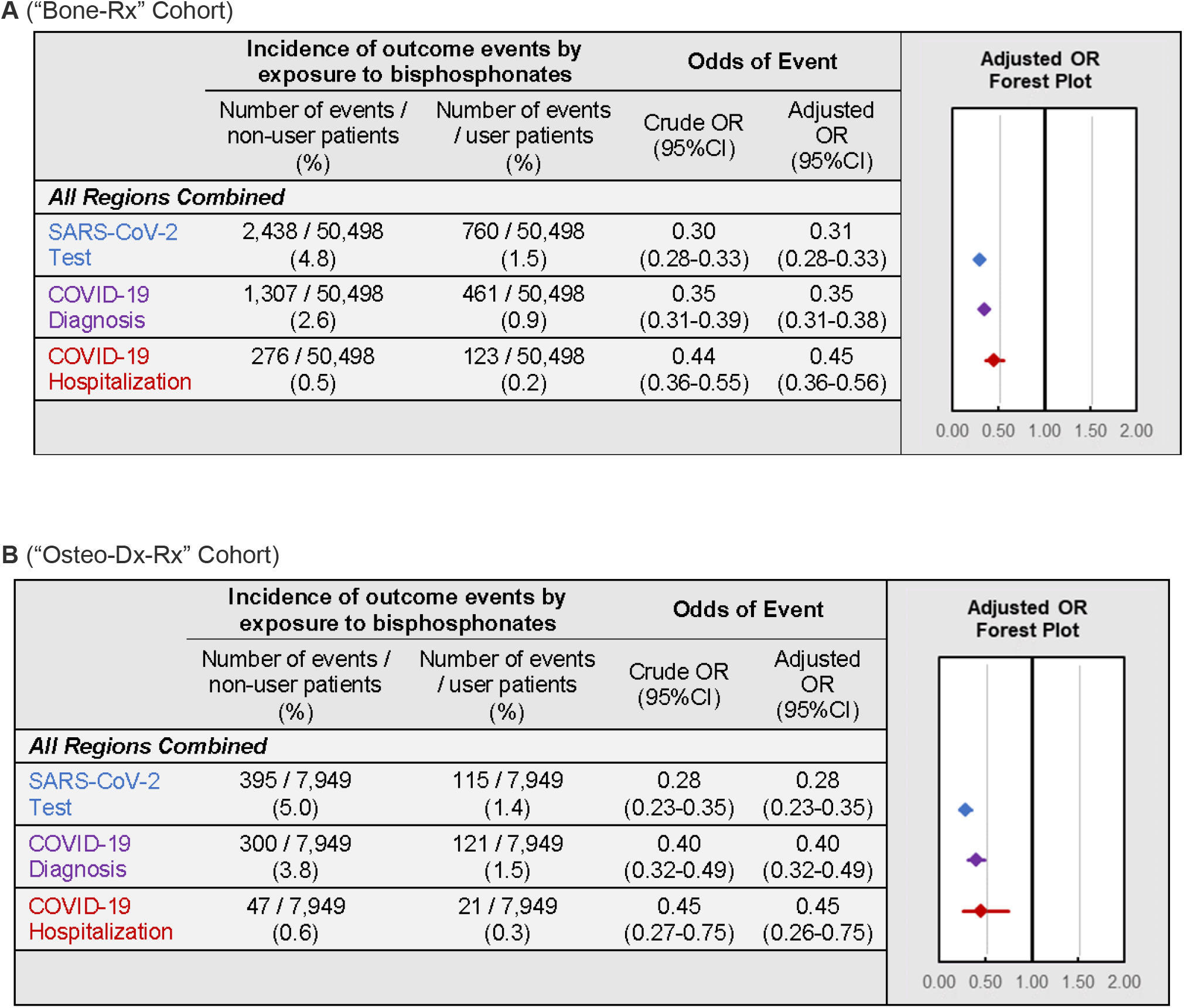
COVID-19-Related Outcomes Among the Bone-RX and Osteo-Dx-Rx Restricted Cohorts. Incidence and forest plots summarizing regression-adjusted odds ratios of SARS-CoV-2 testing (blue), COVID-19 diagnosis (purple), ANDD COVID-19-related hospitalizations (red) in the (**A**) “*Bone-Rx*” and (**B**) “*Osteo-Dx-Rx*” sensitivity analysis cohorts.

### Sensitivity Analysis 2: COVID-19-Related Outcomes Among Users of Anti-Resorptive Medications with a Diagnosis of Osteoporosis (“Osteo-Dx-Rx” Cohort)

The second sensitivity analysis was performed to address the fact that, even after restricting the comparator cohort to users of anti-resorptive medications, differences may still exist between patient cohorts that could affect COVID-19-related outcomes, including different indications for anti-resorptive medication use and other uncontrolled patient characteristics. To address this, the association between BP use and COVID-19 related outcomes were examined in a cohort restricted to female patients over 50 years old, with a diagnosis of osteoporosis, using either a BP or a non-BP anti-resorptive bone medication, matched within insurance-type as a proxy for socioeconomic status, and selected from four states (NY, IL, FL, CA) with high incidences of COVID-19 cases during the observation period^23^ (“*Osteo-Dx-Rx*” cohort). In agreement with the results reported above, the decrease in odds of COVID-19-related outcomes in BP users remained robust for testing (OR=0.28; 95%CI:0.23-0.35; p<0.001), diagnosis (OR=0.40; 95%CI:0.32-0.49; p<0.001), and hospitalizations (OR=0.45; 95%CI:0.26-0.75; p=0.003) (Figure 4B).

### Sensitivity Analysis 3: Association of BP-use with Exploratory Negative Control Outcomes

While the above sensitivity analyses were performed to control for health status differences between BP users and BP non-users during the pre-observation period, it remained theoretically possible that BP users exhibited differential health care access that might have impacted COVID-19-related outcomes. To examine such residual confounding, the third sensitivity analysis evaluated the incidence of exploratory outcomes not hypothesized to be modulated by BP-use in the primary, “*Bone-Rx*”, and “*Osteo-Dx-Rx*” cohorts. Outcomes were measured during the observation period, as well as during a pre-pandemic period (the second half of 2019), to control for potential reduction in the use of healthcare services^26, 27^ during the COVID-19 pandemic. The associations between BP-use and the following exploratory outcomes, which were anticipated to not be impacted by BP-use, were assessed: any acute cholecystitis or acute pancreatitis-related inpatient/ER service. Regression modelling found BP users in the primary analysis cohort to have decreased odds of any inpatient/ER service for acute cholecystitis (OR=0.55; 95%CI:0.44-0.69; p<0.001) and acute pancreatitis (OR=0.58; 95%CI:0.49-0.69; p<0.001) during the observation period (Fig. 5A), as well as decreased odds of acute pancreatitis (OR=0.83; 95%CI:0.71-0.96; p=0.01) during the second half of 2019. BP users in the “*Bone-Rx*” cohort displayed a decreased odds of service for acute pancreatitis (OR=0.55; 95%CI:0.36-0.87; p=0.01) only during the study observation period and no significant difference in odds of service for acute cholecystitis during either periods. Lastly, there was no difference in any outcome between BP users and BP non-users in the “*Osteo-Dx-Rx*” cohort, although there were very few events in this cohort.

**Figure 5:**
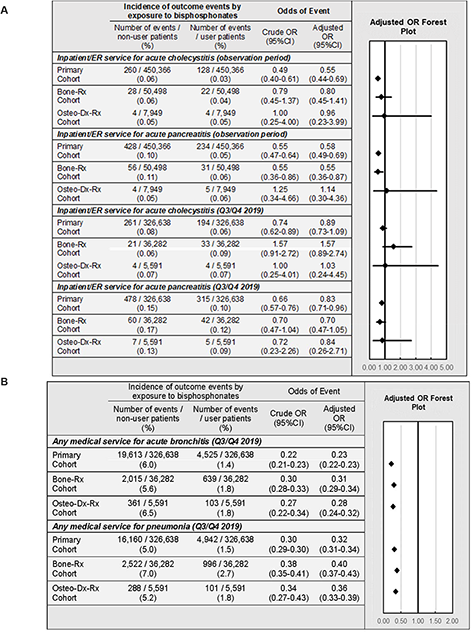
Exploratory Outcomes among BP Users versus BP Non-users. Incidence and adjusted odds ratios of (**A**) expected negative exploratory outcomes and (**B**) other respiratory infections, in the primary, “*Bone-Rx*”, and “*Osteo-Dx-Rx*” cohorts. *CI: confidence interval; ER: emergency room; OR: odds ratio*.

### Sensitivity Analysis 4: Association of BP-use with Exploratory Positive Control Outcomes

The fourth sensitivity analysis was performed to assess if there is an association between BP-use and incidence of other respiratory infections, which has been previously reported^16^. Medical services for acute bronchitis or pneumonia were measured during the second half of 2019, prior to the advent of COVID-19, in the primary, “*Bone-Rx*”, and “*Osteo-Dx-Rx*” cohorts. Regression modelling found that, among all cohort variations modelled, BP users had a decreased odds of any medical service related to acute bronchitis (point estimates of ORs ranged from 0.23 to 0.28) and pneumonia (point estimates of ORs ranged from 0.32 to 0.36) (Figure 5B).

### Sensitivity Analysis 5: Association of Other Preventive Drugs with COVID-19-Related Outcomes

A potential pitfall in the interpretation of apparent effects of preventive medications on health outcomes is the so-called healthy adherer effect, whereby patients may have better outcomes due to their overall healthier behaviours and not due to active drug treatment itself^25^. To address this possibility of unmeasured confounding, a final sensitivity analysis was performed to evaluate the association between control exposures (i.e. use of other preventive medications such as statins, antihypertensives, antidiabetics, and antidepressants) and COVID-19-related outcomes (Figure 6A). In comparison to BPs, the impact of other preventive drug classes on COVID-19-related outcomes was much weaker overall (Figure 6B-E) and varied between geographic regions in terms of magnitude or direction (Table S5A-D). Furthermore, when assessing the impact of BP-use within matched user/non-user preventive drug cohorts (e.g. BP users compared to BP non-users among the matched statin user and statin non-user populations), we found BP-use to be consistently associated with lower odds of testing (point estimates of ORs ranged from 0.21 to 0.27), diagnosis (point estimates of ORs ranged from 0.22 to 0.30), and hospitalizations (point estimates of ORs ranged from 0.25 to 0.33) across all stratified preventive user/non-user cohorts (Figure 6B-E).

**Figure 6:**
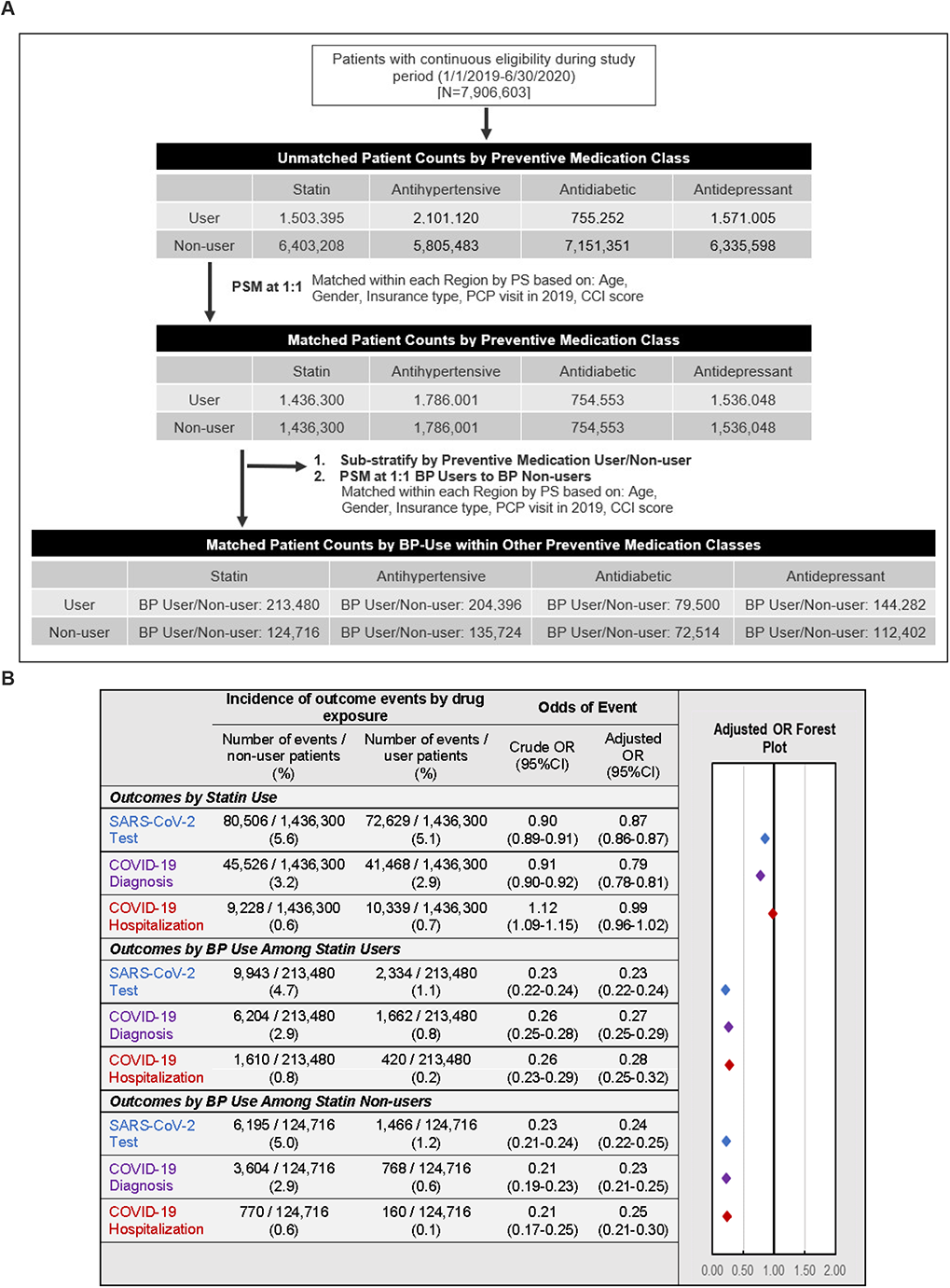
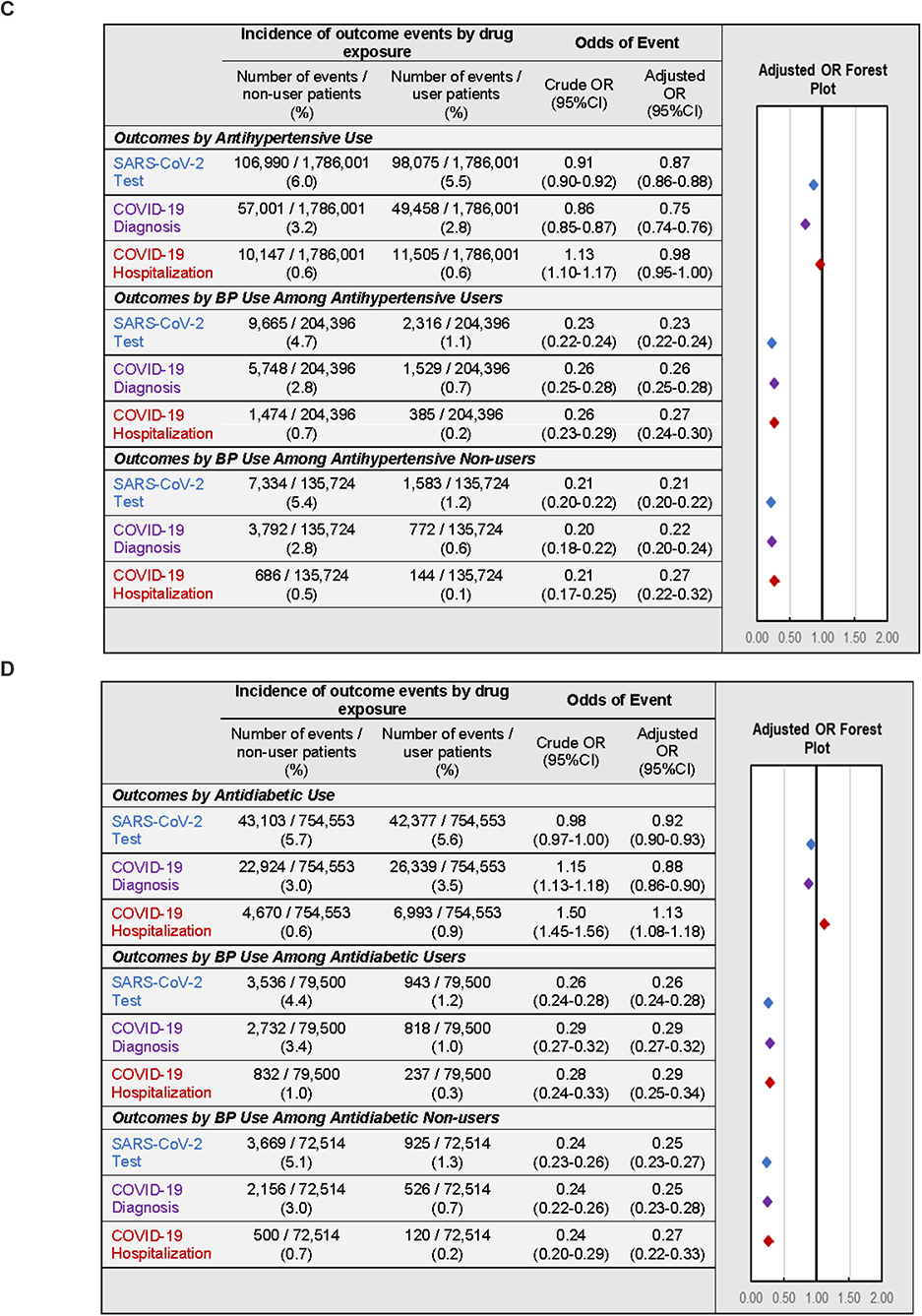
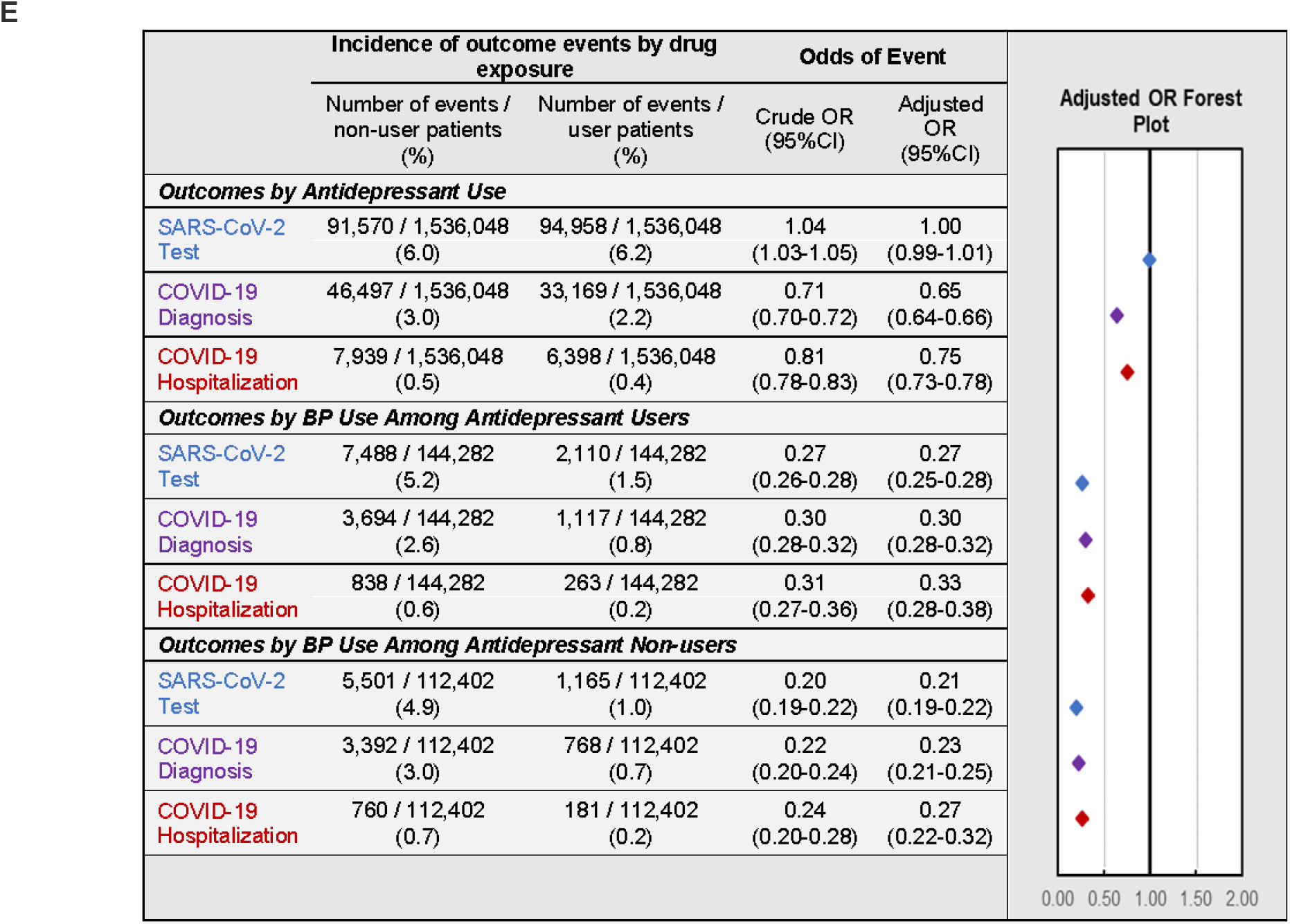
Association of Other Preventive Drugs with COVID-19-Related Outcomes. **A.** Schematic illustrating the identification of study populations and matched controls for each drug class. **B-E.** Incidence and adjusted odds ratios of SARS-CoV-2 testing (blue), COVID-19 diagnosis (purple), and COVID-19-related hospitalizations (red) in users and non-users of (**B**) statins, (**C**) antihypertensive medications, (**D**) non-insulin antidiabetic medications, and (**E**) antidepressant medications. For each class of preventive medications, further analysis was performed comparing BP users and BP non-users within matched cohorts of medication users (middle) and medication non-users (bottom). *BP: bisphosphonate; CCI: Charlson comorbidity index; CI: confidence interval; COPD: chronic obstructive pulmonary disease; OR: odds ratio; PCP: primary care physician; PS: propensity score; PSM: propensity score match*

## DISCUSSION

This study examined the association between recent exposure to BPs and subsequent COVID-19-related outcomes during the initial outbreak of the COVID-19 pandemic in the U.S. Our findings demonstrate that amino-BP users experienced a three- to five-fold reduced incidence of SARS-CoV-2 testing, COVID-19 diagnosis, and COVID-19-related hospitalization during this period. This dramatic difference in outcomes was consistently observed when comparing BP users to BP non-users in a propensity score-matched general population, when comparing to users of other anti-resorptive bone medications, when further restricting the latter cohort to female osteoporosis patients that were matched by comorbidities within state of residence and by insurance type, and when comparing BP users to BP non-users stratified by use of other preventive medications. Therefore, although there are confounding-related limitations inherent within retrospective studies, the consistency and strength of our observed associations when using various methods to control for unmeasured confounding support the contention that further prospective research should be performed to determine the true magnitude of the potential immunomodulatory effects of BP use.

Our findings are consistent with previous observational studies, prior to the advent of COVID-19, that had reported associations between BP use and reduced incidence of pneumonia and pneumonia-related mortality^16, 28, 29^. Accordingly, we observed in our population that BP use was associated with decreased odds of medical services for acute bronchitis and pneumonia during the second half of 2019. Taken together, these findings suggest that BPs may play a protective role in respiratory tract infections from a variety of causes, including SARS-CoV-2.

Other recent retrospective studies have explored, to some extent, associations of anti-resorptive medication use and COVID-19-related outcomes, albeit in much smaller patient populations than were analysed here. One study found no differences in the COVID-19-related risk of hospitalization, ICU admission, and mortality among 1,997 female patients diagnosed with COVID-19 who received anti-osteoporosis medication as compared to propensity score-matched COVID-19 patients who were not receiving such drugs^30^. This study did not examine the incidence of COVID-19 among BP users, but it raises the possibility that the subset of BP users who do develop sufficient pathology to be diagnosed with COVID-19 may have a similar clinical course as BP non-users. Another retrospective cohort study in Italy examining the association of oral amino-BP use and incidence of COVID-19-related hospitalization and mortality found no difference between BP users and BP non-users or users of non-BP anti-resorptive medications^31^. However, the overall incidence of COVID-19 hospitalization in the primary cohort (151/126,370 patients, or 0.12%) of this study was markedly lower than in the present analysis (3,710/900,732 patients, or 0.41%). A third study examined the influence of various anti-osteoporosis drugs, including BPs, on the cumulative incidence of COVID-19 in 2,102 patients with non-inflammatory rheumatic conditions that were compared to population estimates in the same geographic region^32^. In this analysis, users of non-BP anti-resorptive medications and zoledronate, but not users of oral BPs, had a lower incidence and relative risk of COVID-19 diagnosis and hospitalization. The observations with zoledronate are consistent with the findings reported here. However, we did not detect a significant impact of non-BP anti-resorptive medications in comparison to BPs, and we found a robust association between oral BP use and lower odds of COVID-19 diagnosis and related hospitalization. The reason for these discrepancies is unclear but could potentially reflect the large disparity in sample size between our study, which differed by more than three orders of magnitude. A fourth study, which used Israeli insurance data to perform an analysis involving two separate case-control matched cohorts to assess the risk of COVID-19 hospitalizations when stratified by recent medication use, also found that the odds COVID-19-related hospitalizations were lower among users of BPs, and ranged from an OR of 0.705 (95%CI: 0.522 to 0.935) to 0.567 (95%CI:0.400 to 0.789)^33^.

The large size of our dataset allowed for a range of fully powered, stratified analyses to be performed to explore the robustness of our findings and to address unmeasured confounding factors and other sources of potential bias that can occur in retrospective studies using insurance claims data. These sensitivity analyses included: (1) comparing the effect of BP use and active control exposure, namely non-BP anti-resorptive bone medication; (2) further reducing unmeasured confounding by restricting sensitivity analysis (1) to females older than 50 years, diagnosed with osteoporosis, matched within state and insurance type and matched by comorbid conditions thought to impact COVID-19 severity; (3) assessing the effect of BP-use on exploratory negative control outcomes (acute cholecystitis or pancreatitis related inpatient/ER service); (4) evaluating the effect of BP-use on exploratory positive control outcomes (other respiratory infections); and (5) evaluating the potential bias related to the ‘healthy adherer effect’ by assessing (a) the effect of other preventive medication use on COVID-19-related outcomes (as negative control exposures) and (b) the BP effect in users and non-users of other preventive medications.

Notwithstanding, a retrospective analysis of insurance claims data has inevitable limitations that should be considered. For example, the interpretation of our findings depends, in part, on the assumption that BP users and non-users had a similar risk of SARS-CoV-2 infection during the observation period. However, our dataset does not allow us to restrict patient observations to those with known exposure to SARS-CoV-2. Therefore, to minimize potential differences in SARS-CoV-2 exposure between BP users and non-users in our primary study cohort, we implemented additional analytical strategies, including the sensitivity analyses discussed above, as well as matching BP users to BP non-users within geographical regions and specific states.

Despite these efforts, it is important to note that we have limited information to assess and match BP users to BP non-users by sociodemographic risk factors, such as socio-economic status and racial/ethnic minority status, that are associated with COVID-19 incidence and mortality^34, 35^. In particular, our dataset does not include information on patients’ race or ethnic background. This is a relevant limitation given the documented impact of racial/ethnic disparities on COVID-19 related outcomes. Specifically, Black/African-American and Hispanic patients have been shown to have significantly higher test positivity rates^36–39^ and severity of disease at the time of testing^39^. Furthermore, Black/African American^40^ and Hispanic patients were found to have a higher incidence of COVID-19 infection^37, 41^ and odds of COVID-19 related hospital admission even after adjustment for comorbidities^42^, residence in a low-income area^38^, and insurance plan^40, 43, 44^. The greater COVID-19 burden in these groups is likely due to a combination of systemic health inequities as well as a disproportionate representation among essential workers^45, 46^, which could potentially increase their exposure risk to SARS-CoV-2. In addition, there are known variations in the prevalence of osteoporosis between different racial groups, which could potentially result in disproportionate frequencies of BP prescriptions^47^. The potential confounding due to socio-economic status and differential prevalence of osteoporosis among racial/ethnic groups was addressed in our analysis of the *“Osteo-Dx-Rx”* cohort where we compared BP users to non-users after restricting to female patients with a diagnosis of osteoporosis, all using anti-resorptive bone medications, and matched by insurance type (proportion of Medicaid and dual Medicare/Medicaid users) as a proxy for social-economic status (Figure 4B). Nevertheless, this strategy cannot rigorously rule out a potential under-representation of groups with higher sociodemographic risk factors among BP users that could have contributed to the observed decreased odds of COVID-19 related outcomes in our primary analyses.

The potential bias introduced by a putative differential racial/ethnic group composition of BP users *versus* BP non-users is at least partially addressed by a recent study of a large Californian cohort of female BP users^48^. Compared to the racial composition of California at-large (a proxy for BP non-users)^49^, BP users were predominantly Non-Hispanic White (36.5% in California *versus* 53.3% among BP users). The proportions of Black/African-Americans and Asians among BP users in that study were similar to those in California at-large, whereas Hispanic patients represented a smaller percentage (24%) of BP users as compared to Hispanics in the state’s general population (39.4%). Based on these findings and the reported differential case rates of COVID-19 infections among racial groups in California^50^, we can estimate the race-adjusted incidence of COVID-19 in populations reflecting the composition of BP users and non-users^48^ to be 1.7% and 2.1%, respectively. By comparison, in our study the actual rate of COVID-19 diagnosis in the Western US was 2.5% for BP non-users *versus* 0.46% for BP users (Fig. 2), indicating that the uneven representation of ethnic/racial groups cannot fully explain the observed differences in COVID-19 related outcomes. Moreover, we note that racial/ethnic minorities are also under-represented among statin users^51^, but statin-users in our primary cohort had similar odds of COVID-19 hospitalization as statin non-users (Figure 6B). Similarly, Black/African-Americans and Hispanics have lower utilization rates of antidepressants^52^ and Hispanics were also reported to be undertreated with antihypertensive medications^53^. Our analysis of COVID-19-related outcomes among users and non-users of antihypertensives showed a modest decrease in COVID-19 diagnosis and minimal association with COVID-19-related hospitalization (Figure 6C). By contrast, users of antidepressants had uniformly lower odds for both endpoints (Figures 6E), which is consistent with other recent studies^33, 54, 55^. However, regardless of the class of non-BP preventive drugs analysed, concomitant BP use was consistently associated with dramatically decreased odds of COVID-19 diagnosis and hospitalization as well as testing for SARS-CoV-2 (Figure 6B-E).

Furthermore, specifically looking at the rate of SARS-CoV-2 testing in California^37, 38^ or nation-wide^36^, the proportions of different racial and ethnic groups among tested patients were nearly identical to estimates for the state or national population. Thus, the observed association between BP use and reduced testing for SARS-CoV-2 infection in our nation-wide cohorts is unlikely to be explained by potential differences in racial composition between BP users and non-users. It also seems unlikely that exposure to BPs reduces the actual incidence of SARS-CoV-2 infections. More likely, we propose that immune-modulatory effects of BPs may enhance the anti-viral response of BP users to SARS-CoV-2 and mitigate the development of symptoms. Milder or absent symptoms may have caused infected BP users to be less likely to seek testing. Moreover, because there was a nationwide shortage of available tests for SARS-CoV-2 during the observation period, patients needed to present with sufficiently severe disease symptoms to be eligible for testing, so fewer test-seeking BP users may have qualified. Consequently, a larger proportion of uncaptured ‘silent’ infections among BP users could explain why fewer diagnoses and hospitalizations were observed in this group.

The scarceness of COVID-19 tests combined with the strain on healthcare systems during the observation period could potentially have resulted in a misclassification bias whereby some patients may have been falsely diagnosed and/or hospitalized with COVID-19 without having received a confirmatory test. However, this bias should equally affect BP users and BP non-users and bias our findings towards the null. Relatedly, limited hospital capacity during the observation period could have led to rationing of inpatient hospital beds based on severity of disease and likelihood to survive ^56^. However, matching by age and comorbidities should produce patient populations with similar characteristics used for rationing.

A further limitation of our study is the lack of information on the result of COVID-19 tests received by patients. Therefore, as discussed above, the incidence and odds of COVID-19 testing should not be viewed as a proxy for the rate of infection, but rather reflects the incidence of patients with severe enough symptoms or exposure to warrant testing. Another potential source of confounding is the possibility that some patients in our study were classified as BP non-users due to the absence of BP exposure during the pre-observation period but may have received a BP during the observation period. The potential misclassification of BP non-users, however, would bias towards the null hypothesis, and was only seen in 1.92% of the matched BP non-user population.

An additional limitation is potential censoring of patients who died during the observation period, resulting in truncated insurance eligibility and exclusion based on the continuous insurance eligibility requirement. However, modelling the impact of censoring by using death rates observed in BP users and non-users in the first six months of 2020 and attributing all deaths as COVID-19-related did not significantly alter the decreased odds of COVID-19 diagnosis in BP users (see Appendix 3).

Additional limitations could also arise from comorbidities and misclassification bias due to the diagnostic and procedure codes used to identify study outcomes. Regarding comorbidities, one potential limitation in both the primary and “*Bone-Rx*” cohorts is imbalanced comorbidity burden in BP user and non-user cohorts post-match. Table 1 shows there is differential prevalence of most co-morbid diseases despite matched cumulative CCI score between BP user and BP non-user cohorts. However, this limitation is in part addressed given (1) these covariates were controlled for during regression analyses on study outcomes, and (2) that the key study findings were also observed in the “*Osteo-Dx-Rx*” cohort, which matched based on individual comorbidities. Another potential limitation, misclassification bias of study outcomes, may be present due to the specific procedure/diagnostic codes used. For SARS-CoV-2 testing, procedure codes were limited to those testing for active infection, and therefore observations could be missed if they were captured via antibody testing (CPT 86318, 86328). These codes were excluded a priori due to the focus on the symptomatic COVID-19 population. Furthermore, for the COVID-19 diagnosis and hospitalization outcomes, all events were identified using the ICD-10 code for lab-confirmed COVID-19 (U07.1), and therefore events with an associated diagnosis code for suspected COVID-19 (U07.2) were not included. This was done to have a more stringent algorithm when identifying COVID-19-related events, and any impact of events identified using U07.2 is considered minimal, as previous studies of the early COVID-19 outbreak have found that U07.1 alone has a positive predictive value of 94%^57^, and for this study U07.1 captured 99.2%, 99.0%, and 97.5% of all COVID-19 patient-diagnoses for the primary, “*Bone-Rx*”, and “*Osteo-Dx-Rx*” cohorts, respectively.

One large potential bias to consider when comparing BP users to BP non-users is the healthy adherer effect, whereby adherence to drug therapy is associated with overall healthier behavior^58, 59^. During the COVID-19 pandemic, this could have potentially resulted in differences between BP users and non-users such as, for example, adherence to mask-wearing, hand washing, or social distancing. However, if this effect accounted for the observed association between BP use and COVID-19-related outcomes, one would expect that BP users would have distinct odds for outcomes not predicted to be modulated by BPs and/or that users of other preventive medications would show similar associations. The sensitivity analysis of exploratory outcomes in our primary cohort indicates that BP users had decreased odds of any ER/inpatient visit for acute pancreatitis or acute cholecystitis during the observation period and decreased odds of ER/inpatient visits for acute pancreatitis prior to the COVID-19 pandemic in 2019 (Figure 5A). Plausible (non-exclusive) explanations for BP use association with decreased odds of these exploratory outcomes could be that: (1) BP users may be healthier at baseline; (2) BP users may have fewer risk factors for cholecystitis and pancreatitis such as smoking^60^, which can modify COVID-19-related outcomes^61^ but was not captured in our propensity score matching; (3) BPs directly decreased the risk of hypercalcemia-induced cholecystitis^62^ and pancreatitis^63^; and/or (4) BP users may have preferentially avoided or postponed use of medical services during the pandemic.

Thus, the healthy adherer effect could have potentially contributed to the association of BP use with a lower incidence of exploratory outcomes. However, as discussed above, other preventive drug classes had a variable directional impact on the odds of COVID-19-related events, and sub-analyses within each drug class identified a strong association between concomitant BP use and decreased COVID-19-related events (Figures 6B-E). Taken together, these results suggest the observed association between BP use and COVID-19-related outcomes cannot solely be attributed to general behaviors associated with the healthy adherer effect.

Notably, several observational studies have reported that the use of one of our comparator preventive drug classes, statins, is associated with a lower risk of mortality in hospitalized COVID-19 patients^33 64, 65^. Indeed, statins are currently being tested as an adjunct therapy for COVID-19 (NCT04380402). In our study population, statin use was associated with moderately decreased odds of SARS-CoV-2 testing and COVID-19 diagnosis, though at a much smaller magnitude than BPs, and was not consistently associated with reduced odds of COVID-19-related hospitalizations. Our analysis did not address the clinical course of hospitalized patients, so these results are not necessarily conflicting. However, we note that in our primary cohort, as many as 15.2% of statin users concomitantly used a BP. Indeed, within statin users, stratification by BP use revealed that the decreased odds of SARS-CoV-2 testing, COVID-19 diagnosis, and COVID-19-related hospitalizations remained regardless of statin use. Future studies on disease outcomes of hospitalized COVID-19 patients with antecedent use of BPs and statins alone or in combination are needed to clarify the effects of each drug class.

The differential association of amino-BPs *versus* statins with COVID-19 related outcomes is somewhat unexpected because both target the same biochemical pathway, albeit at different enzymatic steps^13^. Statins block HMG-CoA reductase, the first and key rate-limiting enzyme in the mevalonate pathway^66^. Amino-BPs, which account for >99% of BPs prescribed in our study, inhibit a downstream enzyme in the same metabolic pathway, farnesyl pyrophosphate synthase (FPPS), which converts geranyl pyrophosphate to farnesyl pyrophosphate^67^. FPPS blockade disrupts protein prenylation and interferes with cytoskeletal rearrangement, membrane ruffling and vesicular trafficking in osteoclasts, thus preventing bone resorption ^68^. However, the anti-osteolytic activity of BPs *per se* is unlikely to account for the observed association between BP use and decreased incidence of COVID-19 and, more broadly, respiratory tract infections, because patients treated with non-BP anti-resorptive bone health medications have higher odds of respiratory infections (^16^ and this study).

Another consequence of mevalonate pathway inhibition by both statins and amino-BPs is arrested endosomal maturation in antigen-presenting cells resulting in enhanced antigen presentation, T cell activation and humoral immunity^13^. In addition to this adjuvant-like effect, FPPS blockade by amino-BPs causes the intracellular accumulation of the enzyme’s substrate, isopentyl diphosphate (IPP), in myeloid leukocytes, which then stimulate Vγ9Vδ2 T cells^69, 70^, a large population of migratory innate lymphocytes in humans that are thought to play an important role in host defense against infectious pathogens^71^, including SARS-CoV-1 (ref. ^6^). Experiments in humanized mice that were challenged with influenza viruses have shown that amino-BP-induced expansion of Vγ9Vδ2 T cells markedly improves viral control and mitigates disease severity and mortality^8, 72^. However, since statins act upstream of FPPS, they are expected to inhibit IPP synthesis and, hence, have been shown to counteract the stimulatory effect of amino-BPs on Vγ9Vδ2 T cells^69^. However, statins and amino-BPs do not always antagonize each other. *In vitro*, concomitant statin and amino-BP use has been shown to be synergistic in inhibition of cancer cell growth, but mainly through downstream inhibition of geranylgeranyl transferases and subsequent protein prenylation by statins^73^. The fact that the observed reduction in COVID-19-related outcomes in BP users was not altered by concomitant statin use implies that the apparent protective effects of amino-BPs may not rely solely on stimulation of Vγ9Vδ2 T cells. Indeed, in mice (in which BPs are not known to stimulate γδ T cells), BPs potently boost systemic and mucosal antiviral antibody and T cell responses^14^. This effect was also seen with non-nitrogenous BPs, which do not antagonize FPPS ^14^. In the present study, the number of patients who used non-nitrogenous BPs was less than 20, and therefore too small to determine any impact on COVID-19-related outcomes. Nevertheless, in aggregate, these clinical and pre-clinical findings raise the possibility that BPs may exert (at least some) immuno-stimulatory effects by engaging an as yet unidentified additional pathway, regardless of their nitrogen content.

Irrespective of the precise molecular mechanism of action, BPs have been reported to exert a plethora of effects on additional immune cell populations in humans, including NK cells^74^ and regulatory T cells^75^. Moreover, studies of patients treated with amino-BPs found impaired chemotaxis and generation of reactive oxygen species by neutrophils^76, 77^, a population of inflammatory cells whose dysregulated recruitment and activation are strongly implicated in the pathogenesis of severe COVID-19 (refs. ^78, 79^). Thus, BPs may provide therapeutic benefits during infections with SARS-CoV-2 through modulation of both innate and adaptive immune responses. However, further studies to directly test these pleiotropic immuno-modulatory effects of BPs and to assess their relative contribution to the host response to SARS-CoV-2 infection are needed.

We conclude that, despite a number of caveats discussed above, the association between BP use and decreased odds of COVID-19-related endpoints was robust in analyses comparing BP users to BP non-users. Large differences were detected regardless of age, sex or geographic location that remained robust when using multiple approaches to address unmeasured confounding and/or potential sources of bias. These retrospective findings strongly suggest that BPs should be considered for prophylactic and/or therapeutic use in individuals at risk of SARS-CoV-2 infection. However, additional well-controlled prospective clinical studies will be needed to rigorously assess whether the observed reduction in COVID-19-related outcomes is directly caused by BPs and remains true in patient populations not commonly prescribed BPs.

A number of BPs are globally available as relatively affordable generics that are generally well tolerated and could be prescribed for off-label use. Rare, but severe adverse events that have been linked to BP use include osteonecrosis of the jaw^80^ and atypical femur fractures^81^, which are both associated with long-term BP therapy. In this context, it is important to consider the relationship between the timing of BP exposure and COVID-19-related outcomes. Remarkably, BP users of alendronic acid whose prescription ended more than eight months prior to the observation period, as well as users who initiated alendronic acid therapy immediately preceding the observation period, had similarly decreased odds of COVID-19-related outcomes (Figure 3B). A likely explanation for the observed long-term protection after transient BP use may be the well-documented retention of BPs in bone resulting in half-lives of several years^20^. Small amounts of stored BPs are continuously released, especially in regions of high bone turnover, which may result in persistent exposure of immune cells either systemically or preferentially in bone marrow, a site of active immune cell trafficking^82, 83^ where anti-viral immune responses can be initiated in response to respiratory infection^84^. Thus, BP use at the time of infection may not be necessary for protection against COVID-19. Rather, our results suggest that prophylactic BP therapy may be sufficient to achieve a potentially rapid and sustained immune modulation resulting in profound mitigation of the incidence and/or severity of infections by SARS-CoV-2.

## Acknowledgements

The authors acknowledge Ziqi Chen, Paris Pallis, and Flora Tierney for helpful discussions on the interpretation of study results.

We are grateful to Komodo Health who provided all data used in this analysis at no cost, and we thank Vicki Guan and Ben Cohen from Komodo Health for facilitating this research.

Special thanks to Kantar Health (now Cerner Enviza) who provided the support needed to complete this study with no associated financial requirements.

This study was supported by NIH grants AR068383 and AI155865 (to U.H.v.A.) and a CRI Irvington postdoctoral fellowship CRI2453 (to P.H.).

## APPENDIX

## APPENDIX 1: Study Methods

### Section 1: Variable Assignment

#### Outcomes

The following details the identification algorithms and associated codes that were used to identify outcomes of interest, including COVID-19-related as well as the exploratory outcomes that were assessed during sensitivity analyses.

##### Primary Outcomes

<u>SARS-CoV-2 Testing</u>

- Any medical services claim with a procedure code indicating polymerase chain reaction (PCR) testing for active SARS-CoV-2 infection 3/1/2020-6/30/2020
- Identified using HCPCS codes: 87635, 87636, 87637

<u>COVID-19 Diagnosis</u>

- Any medical services claim with a diagnosis code indicating COVID-19 3/1/2020-6/30/2020
- Identified using ICD-10 code U07.1x

<u>COVID-19-Related Hospitalization</u>

- Any medical services claim occurring in an inpatient setting with a diagnosis code indicating COVID-19 3/1/2020-6/30/2020
- Identified using ICD-10 code U07.1x

##### **Exploratory Outcomes** (study observation period)

<u>Acute Cholecystitis-Related Service</u>

- Any medical services claim occurring in an emergency room/inpatient setting with a diagnosis indicating acute cholecystitis 3/1/2020-6/30/2020
- Identified using ICD-10 codes K81.0x

<u>Acute Pancreatitis-Related Service</u>

- Any medical services claim occurring in an emergency room/inpatient setting with a diagnosis indicating acute pancreatitis 3/1/2020-6/30/2020
- Identified using ICD-10 codes K85.x

##### Exploratory Outcomes (2019)

<u>Acute Cholecystitis-Related Service</u>

- Any medical services claim occurring in an emergency room/inpatient setting with a diagnosis indicating acute cholecystitis 7/1/2019-12/31/2019
- Identified using ICD-10 codes K81.0x

<u>Acute Pancreatitis-Related Service</u>

- Any medical services claim occurring in an emergency room/inpatient setting with a diagnosis indicating acute pancreatitis 7/1/2019-12/31/2019
- Identified using ICD-10 codes K85.x

<u>Acute Bronchitis-Related Service</u>

- Any medical services claim with a diagnosis indicating acute bronchitis 7/1/2019-12/31/2019
- Identified using ICD-10 codes J20.x-J21.x

<u>Acute Pneumonia-Related Service</u>

- Any medical services claim with a diagnosis indicating acute bronchitis 7/1/2019-12/31/2019
- Identified using ICD-10 codes J13.x-J18.x

##### Osteonecrosis

<u>Osteonecrosis</u>

- Any medical services claim with a diagnosis indicating drug-induced osteonecrosis 1/1/2019-6/30/2020
- Identified using ICD-10 codes M87.1x

#### Drug-Exposure Assignment

The following details the identification algorithms and associated inputs used for drug-exposure classification of study subjects into users/non-users of bisphosphonates, non-bisphosphonates osteoporosis medications, statins, antihypertensives, non-insulin antidiabetics, and antidepressants.

<u>Bisphosphonates</u>

- Any outpatient prescription or in-office dispensing 1/1/2019-2/29/2020
- Drugs included: alendronate, alendronic acid, etidronate, ibandronate, ibandronic acid, pamidronate, risedronate, and zoledronic acid

<u>Non-BP Anti-Resorptive Bone Health Medications</u>

- Any outpatient prescription or in-office dispensing 1/1/2019-2/29/2020
- Drugs included: denosumab, calcitonin, raloxifene, romosozumab-aqqg, teriparatide, abaloparatide, or bazedoxifene

<u>Statins</u>

- Any outpatient prescription 1/1/2019-2/29/2020
- Drugs included: pravastatin, rosuvastatin, fluvastatin, atorvastatin, pitavastatin, or simvastatin

<u>Antihypertensives</u>

- Any non-ophthalmic, non-injection, outpatient prescription claim for a beta-blocker, calcium channel blocker, or renin angiotensin system antagonist 1/1/2019-2/29/2020
- Drugs included: acebutolol, atenolol, betaxolol, bisoprolol, carvedilol, labetalol, metoprolol, nadolol, nebivolol, penbutolol, pindolol, propranolol, timolol, amlodipine, diltiazem, felodipine, isradipine, nicardipine, nifedipine, nisoldipine, verapamil, aliskiren, azilsartan, benazepril, candesartan, captopril, enalapril, eprosartan, fosinopril, irbesartan, lisinopril, losartan, moexipril, olmesartan, perindopril, quinapril, ramipril, sacubitril, telmisartan, trandolapril, valsartan

<u>Antidiabetics</u>

- Any outpatient prescription claim for a non-insulin antidiabetic medication 1/1/2019-2/29/2020
- Drugs included: metformin, chlorpropamide, glimepiride, glipizide, glyburide, tolazamide, tolbutamide, pioglitazone, rosiglitazone, alogliptin, linagliptin, saxagliptin, sitagliptin, albiglutide, dulaglutide, exenatide, liraglutide, lixisenatide, semaglutide, nateglinide, repaglinide, canagliflozin, dapagliflozin, empagliflozin, ertugliflozin

<u>Antidepressants</u>

- Any outpatient prescription claim for a selective serotonin reuptake inhibitor, norepinephrine-dopamine reuptake inhibitor, serotonin-norepinephrine reuptake inhibitor, tricyclic, tetracyclic, modified cyclic, or MAO inhibitor medication 1/1/2019-2/29/2020
- Drugs included: amoxapine, bupropion, citalopram, clomipramine, desipramine, desvenlafaxine, doxepin, duloxetine, escitalopram, esketamine, fluoxetine, fluvoxamine, imipramine, isocarboxazid, levomilnacipran, maprotiline, mirtazapine, nefazodone, nortriptyline, paroxetine, phenelzine, protriptyline, selegiline, sertraline, tranylcypromine, trazodone, trimipramine, venlafaxine, vilazodone, vortioxetine

#### Charlson Comorbidity Condition Assignment

The following ICD-10 codes were used to assign the CCI condition-specific indicators that are used to calculate the overall CCI score. The time period used for identification of condition-specific indicators was the entire pre-observation period (1/1/2019-2/29/2020).

<u>Myocardial infarction</u>

- ICD-10 codes: I21.x, I22.x, I25.2

<u>Congestive heart failure</u>

- ICD-10 codes: I09.9, I11.0, I13.0, I13.2, I25.5, I42.0, I42.5 - I42.9, I43.x, I50.x, P29.0

<u>Peripheral vascular disease</u>

- ICD-10 codes: I70.x, I71.x, I73.8, I73.9, I77.1, I79.0, I79.2, K55.1, K55.8, K55.9, Z95.8, Z95.9

<u>Cerebrovascular disease</u>

- ICD-10 codes: G45.x, G46.x, H34.0, I60.x-I69.x

<u>Dementia</u>

- ICD-10 codes: F00.x - F03.x, F05.1, G30.x, G31.1

<u>Chronic pulmonary disease</u>

- ICD-10 codes: I27.8, I27.9, J40.x - J47.x, J60.x - J67.x, J68.4, J70.1, J70.3

<u>Rheumatologic disease</u>

- ICD-10 codes: M05.x, M06.x, M31.5, M32.x - M34.x, M35.1, M35.3, M36.0

<u>Peptic ulcer disease</u>

- ICD-10 codes: K25.x-K28.x

<u>Mild liver disease</u>

- ICD-10 codes: B18.x, K70.0 - K70.3, K70.9, K71.3 - K71.5, K71.7, K73.x, K74.x, K76.0, K76.2 - K76.4, K76.8, K76.9, Z94.4

<u>Diabetes without chronic complications</u>

- ICD-10 codes: E10.0, E10.1, E10.6, E10.8, E10.9, E11.0, E11.1, E11.6, E11.8, E11.9, E12.0, E12.1, E12.6, E12.8, E12.9, E13.0, E13.1, E13.6, E13.8, E13.9, E14.0, E14.1, E14.6, E14.8, E14.9

<u>Diabetes with chronic complications</u>

- ICD-10 codes: E10.2 - E10.5, E10.7, E11.2 - E11.5, E11.7, E12.2 - E12.5, E12.7, E13.2 - E13.5, E13.7, E14.2 - E14.5, E14.7

<u>Hemiplegia or paraplegia</u>

- ICD-10 codes: G04.1, G11.4, G80.1, G80.2, G81.x, G82.x, G83.0 - G83.4, G83.9

<u>Renal disease</u>

- ICD-10 codes: I12.0, I13.1, N03.2 - N03.7, N05.2 - N05.7, N18.x, N19.x, N25.0, Z49.0 - Z49.2, Z94.0, Z99.2

<u>Any tumor, leukemia, or lymphoma</u>

- ICD-10 codes: C00.x - C26.x, C30.x - C34.x, C37.x - C41.x, C43.x, C45.x - C58.x, C60.x - C76.x, C81.x - C85.x, C88.x, C90.x - C97.x

<u>Moderate or severe liver disease</u>

- ICD-10 codes: I85.0, I85.9, I86.4, I98.2, K70.4, K71.1, K72.1, K72.9, K76.5, K76.6, K76.7

<u>Metastatic solid tumor</u>

- ICD-10 codes: C77.x - C80.x

<u>AIDS/HIV</u>

- ICD-10 codes: B20.x - B22.x, B24.x

#### Additional Condition Covariate Assignment

The following details the ICD-10 diagnosis codes that were used to identify comorbid conditions. For all condition indicators classification was based on all medical claims occurring during the pre-observation period (1/1/2019-2/29/2020).

<u>Osteoporosis:</u> M80.x, M81.x, M82.x

<u>Cardiovascular Disease Overall:</u> I3x.x-I4x.x, I20.x-I28.x, I50.x-I52.x Cancer: C0x.x - C9x.x

<u>Chronic Kidney Disease (CKD)/ End-Stage Renal Disease (ESRD):</u> I12.0, I13.1, N03.2 - N03.7, N05.2 - N05.7, N18.x, N19.x, N25.0, Z49.0 - Z49.2, Z94.0, Z99.2

<u>Chronic Obstructive Pulmonary Disease (COPD):</u> J43.x, J44.x Dementia: F00.x - F03.x, F05.1, G30.x, G31.1

<u>Depression:</u> F32.x, F33.x Dyslipidemia: E78.x

<u>Heart Failure:</u> I50.x, I11.0xx, I13.0xx, I13.2xx HIV/AIDS: B20.x - B22.x, B24.x

<u>Hypertension:</u> I10.x, I12.x, I11.9xx, I13.1xx

<u>Liver Disease:</u> B18.x, K70.0 - K70.3, K70.9, K71.3 - K71.5, K71.7, K73.x, K74.x, K76.0, K76.2 - K76.4, K76.8, K76.9, Z94.4, I85.0, I85.9, I86.4, I98.2, K70.4, K71.1, K72.1, K72.9, K76.5, K76.6, K76.7

<u>Obesity:</u> E66.x

<u>Sickle Cell Disease:</u> D57.x Stroke: I63.x

<u>Type 2 Diabetes:</u> E11.x

### Section 2: Sensitivity Analyses Methodologies

#### Sensitivity Analysis (1): COVID-19-Related Outcomes in “*Bone-Rx*” Cohort

##### Overview & Rationale

- The first sensitivity analysis was performed to validate the robustness of the primary findings by limiting all BP non-users to those who had used non-BP anti-resorptive bone health medications during the pre-observation period, thus yielding a more comparable comparator cohort that was also receiving bone health medication therapy.
- The use of an active-comparator cohort was done to reduce the impact of unmeasured confounding that may have occurred in the primary analysis due to the use of the derived Charlson Comorbidity Index composite score as the only comorbidity matching covariate. Restriction of the patient population to users of any non-BP anti-resorptive bone health medication prior to propensity-score matching improves the probability of having drug user/non-user matches with more similar clinical characteristics.
- This sensitivity analysis, further, also acted to increase the robustness and reliability of the matched user/non-user outcome comparisons since non-BP anti-resorptive bone health medication users represented the smaller portion of the total bone health medication-user population (“*Bone-Rx*” cohort) and therefore were matched to their best BP-user pair.

##### Analysis Cohort Definition(s)

- Continuous medical and prescription insurance coverage 1/1/2019-6/30/2020
- Patients with ≥1 claim for any anti-resorptive bone health medication 1/1/2019-2/29/2020

##### Exposures of Interest

- Patients were assigned into the BP user cohort if they had any claim 1/1/2019-2/29/2020 for one of the following: alendronate, alendronic acid, etidronate, ibandronate, ibandronic acid, pamidronate, risedronate, and zoledronic acid.
- Patients were assigned into the non-BP any anti-resorptive bone health medication user cohort if: (1) they had any claim 1/1/2019-2/29/2020 for one of the following: denosumab, calcitonin, raloxifene, romosozumab-aqqg, teriparatide, abaloparatide, or bazedoxifene; and (2) they had no BP claims 1/1/2019-2/29/2020.

##### Outcomes

- SARS-CoV-2 testing, COVID-19 diagnosis, and COVID-19-related hospitalizations

##### Cohort Matching

- Non-BP anti-resorptive bone health medication users were matched to BP users based on age, gender, insurance type, any PCP visit in 2019, and comorbidity score. Matching was performed within each region separately (northeast, midwest, south, west) and then combined as well as in NY-state alone.

##### Statistical Analyses

- Same as was performed for the primary analysis cohort.

#### Sensitivity Analysis (2): COVID-19-Related Outcomes in “*Osteo-Dx-Rx*” Cohort

##### Overview & Rationale

- The second sensitivity analysis was performed to further assess the robustness of the primary analysis findings by performing a highly restricted comparator cohort matching that included patients diagnosed and treated for osteoporosis (“*Osteo-Dx-Rx*” cohort).
- The relationship between COVID-19-related outcomes and BP-exposure was modelled after restricting anti-resorptive bone health medication users to those most likely to use BPs and matching BP non-users to BP users based on the presence of comorbid diagnoses within insurance type in four states with early COVID-19 spread representing each to further reduce confounding related to differences in demographic/clinical characteristics amongst BP users/non-users, confounding due to socioeconomic status (insurance type as proxy), and confounding due to differences in COVID-19-exposure risk based on geography.

##### Analysis Cohort Definition(s)

- Continuous medical and prescription insurance coverage 1/1/2019-6/30/2020
- Patients with ≥1 claim for any osteoporosis medication 1/1/2019-2/29/2020 who also met the following criteria: (i) female; (ii) age 51 or older; (iii) identified as residing in New York, Illinois, Florida, or California; and (iv) had ≥1 medical claim indicating a diagnosis of osteoporosis 1/1/2019-2/29/2020

##### Exposures of Interest

- Patients were assigned into the BP user cohort if they had any claim 1/1/2019-2/29/2020 for one of the following: alendronate, alendronic acid, etidronate, ibandronate, ibandronic acid, pamidronate, risedronate, and zoledronic acid.
- Patients were assigned into the non-BP anti-resorptive bone health medication user cohort if: (1) they had any claim 1/1/2019-2/29/2020 for one of the following: denosumab, calcitonin, raloxifene, romosozumab-aqqg, teriparatide, abaloparatide, or bazedoxifene; and (2) they had no BP claims 1/1/2019-2/29/2020.

##### Outcomes

- SARS-CoV-2 testing, COVID-19 diagnosis, and COVID-19-related hospitalizations

##### Cohort Matching

- Non-anti-resorptive bone health medication users were matched to BP users based on age, PCP visit in 2019, and the presence of the following comorbid conditions (assigned using ICD-10 codes on claims occurring 1/1/2019-2/29/2020): cancer, chronic obstructive pulmonary disease, depression, dyslipidaemia, heart failure, hypertension, obesity, and type 2 diabetes.
- Matching was performed within each state when stratified by insurance type (commercial, dual, Medicaid, Medicare).

##### Statistical Analyses

- Multivariate logistic regression analyses, modelled separately for each COVID-19-related outcome of interest, were performed on the unmatched and matched samples after combining all patient observations. In addition to the key exposure variable (indicating BP user versus non-BP user), the regression model also included demographic/clinical covariate for age group, region, insurance type, PCP visit in 2019, and the following comorbid conditions: osteoporosis, cancer, chronic obstructive pulmonary disease, depression, dyslipidaemia, hypertension, obesity, type 2 diabetes, cardiovascular disease overall, sickle cell anemia, stroke, dementia, HIV/AIDS, chronic kidney disease/end-stage renal disease, and liver disease.

#### Sensitivity Analysis (3): Association of BP-use with Exploratory Negative Control Outcomes

##### Overview & Rationale

- The third sensitivity analysis was performed to assess the relationship between BP-use and outcomes not anticipated to be impacted by the pharmacological mechanism of BPs.
- This was performed by modelling the relationship between BP-exposure and other outcomes occurring (1) during the study observation, and (2) during the second half of 2019 among BP users with claims during the first half of 2019 and their previously-assigned BP non-user matched pair, in the primary, “*Bone-Rx*”, and “*Osteo-Dx-Rx*” cohorts.
- Outcomes modelled included any acute cholecystitis-related or acute pancreatitis-related inpatient/emergency-room (ER) service, used as exploratory outcomes not predicted to be modulated by BP exposure to assess the validity of the core COVID-19-related outcomes.

##### Analysis Cohort Definition(s)

- Patients who were included in the primary analysis cohort for assessment of (1) outcomes occurring during the study observation period; for (2) outcomes assessed during the second half of 2019 the cohort was restricted to among BP users with claims during the first half of 2019 and their previously-assigned BP non-user matched pair.
- Patients who met all eligibility criteria to be included in the ‘*Bone-Rx’* cohort for assessment of (1) outcomes occurring during the study observation period; for (2) outcomes assessed during the second half of 2019 the cohort was restricted to among BP users with claims during the first half of 2019 and their previously-assigned BP non-user matched pair.
- Patients who met all eligibility criteria to be included in the ‘*Osteo-Dx-Rx’* cohort for assessment of (1) outcomes occurring during the study observation period; for (2) outcomes assessed during the second half of 2019 the cohort was restricted to among BP users with claims during the first half of 2019 and their previously-assigned BP non-user matched pair.

##### Exposures of Interest

- For the primary analysis cohort, the BP user / BP non-user assignment was the same as used in the core analyses.
- For the “*Bone-Rx*” and “*Osteo-Dx-Rx*” cohorts, assignment was the same as used in those analyses stratifying medication users into BP users and non-BP medication users.

##### Outcomes

- Any medical claim from an ER/inpatient setting with a diagnosis indicating acute cholecystitis (ICD-10 code K81.0x) occurring 3/1/2020-6/30/2020 (observation period)
- Any medical claim from an ER/inpatient setting with a diagnosis indicating acute pancreatitis (ICD-10 code K85.x) occurring 3/1/2020-6/30/2020 (observation period)
- Any medical claim from an ER/inpatient setting with a diagnosis indicating acute cholecystitis (ICD-10 code K81.0x) occurring 7/1/2019-12/31/2019 (2019)
- Any medical claim from an ER/inpatient setting with a diagnosis indicating acute pancreatitis (ICD-10 code K85.x) occurring 7/1/2019-12/31/2019 (2019)

##### Cohort Matching

- NA; all cohorts previously matched.

##### Statistical Analyses

- Multivariate logistic regression analyses were performed using the same methodologies employed when assessing COVID-19 outcomes that were cohort-build-specific (i.e. followed previous approach detailed for each respective cohort build) to assess the odds of acute cholecystitis or acute pancreatitis.

#### Sensitivity Analysis (4): Association of BP-use with Exploratory Positive Control Outcomes in 2019

##### Overview & Rationale

- The fourth sensitivity analysis was performed to assess the relationship between BP-use and select outcomes occurring in 2019 to validate the theorized BP mechanism of action.
- This was performed by modelling the relationship between BP-exposure in the first half of 2019 and other outcomes occurring during the second half of 2019 in the primary, “*Bone-Rx*”, and “*Osteo-Dx-Rx*” cohorts, specifically medical services for other infectious respiratory conditions (acute bronchitis, pneumonia), used to assess the validity of the relationship between BP-use and decreased respiratory infections.

##### Analysis Cohort Definition(s)

- The following criteria were applied to all three cohort build variations (primary analysis cohort, “*Bone-Rx*” cohort, “*Osteo-Dx-Rx*” cohort): (i) BP users were restricted to those with any BP claim 1/1/2019-6/30/2019, and the remaining previously-classified BP-user patients with their first BP-claim date occurring on/after 7/1/2019 were excluded; (ii) BP non-users were restricted to their BP-user matched-pair previously assigned.

##### Exposures of Interest

- In all cohort build variations, the previously-classified BP user cohorts were restricted to those with any BP-claim 1/1/2019-6/30/2019; all other previously-classified BP users were excluded.

##### Outcomes

- Any medical claim with a diagnosis indicating acute bronchitis (ICD-10 code J20.x-J21.x) occurring 7/1/2019-12/31/2019
- Any medical claim with a diagnosis indicating pneumonia (ICD-10 code J13.x-J18.x) occurring 7/1/2019-12/31/2019

##### Cohort Matching

- NA; all cohorts previously matched.

##### Statistical Analyses

- Multivariate logistic regression analyses were performed using the same methodologies employed when assessing COVID-19-related outcomes that were cohort-build-specific (i.e. followed previous approach detailed for each respective cohort build) to assess the odds of acute bronchitis, or pneumonia.

#### Sensitivity Analysis (5): Association between use of Other Drug Classes and COVID-19-Related Outcomes

##### Overview & Rationale

- The fifth sensitivity analysis was performed to assess whether the observed protective effect of BPs may be associated with general healthier behaviours in patients using any medication rather than specifically BP use. To assess this unmeasured confounding due to the healthy adherer effect, which is a type of potential bias where patients may have better outcomes due to their heathier behaviours and not better outcomes related to active drug treatment itself, the first sensitivity analysis evaluated the association between use of other preventive medications (statin, antihypertensive, antidiabetic, antidepressant) and COVID-19-related outcomes were evaluated.
- This was performed following the same techniques used in the primary cohort matching and analyses but when assigned drug exposure cohorts based on the use of statin, antihypertensive, antidiabetic, or antidepressant medications. The consistency of methods was done to permit direct comparison on the association between drug-use and COVID-19-related outcomes to assess whether the healthy adherer effect alone accounts for the decrease in the odds of COVID-19 outcomes when comparing BP users to non-users in the primary analysis. Evidence to support the contention that the HAE is a significant source of unmeasured confounding would necessitate that other drug classes display a similar statistically significant trend and/or magnitude when comparing drug users to non-users. Variability in directional impact, magnitude, and/or statistical significance would, conversely, suggest that the healthy adherer effect itself does not account for the differences seen when comparing BP users to BP non-users.
- This sensitivity analysis, additionally, also employed a unique nested-matching technique wherein BP users were matched to BP non-users within the other-medication-class matched populations when stratified into the already matched but mutually exclusive user/non-user cohorts. This was performed to: (1) assess whether the decreased odds of COVID-19-realted outcomes in BP users compared to BP non-users was robust, even amongst cohorts displaying an increase in the odds of COVID-19-related outcomes; and (2) to assess whether the magnitude of decrease in odds of COVID-19-related outcomes amongst BP users compared to BP non-users seen in the primary analysis is impacted by use of other medication classes, including some that have also been identified as being associated with a reduced incidence and/or severity of COVID-19-related outcomes.

##### Analysis Cohort Definition(s)

- Continuous medical and prescription insurance coverage 1/1/2019-6/30/2020 (*all*)
- Patients with any claim for another drug class of interest (statin, antihypertensive, antidiabetic, antidepressant) medication 1/1/2019-2/29/2020 were classified users
- Among the propensity-score matched drug user/non-user cohorts, a further stratification and propensity-score matching based on BP use 1/1/2019-2/29/2020 to yield the following: (i) drug user/BP user matched to drug user/BP non-user, (ii) drug non-user/BP user matched to drug non-user/BP non-user.

##### Exposures of Interest

- Patients were assigned into the statin user cohort if they had any claim 1/1/2019-2/29/2020 for one of the following: pravastatin, rosuvastatin, fluvastatin, atorvastatin, pitavastatin, or simvastatin
- Patients were assigned into the antihypertensive user cohort if they had any non-ophthalmic, non-injection claim 1/1/2019-2/29/2020 for a beta blocker, calcium channel blocker, or renin-angiotensin system antagonist medication.
- Patients were assigned into the antidiabetic user cohort if they had any claim 1/1/2019-2/29/2020 for one of the following non-insulin medications: metformin, chlorpropamide, glimepiride, glipizide, glyburide, tolazamide, tolbutamide, pioglitazone, rosiglitazone, alogliptin, linagliptin, saxagliptin, sitagliptin, albiglutide, dulaglutide, exenatide, liraglutide, lixisenatide, semaglutide, nateglinide, repaglinide, canagliflozin, dapagliflozin, empagliflozin, ertugliflozin
- Patients were assigned into the antidepressant user cohort if they had any claim 1/1/2019-2/29/2020 for one of the following: amoxapine, bupropion, citalopram, clomipramine, desipramine, desvenlafaxine, doxepin, duloxetine, escitalopram, esketamine, fluoxetine, fluvoxamine, imipramine, isocarboxazid, levomilnacipran, maprotiline, mirtazapine, nefazodone, nortriptyline, paroxetine, phenelzine, protriptyline, selegiline, sertraline, tranylcypromine, trazodone, trimipramine, venlafaxine, vilazodone, vortioxetine

##### Outcomes

- SARS-CoV-2 testing, COVID-19 diagnosis, and COVID-19-related hospitalizations

##### Cohort Matching

- For the larger drug-class analyses, matching was performed following the same methods used in the primary analysis: users were matched to non-users based on age, gender, insurance type, any PCP visit in 2019, and comorbidity score. Matching was performed within each region separately (northeast, midwest, south, west) and then combined, as well as in NY-state alone.
- Following this matching procedure, a nested BP user to BP non-user propensity score match was then performed on the aforementioned matched populations (i.e. within the separate and already matched statin user and statin non-user populations). Matching was performed using the same list of demographic/clinical characteristics, and was also performed within each region separately (northeast, midwest, south, west) and then combined as well as in NY-state alone.

##### Statistical Analyses

- Same as was performed for the primary analysis cohort.

## APPENDIX 2: Additional Study Results; Cohort Characteristics Pre/Post Match

### Primary Analysis Study Population

#### Northeast Region

A total of 2,152,560 patients identified as residing in the northeast were included in the unmatched primary analysis cohort comparisons, of which 119,728 (5.6%) and 2,032,832 (94.4%) were classified as BP users and BP non-users, respectively (Table S2a). Prior to propensity-score matching, there were significant differences across all demographic and clinical characteristics. Compared to BP non-users, BP users were older (97.5% age ≥51 versus 49.8%; p<0.001), predominantly female (90.5% versus 57.4%; p<0.001), with higher comorbidity burden (mean CCI=0.93 versus 0.65; p<0.001), insured by Medicare (46.5% versus 18.0%; p<0.001), and have had a primary-care physician (PCP) visit in 2019 (58.3% versus 42.8%; p<0.001). Propensity-score matching yielded 119,494 BP users and 119,494 BP non-users with no significant differences across examined characteristics. A total of 234 BP users from the northeast region in the unmatched primary analysis cohort were not assigned an applicable BP non-user pair during the matching procedure and were excluded from the matched BP user population.

#### Midwest Region

A total of 1,467,802 patients identified as residing in the midwest were included in the unmatched primary analysis cohort comparisons, of which 75,967 (5.2%) and 1,391,835 (94.8%) were classified as BP users and BP non-users, respectively (Table S2b). Prior to propensity-score matching, there were significant differences across all demographic and clinical characteristics. Compared to BP non-users, BP users were older (96.6% age ≥51 versus 44.0%; p<0.001), predominantly female (90.3% versus 57.1%; p<0.001), with higher comorbidity burden (mean CCI=0.99 versus 0.56; p<0.001), insured by Medicare (43.6% versus 14.5%; p<0.001), and have had a primary-care physician (PCP) visit in 2019 (62.2% versus 51.0%; p<0.001). Propensity-score matching yielded 75,901 BP users and 75,901 BP non-users with no significant differences across examined characteristics. A total of 66 BP users from the midwest region in the unmatched primary analysis cohort were not assigned an applicable BP non-user pair during the matching procedure and were excluded from the matched BP user population.

#### South Region

A total of 3,042,604 patients identified as residing in the south were included in the unmatched primary analysis cohort comparisons, of which 160,886 (5.3%) and 2,881,718 (94.7%) were classified as BP users and BP non-users, respectively (Table S2c). Prior to propensity-score matching, there were significant differences across all demographic and clinical characteristics. Compared to BP non-users, BP users were older (96.8% age ≥51 versus 39.2%; p<0.001), predominantly female (90.6% versus 57.4%; p<0.001), with higher comorbidity burden (mean CCI=0.86 versus 0.55; p<0.001), insured by Medicare (41.0% versus 11.3%; p<0.001), and have had a primary-care physician (PCP) visit in 2019 (66.1% versus 49.2%; p<0.001). Propensity-score matching yielded 159,704 BP users and 159,704 BP non-users with no significant differences across examined characteristics. A total of 1,182 BP users from the south region in the unmatched primary analysis cohort were not assigned an applicable BP non-user pair during the matching procedure and were excluded from the matched BP user population.

#### West Region

A total of 1,243,637 patients identified as residing in the west were included in the unmatched primary analysis cohort comparisons, of which 95,470 (7.7%) and 1,148,167 (92.3%) were classified as BP users and BP non-users, respectively (Table S2d). Prior to propensity-score matching, there were significant differences across all demographic and clinical characteristics. Compared to BP non-users, BP users were older (97.8% age ≥51 versus 43.5%; p<0.001), predominantly female (88.7% versus 56.4%; p<0.001), with higher comorbidity burden (mean CCI=1.08 versus 0.66; p<0.001), insured by Medicare (43.5% versus 11.0%; p<0.001), and have had a primary-care physician (PCP) visit in 2019 (67.7% versus 45.3%; p<0.001). Propensity-score matching yielded 95,267 BP users and 95,267 BP non-users with no significant differences across examined characteristics. A total of 203 BP users from the west region in the unmatched primary analysis cohort were not assigned an applicable BP non-user pair during the matching procedure and were excluded from the matched BP user population.

#### New York State

A total of 968,296 patients identified as residing in New York state were included in the unmatched primary analysis NY-state restricted cohort, of which 50,035 (5.2%) and 918,261 (94.8%) were classified as BP users and BP non-users, respectively (Table S2e). Prior to propensity-score matching, there were significant differences across all demographic and clinical characteristics. Compared to BP non-users, BP users were older (98.1% age ≥51 versus 50.7%; p<0.001), predominantly female (90.9% versus 57.5%; p<0.001), with higher comorbidity burden (mean CCI=0.95 versus 0.63; p<0.001), insured by Medicare (57.7% versus 19.5%; p<0.001), and have had a primary-care physician (PCP) visit in 2019 (62.7% versus 45.3%; p<0.001). Propensity-score matching yielded 49,862 BP users and 49,862 BP non-users with no significant differences across examined characteristics. A total of 173 BP users from the unmatched New York state primary analysis cohort were not assigned an applicable BP non-user pair during the matching procedure and were excluded from the matched BP user population.

### Bone-Rx Analysis Study Population

#### All Observations (all regions combined)

A total of 502,895 patients were included in the unmatched “*Bone-Rx*” analysis cohort comparisons, of which 452,051 (89.9%) and 50,844 (10.1%) were classified as BP users and BP non-users, respectively (Table S6a). Prior to propensity-score matching, there were significant differences across all demographic and clinical characteristics. Compared to BP non-users, BP users were younger (47.9% age ≥71 versus 55.2%; p<0.001), predominantly female (90.1% versus 87.2%; p<0.001), with a lower comorbidity burden (mean CCI=0.95 versus 1.99; p<0.001), with a larger proportion of patients residing in the west (21.1% versus 15.8%; p<0.001), a lower proportion covered by Medicare (43.4% versus 47.5%; p<0.001), and a lower proportion have had a primary-care physician (PCP) visit in 2019 (63.8% versus 64.3%; p=0.009). Propensity-score matching yielded 50,498 BP users and 50,498 BP non-users with no significant differences across examined characteristics. A total of 346 BP non-users from the unmatched “*Bone-Rx*” analysis cohort were not assigned an applicable BP user pair during the matching procedure and were excluded from the matched BP non-user population.

#### Northeast Region

A total of 135,867 patients identified as residing in the northeast were included in the unmatched “*Bone-Rx*” analysis cohort comparisons, of which 119,728 (88.1%) and 16,139 (11.9%) were classified as BP users and BP non-users, respectively (Table S6b). Prior to propensity-score matching based on BP-use, there were significant differences across all demographic and clinical characteristics except for any PCP visit in 2019 (p=0.95). Compared to BP non-users, BP users were younger (48.1% age ≥71 versus 54.8%; p<0.001), predominantly female (90.5% versus 87.5%; p<0.001), with a lower comorbidity burden (mean CCI=0.93 versus 1.97; p<0.001), and a lower proportion insured by Medicare (46.5% versus 54.0%; p<0.001). Propensity-score matching yielded 15,993 BP users and 15,993 BP non-users with no significant differences across examined characteristics. A total of 146 BP non-users from the northeast region in the unmatched “*Bone-Rx*” analysis cohort were not assigned an applicable BP user pair during the matching procedure and were excluded from the matched BP non-user population.

#### Midwest Region

A total of 85,391 patients identified as residing in the midwest were included in the unmatched “*Bone-Rx*” analysis cohort comparisons, of which 75,967 (89.0%) and 9,424 (11.0%) were classified as BP users and BP non-users, respectively (Table S6c). Prior to propensity-score matching, there were significant differences across all demographic and clinical characteristics. Compared to BP non-users, BP users were younger (43.0% age ≥71 versus 54.1%; p<0.001), predominantly female (90.3% versus 86.1%; p<0.001), with a lower comorbidity burden (mean CCI=0.99 versus 2.12; p<0.001), had a lower proportion insured by Medicare (43.6% versus 51.9%; p<0.001), with a lower proportion having a primary-care physician (PCP) visit in 2019 (62.2% versus 64.7%; p<0.001). Propensity-score matching yielded 9,360 BP users and 9,360 BP non-users with no significant differences across examined characteristics. A total of 64 BP non-users from the midwest region in the unmatched “*Bone-Rx*” analysis cohort were not assigned an applicable BP user pair during the matching procedure and were excluded from the matched BP non-user population.

#### South Region

A total of 178,118 patients identified as residing in the south were included in the unmatched “*Bone-Rx*” analysis cohort comparisons, of which 160,886 (90.3%) and 17,232 (9.7%) were classified as BP users and BP non-users, respectively (Table S6d). Prior to propensity-score matching, there were significant differences across all demographic and clinical characteristics except for any PCP visit in 2019 (p=0.45). Compared to BP non-users, BP users were younger (46.6% age ≥71 versus 53.3%; p<0.001), predominantly female (90.6% versus 88.1%; p<0.001), with a lower comorbidity burden (mean CCI=0.86 versus 1.86; p<0.001), and a lower proportion insured by Medicare (41.0% versus 44.0%; p<0.001). Propensity-score matching yielded 17,140 BP users and 17,140 BP non-users with no significant differences across examined characteristics. A total of 92 BP non-users from the south region in the unmatched “*Bone-Rx*” analysis cohort were not assigned an applicable BP user pair during the matching procedure and were excluded from the matched BP non-user population.

#### West Region

A total of 103,519 patients identified as residing in the west were included in the unmatched “*Bone-Rx*” analysis cohort comparisons, of which 95,470 (92.2%) and 8,049 (7.8%) were classified as BP users and BP non-users, respectively (Table S6e). Prior to propensity-score matching, there were significant differences across all demographic and clinical characteristics. Compared to BP non-users, BP users were younger (54.1% age ≥71 versus 61.6%; p<0.001), predominantly female (88.7% versus 86.2%; p<0.001), with a lower comorbidity burden (mean CCI=1.08 versus 2.17; p<0.001), insured by Medicare (43.5% versus 36.9%; p<0.001), with a lower proportion having a primary-care physician (PCP) visit in 2019 (67.7% versus 71.6%; p<0.001). Propensity-score matching yielded 8,005 BP users and 8,005 BP non-users with no significant differences across examined characteristics. A total of 44 BP non-users from the west region in the unmatched “*Bone-Rx*” analysis cohort were not assigned an applicable BP user pair during the matching procedure and were excluded from the matched BP non-user population.

#### New York State

A total of 57,397 patients identified as residing in New York state were included in the unmatched “*Bone-Rx*” analysis NY-state restricted cohort, of which 50,035 (87.2%) and 7,362 (12.8%) were classified as BP users and BP non-users, respectively (Table S6f). Prior to propensity-score matching, there were significant differences across all demographic and clinical characteristics except for any PCP visit in 2019 (p=0.35). Compared to BP non-users, BP users were younger (53.2% age ≥11 versus 54.5%; p<0.001), predominantly female (90.9% versus 89.5%; p<0.001), with a lower comorbidity burden (mean CCI=0.95 versus 1.81; p<0.001), and a higher proportion insured by Medicaid (18.3% versus 13.8%; p<0.001). Propensity-score matching yielded 7,254 BP users and 7,254 BP non-users with no significant differences across examined characteristics. A total of 108 BP non-users from the unmatched New York state “*Bone-Rx*” analysis cohort were not assigned an applicable BP user pair during the matching procedure and were excluded from the matched BP non-user population.

### Osteo-Dx-Rx Analysis Study Population

A total of 60,043 female patients age ≥51 with a diagnosis of osteoporosis who resided in New York (NY), Illinois (IL), Florida (FL), or California (CA) were included in the unmatched “*Osteo-Dx-Rx*” analysis cohort comparison, of which 51,651 (86.0%) and 8,392 (14.0%) were classified as BP users and BP non-users, respectively (Table S7). Prior to propensity-score matching, which was performed within each state by insurance type, there were significant differences across all demographic and clinical characteristics except the proportion of patients with a diagnosis of dyslipidemia (p=0.08). Compared to BP non-users, BP users were younger (18.8% age ≥81 versus 26.0%; p<0.001), with a larger proportion of patients residing in CA (42.5% versus 30.5%; p<0.001), insured by Medicaid (23.1% versus 21.3%; p<0.001), have had a primary-care physician (PCP) visit in 2019 (77.4% versus 71.1%; p<0.001), had a higher proportion with a diagnosis of obesity (11.2% versus 9.6%; p<0.001, and had a lower proportion diagnosed with the following: cancer (11.8% versus 19.4%; p<0.001), COPD (10.1% versus 16.2%; p<0.001), heart failure (6.1% versus 10.7%; p<0.001), hypertension (58.0% versus 60.9%; p<0.001), type 2 diabetes (25.6% versus 26.9%; p<0.01), and depression (13.9% versus 15.2%; p<0.001). Propensity-score matching yielded 7,949 BP users and 7,949 BP non-users with no significant differences across examined characteristics. A total of 443 BP non-users from the unmatched “*Osteo-Dx-Rx*” analysis cohort were not assigned an applicable BP user pair during the matching procedure and were excluded from the matched BP non-user population.

### Statin User/Non-User Analysis

#### Statin-Use Comparison: All Observations (all regions combined)

A total of 7,906,603 patients were included in the unmatched analysis cohort comparison of statin-use, of which 1,503,395 (19.0%) and 6,403,208 (81.0%) were classified as statin users and statin non-users, respectively (Table S8a). Prior to propensity-score matching, there were significant differences across all demographic and clinical characteristics. Compared to statin non-users, statin users were older (87.9% age ≥51 versus 37.1%; p<0.001), with a higher proportion of males (41.1% versus 40.9%; p<0.001), from the northeast (29.7% versus 26.6%; p<0.001), with higher comorbidity burden (mean CCI=1.15 versus 0.49; p<0.001), insured by Medicare (32.7% versus 11.3%; p<0.001), and have had a primary-care physician (PCP) visit in 2019 (66.1% versus 44.1%; p<0.001). Propensity-score matching yielded 1,436,300 statin users and 1,436,300 statin non-users with no significant differences across age group, region, insurance type, and having had any PCP visit in 2019. The final matched population did, however, display statistically significant differences between statin users and statin non-users for gender (58.7% versus 58.4% male; p<0.001) and mean CCI (1.11 versus 1.12; p<0.001). These differences, however, are small in magnitude, and were statistically significant due to the underlying statistical power associated with the large sample size. A total of 67,095 statin users from the unmatched analysis cohort were not assigned an applicable statin non-user pair during the matching procedure and were excluded from the matched statin user population.

#### Statin-Use Comparison: New York State

A total of 968,296 patients identified as residing in New York state were included in the unmatched analysis cohort comparison of statin-use, of which 206,301 (21.3%) and 761,995 (78.7%) were classified as statin users and statin non-users, respectively (Table S8b). Prior to propensity-score matching, there were significant differences across all demographic and clinical characteristics. Compared to statin non-users, statin users were older (90.3% age ≥51 versus 43.1%; p<0.001), with a higher proportion of males (42.0% versus 40.4%; p<0.001), with higher comorbidity burden (mean CCI=1.17 versus 0.51; p<0.001), insured by Medicare (47.4% versus 14.5%; p<0.001), and have had a primary-care physician (PCP) visit in 2019 (64.0% versus 41.3%; p<0.001). Propensity-score matching yielded 185,536 statin users and 185,536 statin non-users with no significant differences across age group, gender, insurance type, and having had any PCP visit in 2019. The final matched population did, however, display statistically significant differences between statin users and statin non-users for mean CCI (1.06 versus 1.08; p<0.001). This difference, however, is small in magnitude, and was statistically significant due to the underlying statistical power associated with the large sample size. A total of 20,765 statin users from the unmatched analysis cohort were not assigned an applicable statin non-user pair during the matching procedure and were excluded from the matched statin user population.

#### BP-Use Comparison within Statin Users: All Regions Combined

Of the 1,436,300 statin users from the statin user/non-user propensity-score matching analysis, a total of 217,981 (15.2%) and 1,218,319 (84.8%) were classified as BP users and BP non-users, respectively (Table S8c). Prior to propensity-score matching based on BP-use, there were significant differences across all demographic and clinical characteristics except for any PCP visit in 2019 (p=0.27). Compared to BP non-users, BP users were older (98.9% age ≥51 versus 85.3%; p<0.001), with a higher proportion of females (90.1% versus 53.1%; p<0.001), from the west (21.7% versus 14.0%; p<0.001), with lower comorbidity burden (mean CCI=0.95 versus 1.13; p<0.001), and insured by Medicare (50.8% versus 29.7%; p<0.001). Propensity-score matching yielded 213,480 BP users and 213,480 BP non-users with no significant differences across examined characteristics. A total of 4,501 BP users were not assigned an applicable BP non-user pair during the matching procedure and were excluded from the matched BP user population.

#### BP-Use Comparison within Statin Users: New York State

Of the 185,536 statin users from the statin user/non-user propensity-score matching analysis on patients residing in New York state, a total of 23,863 (12.9%) and 161,673 (87.1%) were classified as BP users and BP non-users, respectively (Table S8d). Prior to propensity-score matching based on BP-use, there were significant differences across all demographic and clinical characteristics except for any PCP visit in 2019 (p=0.33). Compared to BP non-users, BP users were older (99.3% age ≥51 versus 87.7%; p<0.001), with a higher proportion of females (91.2% versus 53.3%; p<0.001), with lower comorbidity burden (mean CCI=0.92 versus 1.08; p<0.001), and insured by Medicare (66.4% versus 41.9%; p<0.001). Propensity-score matching yielded 23,736 BP users and 23,736 BP non-users with no significant differences across examined characteristics. A total of 127 BP users were not assigned an applicable BP non-user pair during the matching procedure and were excluded from the matched BP user population.

#### BP-Use Comparison within Statin Non-users: All Regions Combined

Of the 1,436,300 statin non-users from the statin user/non-user propensity-score matching analysis, a total of 124,843 (8.7%) and 1,311,457 (91.3%) were classified as BP users and BP non-users, respectively (Table S8e). Prior to propensity-score matching based on BP-use, there were significant differences across all demographic and clinical characteristics. Compared to BP non-users, BP users were older (98.7% age ≥51 versus 86.3%; p<0.001), with a higher proportion of females (89.6% versus 55.5%; p<0.001), from the west (21.4% versus 14.6%; p<0.001), with lower comorbidity burden (mean CCI=1.02 versus 1.13; p<0.001), insured by Medicare (45.8% versus 31.7%; p<0.001), and have had a primary-care physician (PCP) visit in 2019 (71.7% versus 63.9%; p<0.001). Propensity-score matching yielded 124,716 BP users and 124,716 BP non-users with no significant differences across examined characteristics. A total of 127 BP users were not assigned an applicable BP non-user pair during the matching procedure and were excluded from the matched BP user population.

#### BP-Use Comparison within Statin Non-users: New York State

Of the 185,536 statin non-users from the statin user/non-user propensity-score matching analysis on patients residing in New York state, a total of 14,546 (7.8%) and 170,990 (92.2%) were classified as BP users and BP non-users, respectively (Table S8f). Prior to propensity-score matching based on BP-use, there were significant differences across all demographic and clinical characteristics. Compared to BP non-users, BP users were older (99.2% age ≥51 versus 88.4%; p<0.001), with a higher proportion of females (90.6% versus 55.1%; p<0.001), with lower comorbidity burden (mean CCI=0.95 versus 1.09; p<0.001), insured by Medicare (59.7% versus 43.7%; p<0.001), and have had a primary-care physician (PCP) visit in 2019 (70.5% versus 59.4%; p<0.001). Propensity-score matching yielded 14,521 BP users and 14,521 BP non-users with no significant differences across examined characteristics. A total of 25 BP users were not assigned an applicable BP non-user pair during the matching procedure and were excluded from the matched BP user population.

### Antihypertensive User/Non-User Analysis

#### Antihypertensive-Use Comparison: All Observations (all regions combined)

A total of 7,906,603 patients were included in the unmatched analysis cohort comparison of antihypertensive-use, of which 2,101,120 (26.6%) and 5,805,483 (73.4%) were classified as antihypertensive users and antihypertensive non-users, respectively (Table S9a). Prior to propensity-score matching, there were significant differences across all demographic and clinical characteristics. Compared to antihypertensive non-users, antihypertensive users were older (80.8% age ≥51 versus 34.4%; p<0.001), with a higher proportion of females (60.4% versus 58.6%; p<0.001), from the northeast (27.8% versus 27.0%; p<0.001), with higher comorbidity burden (mean CCI=1.13 versus 0.43; p<0.001), insured by Medicare (29.5% versus 10.3%; p<0.001), and have had a primary-care physician (PCP) visit in 2019 (64.2% versus 39.2%; p<0.001). Propensity-score matching yielded 1,786,001 antihypertensive users and 1,786,001 antihypertensive non-users with no significant differences across age group, gender, region, insurance type, and having had any PCP visit in 2019. The final matched population did, however, display statistically significant difference between antihypertensive users and antihypertensive non-users for mean CCI (1.64 versus 1.66; p<0.05). This difference, however, is small in magnitude, and was statistically significant due to the underlying statistical power associated with the large sample size. A total of 315,119 antihypertensive users from the unmatched analysis cohort were not assigned an applicable antihypertensive non-user pair during the matching procedure and were excluded from the matched antihypertensive user population.

#### Antihypertensive-Use Comparison: New York State

A total of 968,296 patients identified as residing in New York state were included in the unmatched analysis cohort comparison of antihypertensive-use, of which 258,652 (26.7%) and 709,644 (73.3%) were classified as antihypertensive users and antihypertensive non-users, respectively (Table S9b). Prior to propensity-score matching, there were significant differences across all demographic and clinical characteristics. Compared to antihypertensive non-users, antihypertensive users were older (86.6% age ≥51 versus 40.9%; p<0.001), with a higher proportion of females (59.4% versus 59.2%; p=0.02), with higher comorbidity burden (mean CCI=1.17 versus 0.46; p<0.001), insured by Medicare (45.9% versus 12.6%; p<0.001), and have had a primary-care physician (PCP) visit in 2019 (62.4% versus 40.3%; p<0.001). Propensity-score matching yielded 203,624 antihypertensive users and 203,624 antihypertensive non-users with no significant differences across examined characteristics. A total of 55,028 antihypertensive users from the unmatched analysis cohort were not assigned an applicable antihypertensive non-user pair during the matching procedure and were excluded from the matched antihypertensive user population.

#### BP-Use Comparison within Antihypertensive Users: All Regions Combined

Of the 1,786,001 antihypertensive users from the antihypertensive user/non-user propensity-score matching analysis, a total of 206,613 (11.6%) and 1,579,388 (88.4%) were classified as BP users and BP non-users, respectively (Table S9c). Prior to propensity-score matching based on BP-use, there were significant differences across all demographic and clinical characteristics. Compared to BP non-users, BP users were older (98.2% age ≥51 versus 75.2%; p<0.001), with a higher proportion of females (89.7% versus 56.6%; p<0.001), from the west (22.0% versus 14.3%; p<0.001), with lower comorbidity burden (mean CCI=0.94 versus 0.95; p=0.02), insured by Medicare (48.6% versus 24.4%; p<0.001), and have not had a primary-care physician (PCP) visit in 2019 (41.2% versus 40.1%; p<0.001). Propensity-score matching yielded 204,396 BP users and 204,396 BP non-users with no significant differences across examined characteristics. A total of 2,217 BP users were not assigned an applicable BP non-user pair during the matching procedure and were excluded from the matched BP user population.

#### BP-Use Comparison within Antihypertensive Users: New York State

Of the 203,624 antihypertensive users from the antihypertensive user/non-user propensity-score matching analysis on patients residing in New York state, a total of 21,213 (10.4%) and 182,411 (89.6%) were classified as BP users and BP non-users, respectively (Table S9d). Prior to propensity-score matching based on BP-use, there were significant differences across all demographic and clinical characteristics. Compared to BP non-users, BP users were older (98.8% age ≥51 versus 81.4%; p<0.001), with a higher proportion of females (90.9% versus 55.5%; p<0.001), with lower comorbidity burden (mean CCI=0.88 versus 0.95; p<0.001), insured by Medicare (64.1% versus 35.9%; p<0.001), and have not had a primary-care physician (PCP) visit in 2019 (53.4% versus 55.7%; p<0.001). Propensity-score matching yielded 21,126 BP users and 21,126 BP non-users with no significant differences across examined characteristics. A total of 87 BP users were not assigned an applicable BP non-user pair during the matching procedure and were excluded from the matched BP user population.

#### BP-Use Comparison within Antihypertensive Non-users: All Regions Combined

Of the 1,786,001 antihypertensive non-users from the antihypertensive user/non-user propensity-score matching analysis, a total of 136,016 (7.6%) and 1,649,985 (92.4%) were classified as BP users and BP non-users, respectively (Table S9e). Prior to propensity-score matching based on BP-use, there were significant differences across all demographic and clinical characteristics. Compared to BP non-users, BP users were older (97.7% age ≥51 versus 76.3%; p<0.001), with a higher proportion of females (90.5% versus 58.0%; p<0.001), from the west (20.3% versus 14.8%; p<0.001), with lower comorbidity burden (mean CCI=0.88 versus 0.96; p<0.001), insured by Medicare (40.7% versus 26.0%; p<0.001), and have had a primary-care physician (PCP) visit in 2019 (68.0% versus 59.0%; p<0.001). Propensity-score matching yielded 135,724 BP users and 135,724 BP non-users with no significant differences across examined characteristics. A total of 292 BP users were not assigned an applicable BP non-user pair during the matching procedure and were excluded from the matched BP user population.

#### BP-Use Comparison within Antihypertensive Non-users: New York State

Of the 203,624 antihypertensive non-users from the antihypertensive user/non-user propensity-score matching analysis on patients residing in New York state, a total of 14,051 (6.9%) and 189,573 (93.1%) were classified as BP users and BP non-users, respectively (Table S9f). Prior to propensity-score matching based on BP-use, there were significant differences across all demographic and clinical characteristics. Compared to BP non-users, BP users were older (98.7% age ≥51 versus 82.1%; p<0.001), with a higher proportion of females (91.3% versus 56.8%; p<0.001), with lower comorbidity burden (mean CCI=0.81 versus 0.96; p<0.001), insured by Medicare (54.9% versus 37.7%; p<0.001), and have had a primary-care physician (PCP) visit in 2019 (66.3% versus 54.7%; p<0.001). Propensity-score matching yielded 13,983 BP users and 13,983 BP non-users with no significant differences across examined characteristics. A total of 68 BP users were not assigned an applicable BP non-user pair during the matching procedure and were excluded from the matched BP user population.

### Antidiabetic User/Non-User Analysis

#### Antidiabetic-Use Comparison: All Observations (all regions combined)

A total of 7,906,603 patients were included in the unmatched analysis cohort comparison of antidiabetic-use, of which 755,252 (9.6%) and 7,151,351 (90.4%) were classified as antidiabetic users and antidiabetic non-users, respectively (Table S10a). Prior to propensity-score matching, there were significant differences across all demographic and clinical characteristics. Compared to antidiabetic non-users, antidiabetic users were older (79.4% age ≥51 versus 43.3%; p<0.001), with a higher proportion of females (60.8% versus 58.9%; p<0.001), from the northeast (28.8% versus 27.1%; p<0.001), with higher comorbidity burden (mean CCI=1.25 versus 0.55; p<0.001), insured by Medicare (26.2% versus 14.2%; p<0.001), and have had a primary-care physician (PCP) visit in 2019 (66.5% versus 43.6%; p<0.001). Propensity-score matching yielded 754,553 antidiabetic users and 754,553 antidiabetic non-users with no significant differences across examined characteristics. A total of 699 antidiabetic users from the unmatched analysis cohort were not assigned an applicable antidiabetic non-user pair during the matching procedure and were excluded from the matched antidiabetic user population.

#### Antidiabetic-Use Comparison: New York State

A total of 968,296 patients identified as residing in New York state were included in the unmatched analysis cohort comparison of antidiabetic-use, of which 105,117 (10.9%) and 863,179 (89.1%) were classified as antidiabetic users and antidiabetic non-users, respectively (Table S10b). Prior to propensity-score matching, there were significant differences across all demographic and clinical characteristics. Compared to antidiabetic non-users, antidiabetic users were older (83.8% age ≥51 versus 49.4%; p<0.001), with a higher proportion of males (42.2% versus 40.6%; p<0.001), with higher comorbidity burden (mean CCI=1.34 versus 0.56; p<0.001), insured by Medicare (40.5% versus 19.2%; p<0.001), and have had a primary-care physician (PCP) visit in 2019 (64.6% versus 43.9%; p<0.001). Propensity-score matching yielded 104,691 antidiabetic users and 104,691 antidiabetic non-users with no significant differences across examined characteristics. A total of 426 antidiabetic users from the unmatched analysis cohort were not assigned an applicable antidiabetic non-user pair during the matching procedure and were excluded from the matched antidiabetic user population.

#### BP-Use Comparison within Antidiabetic Users: All Regions Combined

Of the 754,553 antidiabetic users from the antidiabetic user/non-user propensity-score matching analysis, a total of 80,529 (10.7%) and 674,024 (89.3%) were classified as BP users and BP non-users, respectively (Table S10c). Prior to propensity-score matching based on BP-use, there were significant differences across all demographic and clinical characteristics. Compared to BP non-users, BP users were older (98.2% age ≥51 versus 75.2%; p<0.001), with a higher proportion of females (98.5% versus 77.1%; p<0.001), from the west (22.2% versus 14.2%; p<0.001), with a higher comorbidity burden (mean CCI=1.32 versus 1.23; p<0.001), insured by Medicare (45.2% versus 24.0%; p<0.001), and have had a primary-care physician (PCP) visit in 2019 (69.5% versus 66.1%; p<0.001). Propensity-score matching yielded 79,500 BP users and 79,500 BP non-users with no significant differences across examined characteristics. A total of 1,029 BP users were not assigned an applicable BP non-user pair during the matching procedure and were excluded from the matched BP user population.

#### BP-Use Comparison within Antidiabetic Users: New York State

Of the 104,691 antidiabetic users from the antidiabetic user/non-user propensity-score matching analysis on patients residing in New York state, a total of 9,529 (9.1%) and 95,162 (90.9%) were classified as BP users and BP non-users, respectively (Table S10d). Prior to propensity-score matching based on BP-use, there were significant differences across all demographic and clinical characteristics. Compared to BP non-users, BP users were older (99.1% age ≥51 versus 82.2%; p<0.001), with a higher proportion of females (90.1% versus 54.5%; p<0.001), with a higher comorbidity burden (mean CCI=1.46 versus 1.31; p<0.001), insured by Medicare (64.6% versus 38.2%; p<0.001), and have had a primary-care physician (PCP) visit in 2019 (66.3% versus 64.4%; p<0.001). Propensity-score matching yielded 9,456 BP users and 9,456 BP non-users with no significant differences across examined characteristics. A total of 73 BP users were not assigned an applicable BP non-user pair during the matching procedure and were excluded from the matched BP user population.

#### BP-Use Comparison within Antidiabetic Non-users: All Regions Combined

Of the 754,553 antidiabetic non-users from the antidiabetic user/non-user propensity-score matching analysis, a total of 73,173 (9.7%) and 681,380 (90.3%) were classified as BP users and BP non-users, respectively (Table S10e). Prior to propensity-score matching based on BP-use, there were significant differences across all demographic characteristics, but no difference was seen in mean CCI (1.24 versus 1.24; p=0.92). Compared to BP non-users, BP users were older (98.0% age ≥51 versus 77.3%; p<0.001), with a higher proportion of females (88.9% versus 57.7%; p<0.001), from the west (20.1% versus 14.5%; p<0.001), insured by Medicare (40.0% versus 24.8%; p<0.001), and have had a primary-care physician (PCP) visit in 2019 (74.1% versus 65.7%; p<0.001). Propensity-score matching yielded 72,514 BP users and 72,514 BP non-users with no significant differences across examined characteristics. A total of 659 BP users were not assigned an applicable BP non-user pair during the matching procedure and were excluded from the matched BP user population.

#### BP-Use Comparison within Antidiabetic Non-users: New York State

Of the 104,691 antidiabetic non-users from the antidiabetic user/non-user propensity-score matching analysis on patients residing in New York state, a total of 9,275 (8.9%) and 95,416 (91.1%) were classified as BP users and BP non-users, respectively (Table S10f). Prior to propensity-score matching based on BP-use, there were significant differences across all demographic and clinical characteristics. Compared to BP non-users, BP users were older (99.0% age ≥51 versus 82.2%; p<0.001), with a higher proportion of females (89.2% versus 54.7%; p<0.001), with a higher comorbidity burden (mean CCI=1.37 versus 1.32; p<0.01), insured by Medicare (57.7% versus 38.9%; p<0.001), and have had a primary-care physician (PCP) visit in 2019 (72.5% versus 63.8%; p<0.001). Propensity-score matching yielded 13,983 BP users and 13,983 BP non-users with no significant differences across examined characteristics. A total of 131 BP users were not assigned an applicable BP non-user pair during the matching procedure and were excluded from the matched BP user population.

### Antidepressant User/Non-User Analysis

#### Antidepressant-Use Comparison: All Observations (all regions combined)

A total of 7,906,603 patients were included in the unmatched analysis cohort comparison of antidepressant-use, of which 1,571,005 (19.9%) and 6,335,598 (80.1%) were classified as antidepressant users and antidepressant non-users, respectively (Table S11a). Prior to propensity-score matching, there were significant differences across all demographic and clinical characteristics. Compared to antidepressant non-users, antidepressant users were older (58.6% age ≥51 versus 43.8%; p<0.001), with a higher proportion of females (72.8% versus 55.7%; p<0.001), from the midwest (22.1% versus 17.7%; p<0.001), with higher comorbidity burden (mean CCI=0.90 versus 0.55; p<0.001), insured by Medicare (18.5% versus 14.6%; p<0.001), and have had a primary-care physician (PCP) visit in 2019 (61.1% versus 42.0%; p<0.001). Propensity-score matching yielded 1,536,048 antidepressant users and 1,536,048 antidepressant non-users with no significant differences across examined characteristics. A total of 34,957 antidepressant users from the unmatched analysis cohort were not assigned an applicable antidepressant non-user pair during the matching procedure and were excluded from the matched antidepressant user population.

#### Antidepressant-Use Comparison: New York State

A total of 968,296 patients identified as residing in New York state were included in the unmatched analysis cohort comparison of antidepressant-use, of which 136,081 (14.1%) and 832,215 (85.9%) were classified as antidepressant users and antidepressant non-users, respectively (Table S11b). Prior to propensity-score matching, there were significant differences across all demographic and clinical characteristics. Compared to antidepressant non-users, antidepressant users were older (66.3% age ≥51 versus 51.0%; p<0.001), with a higher proportion of females (71.2% versus 57.3%; p<0.001), with higher comorbidity burden (mean CCI=0.98 versus 0.59; p<0.001), insured by Medicare (32.2% versus 19.8%; p<0.001), and have had a primary-care physician (PCP) visit in 2019 (60.7% versus 43.8%; p<0.001). Propensity-score matching yielded 135,516 antidepressant users and 135,516 antidepressant non-users with no significant differences across examined characteristics. A total of 565 antidepressant users from the unmatched analysis cohort were not assigned an applicable antidepressant non-user pair during the matching procedure and were excluded from the matched antidepressant user population.

#### BP-Use Comparison within Antidepressant Users: All Regions Combined

Of the 1,536,048 antidepressant users from the antidepressant user/non-user propensity-score matching analysis, a total of 145,109 (9.4%) and 1,390,939 (90.6%) were classified as BP users and BP non-users, respectively (Table S11c). Prior to propensity-score matching based on BP-use, there were significant differences across all demographic and clinical characteristics. Compared to BP non-users, BP users were older (96.7% age ≥51 versus 54.4%; p<0.001), with a higher proportion of females (91.9% versus 70.2%; p<0.001), from the west (19.6% versus 13.9%; p<0.001), with a higher comorbidity burden (mean CCI=1.09 versus 0.84; p<0.001), insured by Medicare (42.4% versus 16.2%; p<0.001), and have had a primary-care physician (PCP) visit in 2019 (64.6% versus 60.2%; p<0.001). Propensity-score matching yielded 144,282 BP users and 144,282 BP non-users with no significant differences across examined characteristics. A total of 827 BP users were not assigned an applicable BP non-user pair during the matching procedure and were excluded from the matched BP user population.

#### BP-Use Comparison within Antidepressant Users: New York State

Of the 135,516 antidepressant users from the antidepressant user/non-user propensity-score matching analysis on patients residing in New York state, a total of 12,950 (9.6%) and 122,566 (90.4%) were classified as BP users and BP non-users, respectively (Table S11d). Prior to propensity-score matching based on BP-use, there were significant differences across all demographic and clinical characteristics. Compared to BP non-users, BP users were older (97.8% age ≥51 versus 63.0%; p<0.001), with a higher proportion of females (92.6% versus 68.9%; p<0.001), with a higher comorbidity burden (mean CCI=1.13 versus 0.95; p<0.001), insured by Medicare (60.8% versus 29.1%; p<0.001), and have had a primary-care physician (PCP) visit in 2019 (65.3% versus 60.1%; p<0.001). Propensity-score matching yielded 12,859 BP users and 12,859 BP non-users with no significant differences across examined characteristics. A total of 91 BP users were not assigned an applicable BP non-user pair during the matching procedure and were excluded from the matched BP user population.

#### BP-Use Comparison within Antidepressant Non-users: All Regions Combined

Of the 1,536,048 antidepressant non-users from the antidepressant user/non-user propensity-score matching analysis, a total of 113,110 (7.4%) and 1,422,938 (92.6%) were classified as BP users and BP non-users, respectively (Table S11e). Prior to propensity-score matching based on BP-use, there were significant differences across all demographic characteristics. Compared to BP non-users, BP users were older (97.1% age ≥51 versus 55.4%; p<0.001), with a higher proportion of females (93.2% versus 70.6%; p<0.001), from the west (20.0% versus 14.0%; p<0.001), with a higher comorbidity burden (mean CCI=1.06 versus 0.85; p<0.001), insured by Medicare (40.4% versus 17.0%; p<0.001), and have had a primary-care physician (PCP) visit in 2019 (71.2% versus 59.8%; p<0.001). Propensity-score matching yielded 112,402 BP users and 112,402 BP non-users with no significant differences across examined characteristics. A total of 708 BP users were not assigned an applicable BP non-user pair during the matching procedure and were excluded from the matched BP user population.

#### BP-Use Comparison within Antidepressant Non-users: New York State

Of the 135,516 antidepressant non-users from the antidepressant user/non-user propensity-score matching analysis on patients residing in New York state, a total of 10,174 (7.5%) and 125,342 (92.5%) were classified as BP users and BP non-users, respectively (Table S11f). Prior to propensity-score matching based on BP-use, there were significant differences across all demographic and clinical characteristics. Compared to BP non-users, BP users were older (98.4% age ≥51 versus 63.7%; p<0.001), with a higher proportion of females (93.6% versus 69.4%; p<0.001), with a higher comorbidity burden (mean CCI=1.13 versus 0.95; p<0.01), insured by Medicare (60.0% versus 29.9%; p<0.001), and have had a primary-care physician (PCP) visit in 2019 (71.7% versus 59.7%; p<0.001). Propensity-score matching yielded 10,091 BP users and 10,091 BP non-users with no significant differences across examined characteristics. A total of 83 BP users were not assigned an applicable BP non-user pair during the matching procedure and were excluded from the matched BP user population.

## APPENDIX 3: Post-hoc Analysis on the Impact of Censoring due to Death

### Background

Following completion of all core study analyses, an additional post-hoc investigation was performed to assess whether censoring bias due to patient death could impact our current findings of a decrease in the odds of COVID-19 outcomes seen amongst BP users. Typically, it is very difficult to perform assessments on this type of bias due to the fact that insurance claims databases in the United States do not include this information. Some claims database providers, including Komodo Health, do have the capability to ‘link’ their de-identified claims data with external sources on decedent enrollees, but at the time of study initiation and data extraction there were enhanced HIPAA constraints associated with claims datasets that included COVID-identifying diagnosis/treatment codes due to the heightened risk of patient re-identification due to the then lower prevalence and high visibility associated for patients with COVID-19. Eventually the increased prevalence of COVID-19 reduced the HIPAA concerns on working with claims data that include COVID-19-identifiers, and in support of this analysis and the potentially significant public health implications of our findings, Komodo Health linked their COVID-identifiable dataset with mortality data sources that account for roughly 80-85% of available death records. In conjunction with Komodo Health, queries on this mortality-linked COVID-19-identifiable dataset were performed to determine whether bias caused by patient censoring due to death could have impacted the validity and/or reliability of our current findings

### Methodological Concerns of Patient Censoring due to Death

The single motivating factor for initiation of this post-hoc analysis was the fact that the decrease in odds of COVID-19 outcomes among BP users in this study was found to be statistically significant, large in magnitude, and robust across almost all analysis variations performed. The exhaustive use of methodological techniques to control for unmeasured confounding and/or outside sources of bias employed in this current study were undertaken not in search of statistical significance, but in search of non-significance. This was undertaken because the consistency seen in statistical significance, in addition to the magnitude of the decrease in the odds of our outcomes of interest, are typically not seen to this degree. As such, the next logical step after exhausting all methodological techniques is to search for other sources that could induce a large-enough bias on the underlying patient population itself, such as censoring of the target study cohort, that could drastically alter the typical composition of the overall sample and thus impact the reliability and validity of outcomes measured.

The high rate of death associated with COVID-19 infection, which was even worse during the early months of the pandemic, represents such an instance where outside influences could impact the underlying data, and as such, the validity of research performed on that data. The primary concern is whether patients who have died are censored from the analytical sample due to the application of one of the most fundamental inclusion/exclusion criteria used in claims-based research, the requirement for continuous insurance eligibility over the entire study period that is needed so that healthcare resource utilization events from all subjects are captured and available in the data for analysis. If in our current sample, a larger number of BP users died after contracting COVID-19 and were censored due to insurance eligibility, and a lower number of BP non-users survived and thus met the insurance eligibility criteria, then the remaining study sample would be comprised of healthier-looking BP users and a higher number of BP non-users with COVID-19 related healthcare services.

The potential for such a censoring bias in this current study sample, and the impact of that bias on the magnitude and statistical significance of our core study findings, was assessed in this post-hoc analysis by: (1) adjusting eligibility criteria to prevent the censoring of patients that may have died during the first half of 2020; (2) replicating key exposure (BP-use, use of other non-BP bone health medications) and outcomes (COVID-19 diagnosis) in this expanded sample that aligns with the core study methods; (3) analysing the impact on study findings that would result from the retention and inclusion of deceased-patient observations in the core study sample on the odds of COVID-19 diagnosis; and (4) calculating the number of missing patient observations censored due to death that would be required to reach a statistically non-significant difference in the odds of COVID-19.

### Post-Hoc Analysis

#### Methods

##### Cohort Definition

- Continuous insurance eligibility 1/1/2019-12/31/2019; used to ensure that any censoring due to death occurs during the observation period of 1/1/2020-6/30/2020
- BP users compared to BP non-users to produce a cohort comparison similar to the primary analysis cohort
- BP users compared to users of non-BP anti-resorptive bone health medications to produce a cohort comparison similar to the “*Bone-Rx*” active comparator analysis

##### Exposures of Interest

- Patients were assigned into the BP user cohort if they had any claim 1/1/2019-2/29/2020 for one of the following: alendronate, alendronic acid, etidronate, ibandronate, ibandronic acid, pamidronate, risedronate, and zoledronic acid; for the cohort comparison of all osteoporosis medication users BP users were further restricted to those that had no claims for a non-BP anti-resorptive bone health medication 1/1/2019-2/29/2020.
- Patients were assigned into the non-BP anti-resorptive bone health medication user cohort if: (1) they had any claim 1/1/2019-2/29/2020 for one of the following: denosumab, calcitonin, raloxifene, romosozumab-aqqg, teriparatide, abaloparatide, or bazedoxifene; and (2) they had no BP claims

##### Outcomes / Endpoints

- Patients were assigned into the COVID-19 diagnosis cohort based on any medical service claim with an ICD-10 diagnosis code of U07.1 occurring 1/1/2200-6/30/2020
- Patients with a date-of-death between 1/1/2020-6/30/2020 were classified into the deceased cohort

##### Statistical Analysis

- Chi-square testing was used to assess whether statistically significant differences exist between BP users and BP non-users in the unadjusted odds of having any COVID-19 diagnosis during the first half of 2020 among cohorts that approximate the primary analysis and “*Bone-Rx*” study cohorts for the following:

1. Among all patient-observations with a COVID-19 diagnosis to assess the potential ‘true’ comparison that would occur
2. With deceased patient-observations that had a known COVID-19 diagnosis removed prior to testing to replicate findings that would occur if these observations were censored
3. When making the assumption that all patients who died during this period died due to COVID-19, and thus should be classified as having a COVID-19 diagnosis
- An additional analysis was performed on the last variation modelled (assuming all patients died due to COVID-19) to determine the additional BP user patient observations that would be needed to be classified as having had a COVID-19 diagnosis to yield a similar distribution of COVID-19 diagnosis (yes/no) as was seen in the BP non-user cohort to yield an odds ratio ∼1.0
- Finally, the impact on odds ratio testing results comparing BP users to BP non-users was modelled based on the additional number of BP users needed to be classified as having been diagnosed with COVID-19 to reach statistical non-significance

#### Results

##### Patient Count Distribution

- Among the full sample a decreased rate of COVID-19 among BP users compared to BP non-users was seen in both the full sample population (1.2% versus 4.7%) as well as when restricted to users of non-BP anti-resorptive bone health medications (1.2% versus 4.3%) (Table S12a)

##### Unadjusted Chi-Square Comparison Inclusive of Deceased Patients

- The decrease in the odds of any COVID-19 diagnosis amongst BP users compared to BP non-users was found to be robust in both the full (OR=0.24) and “*Bone-Rx*” (OR=0.35) comparisons when including deceased patients with a known COVID-19 diagnosis (Table S12b)

##### Unadjusted Chi-Square Comparison with Deceased Patients Removed

- The decrease in the odds of any COVID-19 diagnosis amongst BP users compared to BP non-users was found to be robust in both the full (OR=0.23) and “*Bone-Rx*” (OR=0.26) comparisons when removing deceased patients with a known COVID-19 diagnosis (Table S12c)

##### Unadjusted Chi-Square Comparison Assuming all Deceased Patients had COVID-19

- The decrease in the odds of any COVID-19 diagnosis amongst BP users compared to BP non-users was found to be robust in both the full (OR=0.39) and “*Bone-Rx*” (OR=0.29) comparisons when assuming that all deceased patients had a COVID-19 diagnosis (Table S12d)
- Among this final analysis that assumes all deceased patients had a diagnosis of COVID-19, the percentage of BP non-users with an assumed COVID-19 diagnosis was 5.5% and 7.2% for the full and OPRX comparisons, respectively.
- These proportions were then used to estimate the number of additional BP users with a COVID-19 diagnosis that would be needed to have the same distribution and thus an odds ratio ∼1.0 (Table S12e)
- It would require an additional 22,235 (37,095-14,860) BP-user patient observations from the full cohort comparison to be classified as having a COVID-19 diagnosis to have an equivalent odds of being diagnosed with COVID-19 as was seen among the BP non-user cohort
- It would require an additional 32,598 (46,637-14,039) BP-user patient observations from the “*Bone-Rx*” cohort comparison to be classified as having a COVID-19 diagnosis to have an equivalent odds of being diagnosed with COVID-19 as was seen among the BP non-user cohort
- In the full (all observations) comparison, the minimum number of additional BP users classified as having a COVID-19 diagnosis needed to reach statistical non-significance for the calculated unadjusted odds ratio was 21,860 (Figure S1)
- In the “*Bone-Rx*” comparison, the minimum number of additional BP users classified as having a COVID-19 diagnosis needed to reach statistical non-significance for the calculated unadjusted odds ratio was 31,360 (Figure S2)

---

**Figure S1:**
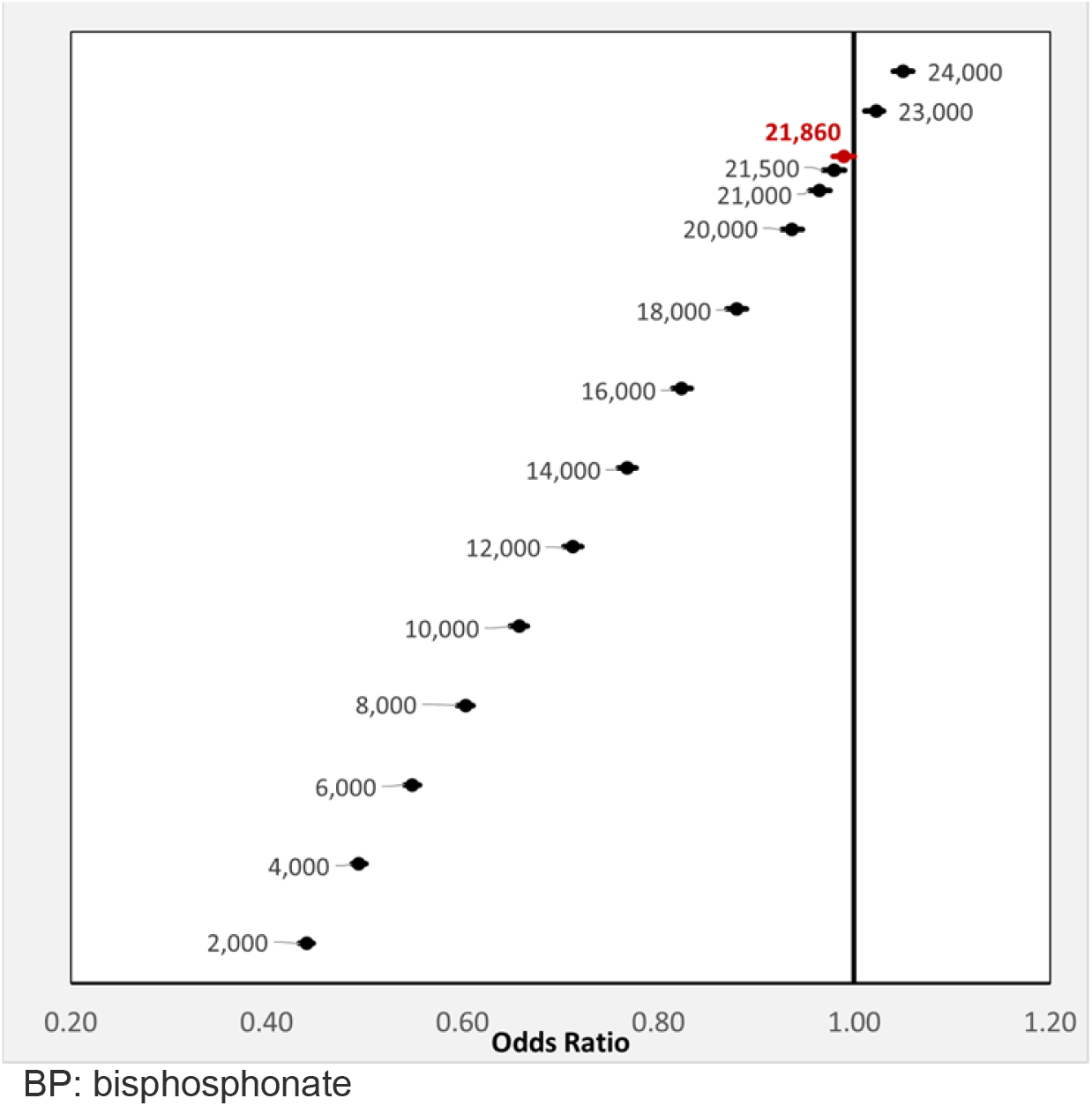
Full Cohort: Odds Ratio by Additional Number of BP Users Classified as having COVID-19 Diagnosis. <u>Legend:</u> Shown in Figure S1 is the forest plot of the change in the crude odds ratio (OR) of BP users having a COVID-19 diagnosis as a factor of the additional number of BP users needed to be classified as having a COVID-19 diagnosis to reach statistical non-significance for all observations.

**Figure S2:**
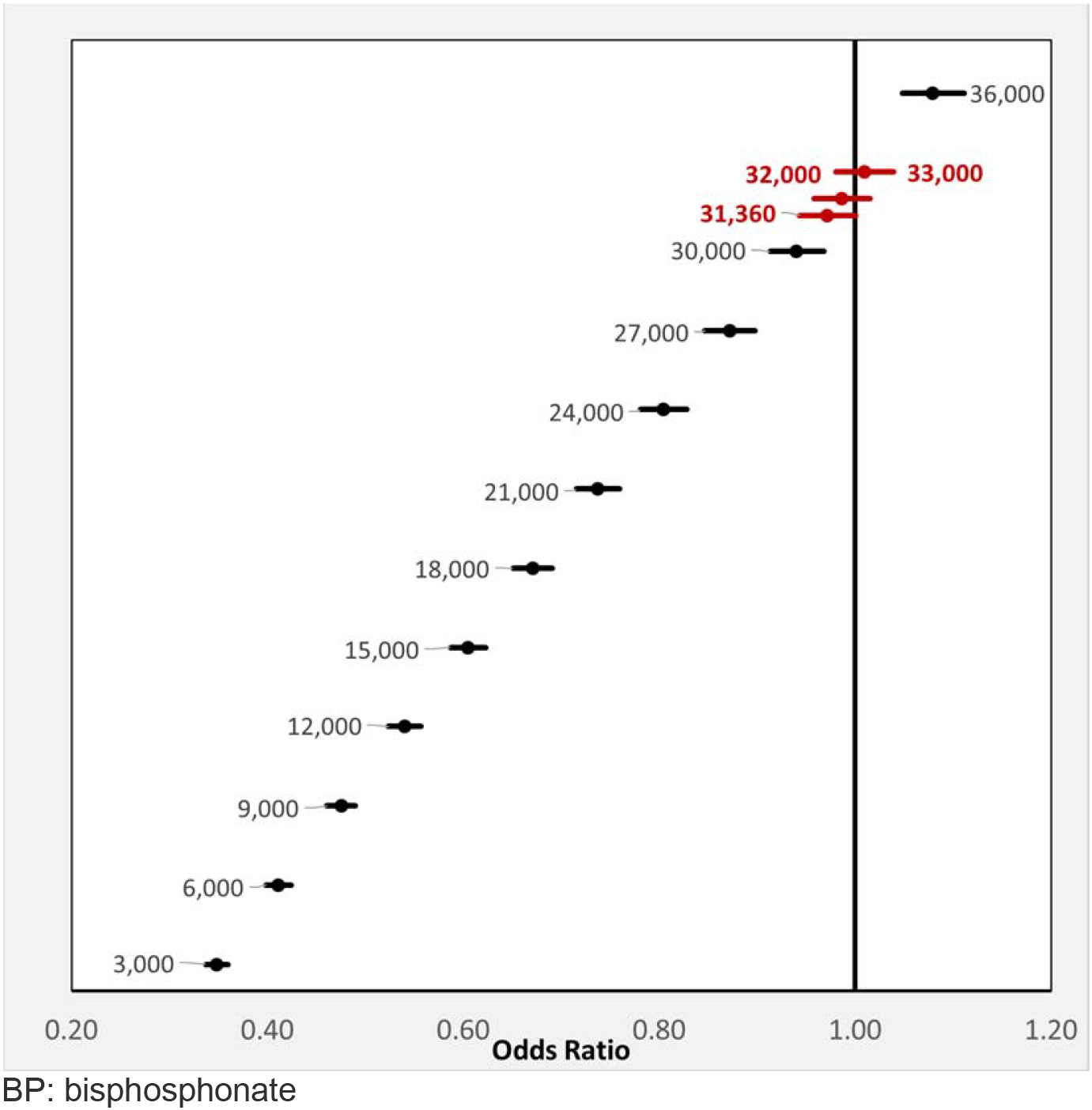
Bone-Rx Cohort: Odds Ratio by Additional Number of BP Users Classified as having COVID-19 Diagnosis. <u>Legend:</u> Shown in Figure S2 is the forest plot of the change in the crude odds ratio (OR) of BP users having a COVID-19 diagnosis as a factor of the additional number of BP users needed to be classified as having a COVID-19 diagnosis to reach statistical non-significance when comparing BP users to users of non-BP anti-resorptive bone medications.

## APPENDIX TABLES

**Table S1:**
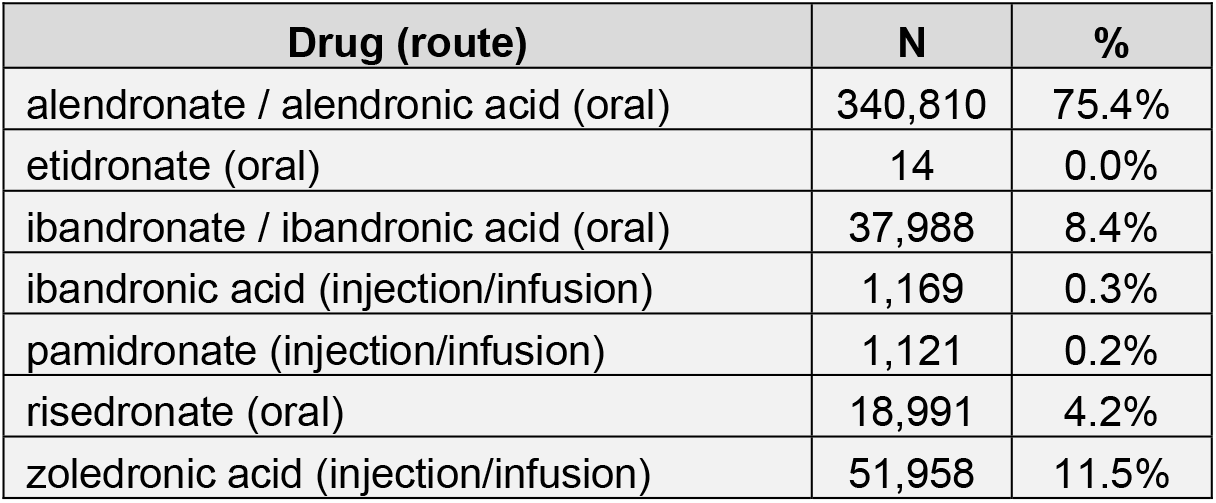
Most Recent Bisphosphonate Claim Among all Users

**Table S2a:** Primary Analysis Cohort (Region=Northeast), Patient Characteristics Pre/Post Match

|  | Region=Northeast Unmatched |  |  |  |  |  |  | Region=Northeast Matched |  |  |  |  |  |  |
| --- | --- | --- | --- | --- | --- | --- | --- | --- | --- | --- | --- | --- | --- | --- |
|  | All |  | BP Non-users |  | BP Users |  | p-value | All |  | BP Non-users |  | BP Users |  | p-value |
|  | N | % | N | % | N | % |  | N | % | N | % | N | % |  |
| All Patients | 2,152,560 | 100.0% | 2,032,832 | 94.4% | 119,728 | 5.6% |  | 238,988 | 100.0% | 119,494 | 50.0% | 119,494 | 50.0% |  |
| Age |  |  |  |  |  |  | <0.001 |  |  |  |  |  |  | 1.00 |
| ≤20 | 363,637 | 16.9% | 363,401 | 17.9% | 236 | 0.2% |  | 474 | 0.2% | 238 | 0.2% | 236 | 0.2% |  |
| 21-40 | 397,377 | 18.5% | 396,613 | 19.5% | 764 | 0.6% |  | 1,528 | 0.6% | 764 | 0.6% | 764 | 0.6% |  |
| 41-50 | 261,570 | 12.2% | 259,528 | 12.8% | 2,042 | 1.7% |  | 4,084 | 1.7% | 2,042 | 1.7% | 2,042 | 1.7% |  |
| 51-60 | 372,238 | 17.3% | 354,228 | 17.4% | 18,010 | 15.0% |  | 36,020 | 15.1% | 18,010 | 15.1% | 18,010 | 15.1% |  |
| 61-70 | 354,331 | 16.5% | 313,237 | 15.4% | 41,094 | 34.3% |  | 82,233 | 34.4% | 41,139 | 34.4% | 41,094 | 34.4% |  |
| 71-80 | 252,712 | 11.7% | 215,151 | 10.6% | 37,561 | 31.4% |  | 74,831 | 31.3% | 37,393 | 31.3% | 37,438 | 31.3% |  |
| ≥81 | 150,695 | 7.0% | 130,674 | 6.4% | 20,021 | 16.7% |  | 39,818 | 16.7% | 19,908 | 16.7% | 19,910 | 16.7% |  |
| Gender |  |  |  |  |  |  |  |  |  |  |  |  |  |  |
| Female | 1,275,611 | 59.3% | 1,167,241 | 57.4% | 108,370 | 90.5% | <0.001 | 216,273 | 90.5% | 108,137 | 90.5% | 108,136 | 90.5% | 0.99 |
| Male | 876,949 | 40.7% | 865,591 | 42.6% | 11,358 | 9.5% |  | 22,715 | 9.5% | 11,357 | 9.5% | 11,358 | 9.5% |  |
| Insurance |  |  |  |  |  |  |  |  |  |  |  |  |  |  |
| Commercial | 1,050,795 | 48.8% | 1,017,502 | 50.1% | 33,293 | 27.8% | <0.001 | 66,552 | 27.8% | 33,259 | 27.8% | 33,293 | 27.9% | 0.99 |
| Dual | 47,773 | 2.2% | 40,168 | 2.0% | 7,605 | 6.4% |  | 15,114 | 6.3% | 7,576 | 6.3% | 7,538 | 6.3% |  |
| Medicaid | 631,863 | 29.4% | 608,649 | 29.9% | 23,214 | 19.4% |  | 46,094 | 19.3% | 23,047 | 19.3% | 23,047 | 19.3% |  |
| Medicare | 422,129 | 19.6% | 366,513 | 18.0% | 55,616 | 46.5% |  | 111,228 | 46.5% | 55,612 | 46.5% | 55,616 | 46.5% |  |
| PCP Visit 2019 |  |  |  |  |  |  |  |  |  |  |  |  |  |  |
| No | 1,212,394 | 56.3% | 1,162,527 | 57.2% | 49,867 | 41.7% | <0.001 | 99,741 | 41.7% | 49,874 | 41.7% | 49,867 | 41.7% | 0.98 |
| Yes | 940,166 | 43.7% | 870,305 | 42.8% | 69,861 | 58.3% |  | 139,247 | 58.3% | 69,620 | 58.3% | 69,627 | 58.3% |  |
| Continuous Outcomes |  |  |  |  |  |  |  |  |  |  |  |  |  |  |
|  | mean | SD | mean | SD | mean | SD | p-value | mean | SD | mean | SD | mean | SD | p-value |
| CCI | 0.67 | 1.42 | 0.65 | 1.40 | 0.93 | 1.71 | <0.001 | 0.93 | 1.71 | 0.93 | 1.71 | 0.93 | 1.71 | 0.96 |

BP: bisphosphonate; CCI: Charlson Comorbidity Index; PCP: primary care physician; SD: standard deviation

**Table S2b:**
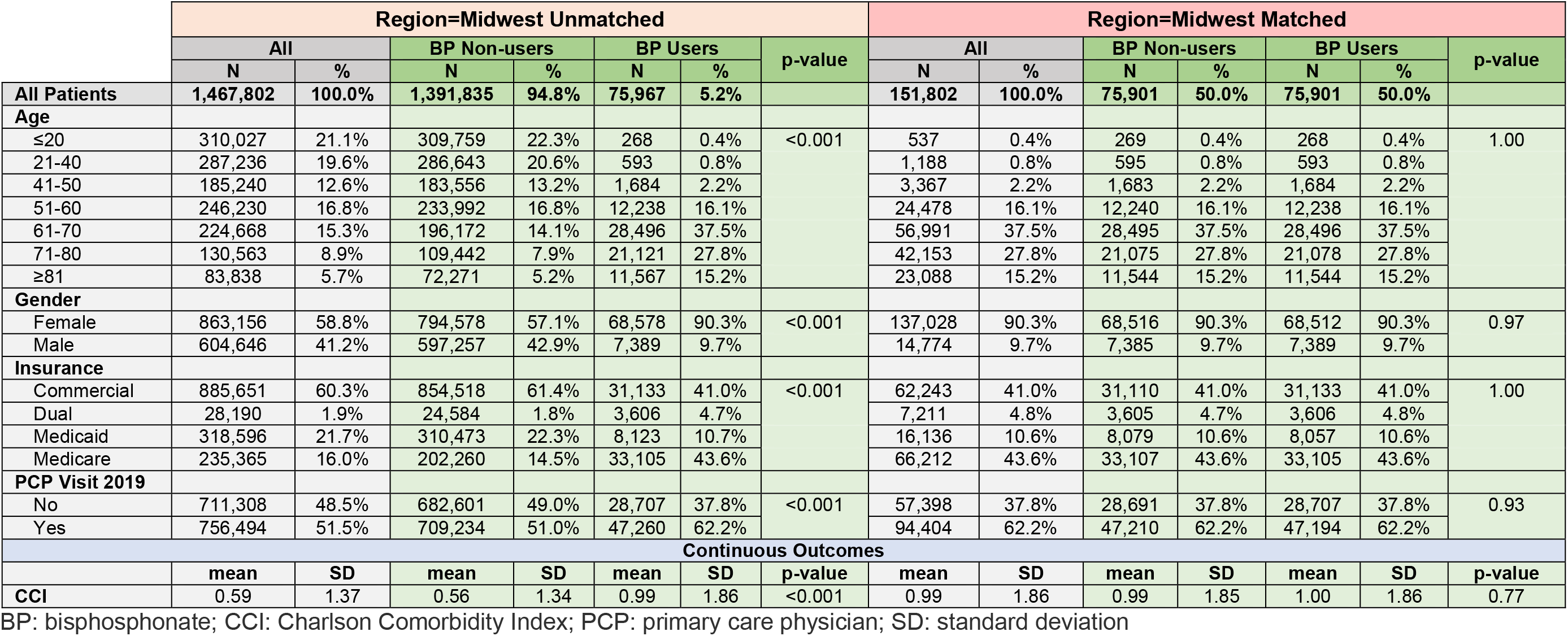
Primary Analysis Cohort (Region=Midwest), Patient Characteristics Pre/Post Match

**Table S2c:** Primary Analysis Cohort (Region=South), Patient Characteristics Pre/Post Match

|  | Region=South Unmatched |  |  |  |  |  |  | Region=South Matched |  |  |  |  |  |  |
| --- | --- | --- | --- | --- | --- | --- | --- | --- | --- | --- | --- | --- | --- | --- |
|  | All |  | BP Non-users |  | BP Users |  | p-value | All |  | BP Non-users |  | BP Users |  | p-value |
|  | N | % | N | % | N | % |  | N | % | N | % | N | % |  |
| All Patients | 3,042,604 | 100.0% | 2,881,718 | 94.7% | 160,886 | 5.3% |  | 319,408 | 100.0% | 159,704 | 50.0% | 159,704 | 50.0% |  |
| Age |  |  |  |  |  |  | <0.001 |  |  |  |  |  |  | 1.00 |
| ≤20 | 890,677 | 29.3% | 890,203 | 30.9% | 474 | 0.3% |  | 943 | 0.3% | 469 | 0.3% | 474 | 0.3% |  |
| 21-40 | 527,971 | 17.4% | 526,794 | 18.3% | 1,177 | 0.7% |  | 2,364 | 0.7% | 1,187 | 0.7% | 1,177 | 0.7% |  |
| 41-50 | 338,262 | 11.1% | 334,841 | 11.6% | 3,421 | 2.1% |  | 6,839 | 2.1% | 3,418 | 2.1% | 3,421 | 2.1% |  |
| 51-60 | 442,757 | 14.6% | 417,664 | 14.5% | 25,093 | 15.6% |  | 50,186 | 15.7% | 25,093 | 15.7% | 25,093 | 15.7% |  |
| 61-70 | 409,854 | 13.5% | 353,958 | 12.3% | 55,896 | 34.7% |  | 111,800 | 35.0% | 55,904 | 35.0% | 55,896 | 35.0% |  |
| 71-80 | 272,761 | 9.0% | 222,156 | 7.7% | 50,605 | 31.5% |  | 99,223 | 31.1% | 49,605 | 31.1% | 49,618 | 31.1% |  |
| ≥81 | 160,322 | 5.3% | 136,102 | 4.7% | 24,220 | 15.1% |  | 48,053 | 15.0% | 24,028 | 15.0% | 24,025 | 15.0% |  |
| Gender |  |  |  |  |  |  |  |  |  |  |  |  |  |  |
| Female | 1,800,166 | 59.2% | 1,654,351 | 57.4% | 145,815 | 90.6% | <0.001 | 289,263 | 90.6% | 144,630 | 90.6% | 144,633 | 90.6% | 0.99 |
| Male | 1,242,438 | 40.8% | 1,227,367 | 42.6% | 15,071 | 9.4% |  | 30,145 | 9.4% | 15,074 | 9.4% | 15,071 | 9.4% |  |
| Insurance |  |  |  |  |  |  |  |  |  |  |  |  |  |  |
| Commercial | 1,475,456 | 48.5% | 1,416,166 | 49.1% | 59,290 | 36.9% | <0.001 | 118,587 | 37.1% | 59,297 | 37.1% | 59,290 | 37.1% | 1.00 |
| Dual | 53,474 | 1.8% | 39,414 | 1.4% | 14,060 | 8.7% |  | 25,752 | 8.1% | 12,874 | 8.1% | 12,878 | 8.1% |  |
| Medicaid | 1,121,606 | 36.9% | 1,099,957 | 38.2% | 21,649 | 13.5% |  | 43,299 | 13.6% | 21,650 | 13.6% | 21,649 | 13.6% |  |
| Medicare | 392,068 | 12.9% | 326,181 | 11.3% | 65,887 | 41.0% |  | 131,770 | 41.3% | 65,883 | 41.3% | 65,887 | 41.3% |  |
| PCP Visit 2019 |  |  |  |  |  |  |  |  |  |  |  |  |  |  |
| No | 1,701,040 | 55.9% | 1,646,572 | 57.1% | 54,468 | 33.9% | <0.001 | 108,601 | 34.0% | 54,275 | 34.0% | 54,326 | 34.0% | 0.85 |
| Yes | 1,341,564 | 44.1% | 1,235,146 | 42.9% | 106,418 | 66.1% |  | 210,807 | 66.0% | 105,429 | 66.0% | 105,378 | 66.0% |  |
| Continuous Outcomes |  |  |  |  |  |  |  |  |  |  |  |  |  |  |
|  | mean | SD | mean | SD | mean | SD | p-value | mean | SD | mean | SD | mean | SD | p-value |
| CCI | 0.57 | 1.31 | 0.55 | 1.28 | 0.86 | 1.70 | <0.001 | 0.86 | 1.70 | 0.86 | 1.70 | 0.86 | 1.71 | 0.84 |

BP: bisphosphonate; CCI: Charlson Comorbidity Index; PCP: primary care physician; SD: standard deviation

**Table S2d:**
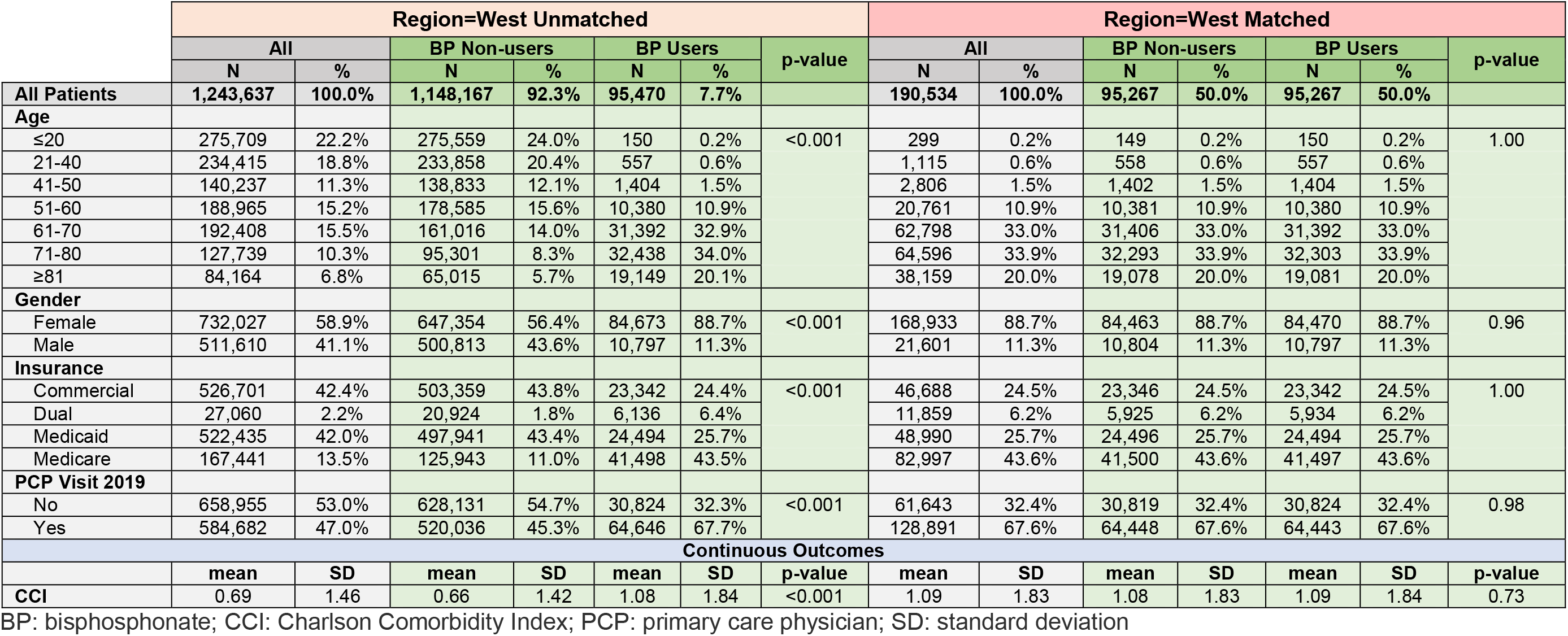
Primary Analysis Cohort (Region=West), Patient Characteristics Pre/Post Match

**Table S2e:** Primary Analysis Cohort (Region=New York State), Patient Characteristics Pre/Post Match

|  | Region=New York State Unmatched |  |  |  |  |  |  | Region=New York State Matched |  |  |  |  |  |  |
| --- | --- | --- | --- | --- | --- | --- | --- | --- | --- | --- | --- | --- | --- | --- |
|  | All |  | BP Non-users |  | BP Users |  | p-value | All |  | BP Non-users |  | BP Users |  | p-value |
|  | N | % | N | % | N | % |  | N | % | N | % | N | % |  |
| All Patients | 968,296 | 100.0% | 918,261 | 94.8% | 50,035 | 5.2% |  | 99,724 | 100.0% | 49,862 | 50.0% | 49,862 | 50.0% |  |
| Age |  |  |  |  |  |  | <0.001 |  |  |  |  |  |  | 1.00 |
| ≤20 | 133,178 | 13.8% | 133,128 | 14.5% | 50 | 0.1% |  | 102 | 0.1% | 52 | 0.1% | 50 | 0.1% |  |
| 21-40 | 192,959 | 19.9% | 192,731 | 21.0% | 228 | 0.5% |  | 453 | 0.5% | 225 | 0.5% | 228 | 0.5% |  |
| 41-50 | 127,794 | 13.2% | 127,139 | 13.8% | 655 | 1.3% |  | 1,311 | 1.3% | 656 | 1.3% | 655 | 1.3% |  |
| 51-60 | 172,444 | 17.8% | 166,080 | 18.1% | 6,364 | 12.7% |  | 12,732 | 12.8% | 6,368 | 12.8% | 6,364 | 12.8% |  |
| 61-70 | 159,912 | 16.5% | 143,776 | 15.7% | 16,136 | 32.2% |  | 32,265 | 32.4% | 16,129 | 32.3% | 16,136 | 32.4% |  |
| 71-80 | 120,117 | 12.4% | 102,655 | 11.2% | 17,462 | 34.9% |  | 34,693 | 34.8% | 17,352 | 34.8% | 17,341 | 34.8% |  |
| ≥81 | 61,892 | 6.4% | 52,752 | 5.7% | 9,140 | 18.3% |  | 18,168 | 18.2% | 9,080 | 18.2% | 9,088 | 18.2% |  |
| Gender |  |  |  |  |  |  | <0.001 |  |  |  |  |  |  | 0.97 |
| Female | 573,610 | 59.2% | 528,152 | 57.5% | 45,458 | 90.9% |  | 90,567 | 90.8% | 45,282 | 90.8% | 45,285 | 90.8% |  |
| Male | 394,686 | 40.8% | 390,109 | 42.5% | 4,577 | 9.1% |  | 9,157 | 9.2% | 4,580 | 9.2% | 4,577 | 9.2% |  |
| Insurance |  |  |  |  |  |  | <0.001 |  |  |  |  |  |  | 1.00 |
| Commercial | 500,918 | 51.7% | 490,503 | 53.4% | 10,415 | 20.8% |  | 20,830 | 20.9% | 10,415 | 20.9% | 10,415 | 20.9% |  |
| Dual | 6,814 | 0.7% | 5,218 | 0.6% | 1,596 | 3.2% |  | 3,154 | 3.2% | 1,581 | 3.2% | 1,573 | 3.2% |  |
| Medicaid | 252,366 | 26.1% | 243,191 | 26.5% | 9,175 | 18.3% |  | 18,044 | 18.1% | 9,019 | 18.1% | 9,025 | 18.1% |  |
| Medicare | 208,198 | 21.5% | 179,349 | 19.5% | 28,849 | 57.7% |  | 57,696 | 57.9% | 28,847 | 57.9% | 28,849 | 57.9% |  |
| PCP Visit 2019 |  |  |  |  |  |  | <0.001 |  |  |  |  |  |  | 0.89 |
| No | 521,282 | 53.8% | 502,609 | 54.7% | 18,673 | 37.3% |  | 37,253 | 37.4% | 18,616 | 37.3% | 18,637 | 37.4% |  |
| Yes | 447,014 | 46.2% | 415,652 | 45.3% | 31,362 | 62.7% |  | 62,471 | 62.6% | 31,246 | 62.7% | 31,225 | 62.6% |  |
| Continuous Outcomes |  |  |  |  |  |  |  |  |  |  |  |  |  |  |
|  | mean | SD | mean | SD | mean | SD | p-value | mean | SD | mean | SD | mean | SD | p-value |
| CCI | 0.65 | 1.39 | 0.63 | 1.37 | 0.95 | 1.68 | <0.001 | 0.95 | 1.68 | 0.95 | 1.67 | 0.95 | 1.68 | 0.93 |

BP: bisphosphonate; CCI: Charlson Comorbidity Index; PCP: primary care physician; SD: standard deviation

**Table S3a:**
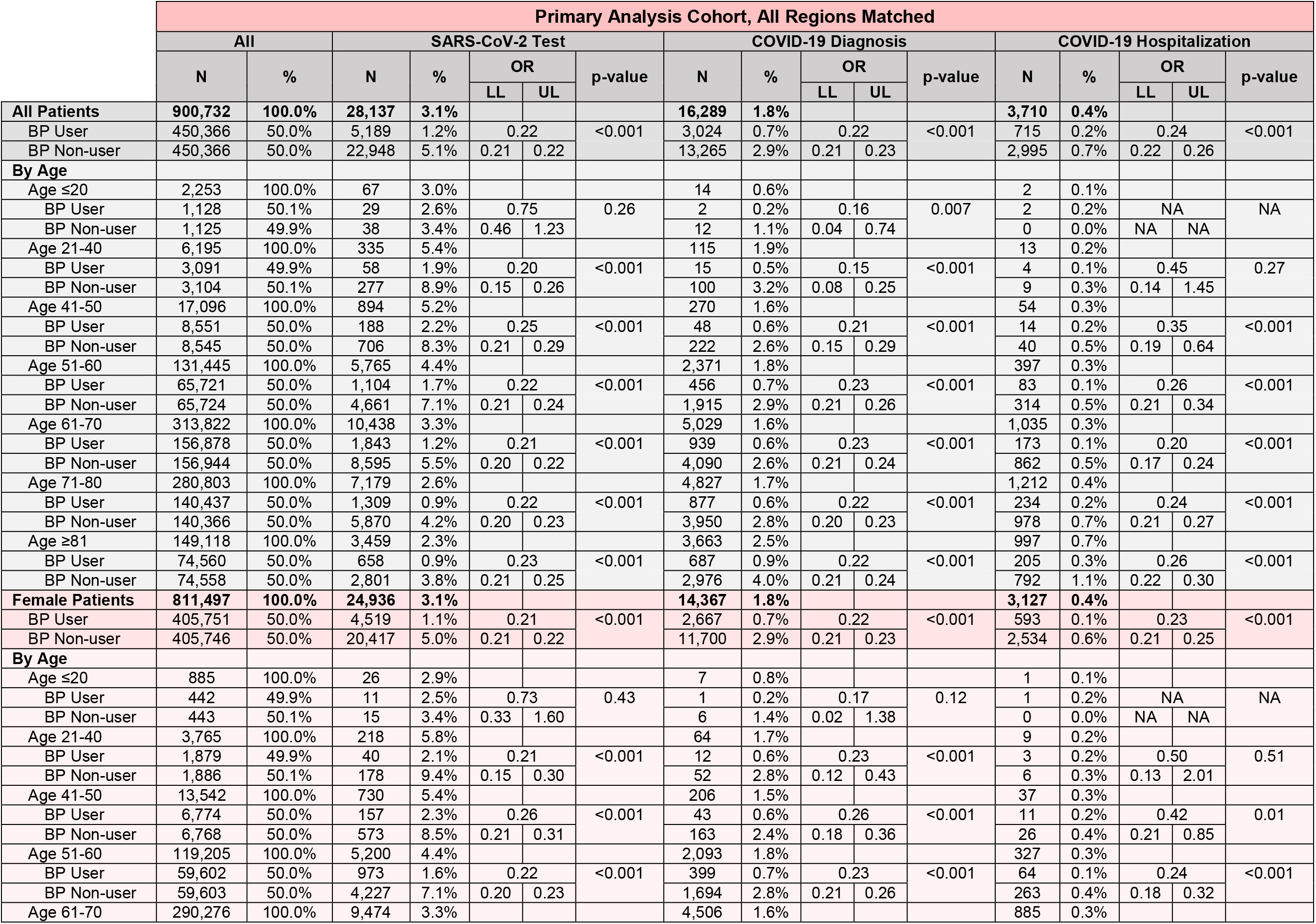
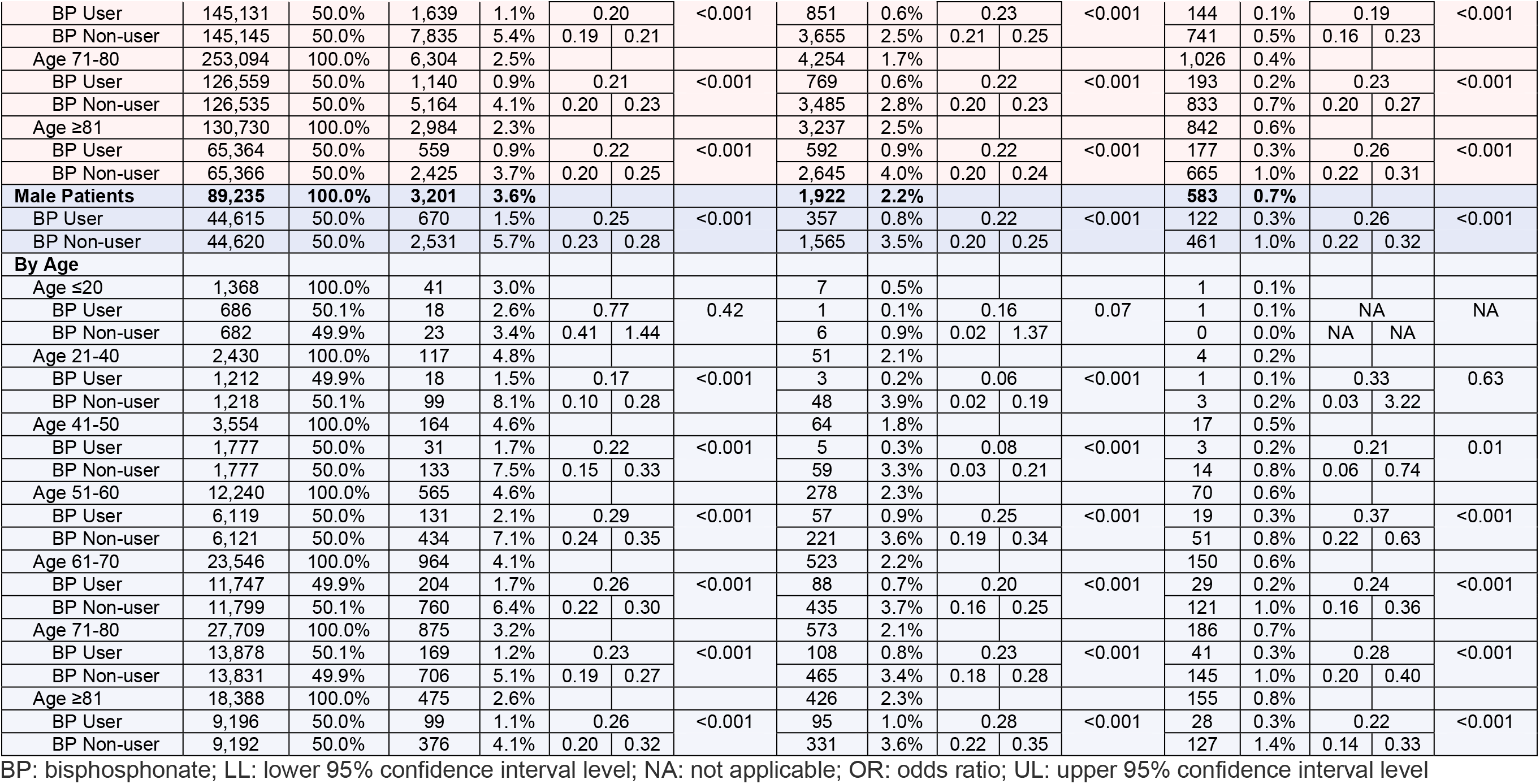
Unadjusted COVID-19-Related Outcomes Stratified by Age, Sex, & Age by Sex; Matched Primary Analysis Cohort, All-Regions Combined

**Table S3b:**
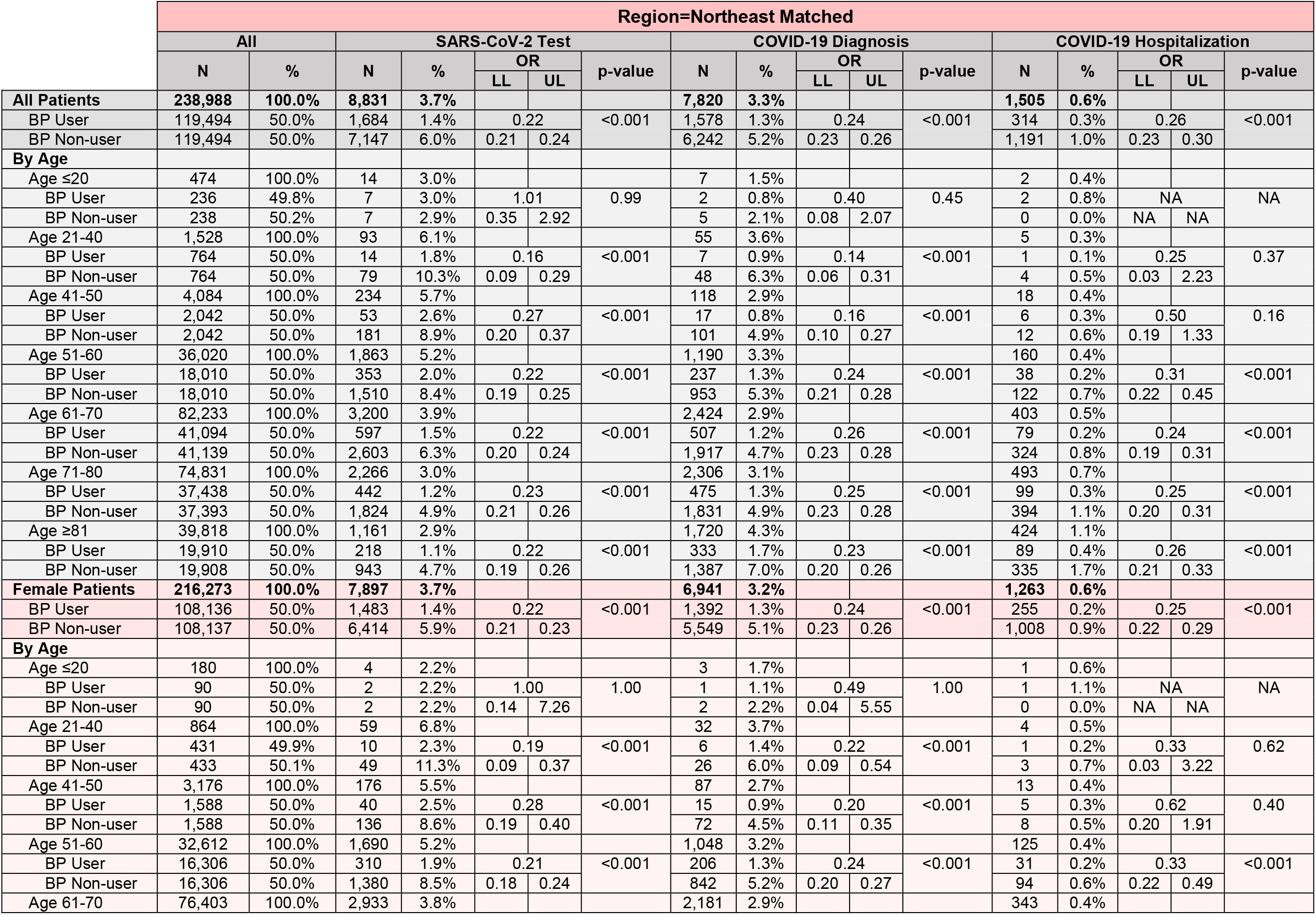
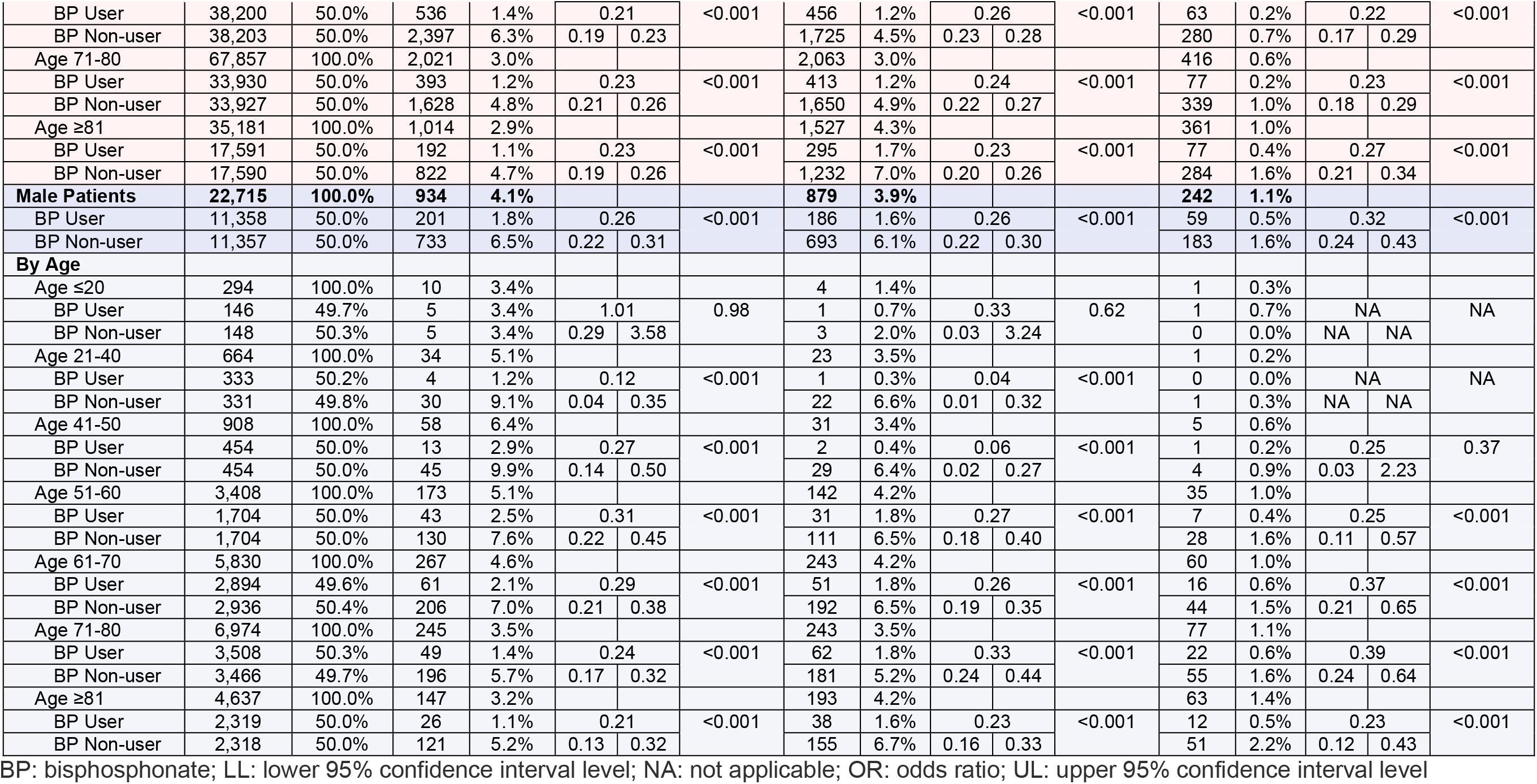
Unadjusted COVID-19-Related Outcomes Stratified by Age, Sex, & Age by Sex; Matched Primary Analysis Cohort, Region=Northeast

**Table S3c:**
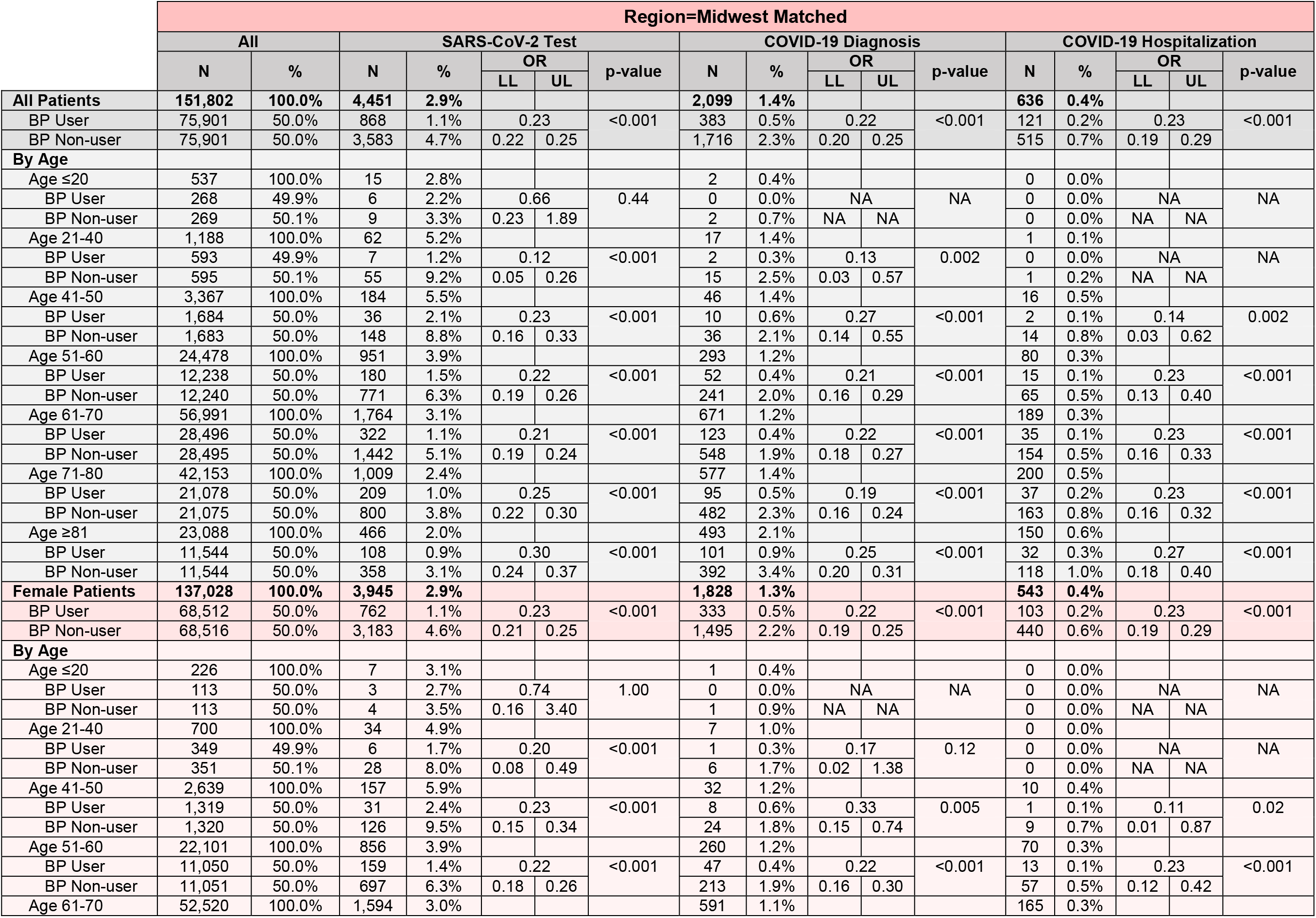
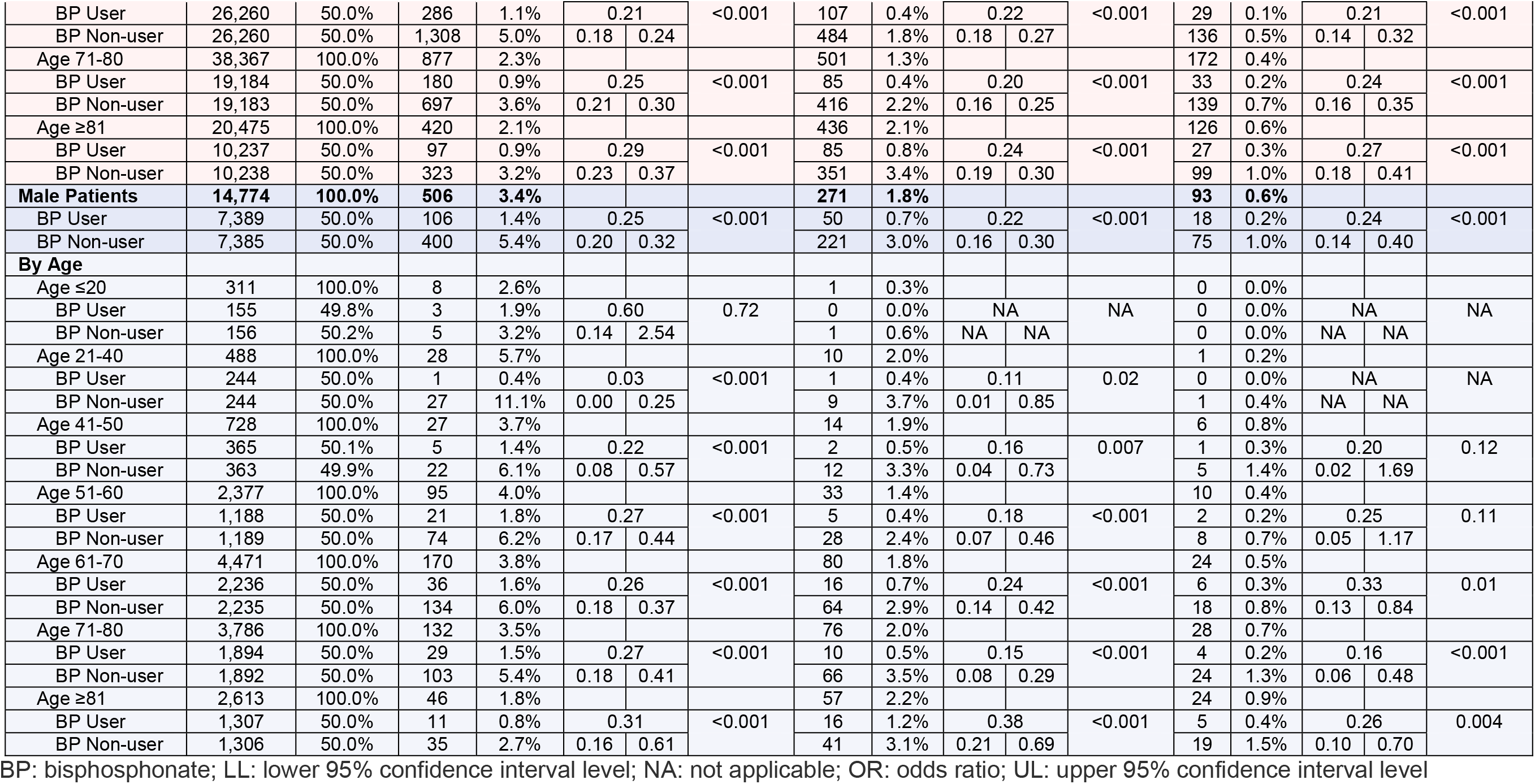
Unadjusted COVID-19-Related Outcomes Stratified by Age, Sex, & Age by Sex; Matched Primary Analysis Cohort, Region=Midwest

**Table S3d:** Unadjusted COVID-19-Related Outcomes Stratified by Age, Sex, & Age by Sex; Matched Primary Analysis Cohort, Region=South

|  | Region=South Matched |  |  |  |  |  |  |  |  |  |  |  |  |  |  |  |  |
| --- | --- | --- | --- | --- | --- | --- | --- | --- | --- | --- | --- | --- | --- | --- | --- | --- | --- |
|  | All |  | SARS-CoV-2 Test |  |  |  | p-value | COVID-19 Diagnosis |  |  |  |  | COVID-19 Hospitalization |  |  |  |  |
|  | N | % | N | % | OR |  |  | N | % | OR |  | p-value | N | % | OR |  | p-value |
|  |  |  |  |  | LL | UL |  |  |  | LL | UL |  |  |  | LL | UL |  |
| All Patients | 319,408 | 100.0% | 8,418 | 2.6% |  |  |  | 3,535 | 1.1% |  |  |  | 849 | 0.3% |  |  |  |
| BP User | 159,704 | 50.0% | 1,553 | 1.0% | 0.22 |  | <0.001 | 624 | 0.4% | 0.21 |  | <0.001 | 167 | 0.1% | 0.24 |  | <0.001 |
| BP Non-user | 159,704 | 50.0% | 6,865 | 4.3% | 0.21 | 0.23 |  | 2,911 | 1.8% | 0.19 | 0.23 |  | 682 | 0.4% | 0.21 | 0.29 |  |
| By Age |  |  |  |  |  |  |  |  |  |  |  |  |  |  |  |  |  |
| Age ≤20 | 943 | 100.0% | 29 | 3.1% |  |  |  | 4 | 0.4% |  |  |  | 0 | 0.0% |  |  |  |
| BP User | 474 | 50.3% | 15 | 3.2% | 1.06 |  | 0.87 | 0 | 0.0% | NA |  | NA | 0 | 0.0% | NA |  | NA |
| BP Non-user | 469 | 49.7% | 14 | 3.0% | 0.51 | 2.23 |  | 4 | 0.9% | NA | NA |  | 0 | 0.0% | NA | NA |  |
| Age 21-40 | 2,364 | 100.0% | 113 | 4.8% |  |  |  | 25 | 1.1% |  |  |  | 4 | 0.2% |  |  |  |
| BP User | 1,177 | 49.8% | 20 | 1.7% | 0.20 |  | <0.001 | 4 | 0.3% | 0.19 |  | <0.001 | 2 | 0.2% | 1.01 |  | 1.00 |
| BP Non-user | 1,187 | 50.2% | 93 | 7.8% | 0.12 | 0.33 |  | 21 | 1.8% | 0.06 | 0.55 |  | 2 | 0.2% | 0.14 | 7.17 |  |
| Age 41-50 | 6,839 | 100.0% | 329 | 4.8% |  |  |  | 73 | 1.1% |  |  |  | 10 | 0.1% |  |  |  |
| BP User | 3,421 | 50.0% | 72 | 2.1% | 0.26 |  | <0.001 | 18 | 0.5% | 0.32 |  | <0.001 | 5 | 0.1% | 1.00 |  | 0.99 |
| BP Non-user | 3,418 | 50.0% | 257 | 7.5% | 0.20 | 0.34 |  | 55 | 1.6% | 0.19 | 0.55 |  | 5 | 0.1% | 0.29 | 3.45 |  |
| Age 51-60 | 50,186 | 100.0% | 1,999 | 4.0% |  |  |  | 584 | 1.2% |  |  |  | 103 | 0.2% |  |  |  |
| BP User | 25,093 | 50.0% | 393 | 1.6% | 0.23 |  | <0.001 | 114 | 0.5% | 0.24 |  | <0.001 | 23 | 0.1% | 0.29 |  | <0.001 |
| BP Non-user | 25,093 | 50.0% | 1,606 | 6.4% | 0.21 | 0.26 |  | 470 | 1.9% | 0.19 | 0.29 |  | 80 | 0.3% | 0.18 | 0.46 |  |
| Age 61-70 | 111,800 | 100.0% | 3,246 | 2.9% |  |  |  | 1,106 | 1.0% |  |  |  | 247 | 0.2% |  |  |  |
| BP User | 55,896 | 50.0% | 583 | 1.0% | 0.21 |  | <0.001 | 191 | 0.3% | 0.21 |  | <0.001 | 38 | 0.1% | 0.18 |  | <0.001 |
| BP Non-user | 55,904 | 50.0% | 2,663 | 4.8% | 0.19 | 0.23 |  | 915 | 1.6% | 0.18 | 0.24 |  | 209 | 0.4% | 0.13 | 0.26 |  |
| Age 71-80 | 99,223 | 100.0% | 1,942 | 2.0% |  |  |  | 1,029 | 1.0% |  |  |  | 260 | 0.3% |  |  |  |
| BP User | 49,618 | 50.0% | 322 | 0.6% | 0.19 |  | <0.001 | 170 | 0.3% | 0.20 |  | <0.001 | 55 | 0.1% | 0.27 |  | <0.001 |
| BP Non-user | 49,605 | 50.0% | 1,620 | 3.3% | 0.17 | 0.22 |  | 859 | 1.7% | 0.17 | 0.23 |  | 205 | 0.4% | 0.20 | 0.36 |  |
| Age ≥81 | 48,053 | 100.0% | 760 | 1.6% |  |  |  | 714 | 1.5% |  |  |  | 225 | 0.5% |  |  |  |
| BP User | 24,025 | 50.0% | 148 | 0.6% | 0.24 |  | <0.001 | 127 | 0.5% | 0.21 |  | <0.001 | 44 | 0.2% | 0.24 |  | <0.001 |
| BP Non-user | 24,028 | 50.0% | 612 | 2.5% | 0.20 | 0.28 |  | 587 | 2.4% | 0.18 | 0.26 |  | 181 | 0.8% | 0.17 | 0.34 |  |
| Female Patients | 289,263 | 100.0% | 7,519 | 2.6% |  |  |  | 3,159 | 1.1% |  |  |  | 745 | 0.3% |  |  |  |
| BP User | 144,633 | 50.0% | 1,365 | 0.9% | 0.21 |  | <0.001 | 562 | 0.4% | 0.21 |  | <0.001 | 143 | 0.1% | 0.24 |  | <0.001 |
| BP Non-user | 144,630 | 50.0% | 6,154 | 4.3% | 0.20 | 0.23 |  | 2,597 | 1.8% | 0.19 | 0.23 |  | 602 | 0.4% | 0.20 | 0.28 |  |
| By Age |  |  |  |  |  |  |  |  |  |  |  |  |  |  |  |  |  |
| Age ≤20 | 372 | 100.0% | 11 | 3.0% |  |  |  | 3 | 0.8% |  |  |  | 0 | 0.0% |  |  |  |
| BP User | 185 | 49.7% | 6 | 3.2% | 1.22 |  | 0.75 | 0 | 0.0% | NA |  | NA | 0 | 0.0% | NA |  | NA |
| BP Non-user | 187 | 50.3% | 5 | 2.7% | 0.37 | 4.07 |  | 3 | 1.6% | NA | NA |  | 0 | 0.0% | NA | NA |  |
| Age 21-40 | 1,543 | 100.0% | 81 | 5.2% |  |  |  | 16 | 1.0% |  |  |  | 3 | 0.2% |  |  |  |
| BP User | 770 | 49.9% | 14 | 1.8% | 0.20 |  | <0.001 | 4 | 0.5% | 0.33 |  | 0.08 | 2 | 0.3% | 2.01 |  | 0.62 |
| BP Non-user | 773 | 50.1% | 67 | 8.7% | 0.11 | 0.35 |  | 12 | 1.6% | 0.11 | 1.03 |  | 1 | 0.1% | 0.18 | 22.22 |  |
| Age 41-50 | 5,569 | 100.0% | 273 | 4.9% |  |  |  | 66 | 1.2% |  |  |  | 9 | 0.2% |  |  |  |
| BP User | 2,787 | 50.0% | 65 | 2.3% | 0.30 |  | <0.001 | 18 | 0.6% | 0.37 |  | <0.001 | 5 | 0.2% | 1.25 |  | 1.00 |
| BP Non-user | 2,782 | 50.0% | 208 | 7.5% | 0.22 | 0.39 |  | 48 | 1.7% | 0.21 | 0.64 |  | 4 | 0.1% | 0.33 | 4.65 |  |
| Age 51-60 | 46,012 | 100.0% | 1,819 | 4.0% |  |  |  | 521 | 1.1% |  |  |  | 89 | 0.2% |  |  |  |
| BP User | 23,007 | 50.0% | 358 | 1.6% | 0.23 |  | <0.001 | 100 | 0.4% | 0.23 |  | <0.001 | 16 | 0.1% | 0.22 |  | <0.001 |
| BP Non-user | 23,005 | 50.0% | 1,461 | 6.4% | 0.21 | 0.26 |  | 421 | 1.8% | 0.19 | 0.29 |  | 73 | 0.3% | 0.13 | 0.38 |  |
| Age 61-70 | 103,825 | 100.0% | 2,948 | 2.8% |  |  |  | 1,007 | 1.0% |  |  |  | 218 | 0.2% |  |  |  |
| BP User | 51,910 | 50.0% | 517 | 1.0% | 0.20 |  | <0.001 | 177 | 0.3% | 0.21 |  | <0.001 | 33 | 0.1% | 0.18 |  | <0.001 |
| BP Non-user | 51,915 | 50.0% | 2,431 | 4.7% | 0.19 | 0.23 |  | 830 | 1.6% | 0.18 | 0.25 |  | 185 | 0.4% | 0.12 | 0.26 |  |
| Age 71-80 | 89,474 | 100.0% | 1,729 | 1.9% |  |  |  | 915 | 1.0% |  |  |  | 230 | 0.3% |  |  |  |
| BP User | 44,742 | 50.0% | 283 | 0.6% | 0.19 |  | <0.001 | 153 | 0.3% | 0.20 |  | <0.001 | 47 | 0.1% | 0.26 |  | <0.001 |
| BP Non-user | 44,732 | 50.0% | 1,446 | 3.2% | 0.17 | 0.22 |  | 762 | 1.7% | 0.17 | 0.24 |  | 183 | 0.4% | 0.19 | 0.35 |  |
| Age ≥81 | 42,468 | 100.0% | 658 | 1.5% |  |  |  | 631 | 1.5% |  |  |  | 196 | 0.5% |  |  |  |
| BP User | 21,232 | 50.0% | 122 | 0.6% | 0.22 |  | <0.001 | 110 | 0.5% | 0.21 |  | <0.001 | 40 | 0.2% | 0.26 |  | <0.001 |
| BP Non-user | 21,236 | 50.0% | 536 | 2.5% | 0.18 | 0.27 |  | 521 | 2.5% | 0.17 | 0.25 |  | 156 | 0.7% | 0.18 | 0.36 |  |
| **Male Patients** | **30,145** | **100.0%** | **899** | **3.0%** |  |  |  | **376** | **1.2%** |  |  |  | **104** | **0.3%** |  |  |  |
| BP User | 15,071 | 50.0% | 188 | 1.2% | 0.26 |  | <0.001 | 62 | 0.4% | 0.19 |  | <0.001 | 24 | 0.2% | 0.30 |  | <0.001 |
| BP Non-user | 15,074 | 50.0% | 711 | 4.7% | 0.22 | 0.30 |  | 314 | 2.1% | 0.15 | 0.26 |  | 80 | 0.5% | 0.19 | 0.47 |  |
| **By Age** |  |  |  |  |  |  |  |  |  |  |  |  |  |  |  |  |  |
| Age ≤20 | 571 | 100.0% | 18 | 3.2% |  |  |  | 1 | 0.2% |  |  |  | 0 | 0.0% |  |  |  |
| BP User | 289 | 50.6% | 9 | 3.1% | 0.98 |  | 0.96 | 0 | 0.0% | NA |  | NA | 0 | 0.0% | NA |  | NA |
| BP Non-user | 282 | 49.4% | 9 | 3.2% | 0.38 | 2.49 |  | 1 | 0.4% | NA | NA |  | 0 | 0.0% | NA | NA |  |
| Age 21-40 | 821 | 100.0% | 32 | 3.9% |  |  |  | 9 | 1.1% |  |  |  | 1 | 0.1% |  |  |  |
| BP User | 407 | 49.6% | 6 | 1.5% | 0.22 |  | <0.001 | 0 | 0.0% | NA |  | NA | 0 | 0.0% | NA |  | NA |
| BP Non-user | 414 | 50.4% | 26 | 6.3% | 0.09 | 0.55 |  | 9 | 2.2% | NA | NA |  | 1 | 0.2% | NA | NA |  |
| Age 41-50 | 1,270 | 100.0% | 56 | 4.4% |  |  |  | 7 | 0.6% |  |  |  | 1 | 0.1% |  |  |  |
| BP User | 634 | 49.9% | 7 | 1.1% | 0.13 |  | <0.001 | 0 | 0.0% | NA |  | NA | 0 | 0.0% | NA |  | NA |
| BP Non-user | 636 | 50.1% | 49 | 7.7% | 0.06 | 0.30 |  | 7 | 1.1% | NA | NA |  | 1 | 0.2% | NA | NA |  |
| Age 51-60 | 4,174 | 100.0% | 180 | 4.3% |  |  |  | 63 | 1.5% |  |  |  | 14 | 0.3% |  |  |  |
| BP User | 2,086 | 50.0% | 35 | 1.7% | 0.23 |  | <0.001 | 14 | 0.7% | 0.28 |  | <0.001 | 7 | 0.3% | 1.00 |  | 0.99 |
| BP Non-user | 2,088 | 50.0% | 145 | 6.9% | 0.16 | 0.33 |  | 49 | 2.3% | 0.15 | 0.51 |  | 7 | 0.3% | 0.35 | 2.86 |  |
| Age 61-70 | 7,975 | 100.0% | 298 | 3.7% |  |  |  | 99 | 1.2% |  |  |  | 29 | 0.4% |  |  |  |
| BP User | 3,986 | 50.0% | 66 | 1.7% | 0.27 |  | <0.001 | 14 | 0.4% | 0.16 |  | <0.001 | 5 | 0.1% | 0.21 |  | <0.001 |
| BP Non-user | 3,989 | 50.0% | 232 | 5.8% | 0.21 | 0.36 |  | 85 | 2.1% | 0.09 | 0.29 |  | 24 | 0.6% | 0.08 | 0.54 |  |
| Age 71-80 | 9,749 | 100.0% | 213 | 2.2% |  |  |  | 114 | 1.2% |  |  |  | 30 | 0.3% |  |  |  |
| BP User | 4,876 | 50.0% | 39 | 0.8% | 0.22 |  | <0.001 | 17 | 0.3% | 0.17 |  | <0.001 | 8 | 0.2% | 0.36 |  | 0.01 |
| BP Non-user | 4,873 | 50.0% | 174 | 3.6% | 0.15 | 0.31 |  | 97 | 2.0% | 0.10 | 0.29 |  | 22 | 0.5% | 0.16 | 0.81 |  |
| Age ≥81 | 5,585 | 100.0% | 102 | 1.8% |  |  |  | 83 | 1.5% |  |  |  | 29 | 0.5% |  |  |  |
| BP User | 2,793 | 50.0% | 26 | 0.9% | 0.34 |  | <0.001 | 17 | 0.6% | 0.25 |  | <0.001 | 4 | 0.1% | 0.16 |  | <0.001 |
| BP Non-user | 2,792 | 50.0% | 76 | 2.7% | 0.21 | 0.53 |  | 66 | 2.4% | 0.15 | 0.43 |  | 25 | 0.9% | 0.06 | 0.46 |  |

BP: bisphosphonate; LL: lower 95% confidence interval level; NA: not applicable; OR: odds ratio; UL: upper 95% confidence interval level

**Table S3e:**
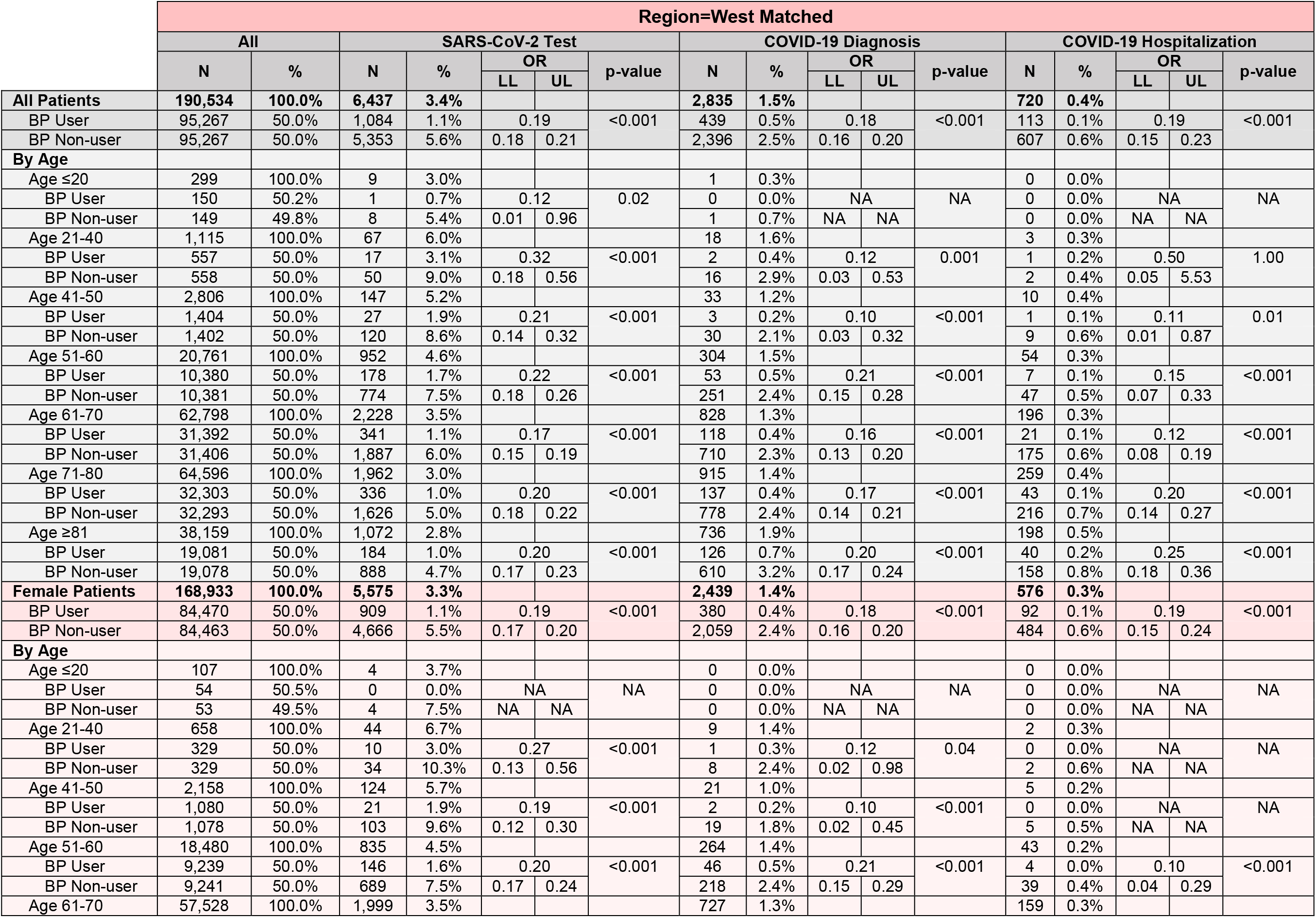
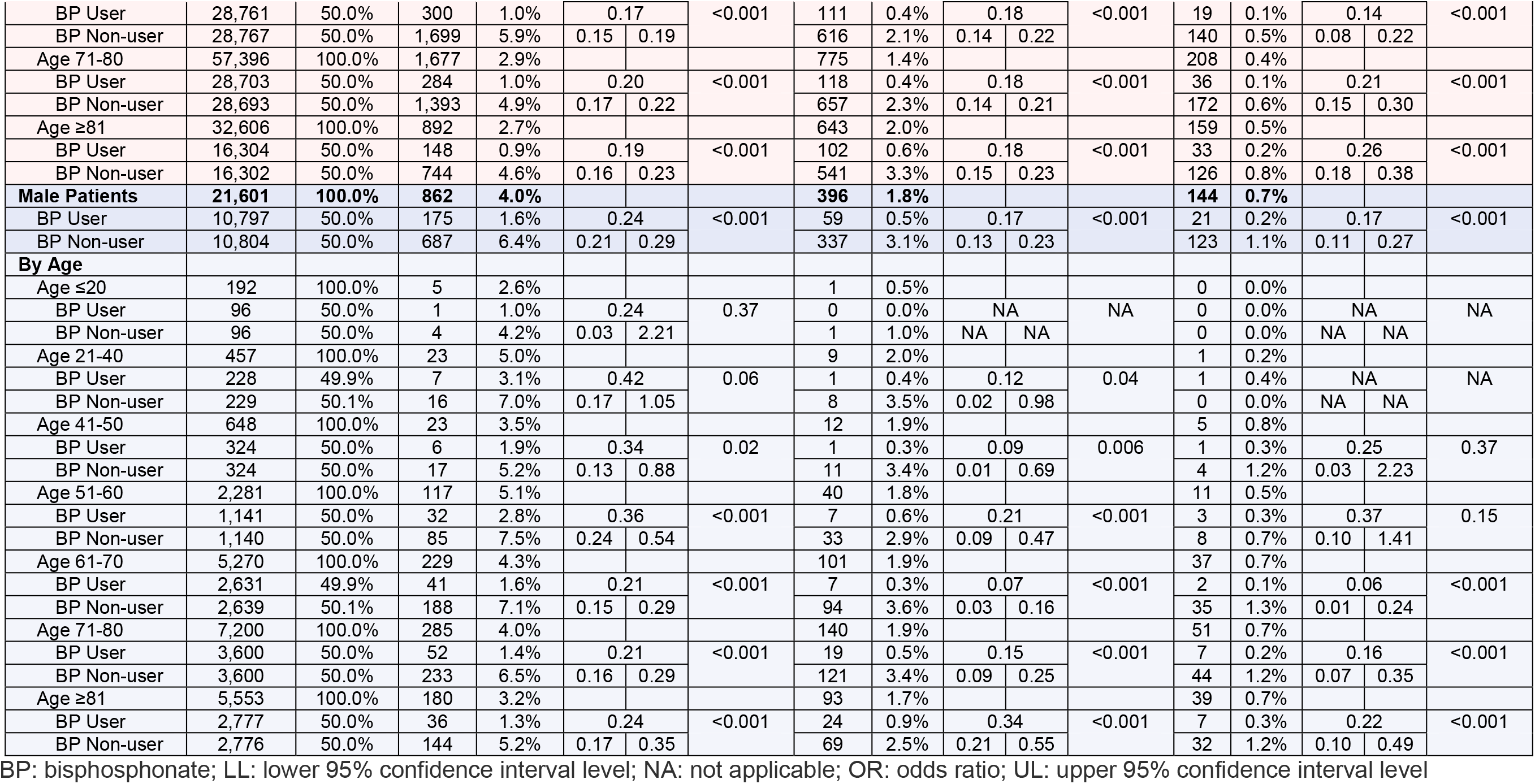
Unadjusted COVID-19-Related Outcomes Stratified by Age, Sex, & Age by Sex; Matched Primary Analysis Cohort, Region=West

**Table S3f:** Unadjusted COVID-19-Related Outcomes Stratified by Age, Sex, & Age by Sex; Matched Primary Analysis Cohort, Region=New York State

|  | Region=New York State Matched |  |  |  |  |  |  |  |  |  |  |  |  |  |  |  |  |
| --- | --- | --- | --- | --- | --- | --- | --- | --- | --- | --- | --- | --- | --- | --- | --- | --- | --- |
|  | All |  | SARS-CoV-2 Test |  |  |  |  | COVID-19 Diagnosis |  |  |  |  | COVID-19 Hospitalization |  |  |  |  |
|  | N | % | N | % | OR |  | p-value | N | % | OR |  | p-value | N | % | OR |  | p-value |
|  |  |  |  |  | LL | UL |  |  |  | LL | UL |  |  |  | LL | UL |  |
| All Patients | 99,724 | 100.0% | 3,598 | 3.6% |  |  |  | 3,607 | 3.6% |  |  |  | 622 | 0.6% |  |  |  |
| BP User | 49,862 | 50.0% | 772 | 1.5% | 0.26 |  | <0.001 | 811 | 1.6% | 0.28 |  | <0.001 | 136 | 0.3% | 0.28 |  | <0.001 |
| BP Non-user | 49,862 | 50.0% | 2,826 | 5.7% | 0.24 | 0.28 |  | 2,796 | 5.6% | 0.26 | 0.30 |  | 486 | 1.0% | 0.23 | 0.34 |  |
| By Age |  |  |  |  |  |  |  |  |  |  |  |  |  |  |  |  |  |
| Age ≤20 | 102 | 100.0% | 4 | 3.9% |  |  |  | 2 | 2.0% |  |  |  | 1 | 1.0% |  |  |  |
| BP User | 50 | 49.0% | 2 | 4.0% | 1.04 |  | 1.00 | 1 | 2.0% | 1.04 |  | 1.00 | 1 | 2.0% | NA |  | NA |
| BP Non-user | 52 | 51.0% | 2 | 3.8% | 0.14 | 7.69 |  | 1 | 1.9% | 0.06 | 17.11 |  | 0 | 0.0% | NA | NA |  |
| Age 21-40 | 453 | 100.0% | 21 | 4.6% |  |  |  | 15 | 3.3% |  |  |  | 1 | 0.2% |  |  |  |
| BP User | 228 | 50.3% | 3 | 1.3% | 0.15 |  | <0.001 | 2 | 0.9% | 0.14 |  | 0.004 | 1 | 0.4% | NA |  | NA |
| BP Non-user | 225 | 49.7% | 18 | 8.0% | 0.04 | 0.53 |  | 13 | 5.8% | 0.03 | 0.65 |  | 0 | 0.0% | NA | NA |  |
| Age 41-50 | 1,311 | 100.0% | 77 | 5.9% |  |  |  | 36 | 2.7% |  |  |  | 4 | 0.3% |  |  |  |
| BP User | 655 | 50.0% | 22 | 3.4% | 0.38 |  | <0.001 | 8 | 1.2% | 0.28 |  | <0.001 | 1 | 0.2% | 0.33 |  | 0.62 |
| BP Non-user | 656 | 50.0% | 55 | 8.4% | 0.23 | 0.63 |  | 28 | 4.3% | 0.13 | 0.61 |  | 3 | 0.5% | 0.03 | 3.21 |  |
| Age 51-60 | 12,732 | 100.0% | 688 | 5.4% |  |  |  | 527 | 4.1% |  |  |  | 58 | 0.5% |  |  |  |
| BP User | 6,364 | 50.0% | 155 | 2.4% | 0.27 |  | <0.001 | 118 | 1.9% | 0.28 |  | <0.001 | 17 | 0.3% | 0.41 |  | 0.002 |
| BP Non-user | 6,368 | 50.0% | 533 | 8.4% | 0.23 | 0.33 |  | 409 | 6.4% | 0.22 | 0.34 |  | 41 | 0.6% | 0.23 | 0.73 |  |
| Age 61-70 | 32,265 | 100.0% | 1,294 | 4.0% |  |  |  | 1,150 | 3.6% |  |  |  | 141 | 0.4% |  |  |  |
| BP User | 16,136 | 50.0% | 277 | 1.7% | 0.26 |  | <0.001 | 267 | 1.7% | 0.29 |  | <0.001 | 27 | 0.2% | 0.24 |  | <0.001 |
| BP Non-user | 16,129 | 50.0% | 1,017 | 6.3% | 0.23 | 0.30 |  | 883 | 5.5% | 0.25 | 0.33 |  | 114 | 0.7% | 0.15 | 0.36 |  |
| Age 71-80 | 34,693 | 100.0% | 957 | 2.8% |  |  |  | 1,196 | 3.4% |  |  |  | 240 | 0.7% |  |  |  |
| BP User | 17,341 | 50.0% | 204 | 1.2% | 0.26 |  | <0.001 | 257 | 1.5% | 0.26 |  | <0.001 | 45 | 0.3% | 0.23 |  | <0.001 |
| BP Non-user | 17,352 | 50.0% | 753 | 4.3% | 0.22 | 0.31 |  | 939 | 5.4% | 0.23 | 0.30 |  | 195 | 1.1% | 0.17 | 0.32 |  |
| Age ≥81 | 18,168 | 100.0% | 557 | 3.1% |  |  |  | 681 | 3.7% |  |  |  | 177 | 1.0% |  |  |  |
| BP User | 9,088 | 50.0% | 109 | 1.2% | 0.23 |  | <0.001 | 158 | 1.7% | 0.29 |  | <0.001 | 44 | 0.5% | 0.33 |  | <0.001 |
| BP Non-user | 9,080 | 50.0% | 448 | 4.9% | 0.19 | 0.29 |  | 523 | 5.8% | 0.24 | 0.35 |  | 133 | 1.5% | 0.23 | 0.46 |  |
| Female Patients | 90,567 | 100.0% | 3,255 | 3.6% |  |  |  | 3,235 | 3.6% |  |  |  | 537 | 0.6% |  |  |  |
| BP User | 45,285 | 50.0% | 687 | 1.5% | 0.26 |  | <0.001 | 726 | 1.6% | 0.28 |  | <0.001 | 108 | 0.2% | 0.25 |  | <0.001 |
| BP Non-user | 45,282 | 50.0% | 2,568 | 5.7% | 0.24 | 0.28 |  | 2,509 | 5.5% | 0.26 | 0.30 |  | 429 | 0.9% | 0.20 | 0.31 |  |
| By Age |  |  |  |  |  |  |  |  |  |  |  |  |  |  |  |  |  |
| Age ≤20 | 33 | 100.0% | 0 | 0.0% |  |  |  | 1 | 3.0% |  |  |  | 1 | 3.0% |  |  |  |
| BP User | 16 | 48.5% | 0 | 0.0% | NA |  | NA | 1 | 6.3% | NA |  | NA | 1 | 6.3% | NA |  | NA |
| BP Non-user | 17 | 51.5% | 0 | 0.0% | NA | NA |  | 0 | 0.0% | NA | NA |  | 0 | 0.0% | NA | NA |  |
| Age 21-40 | 261 | 100.0% | 16 | 6.1% |  |  |  | 8 | 3.1% |  |  |  | 1 | 0.4% |  |  |  |
| BP User | 132 | 50.6% | 2 | 1.5% | 0.13 |  | 0.002 | 2 | 1.5% | 0.32 |  | 0.17 | 1 | 0.8% | NA |  | NA |
| BP Non-user | 129 | 49.4% | 14 | 10.9% | 0.03 | 0.57 |  | 6 | 4.7% | 0.06 | 1.59 |  | 0 | 0.0% | NA | NA |  |
| Age 41-50 | 1,032 | 100.0% | 58 | 5.6% |  |  |  | 28 | 2.7% |  |  |  | 3 | 0.3% |  |  |  |
| BP User | 516 | 50.0% | 18 | 3.5% | 0.43 |  | 0.003 | 7 | 1.4% | 0.32 |  | 0.007 | 0 | 0.0% | NA |  | NA |
| BP Non-user | 516 | 50.0% | 40 | 7.8% | 0.24 | 0.76 |  | 21 | 4.1% | 0.14 | 0.77 |  | 3 | 0.6% | NA | NA |  |
| Age 51-60 | 11,699 | 100.0% | 637 | 5.4% |  |  |  | 482 | 4.1% |  |  |  | 47 | 0.4% |  |  |  |
| BP User | 5,849 | 50.0% | 138 | 2.4% | 0.26 |  | <0.001 | 110 | 1.9% | 0.28 |  | <0.001 | 14 | 0.2% | 0.42 |  | 0.006 |
| BP Non-user | 5,850 | 50.0% | 499 | 8.5% | 0.21 | 0.31 |  | 372 | 6.4% | 0.23 | 0.35 |  | 33 | 0.6% | 0.23 | 0.79 |  |
| Age 61-70 | 30,115 | 100.0% | 1,204 | 4.0% |  |  |  | 1,070 | 3.6% |  |  |  | 126 | 0.4% |  |  |  |
| BP User | 15,060 | 50.0% | 257 | 1.7% | 0.26 |  | <0.001 | 248 | 1.6% | 0.29 |  | <0.001 | 23 | 0.2% | 0.22 |  | <0.001 |
| BP Non-user | 15,055 | 50.0% | 947 | 6.3% | 0.22 | 0.30 |  | 822 | 5.5% | 0.25 | 0.33 |  | 103 | 0.7% | 0.14 | 0.35 |  |
| Age 71-80 | 31,385 | 100.0% | 858 | 2.7% |  |  |  | 1,052 | 3.4% |  |  |  | 208 | 0.7% |  |  |  |
| BP User | 15,688 | 50.0% | 176 | 1.1% | 0.25 |  | <0.001 | 221 | 1.4% | 0.26 |  | <0.001 | 33 | 0.2% | 0.19 |  | <0.001 |
| BP Non-user | 15,697 | 50.0% | 682 | 4.3% | 0.21 | 0.30 |  | 831 | 5.3% | 0.22 | 0.30 |  | 175 | 1.1% | 0.13 | 0.27 |  |
| Age ≥81 | 16,042 | 100.0% | 482 | 3.0% |  |  |  | 594 | 3.7% |  |  |  | 151 | 0.9% |  |  |  |
| BP User | 8,024 | 50.0% | 96 | 1.2% | 0.24 |  | <0.001 | 137 | 1.7% | 0.29 |  | <0.001 | 36 | 0.4% | 0.31 |  | <0.001 |
| BP Non-user | 8,018 | 50.0% | 386 | 4.8% | 0.19 | 0.30 |  | 457 | 5.7% | 0.24 | 0.35 |  | 115 | 1.4% | 0.21 | 0.45 |  |
| **Male Patients** | **9,157** | **100.0%** | **343** | **3.7%** |  |  |  | **372** | **4.1%** |  |  |  | **85** | **0.9%** |  |  |  |
| BP User | 4,577 | 50.0% | 85 | 1.9% | 0.32 |  | <0.001 | 85 | 1.9% | 0.28 |  | <0.001 | 28 | 0.6% | 0.49 |  | 0.002 |
| BP Non-user | 4,580 | 50.0% | 258 | 5.6% | 0.25 | 0.41 |  | 287 | 6.3% | 0.22 | 0.36 |  | 57 | 1.2% | 0.31 | 0.77 |  |
| **By Age** |  |  |  |  |  |  |  |  |  |  |  |  |  |  |  |  |  |
| Age ≤20 | 69 | 100.0% | 4 | 5.8% |  |  |  | 1 | 1.4% |  |  |  | 0 | 0.0% |  |  |  |
| BP User | 34 | 49.3% | 2 | 5.9% | 1.03 |  | 1.00 | 0 | 0.0% | NA |  | NA | 0 | 0.0% | NA |  | NA |
| BP Non-user | 35 | 50.7% | 2 | 5.7% | 0.14 | 7.77 |  | 1 | 2.9% | NA | NA |  | 0 | 0.0% | NA | NA |  |
| Age 21-40 | 192 | 100.0% | 5 | 2.6% |  |  |  | 7 | 3.6% |  |  |  | 0 | 0.0% |  |  |  |
| BP User | 96 | 50.0% | 1 | 1.0% | 0.24 |  | 0.37 | 0 | 0.0% | NA |  | NA | 0 | 0.0% | NA |  | NA |
| BP Non-user | 96 | 50.0% | 4 | 4.2% | 0.03 | 2.21 |  | 7 | 7.3% | NA | NA |  | 0 | 0.0% | NA | NA |  |
| Age 41-50 | 279 | 100.0% | 19 | 6.8% |  |  |  | 8 | 2.9% |  |  |  | 1 | 0.4% |  |  |  |
| BP User | 139 | 49.8% | 4 | 2.9% | 0.25 |  | 0.02 | 1 | 0.7% | 0.14 |  | 0.07 | 1 | 0.7% | NA |  | NA |
| BP Non-user | 140 | 50.2% | 15 | 10.7% | 0.08 | 0.76 |  | 7 | 5.0% | 0.02 | 1.13 |  | 0 | 0.0% | NA | NA |  |
| Age 51-60 | 1,033 | 100.0% | 51 | 4.9% |  |  |  | 45 | 4.4% |  |  |  | 11 | 1.1% |  |  |  |
| BP User | 515 | 49.9% | 17 | 3.3% | 0.49 |  | 0.02 | 8 | 1.6% | 0.21 |  | <0.001 | 3 | 0.6% | 0.37 |  | 0.22 |
| BP Non-user | 518 | 50.1% | 34 | 6.6% | 0.27 | 0.88 |  | 37 | 7.1% | 0.09 | 0.44 |  | 8 | 1.5% | 0.10 | 1.42 |  |
| Age 61-70 | 2,150 | 100.0% | 90 | 4.2% |  |  |  | 80 | 3.7% |  |  |  | 15 | 0.7% |  |  |  |
| BP User | 1,076 | 50.0% | 20 | 1.9% | 0.27 |  | <0.001 | 19 | 1.8% | 0.30 |  | <0.001 | 4 | 0.4% | 0.36 |  | 0.08 |
| BP Non-user | 1,074 | 50.0% | 70 | 6.5% | 0.16 | 0.45 |  | 61 | 5.7% | 0.18 | 0.50 |  | 11 | 1.0% | 0.11 | 1.14 |  |
| Age 71-80 | 3,308 | 100.0% | 99 | 3.0% |  |  |  | 144 | 4.4% |  |  |  | 32 | 1.0% |  |  |  |
| BP User | 1,653 | 50.0% | 28 | 1.7% | 0.38 |  | <0.001 | 36 | 2.2% | 0.32 |  | <0.001 | 12 | 0.7% | 0.60 |  | 0.16 |
| BP Non-user | 1,655 | 50.0% | 71 | 4.3% | 0.25 | 0.60 |  | 108 | 6.5% | 0.22 | 0.47 |  | 20 | 1.2% | 0.29 | 1.23 |  |
| Age ≥81 | 2,126 | 100.0% | 75 | 3.5% |  |  |  | 87 | 4.1% |  |  |  | 26 | 1.2% |  |  |  |
| BP User | 1,064 | 50.0% | 13 | 1.2% | 0.20 |  | <0.001 | 21 | 2.0% | 0.30 |  | <0.001 | 8 | 0.8% | 0.44 |  | 0.05 |
| BP Non-user | 1,062 | 50.0% | 62 | 5.8% | 0.11 | 0.37 |  | 66 | 6.2% | 0.18 | 0.50 |  | 18 | 1.7% | 0.19 | 1.02 |  |

BP: bisphosphonate; LL: lower 95% confidence interval level; NA: not applicable; OR: odds ratio; UL: upper 95% confidence interval level

**Table S4:**
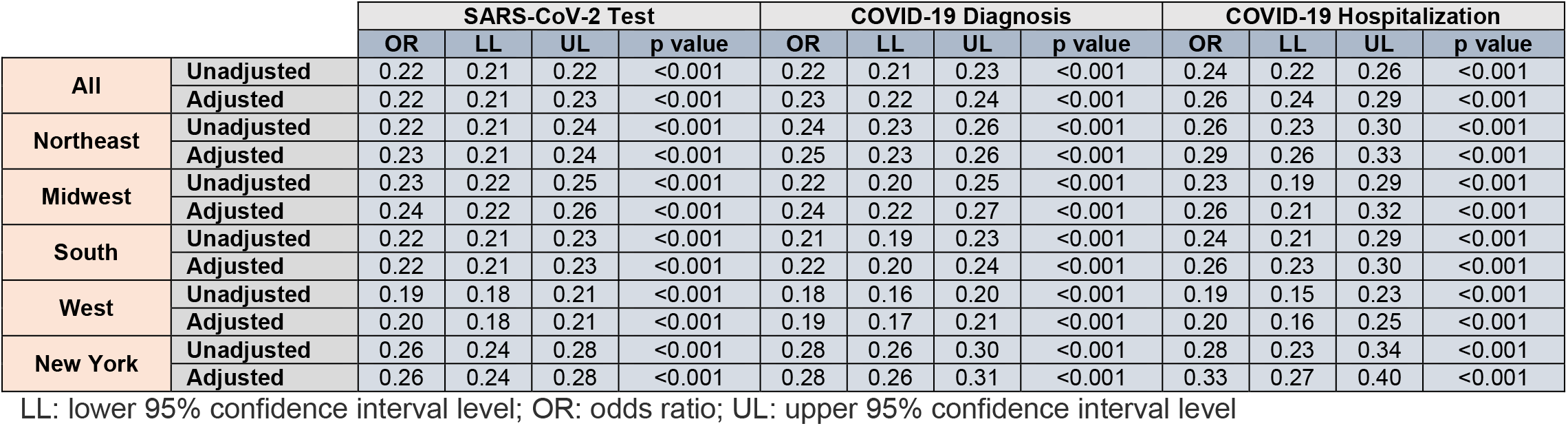
‘*Bone-Rx*’ Cohort Unadjusted/Adjusted Odds Ratio for COVID-19-Related Outcomes, Stratified by Region and New York State

**Table S5a:** Statin Use Sensitivity Analysis, Unadjusted/Adjusted Odds Ratio for COVID-19-Related Outcomes, Stratified by Region and New York State

|  |  | SARS-CoV-2 Test |  |  |  | COVID-19 Diagnosis |  |  |  | COVID-19 Hospitalization |  |  |  |
| --- | --- | --- | --- | --- | --- | --- | --- | --- | --- | --- | --- | --- | --- |
|  |  | Statin Uses versus Non-users |  |  |  |  |  |  |  |  |  |  |  |
|  |  | OR | LL | UL | p value | OR | LL | UL | p value | OR | LL | UL | p value |
| All | Unadjusted | 0.90 | 0.89 | 0.91 | <0.001 | 0.91 | 0.90 | 0.92 | <0.001 | 1.12 | 1.09 | 1.15 | <0.001 |
|  | Adjusted | 0.87 | 0.86 | 0.87 | <0.001 | 0.79 | 0.78 | 0.81 | <0.001 | 0.99 | 0.96 | 1.02 | 0.48 |
| Northeast | Unadjusted | 0.87 | 0.85 | 0.88 | <0.001 | 0.88 | 0.86 | 0.90 | <0.001 | 1.16 | 1.11 | 1.21 | <0.001 |
|  | Adjusted | 0.85 | 0.84 | 0.87 | <0.001 | 0.77 | 0.75 | 0.78 | <0.001 | 1.03 | 0.98 | 1.07 | 0.22 |
| Midwest | Unadjusted | 0.97 | 0.95 | 0.99 | 0.02 | 1.10 | 1.07 | 1.14 | <0.001 | 1.27 | 1.19 | 1.36 | <0.001 |
|  | Adjusted | 0.92 | 0.90 | 0.94 | <0.001 | 0.99 | 0.96 | 1.03 | 0.75 | 1.15 | 1.08 | 1.23 | <0.001 |
| South | Unadjusted | 0.90 | 0.88 | 0.91 | <0.001 | 0.90 | 0.88 | 0.93 | <0.001 | 1.00 | 0.95 | 1.06 | 0.90 |
|  | Adjusted | 0.85 | 0.84 | 0.87 | <0.001 | 0.80 | 0.78 | 0.83 | <0.001 | 0.88 | 0.83 | 0.94 | <0.001 |
| West | Unadjusted | 0.88 | 0.86 | 0.90 | <0.001 | 0.83 | 0.80 | 0.86 | <0.001 | 1.02 | 0.95 | 1.10 | 0.58 |
|  | Adjusted | 0.86 | 0.83 | 0.88 | <0.001 | 0.71 | 0.68 | 0.74 | <0.001 | 0.87 | 0.80 | 0.94 | <0.001 |
| New York | Unadjusted | 0.91 | 0.89 | 0.93 | <0.001 | 0.93 | 0.91 | 0.96 | <0.001 | 1.21 | 1.14 | 1.29 | <0.001 |
|  | Adjusted | 0.92 | 0.90 | 0.95 | <0.001 | 0.79 | 0.77 | 0.82 | <0.001 | 1.05 | 0.98 | 1.13 | 0.15 |
|  |  | BP Users versus BP Non-users among Statin Users |  |  |  |  |  |  |  |  |  |  |  |
|  |  | OR | LL | UL | p value | OR | LL | UL | p value | OR | LL | UL | p value |
| All | Unadjusted | 0.23 | 0.22 | 0.24 | <0.001 | 0.26 | 0.25 | 0.28 | <0.001 | 0.26 | 0.23 | 0.29 | <0.001 |
|  | Adjusted | 0.23 | 0.22 | 0.24 | <0.001 | 0.27 | 0.25 | 0.29 | <0.001 | 0.28 | 0.25 | 0.32 | <0.001 |
| Northeast | Unadjusted | 0.25 | 0.23 | 0.27 | <0.001 | 0.29 | 0.27 | 0.31 | <0.001 | 0.28 | 0.24 | 0.34 | <0.001 |
|  | Adjusted | 0.25 | 0.23 | 0.27 | <0.001 | 0.29 | 0.27 | 0.32 | <0.001 | 0.32 | 0.26 | 0.38 | <0.001 |
| Midwest | Unadjusted | 0.24 | 0.22 | 0.27 | <0.001 | 0.22 | 0.19 | 0.25 | <0.001 | 0.21 | 0.16 | 0.27 | <0.001 |
|  | Adjusted | 0.25 | 0.23 | 0.29 | <0.001 | 0.23 | 0.22 | 0.25 | <0.001 | 0.22 | 0.17 | 0.30 | <0.001 |
| South | Unadjusted | 0.22 | 0.21 | 0.24 | <0.001 | 0.26 | 0.23 | 0.29 | <0.001 | 0.26 | 0.21 | 0.33 | <0.001 |
|  | Adjusted | 0.22 | 0.20 | 0.24 | <0.001 | 0.27 | 0.24 | 0.31 | <0.001 | 0.28 | 0.22 | 0.36 | <0.001 |
| West | Unadjusted | 0.20 | 0.18 | 0.22 | <0.001 | 0.22 | 0.19 | 0.25 | <0.001 | 0.25 | 0.20 | 0.33 | <0.001 |
|  | Adjusted | 0.20 | 0.18 | 0.22 | <0.001 | 0.23 | 0.20 | 0.27 | <0.001 | 0.28 | 0.21 | 0.36 | <0.001 |
| New York | Unadjusted | 0.27 | 0.24 | 0.30 | <0.001 | 0.31 | 0.28 | 0.35 | <0.001 | 0.30 | 0.23 | 0.39 | <0.001 |
|  | Adjusted | 0.28 | 0.25 | 0.32 | <0.001 | 0.31 | 0.28 | 0.35 | <0.001 | 0.33 | 0.25 | 0.44 | <0.001 |
|  |  | BP Users versus BP Non-users among Statin Non-users |  |  |  |  |  |  |  |  |  |  |  |
|  |  | OR | LL | UL | p value | OR | LL | UL | p value | OR | LL | UL | p value |
| All | Unadjusted | 0.23 | 0.21 | 0.24 | <0.001 | 0.21 | 0.19 | 0.23 | <0.001 | 0.21 | 0.17 | 0.25 | <0.001 |
|  | Adjusted | 0.24 | 0.22 | 0.25 | <0.001 | 0.23 | 0.21 | 0.25 | <0.001 | 0.25 | 0.21 | 0.30 | <0.001 |
| Northeast | Unadjusted | 0.25 | 0.22 | 0.27 | <0.001 | 0.22 | 0.20 | 0.25 | <0.001 | 0.24 | 0.19 | 0.31 | <0.001 |
|  | Adjusted | 0.26 | 0.23 | 0.29 | <0.001 | 0.25 | 0.22 | 0.28 | <0.001 | 0.29 | 0.22 | 0.37 | <0.001 |
| Midwest | Unadjusted | 0.24 | 0.21 | 0.28 | <0.001 | 0.22 | 0.18 | 0.27 | <0.001 | 0.21 | 0.14 | 0.31 | <0.001 |
|  | Adjusted | 0.24 | 0.20 | 0.28 | <0.001 | 0.25 | 0.20 | 0.32 | <0.001 | 0.26 | 0.17 | 0.39 | <0.001 |
| South | Unadjusted | 0.23 | 0.21 | 0.25 | <0.001 | 0.19 | 0.15 | 0.22 | <0.001 | 0.18 | 0.12 | 0.27 | <0.001 |
|  | Adjusted | 0.24 | 0.21 | 0.27 | <0.001 | 0.21 | 0.17 | 0.25 | <0.001 | 0.22 | 0.15 | 0.33 | <0.001 |
| West | Unadjusted | 0.19 | 0.17 | 0.22 | <0.001 | 0.18 | 0.15 | 0.22 | <0.001 | 0.16 | 0.11 | 0.25 | <0.001 |
|  | Adjusted | 0.20 | 0.17 | 0.23 | <0.001 | 0.19 | 0.18 | 0.21 | <0.001 | 0.18 | 0.11 | 0.29 | <0.001 |
| New York | Unadjusted | 0.26 | 0.23 | 0.30 | <0.001 | 0.26 | 0.22 | 0.30 | <0.001 | 0.27 | 0.19 | 0.39 | <0.001 |
|  | Adjusted | 0.26 | 0.22 | 0.31 | <0.001 | 0.25 | 0.21 | 0.30 | <0.001 | 0.35 | 0.23 | 0.52 | <0.001 |

LL: lower 95% confidence interval level; OR: odds ratio; UL: upper 95% confidence interval level

**Table S5b:** Antihypertensive Use Sensitivity Analysis, Unadjusted/Adjusted Odds Ratio for COVID-19-Related Outcomes, Stratified by Region and New York State

|  |  | Odds of SARS-CoV-2 Test |  |  |  | Odds of COVID-19 Diagnosis |  |  |  | Odds of COVID-19 Hospitalization |  |  |  |
| --- | --- | --- | --- | --- | --- | --- | --- | --- | --- | --- | --- | --- | --- |
|  |  | Antihypertensive Users versus Non-users |  |  |  |  |  |  |  |  |  |  |  |
|  |  | OR | LL | UL | p value | OR | LL | UL | p value | OR | LL | UL | p value |
| All | Unadjusted | 0.91 | 0.90 | 0.92 | <0.001 | 0.86 | 0.85 | 0.87 | <0.001 | 1.13 | 1.10 | 1.17 | <0.001 |
|  | Adjusted | 0.87 | 0.86 | 0.88 | <0.001 | 0.75 | 0.74 | 0.76 | <0.001 | 0.98 | 0.95 | 1.00 | 0.10 |
| Northeast | Unadjusted | 0.86 | 0.84 | 0.87 | <0.001 | 0.83 | 0.82 | 0.85 | <0.001 | 1.20 | 1.15 | 1.25 | <0.001 |
|  | Adjusted | 0.82 | 0.81 | 0.83 | <0.001 | 0.72 | 0.71 | 0.73 | <0.001 | 1.04 | 0.99 | 1.08 | 0.10 |
| Midwest | Unadjusted | 1.00 | 0.98 | 1.02 | 0.98 | 1.06 | 1.03 | 1.10 | <0.001 | 1.28 | 1.20 | 1.36 | <0.001 |
|  | Adjusted | 0.94 | 0.91 | 0.96 | <0.001 | 0.94 | 0.90 | 0.97 | <0.001 | 1.11 | 1.04 | 1.19 | 0.002 |
| South | Unadjusted | 0.93 | 0.92 | 0.94 | <0.001 | 0.88 | 0.86 | 0.90 | <0.001 | 1.02 | 0.96 | 1.07 | 0.58 |
|  | Adjusted | 0.88 | 0.87 | 0.89 | <0.001 | 0.78 | 0.76 | 0.80 | <0.001 | 0.89 | 0.84 | 0.94 | <0.001 |
| West | Unadjusted | 0.90 | 0.88 | 0.92 | <0.001 | 0.75 | 0.73 | 0.78 | <0.001 | 0.99 | 0.92 | 1.06 | 0.83 |
|  | Adjusted | 0.87 | 0.85 | 0.89 | <0.001 | 0.65 | 0.62 | 0.67 | <0.001 | 0.84 | 0.78 | 0.90 | <0.001 |
| New York | Unadjusted | 0.92 | 0.90 | 0.94 | <0.001 | 0.90 | 0.87 | 0.92 | <0.001 | 1.23 | 1.15 | 1.31 | <0.001 |
|  | Adjusted | 0.90 | 0.87 | 0.92 | <0.001 | 0.75 | 0.73 | 0.77 | <0.001 | 1.01 | 0.95 | 1.09 | 0.70 |
|  |  | BP Users versus BP Non-users among Antihypertensive Users |  |  |  |  |  |  |  |  |  |  |  |
|  |  | OR | LL | UL | p value | OR | LL | UL | p value | OR | LL | UL | p value |
| All | Unadjusted | 0.23 | 0.22 | 0.24 | <0.001 | 0.26 | 0.25 | 0.28 | <0.001 | 0.26 | 0.23 | 0.29 | <0.001 |
|  | Adjusted | 0.23 | 0.22 | 0.24 | <0.001 | 0.26 | 0.25 | 0.28 | <0.001 | 0.27 | 0.24 | 0.30 | <0.001 |
| Northeast | Unadjusted | 0.24 | 0.22 | 0.26 | <0.001 | 0.28 | 0.26 | 0.31 | <0.001 | 0.27 | 0.22 | 0.32 | <0.001 |
|  | Adjusted | 0.23 | 0.21 | 0.26 | <0.001 | 0.28 | 0.26 | 0.31 | <0.001 | 0.29 | 0.24 | 0.34 | <0.001 |
| Midwest | Unadjusted | 0.26 | 0.23 | 0.29 | <0.001 | 0.27 | 0.23 | 0.31 | <0.001 | 0.27 | 0.21 | 0.35 | <0.001 |
|  | Adjusted | 0.27 | 0.24 | 0.30 | <0.001 | 0.28 | 0.26 | 0.30 | <0.001 | 0.27 | 0.20 | 0.35 | <0.001 |
| South | Unadjusted | 0.23 | 0.21 | 0.25 | <0.001 | 0.24 | 0.22 | 0.28 | <0.001 | 0.26 | 0.20 | 0.32 | <0.001 |
|  | Adjusted | 0.23 | 0.21 | 0.25 | <0.001 | 0.24 | 0.21 | 0.28 | <0.001 | 0.25 | 0.20 | 0.32 | <0.001 |
| West | Unadjusted | 0.20 | 0.18 | 0.22 | <0.001 | 0.21 | 0.18 | 0.25 | <0.001 | 0.24 | 0.18 | 0.31 | <0.001 |
|  | Adjusted | 0.20 | 0.18 | 0.22 | <0.001 | 0.22 | 0.18 | 0.25 | <0.001 | 0.24 | 0.18 | 0.33 | <0.001 |
| New York | Unadjusted | 0.26 | 0.23 | 0.29 | <0.001 | 0.30 | 0.26 | 0.33 | <0.001 | 0.29 | 0.22 | 0.38 | <0.001 |
|  | Adjusted | 0.25 | 0.22 | 0.29 | <0.001 | 0.30 | 0.26 | 0.34 | <0.001 | 0.33 | 0.24 | 0.44 | <0.001 |
|  |  | BP Users versus BP Non-users among Antihypertensive Non-users |  |  |  |  |  |  |  |  |  |  |  |
|  |  | OR | LL | UL | p value | OR | LL | UL | p value | OR | LL | UL | p value |
| All | Unadjusted | 0.21 | 0.20 | 0.22 | <0.001 | 0.20 | 0.18 | 0.22 | <0.001 | 0.21 | 0.17 | 0.25 | <0.001 |
|  | Adjusted | 0.21 | 0.20 | 0.22 | <0.001 | 0.22 | 0.20 | 0.24 | <0.001 | 0.27 | 0.22 | 0.32 | <0.001 |
| Northeast | Unadjusted | 0.21 | 0.19 | 0.23 | <0.001 | 0.22 | 0.19 | 0.24 | <0.001 | 0.23 | 0.18 | 0.31 | <0.001 |
|  | Adjusted | 0.22 | 0.20 | 0.25 | <0.001 | 0.25 | 0.22 | 0.28 | <0.001 | 0.30 | 0.22 | 0.40 | <0.001 |
| Midwest | Unadjusted | 0.22 | 0.19 | 0.25 | <0.001 | 0.16 | 0.12 | 0.20 | <0.001 | 0.20 | 0.13 | 0.31 | <0.001 |
|  | Adjusted | 0.21 | 0.18 | 0.25 | <0.001 | 0.18 | 0.14 | 0.23 | <0.001 | 0.26 | 0.16 | 0.42 | <0.001 |
| South | Unadjusted | 0.20 | 0.18 | 0.22 | <0.001 | 0.19 | 0.16 | 0.22 | <0.001 | 0.22 | 0.15 | 0.32 | <0.001 |
|  | Adjusted | 0.20 | 0.18 | 0.22 | <0.001 | 0.21 | 0.17 | 0.25 | <0.001 | 0.28 | 0.19 | 0.41 | <0.001 |
| West | Unadjusted | 0.19 | 0.17 | 0.22 | <0.001 | 0.18 | 0.15 | 0.22 | <0.001 | 0.15 | 0.09 | 0.24 | <0.001 |
|  | Adjusted | 0.20 | 0.17 | 0.22 | <0.001 | 0.20 | 0.16 | 0.25 | <0.001 | 0.19 | 0.11 | 0.32 | <0.001 |
| New York | Unadjusted | 0.26 | 0.23 | 0.31 | <0.001 | 0.25 | 0.21 | 0.29 | <0.001 | 0.23 | 0.15 | 0.36 | <0.001 |
|  | Adjusted | 0.27 | 0.23 | 0.32 | <0.001 | 0.26 | 0.22 | 0.31 | <0.001 | 0.26 | 0.16 | 0.43 | <0.001 |

LL: lower 95% confidence interval level; OR: odds ratio; UL: upper 95% confidence interval level

**Table S5c:**
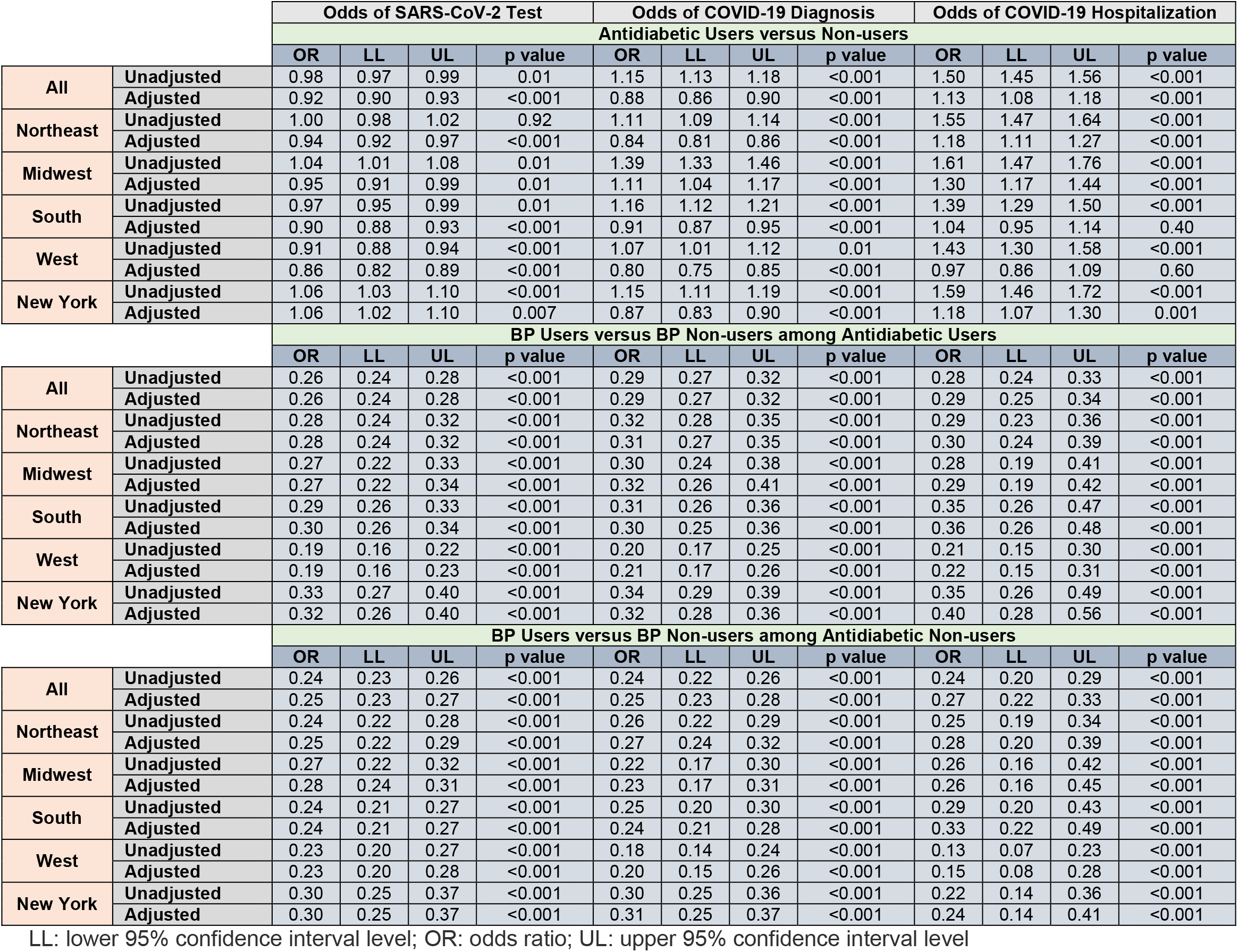
Antidiabetic Use Sensitivity Analysis, Unadjusted/Adjusted Odds Ratio for COVID-19-Related Outcomes, Stratified by Region and New York State

|  |  | Odds of SARS-CoV-2 Test |  |  |  | Odds of COVID-19 Diagnosis |  |  |  | Odds of COVID-19 Hospitalization |  |  |  |
| --- | --- | --- | --- | --- | --- | --- | --- | --- | --- | --- | --- | --- | --- |
|  |  | Antidiabetic Users versus Non-users |  |  |  |  |  |  |  |  |  |  |  |
|  |  | OR | LL | UL | p value | OR | LL | UL | p value | OR | LL | UL | p value |
| All | Unadjusted | 0.98 | 0.97 | 0.99 | 0.01 | 1.15 | 1.13 | 1.18 | <0.001 | 1.50 | 1.45 | 1.56 | <0.001 |
|  | Adjusted | 0.92 | 0.90 | 0.93 | <0.001 | 0.88 | 0.86 | 0.90 | <0.001 | 1.13 | 1.08 | 1.18 | <0.001 |
| Northeast | Unadjusted | 1.00 | 0.98 | 1.02 | 0.92 | 1.11 | 1.09 | 1.14 | <0.001 | 1.55 | 1.47 | 1.64 | <0.001 |
|  | Adjusted | 0.94 | 0.92 | 0.97 | <0.001 | 0.84 | 0.81 | 0.86 | <0.001 | 1.18 | 1.11 | 1.27 | <0.001 |
| Midwest | Unadjusted | 1.04 | 1.01 | 1.08 | 0.01 | 1.39 | 1.33 | 1.46 | <0.001 | 1.61 | 1.47 | 1.76 | <0.001 |
|  | Adjusted | 0.95 | 0.91 | 0.99 | 0.01 | 1.11 | 1.04 | 1.17 | <0.001 | 1.30 | 1.17 | 1.44 | <0.001 |
| South | Unadjusted | 0.97 | 0.95 | 0.99 | 0.01 | 1.16 | 1.12 | 1.21 | <0.001 | 1.39 | 1.29 | 1.50 | <0.001 |
|  | Adjusted | 0.90 | 0.88 | 0.93 | <0.001 | 0.91 | 0.87 | 0.95 | <0.001 | 1.04 | 0.95 | 1.14 | 0.40 |
| West | Unadjusted | 0.91 | 0.88 | 0.94 | <0.001 | 1.07 | 1.01 | 1.12 | 0.01 | 1.43 | 1.30 | 1.58 | <0.001 |
|  | Adjusted | 0.86 | 0.82 | 0.89 | <0.001 | 0.80 | 0.75 | 0.85 | <0.001 | 0.97 | 0.86 | 1.09 | 0.60 |
| New York | Unadjusted | 1.06 | 1.03 | 1.10 | <0.001 | 1.15 | 1.11 | 1.19 | <0.001 | 1.59 | 1.46 | 1.72 | <0.001 |
|  | Adjusted | 1.06 | 1.02 | 1.10 | 0.007 | 0.87 | 0.83 | 0.90 | <0.001 | 1.18 | 1.07 | 1.30 | 0.001 |
|  |  | BP Users versus BP Non-users among Antidiabetic Users |  |  |  |  |  |  |  |  |  |  |  |
|  |  | OR | LL | UL | p value | OR | LL | UL | p value | OR | LL | UL | p value |
| All | Unadjusted | 0.26 | 0.24 | 0.28 | <0.001 | 0.29 | 0.27 | 0.32 | <0.001 | 0.28 | 0.24 | 0.33 | <0.001 |
|  | Adjusted | 0.26 | 0.24 | 0.28 | <0.001 | 0.29 | 0.27 | 0.32 | <0.001 | 0.29 | 0.25 | 0.34 | <0.001 |
| Northeast | Unadjusted | 0.28 | 0.24 | 0.32 | <0.001 | 0.32 | 0.28 | 0.35 | <0.001 | 0.29 | 0.23 | 0.36 | <0.001 |
|  | Adjusted | 0.28 | 0.24 | 0.32 | <0.001 | 0.31 | 0.27 | 0.35 | <0.001 | 0.30 | 0.24 | 0.39 | <0.001 |
| Midwest | Unadjusted | 0.27 | 0.22 | 0.33 | <0.001 | 0.30 | 0.24 | 0.38 | <0.001 | 0.28 | 0.19 | 0.41 | <0.001 |
|  | Adjusted | 0.27 | 0.22 | 0.34 | <0.001 | 0.32 | 0.26 | 0.41 | <0.001 | 0.29 | 0.19 | 0.42 | <0.001 |
| South | Unadjusted | 0.29 | 0.26 | 0.33 | <0.001 | 0.31 | 0.26 | 0.36 | <0.001 | 0.35 | 0.26 | 0.47 | <0.001 |
|  | Adjusted | 0.30 | 0.26 | 0.34 | <0.001 | 0.30 | 0.25 | 0.36 | <0.001 | 0.36 | 0.26 | 0.48 | <0.001 |
| West | Unadjusted | 0.19 | 0.16 | 0.22 | <0.001 | 0.20 | 0.17 | 0.25 | <0.001 | 0.21 | 0.15 | 0.30 | <0.001 |
|  | Adjusted | 0.19 | 0.16 | 0.23 | <0.001 | 0.21 | 0.17 | 0.26 | <0.001 | 0.22 | 0.15 | 0.31 | <0.001 |
| New York | Unadjusted | 0.33 | 0.27 | 0.40 | <0.001 | 0.34 | 0.29 | 0.39 | <0.001 | 0.35 | 0.26 | 0.49 | <0.001 |
|  | Adjusted | 0.32 | 0.26 | 0.40 | <0.001 | 0.32 | 0.28 | 0.36 | <0.001 | 0.40 | 0.28 | 0.56 | <0.001 |
|  |  | BP Users versus BP Non-users among Antidiabetic Non-users |  |  |  |  |  |  |  |  |  |  |  |
|  |  | OR | LL | UL | p value | OR | LL | UL | p value | OR | LL | UL | p value |
| All | Unadjusted | 0.24 | 0.23 | 0.26 | <0.001 | 0.24 | 0.22 | 0.26 | <0.001 | 0.24 | 0.20 | 0.29 | <0.001 |
|  | Adjusted | 0.25 | 0.23 | 0.27 | <0.001 | 0.25 | 0.23 | 0.28 | <0.001 | 0.27 | 0.22 | 0.33 | <0.001 |
| Northeast | Unadjusted | 0.24 | 0.22 | 0.28 | <0.001 | 0.26 | 0.22 | 0.29 | <0.001 | 0.25 | 0.19 | 0.34 | <0.001 |
|  | Adjusted | 0.25 | 0.22 | 0.29 | <0.001 | 0.27 | 0.24 | 0.32 | <0.001 | 0.28 | 0.20 | 0.39 | <0.001 |
| Midwest | Unadjusted | 0.27 | 0.22 | 0.32 | <0.001 | 0.22 | 0.17 | 0.30 | <0.001 | 0.26 | 0.16 | 0.42 | <0.001 |
|  | Adjusted | 0.28 | 0.24 | 0.31 | <0.001 | 0.23 | 0.17 | 0.31 | <0.001 | 0.26 | 0.16 | 0.45 | <0.001 |
| South | Unadjusted | 0.24 | 0.21 | 0.27 | <0.001 | 0.25 | 0.20 | 0.30 | <0.001 | 0.29 | 0.20 | 0.43 | <0.001 |
|  | Adjusted | 0.24 | 0.21 | 0.27 | <0.001 | 0.24 | 0.21 | 0.28 | <0.001 | 0.33 | 0.22 | 0.49 | <0.001 |
| West | Unadjusted | 0.23 | 0.20 | 0.27 | <0.001 | 0.18 | 0.14 | 0.24 | <0.001 | 0.13 | 0.07 | 0.23 | <0.001 |
|  | Adjusted | 0.23 | 0.20 | 0.28 | <0.001 | 0.20 | 0.15 | 0.26 | <0.001 | 0.15 | 0.08 | 0.28 | <0.001 |
| New York | Unadjusted | 0.30 | 0.25 | 0.37 | <0.001 | 0.30 | 0.25 | 0.36 | <0.001 | 0.22 | 0.14 | 0.36 | <0.001 |
|  | Adjusted | 0.30 | 0.25 | 0.37 | <0.001 | 0.31 | 0.25 | 0.37 | <0.001 | 0.24 | 0.14 | 0.41 | <0.001 |

LL: lower 95% confidence interval level; OR: odds ratio; UL: upper 95% confidence interval level

**Table S5d:** Antidepressant Use Sensitivity Analysis, Unadjusted/Adjusted Odds Ratio for COVID-19-Related Outcomes, Stratified by Region and New York State

|  |  | Odds of SARS-CoV-2 Test |  |  |  | Odds of COVID-19 Diagnosis |  |  |  | Odds of COVID-19 Hospitalization |  |  |  |
| --- | --- | --- | --- | --- | --- | --- | --- | --- | --- | --- | --- | --- | --- |
|  |  | Antidepressant Users versus Non-users |  |  |  |  |  |  |  |  |  |  |  |
|  |  | OR | LL | UL | p value | OR | LL | UL | p value | OR | LL | UL | p value |
| All | Unadjusted | 1.04 | 1.03 | 1.05 | <0.001 | 0.71 | 0.70 | 0.72 | <0.001 | 0.81 | 0.78 | 0.83 | <0.001 |
|  | Adjusted | 1.00 | 0.99 | 1.01 | 0.61 | 0.65 | 0.64 | 0.66 | <0.001 | 0.75 | 0.73 | 0.78 | <0.001 |
| Northeast | Unadjusted | 1.01 | 0.99 | 1.02 | 0.54 | 0.71 | 0.69 | 0.72 | <0.001 | 0.84 | 0.80 | 0.88 | <0.001 |
|  | Adjusted | 0.97 | 0.95 | 0.99 | 0.001 | 0.65 | 0.63 | 0.66 | <0.001 | 0.77 | 0.73 | 0.82 | <0.001 |
| Midwest | Unadjusted | 1.10 | 1.08 | 1.12 | <0.001 | 0.75 | 0.72 | 0.78 | <0.001 | 0.84 | 0.78 | 0.90 | <0.001 |
|  | Adjusted | 1.05 | 1.03 | 1.07 | <0.001 | 0.69 | 0.66 | 0.71 | <0.001 | 0.78 | 0.73 | 0.84 | <0.001 |
| South | Unadjusted | 1.04 | 1.02 | 1.05 | <0.001 | 0.68 | 0.66 | 0.70 | <0.001 | 0.74 | 0.70 | 0.79 | <0.001 |
|  | Adjusted | 0.99 | 0.98 | 1.01 | 0.49 | 0.64 | 0.62 | 0.66 | <0.001 | 0.72 | 0.68 | 0.77 | <0.001 |
| West | Unadjusted | 1.04 | 1.02 | 1.06 | 0.00 | 0.70 | 0.67 | 0.73 | <0.001 | 0.77 | 0.70 | 0.84 | <0.001 |
|  | Adjusted | 0.99 | 0.97 | 1.02 | 0.46 | 0.64 | 0.61 | 0.66 | <0.001 | 0.70 | 0.64 | 0.77 | <0.001 |
| New York | Unadjusted | 1.00 | 0.97 | 1.03 | 0.86 | 0.77 | 0.74 | 0.80 | <0.001 | 0.83 | 0.76 | 0.91 | <0.001 |
|  | Adjusted | 0.98 | 0.95 | 1.01 | 0.27 | 0.72 | 0.70 | 0.75 | <0.001 | 0.77 | 0.70 | 0.85 | <0.001 |
|  |  | BP Users versus BP Non-users among Antidepressant Users |  |  |  |  |  |  |  |  |  |  |  |
|  |  | OR | LL | UL | p value | OR | LL | UL | p value | OR | LL | UL | p value |
| All | Unadjusted | 0.27 | 0.26 | 0.28 | <0.001 | 0.30 | 0.28 | 0.32 | <0.001 | 0.31 | 0.27 | 0.36 | <0.001 |
|  | Adjusted | 0.27 | 0.25 | 0.28 | <0.001 | 0.30 | 0.28 | 0.32 | <0.001 | 0.33 | 0.28 | 0.38 | <0.001 |
| Northeast | Unadjusted | 0.28 | 0.26 | 0.31 | <0.001 | 0.33 | 0.30 | 0.37 | <0.001 | 0.36 | 0.29 | 0.45 | <0.001 |
|  | Adjusted | 0.28 | 0.25 | 0.30 | <0.001 | 0.32 | 0.29 | 0.36 | <0.001 | 0.37 | 0.29 | 0.47 | <0.001 |
| Midwest | Unadjusted | 0.30 | 0.27 | 0.34 | <0.001 | 0.26 | 0.22 | 0.31 | <0.001 | 0.25 | 0.18 | 0.34 | <0.001 |
|  | Adjusted | 0.30 | 0.26 | 0.34 | <0.001 | 0.27 | 0.22 | 0.33 | <0.001 | 0.26 | 0.18 | 0.36 | <0.001 |
| South | Unadjusted | 0.26 | 0.24 | 0.29 | <0.001 | 0.27 | 0.23 | 0.31 | <0.001 | 0.32 | 0.24 | 0.41 | <0.001 |
|  | Adjusted | 0.26 | 0.24 | 0.28 | <0.001 | 0.27 | 0.23 | 0.32 | <0.001 | 0.32 | 0.24 | 0.43 | <0.001 |
| West | Unadjusted | 0.25 | 0.22 | 0.28 | <0.001 | 0.27 | 0.22 | 0.32 | <0.001 | 0.29 | 0.20 | 0.41 | <0.001 |
|  | Adjusted | 0.24 | 0.21 | 0.27 | <0.001 | 0.29 | 0.28 | 0.30 | <0.001 | 0.33 | 0.23 | 0.48 | <0.001 |
| New York | Unadjusted | 0.30 | 0.26 | 0.34 | <0.001 | 0.33 | 0.28 | 0.38 | <0.001 | 0.24 | 0.16 | 0.36 | <0.001 |
|  | Adjusted | 0.30 | 0.25 | 0.34 | <0.001 | 0.31 | 0.27 | 0.37 | <0.001 | 0.25 | 0.16 | 0.39 | <0.001 |
|  |  | BP Users versus BP Non-users among Antidepressant Non-users |  |  |  |  |  |  |  |  |  |  |  |
|  |  | OR | LL | UL | p value | OR | LL | UL | p value | OR | LL | UL | p value |
| All | Unadjusted | 0.20 | 0.19 | 0.22 | <0.001 | 0.22 | 0.20 | 0.24 | <0.001 | 0.24 | 0.20 | 0.28 | <0.001 |
|  | Adjusted | 0.21 | 0.19 | 0.22 | <0.001 | 0.23 | 0.21 | 0.25 | <0.001 | 0.27 | 0.22 | 0.32 | <0.001 |
| Northeast | Unadjusted | 0.21 | 0.19 | 0.24 | <0.001 | 0.23 | 0.20 | 0.26 | <0.001 | 0.25 | 0.19 | 0.32 | <0.001 |
|  | Adjusted | 0.22 | 0.19 | 0.25 | <0.001 | 0.24 | 0.22 | 0.25 | <0.001 | 0.29 | 0.22 | 0.39 | <0.001 |
| Midwest | Unadjusted | 0.22 | 0.19 | 0.26 | <0.001 | 0.23 | 0.18 | 0.28 | <0.001 | 0.28 | 0.19 | 0.39 | <0.001 |
|  | Adjusted | 0.21 | 0.18 | 0.25 | <0.001 | 0.26 | 0.24 | 0.27 | <0.001 | 0.32 | 0.22 | 0.47 | <0.001 |
| South | Unadjusted | 0.20 | 0.18 | 0.22 | <0.001 | 0.21 | 0.18 | 0.25 | <0.001 | 0.21 | 0.15 | 0.30 | <0.001 |
|  | Adjusted | 0.20 | 0.18 | 0.23 | <0.001 | 0.23 | 0.19 | 0.27 | <0.001 | 0.22 | 0.16 | 0.32 | <0.001 |
| West | Unadjusted | 0.18 | 0.16 | 0.21 | <0.001 | 0.20 | 0.16 | 0.25 | <0.001 | 0.20 | 0.13 | 0.30 | <0.001 |
|  | Adjusted | 0.19 | 0.16 | 0.22 | <0.001 | 0.20 | 0.20 | 0.21 | <0.001 | 0.22 | 0.14 | 0.35 | <0.001 |
| New York | Unadjusted | 0.26 | 0.22 | 0.32 | <0.001 | 0.27 | 0.23 | 0.32 | <0.001 | 0.29 | 0.19 | 0.43 | <0.001 |
|  | Adjusted | 0.26 | 0.23 | 0.30 | <0.001 | 0.26 | 0.22 | 0.32 | <0.001 | 0.35 | 0.22 | 0.54 | <0.001 |

LL: lower 95% confidence interval level; OR: odds ratio; UL: upper 95% confidence interval level

**Table S6a:** “*Bone-Rx*” Cohort (All Regions), Patient Characteristics Pre/Post Match

|  | "Bone-Rx" Cohort / All Observations Unmatched |  |  |  |  |  |  | "Bone-Rx" Cohort / All Observations Matched |  |  |  |  |  |  |
| --- | --- | --- | --- | --- | --- | --- | --- | --- | --- | --- | --- | --- | --- | --- |
|  | All |  | BP Non-user |  | BP User |  | p-value | All |  | BP Non-user |  | BP User |  | p-value |
|  | N | % | N | % | N | % |  | N | % | N | % | N | % |  |
| All Patients | 502,895 | 100.0% | 50,844 | 10.1% | 452,051 | 89.9% |  | 100,996 | 100.0% | 50,498 | 50.0% | 50,498 | 50.0% |  |
| Age |  |  |  |  |  |  |  |  |  |  |  |  |  |  |
| ≤20 | 1,164 | 0.2% | 36 | 0.1% | 1,128 | 0.2% | <0.001 | 67 | 0.1% | 36 | 0.1% | 31 | 0.1% | 0.97 |
| 21-40 | 3,501 | 0.7% | 410 | 0.8% | 3,091 | 0.7% |  | 790 | 0.8% | 403 | 0.8% | 387 | 0.8% |  |
| 41-50 | 9,631 | 1.9% | 1,080 | 2.1% | 8,551 | 1.9% |  | 2,107 | 2.1% | 1,069 | 2.1% | 1,038 | 2.1% |  |
| 51-60 | 72,139 | 14.3% | 6,418 | 12.6% | 65,721 | 14.5% |  | 12,777 | 12.7% | 6,395 | 12.7% | 6,382 | 12.6% |  |
| 61-70 | 171,687 | 34.1% | 14,809 | 29.1% | 156,878 | 34.7% |  | 29,509 | 29.2% | 14,751 | 29.2% | 14,758 | 29.2% |  |
| 71-80 | 157,877 | 31.4% | 16,152 | 31.8% | 141,725 | 31.4% |  | 32,129 | 31.8% | 16,055 | 31.8% | 16,074 | 31.8% |  |
| ≥81 | 86,896 | 17.3% | 11,939 | 23.5% | 74,957 | 16.6% |  | 23,617 | 23.4% | 11,789 | 23.3% | 11,828 | 23.4% |  |
| Gender |  |  |  |  |  |  |  |  |  |  |  |  |  |  |
| Female | 451,790 | 89.8% | 44,354 | 87.2% | 407,436 | 90.1% | <0.001 | 88,552 | 87.7% | 44,235 | 87.6% | 44,317 | 87.8% | 0.43 |
| Male | 51,105 | 10.2% | 6,490 | 12.8% | 44,615 | 9.9% |  | 12,444 | 12.3% | 6,263 | 12.4% | 6,181 | 12.2% |  |
| Region |  |  |  |  |  |  |  |  |  |  |  |  |  |  |
| Midwest | 85,391 | 17.0% | 9,424 | 18.5% | 75,967 | 16.8% | <0.001 | 18,720 | 18.5% | 9,360 | 18.5% | 9,360 | 18.5% | 1.00 |
| Northeast | 135,867 | 27.0% | 16,139 | 31.7% | 119,728 | 26.5% |  | 31,986 | 31.7% | 15,993 | 31.7% | 15,993 | 31.7% |  |
| South | 178,118 | 35.4% | 17,232 | 33.9% | 160,886 | 35.6% |  | 34,280 | 33.9% | 17,140 | 33.9% | 17,140 | 33.9% |  |
| West | 103,519 | 20.6% | 8,049 | 15.8% | 95,470 | 21.1% |  | 16,010 | 15.9% | 8,005 | 15.9% | 8,005 | 15.9% |  |
| Insurance |  |  |  |  |  |  |  |  |  |  |  |  |  |  |
| Commercial | 164,150 | 32.6% | 17,092 | 33.6% | 147,058 | 32.5% | <0.001 | 33,977 | 33.6% | 16,963 | 33.6% | 17,014 | 33.7% | 0.91 |
| Dual | 33,969 | 6.8% | 2,562 | 5.0% | 31,407 | 6.9% |  | 5,056 | 5.0% | 2,547 | 5.0% | 2,509 | 5.0% |  |
| Medicaid | 84,514 | 16.8% | 7,034 | 13.8% | 77,480 | 17.1% |  | 13,925 | 13.8% | 6,986 | 13.8% | 6,939 | 13.7% |  |
| Medicare | 220,262 | 43.8% | 24,156 | 47.5% | 196,106 | 43.4% |  | 48,038 | 47.6% | 24,002 | 47.5% | 24,036 | 47.6% |  |
| PCP Visit 2019 |  |  |  |  |  |  |  |  |  |  |  |  |  |  |
| No | 181,996 | 36.2% | 18,130 | 35.7% | 163,866 | 36.2% | 0.009 | 35,943 | 35.6% | 17,979 | 35.6% | 17,964 | 35.6% | 0.92 |
| Yes | 320,899 | 63.8% | 32,714 | 64.3% | 288,185 | 63.8% |  | 65,053 | 64.4% | 32,519 | 64.4% | 32,534 | 64.4% |  |
| Continuous Outcomes |  |  |  |  |  |  |  |  |  |  |  |  |  |  |
|  | mean | SD | mean | SD | mean | SD | p-value | mean | SD | mean | SD | mean | SD | p-value |
| CCI | 1.05 | 1.91 | 1.99 | 2.71 | 0.95 | 1.76 | <0.001 | 1.93 | 2.59 | 1.93 | 2.60 | 1.92 | 2.59 | 0.76 |

BP: bisphosphonate; CCI: Charlson Comorbidity Index; PCP: primary care physician; SD: standard deviation

**Table S6b:**
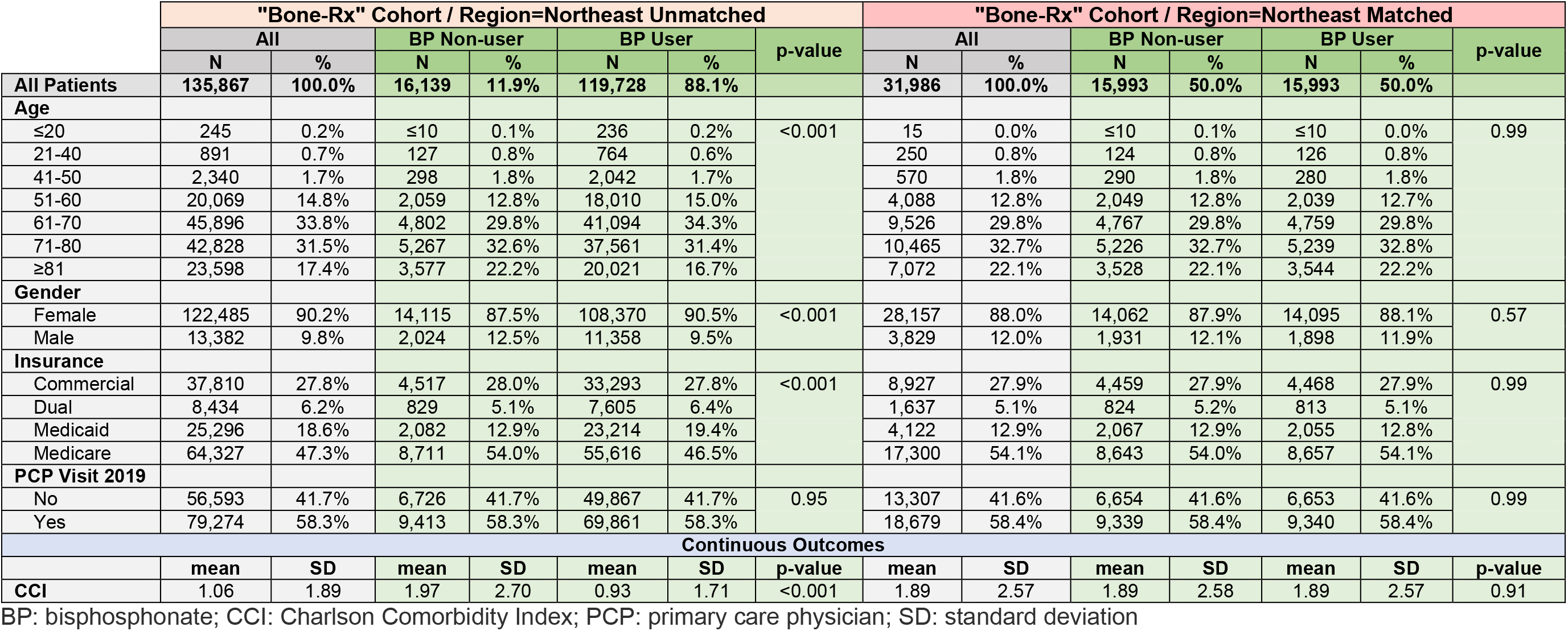
“*Bone-Rx*” Cohort (Region=Northeast), Patient Characteristics Pre/Post Match

|  | "Bone-Rx" Cohort / Region=Northeast Unmatched |  |  |  |  |  |  | "Bone-Rx" Cohort / Region=Northeast Matched |  |  |  |  |  |  |
| --- | --- | --- | --- | --- | --- | --- | --- | --- | --- | --- | --- | --- | --- | --- |
|  | All |  | BP Non-user |  | BP User |  | p-value | All |  | BP Non-user |  | BP User |  | p-value |
|  | N | % | N | % | N | % |  | N | % | N | % | N | % |  |
| All Patients | 135,867 | 100.0% | 16,139 | 11.9% | 119,728 | 88.1% |  | 31,986 | 100.0% | 15,993 | 50.0% | 15,993 | 50.0% |  |
| Age |  |  |  |  |  |  |  |  |  |  |  |  |  |  |
| ≤20 | 245 | 0.2% | ≤10 | 0.1% | 236 | 0.2% | <0.001 | 15 | 0.0% | ≤10 | 0.1% | ≤10 | 0.0% | 0.99 |
| 21-40 | 891 | 0.7% | 127 | 0.8% | 764 | 0.6% |  | 250 | 0.8% | 124 | 0.8% | 126 | 0.8% |  |
| 41-50 | 2,340 | 1.7% | 298 | 1.8% | 2,042 | 1.7% |  | 570 | 1.8% | 290 | 1.8% | 280 | 1.8% |  |
| 51-60 | 20,069 | 14.8% | 2,059 | 12.8% | 18,010 | 15.0% |  | 4,088 | 12.8% | 2,049 | 12.8% | 2,039 | 12.7% |  |
| 61-70 | 45,896 | 33.8% | 4,802 | 29.8% | 41,094 | 34.3% |  | 9,526 | 29.8% | 4,767 | 29.8% | 4,759 | 29.8% |  |
| 71-80 | 42,828 | 31.5% | 5,267 | 32.6% | 37,561 | 31.4% |  | 10,465 | 32.7% | 5,226 | 32.7% | 5,239 | 32.8% |  |
| ≥81 | 23,598 | 17.4% | 3,577 | 22.2% | 20,021 | 16.7% |  | 7,072 | 22.1% | 3,528 | 22.1% | 3,544 | 22.2% |  |
| Gender |  |  |  |  |  |  |  |  |  |  |  |  |  |  |
| Female | 122,485 | 90.2% | 14,115 | 87.5% | 108,370 | 90.5% | <0.001 | 28,157 | 88.0% | 14,062 | 87.9% | 14,095 | 88.1% | 0.57 |
| Male | 13,382 | 9.8% | 2,024 | 12.5% | 11,358 | 9.5% |  | 3,829 | 12.0% | 1,931 | 12.1% | 1,898 | 11.9% |  |
| Insurance |  |  |  |  |  |  |  |  |  |  |  |  |  |  |
| Commercial | 37,810 | 27.8% | 4,517 | 28.0% | 33,293 | 27.8% | <0.001 | 8,927 | 27.9% | 4,459 | 27.9% | 4,468 | 27.9% | 0.99 |
| Dual | 8,434 | 6.2% | 829 | 5.1% | 7,605 | 6.4% |  | 1,637 | 5.1% | 824 | 5.2% | 813 | 5.1% |  |
| Medicaid | 25,296 | 18.6% | 2,082 | 12.9% | 23,214 | 19.4% |  | 4,122 | 12.9% | 2,067 | 12.9% | 2,055 | 12.8% |  |
| Medicare | 64,327 | 47.3% | 8,711 | 54.0% | 55,616 | 46.5% |  | 17,300 | 54.1% | 8,643 | 54.0% | 8,657 | 54.1% |  |
| PCP Visit 2019 |  |  |  |  |  |  |  |  |  |  |  |  |  |  |
| No | 56,593 | 41.7% | 6,726 | 41.7% | 49,867 | 41.7% | 0.95 | 13,307 | 41.6% | 6,654 | 41.6% | 6,653 | 41.6% | 0.99 |
| Yes | 79,274 | 58.3% | 9,413 | 58.3% | 69,861 | 58.3% |  | 18,679 | 58.4% | 9,339 | 58.4% | 9,340 | 58.4% |  |
| Continuous Outcomes |  |  |  |  |  |  |  |  |  |  |  |  |  |  |
|  | mean | SD | mean | SD | mean | SD | p-value | mean | SD | mean | SD | mean | SD | p-value |
| CCI | 1.06 | 1.89 | 1.97 | 2.70 | 0.93 | 1.71 | <0.001 | 1.89 | 2.57 | 1.89 | 2.58 | 1.89 | 2.57 | 0.91 |

BP: bisphosphonate; CCI: Charlson Comorbidity Index; PCP: primary care physician; SD: standard deviation

**Table S6c:**
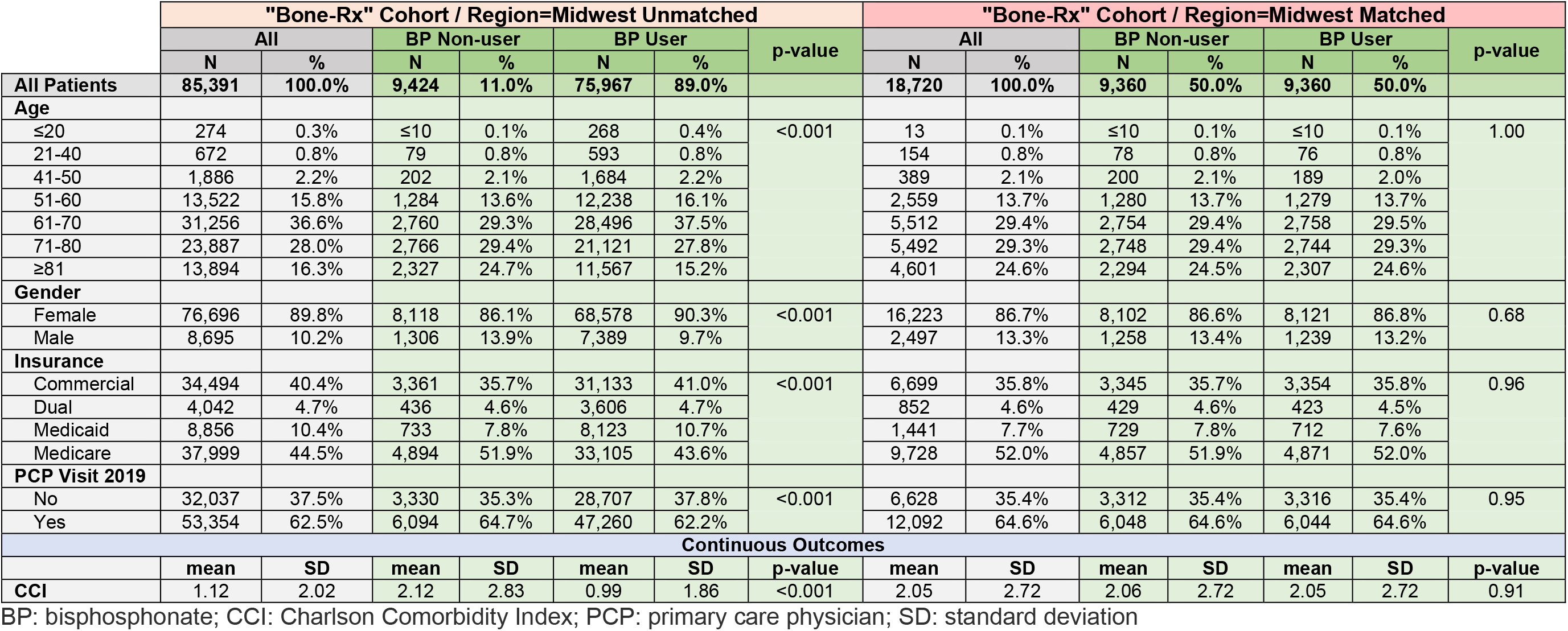
“*Bone-Rx*” Cohort (Region=Midwest), Patient Characteristics Pre/Post Match

**Table S6d:** “*Bone-Rx*” Cohort (Region=South), Patient Characteristics Pre/Post Match

|  | "Bone-Rx" Cohort / Region=South Unmatched |  |  |  |  |  |  | "Bone-Rx" Cohort / Region=South Matched |  |  |  |  |  |  |
| --- | --- | --- | --- | --- | --- | --- | --- | --- | --- | --- | --- | --- | --- | --- |
|  | All |  | BP Non-user |  | BP User |  | p-value | All |  | BP Non-user |  | BP User |  | p-value |
|  | N | % | N | % | N | % |  | N | % | N | % | N | % |  |
| All Patients | 178,118 | 100.0% | 17,232 | 9.7% | 160,886 | 90.3% |  | 34,280 | 100.0% | 17,140 | 50.0% | 17,140 | 50.0% |  |
| Age |  |  |  |  |  |  |  |  |  |  |  |  |  |  |
| ≤20 | 490 | 0.3% | 16 | 0.1% | 474 | 0.3% | <0.001 | 31 | 0.1% | 16 | 0.1% | 15 | 0.1% | 1.00 |
| 21-40 | 1,313 | 0.7% | 136 | 0.8% | 1,177 | 0.7% |  | 262 | 0.8% | 134 | 0.8% | 128 | 0.7% |  |
| 41-50 | 3,866 | 2.2% | 445 | 2.6% | 3,421 | 2.1% |  | 884 | 2.6% | 444 | 2.6% | 440 | 2.6% |  |
| 51-60 | 27,389 | 15.4% | 2,296 | 13.3% | 25,093 | 15.6% |  | 4,574 | 13.3% | 2,290 | 13.4% | 2,284 | 13.3% |  |
| 61-70 | 61,038 | 34.3% | 5,142 | 29.8% | 55,896 | 34.7% |  | 10,271 | 30.0% | 5,129 | 29.9% | 5,142 | 30.0% |  |
| 71-80 | 56,126 | 31.5% | 5,521 | 32.0% | 50,605 | 31.5% |  | 10,990 | 32.1% | 5,493 | 32.0% | 5,497 | 32.1% |  |
| ≥81 | 27,896 | 15.7% | 3,676 | 21.3% | 24,220 | 15.1% |  | 7,268 | 21.2% | 3,634 | 21.2% | 3,634 | 21.2% |  |
| Gender |  |  |  |  |  |  |  |  |  |  |  |  |  |  |
| Female | 160,994 | 90.4% | 15,179 | 88.1% | 145,815 | 90.6% | <0.001 | 30,322 | 88.5% | 15,149 | 88.4% | 15,173 | 88.5% | 0.69 |
| Male | 17,124 | 9.6% | 2,053 | 11.9% | 15,071 | 9.4% |  | 3,958 | 11.5% | 1,991 | 11.6% | 1,967 | 11.5% |  |
| Insurance |  |  |  |  |  |  |  |  |  |  |  |  |  |  |
| Commercial | 66,332 | 37.2% | 7,042 | 40.9% | 59,290 | 36.9% | <0.001 | 14,052 | 41.0% | 7,007 | 40.9% | 7,045 | 41.1% | 0.95 |
| Dual | 14,829 | 8.3% | 769 | 4.5% | 14,060 | 8.7% |  | 1,523 | 4.4% | 769 | 4.5% | 754 | 4.4% |  |
| Medicaid | 23,492 | 13.2% | 1,843 | 10.7% | 21,649 | 13.5% |  | 3,639 | 10.6% | 1,829 | 10.7% | 1,810 | 10.6% |  |
| Medicare | 73,465 | 41.2% | 7,578 | 44.0% | 65,887 | 41.0% |  | 15,066 | 43.9% | 7,535 | 44.0% | 7,531 | 43.9% |  |
| PCP Visit 2019 |  |  |  |  |  |  |  |  |  |  |  |  |  |  |
| No | 60,253 | 33.8% | 5,785 | 33.6% | 54,468 | 33.9% | 0.454 | 11,462 | 33.4% | 5,736 | 33.5% | 5,726 | 33.4% | 0.91 |
| Yes | 117,865 | 66.2% | 11,447 | 66.4% | 106,418 | 66.1% |  | 22,818 | 66.6% | 11,404 | 66.5% | 11,414 | 66.6% |  |
| Continuous Outcomes |  |  |  |  |  |  |  |  |  |  |  |  |  |  |
|  | mean | SD | mean | SD | mean | SD | p-value | mean | SD | mean | SD | mean | SD | p-value |
| CCI | 0.95 | 1.84 | 1.86 | 2.65 | 0.86 | 1.70 | <0.001 | 1.80 | 2.54 | 1.80 | 2.54 | 1.79 | 2.53 | 0.78 |

BP: bisphosphonate; CCI: Charlson Comorbidity Index; PCP: primary care physician; SD: standard deviation

**Table S6e:** “*Bone-Rx*” Cohort (Region=West), Patient Characteristics Pre/Post Match

|  | "Bone-Rx" Cohort / Region=West Unmatched |  |  |  |  |  |  | "Bone-Rx" Cohort / Region=West Matched |  |  |  |  |  |  |
| --- | --- | --- | --- | --- | --- | --- | --- | --- | --- | --- | --- | --- | --- | --- |
|  | All |  | BP Non-user |  | BP User |  | p-value | All |  | BP Non-user |  | BP User |  | p-value |
|  | N | % | N | % | N | % |  | N | % | N | % | N | % |  |
| All Patients | 103,519 | 100.0% | 8,049 | 7.8% | 95,470 | 92.2% |  | 16,010 | 100.0% | 8,005 | 50.0% | 8,005 | 50.0% |  |
| Age |  |  |  |  |  |  |  |  |  |  |  |  |  |  |
| ≤20 | 155 | 0.1% | ≤10 | 0.1% | 150 | 0.2% | <0.001 | ≤10 | 0.0% | ≤10 | 0.1% | ≤10 | 0.0% | 0.96 |
| 21-40 | 625 | 0.6% | 68 | 0.8% | 557 | 0.6% |  | 124 | 0.8% | 67 | 0.8% | 57 | 0.7% |  |
| 41-50 | 1,539 | 1.5% | 135 | 1.7% | 1,404 | 1.5% |  | 264 | 1.6% | 135 | 1.7% | 129 | 1.6% |  |
| 51-60 | 11,159 | 10.8% | 779 | 9.7% | 10,380 | 10.9% |  | 1,556 | 9.7% | 776 | 9.7% | 780 | 9.7% |  |
| 61-70 | 33,497 | 32.4% | 2,105 | 26.2% | 31,392 | 32.9% |  | 4,200 | 26.2% | 2,101 | 26.2% | 2,099 | 26.2% |  |
| 71-80 | 35,036 | 33.8% | 2,598 | 32.3% | 32,438 | 34.0% |  | 5,182 | 32.4% | 2,588 | 32.3% | 2,594 | 32.4% |  |
| ≥81 | 21,508 | 20.8% | 2,359 | 29.3% | 19,149 | 20.1% |  | 4,676 | 29.2% | 2,333 | 29.1% | 2,343 | 29.3% |  |
| Gender |  |  |  |  |  |  |  |  |  |  |  |  |  |  |
| Female | 91,615 | 88.5% | 6,942 | 86.2% | 84,673 | 88.7% | <0.001 | 13,850 | 86.5% | 6,922 | 86.5% | 6,928 | 86.5% | 0.89 |
| Male | 11,904 | 11.5% | 1,107 | 13.8% | 10,797 | 11.3% |  | 2,160 | 13.5% | 1,083 | 13.5% | 1,077 | 13.5% |  |
| Insurance |  |  |  |  |  |  |  |  |  |  |  |  |  |  |
| Commercial | 25,514 | 24.6% | 2,172 | 27.0% | 23,342 | 24.4% | <0.001 | 4,299 | 26.9% | 2,152 | 26.9% | 2,147 | 26.8% | 1.00 |
| Dual | 6,664 | 6.4% | 528 | 6.6% | 6,136 | 6.4% |  | 1,044 | 6.5% | 525 | 6.6% | 519 | 6.5% |  |
| Medicaid | 26,870 | 26.0% | 2,376 | 29.5% | 24,494 | 25.7% |  | 4,723 | 29.5% | 2,361 | 29.5% | 2,362 | 29.5% |  |
| Medicare | 44,471 | 43.0% | 2,973 | 36.9% | 41,498 | 43.5% |  | 5,944 | 37.1% | 2,967 | 37.1% | 2,977 | 37.2% |  |
| PCP Visit 2019 |  |  |  |  |  |  |  |  |  |  |  |  |  |  |
| No | 33,113 | 32.0% | 2,289 | 28.4% | 30,824 | 32.3% | <0.001 | 4,546 | 28.4% | 2,277 | 28.4% | 2,269 | 28.3% | 0.89 |
| Yes | 70,406 | 68.0% | 5,760 | 71.6% | 64,646 | 67.7% |  | 11,464 | 71.6% | 5,728 | 71.6% | 5,736 | 71.7% |  |
| Continuous Outcomes |  |  |  |  |  |  |  |  |  |  |  |  |  |  |
|  | mean | SD | mean | SD | mean | SD | p-value | mean | SD | mean | SD | mean | SD | p-value |
| CCI | 1.17 | 1.94 | 2.17 | 2.67 | 1.08 | 1.84 | <0.001 | 2.12 | 2.59 | 2.12 | 2.59 | 2.12 | 2.59 | 0.93 |

BP: bisphosphonate; CCI: Charlson Comorbidity Index; PCP: primary care physician; SD: standard deviation

**Table S6f:** “*Bone-Rx*” Cohort (Region=New York State), Patient Characteristics Pre/Post Match

|  | "Bone-Rx" Cohort / Region=New York State Unmatched |  |  |  |  |  |  | "Bone-Rx" Cohort / Region=New York State Matched |  |  |  |  |  |  |
| --- | --- | --- | --- | --- | --- | --- | --- | --- | --- | --- | --- | --- | --- | --- |
|  | All |  | BP Non-user |  | BP User |  | p-value | All |  | BP Non-user |  | BP User |  | p-value |
|  | N | % | N | % | N | % |  | N | % | N | % | N | % |  |
| All Patients | 57,397 | 100.0% | 7,362 | 12.8% | 50,035 | 87.2% |  | 14,508 | 100.0% | 7,254 | 50.0% | 7,254 | 50.0% |  |
| Age |  |  |  |  |  |  |  |  |  |  |  |  |  |  |
| ≤20 | 56 | 0.1% | ≤10 | 0.1% | 50 | 0.1% | <0.001 | 11 | 0.1% | ≤10 | 0.1% | ≤10 | 0.1% | 0.96 |
| 21-40 | 272 | 0.5% | 44 | 0.6% | 228 | 0.5% |  | 76 | 0.5% | 42 | 0.6% | 34 | 0.5% |  |
| 41-50 | 775 | 1.4% | 120 | 1.6% | 655 | 1.3% |  | 207 | 1.4% | 107 | 1.5% | 100 | 1.4% |  |
| 51-60 | 7,249 | 12.6% | 885 | 12.0% | 6,364 | 12.7% |  | 1,744 | 12.0% | 871 | 12.0% | 873 | 12.0% |  |
| 61-70 | 18,433 | 32.1% | 2,297 | 31.2% | 16,136 | 32.2% |  | 4,540 | 31.3% | 2,264 | 31.2% | 2,276 | 31.4% |  |
| 71-80 | 19,944 | 34.7% | 2,482 | 33.7% | 17,462 | 34.9% |  | 4,934 | 34.0% | 2,455 | 33.8% | 2,479 | 34.2% |  |
| ≥81 | 10,668 | 18.6% | 1,528 | 20.8% | 9,140 | 18.3% |  | 2,996 | 20.7% | 1,509 | 20.8% | 1,487 | 20.5% |  |
| Gender |  |  |  |  |  |  |  |  |  |  |  |  |  |  |
| Female | 52,047 | 90.7% | 6,589 | 89.5% | 45,458 | 90.9% | <.001 | 13,106 | 90.3% | 6,526 | 90.0% | 6,580 | 90.7% | 0.13 |
| Male | 5,350 | 9.3% | 773 | 10.5% | 4,577 | 9.1% |  | 1,402 | 9.7% | 728 | 10.0% | 674 | 9.3% |  |
| Insurance |  |  |  |  |  |  |  |  |  |  |  |  |  |  |
| Commercial | 12,309 | 21.4% | 1,894 | 25.7% | 10,415 | 20.8% | <0.001 | 3,706 | 25.5% | 1,850 | 25.5% | 1,856 | 25.6% | 1.00 |
| Dual | 1,750 | 3.0% | 154 | 2.1% | 1,596 | 3.2% |  | 307 | 2.1% | 153 | 2.1% | 154 | 2.1% |  |
| Medicaid | 10,191 | 17.8% | 1,016 | 13.8% | 9,175 | 18.3% |  | 1,968 | 13.6% | 987 | 13.6% | 981 | 13.5% |  |
| Medicare | 33,147 | 57.8% | 4,298 | 58.4% | 28,849 | 57.7% |  | 8,527 | 58.8% | 4,264 | 58.8% | 4,263 | 58.8% |  |
| PCP Visit 2019 |  |  |  |  |  |  |  |  |  |  |  |  |  |  |
| No | 21,462 | 37.4% | 2,789 | 37.9% | 18,673 | 37.3% | 0.35 | 5,468 | 37.7% | 2,744 | 37.8% | 2,724 | 37.6% | 0.73 |
| Yes | 35,935 | 62.6% | 4,573 | 62.1% | 31,362 | 62.7% |  | 9,040 | 62.3% | 4,510 | 62.2% | 4,530 | 62.4% |  |
| Continuous Outcomes |  |  |  |  |  |  |  |  |  |  |  |  |  |  |
|  | mean | SD | mean | SD | mean | SD | p-value | mean | SD | mean | SD | mean | SD | p-value |
| CCI | 1.06 | 1.84 | 1.81 | 2.56 | 0.95 | 1.68 | <0.001 | 1.69 | 2.35 | 1.69 | 2.36 | 1.69 | 2.35 | 0.98 |

BP: bisphosphonate; CCI: Charlson Comorbidity Index; PCP: primary care physician; SD: standard deviation

**Table S7:** “*Osteo-Dx-Rx*” Cohort, Patient Characteristics Pre/Post Match

|  | "Osteo-Dx-Rx" Cohort / All Observations Unmatched |  |  |  |  |  |  | "Osteo-Dx-Rx" Cohort / All Observations Matched |  |  |  |  |  |  |
| --- | --- | --- | --- | --- | --- | --- | --- | --- | --- | --- | --- | --- | --- | --- |
|  | All |  | BP Non-user |  | BP User |  | p-value | All |  | BP Non-user |  | BP User |  | p-value |
|  | N | % | N | % | N | % |  | N | % | N | % | N | % |  |
| All Patients | 60,043 | 100.0% | 8,392 | 14.0% | 51,651 | 86.0% |  | 15,898 | 100.0% | 7,949 | 50.0% | 7,949 | 50.0% |  |
| Age |  |  |  |  |  |  |  |  |  |  |  |  |  |  |
| 51-60 | 6,443 | 10.7% | 753 | 9.0% | 5,690 | 11.0% | <0.001 | 1,430 | 9.0% | 723 | 9.1% | 707 | 8.9% | 0.95 |
| 61-70 | 20,187 | 33.6% | 2,492 | 29.7% | 17,695 | 34.3% |  | 4,821 | 30.3% | 2,397 | 30.2% | 2,424 | 30.5% |  |
| 71-80 | 21,545 | 35.9% | 2,964 | 35.3% | 18,581 | 36.0% |  | 5,677 | 35.7% | 2,841 | 35.7% | 2,836 | 35.7% |  |
| ≥81 | 11,868 | 19.8% | 2,183 | 26.0% | 9,685 | 18.8% |  | 3,970 | 25.0% | 1,988 | 25.0% | 1,982 | 24.9% |  |
| State |  |  |  |  |  |  |  |  |  |  |  |  |  |  |
| CA | 24,489 | 40.8% | 2,558 | 30.5% | 21,931 | 42.5% | <0.001 | 4,886 | 30.7% | 2,443 | 30.7% | 2,443 | 30.7% | 1.00 |
| FL | 11,904 | 19.8% | 1,767 | 21.1% | 10,137 | 19.6% |  | 3,256 | 20.5% | 1,628 | 20.5% | 1,628 | 20.5% |  |
| IL | 4,447 | 7.4% | 678 | 8.1% | 3,769 | 7.3% |  | 1,168 | 7.3% | 584 | 7.3% | 584 | 7.3% |  |
| NY | 19,203 | 32.0% | 3,389 | 40.4% | 15,814 | 30.6% |  | 6,588 | 41.4% | 3,294 | 41.4% | 3,294 | 41.4% |  |
| Insurance |  |  |  |  |  |  |  |  |  |  |  |  |  |  |
| Commercial | 12,990 | 21.6% | 2,048 | 24.4% | 10,942 | 21.2% | <0.001 | 3,736 | 23.5% | 1,868 | 23.5% | 1,868 | 23.5% | 1.00 |
| Dual | 3,652 | 6.1% | 313 | 3.7% | 3,339 | 6.5% |  | 554 | 3.5% | 277 | 3.5% | 277 | 3.5% |  |
| Medicaid | 13,698 | 22.8% | 1,785 | 21.3% | 11,913 | 23.1% |  | 3,392 | 21.3% | 1,696 | 21.3% | 1,696 | 21.3% |  |
| Medicare | 29,703 | 49.5% | 4,246 | 50.6% | 25,457 | 49.3% |  | 8,216 | 51.7% | 4,108 | 51.7% | 4,108 | 51.7% |  |
| PCP Visit 2019 |  |  |  |  |  |  |  |  |  |  |  |  |  |  |
| No | 14,089 | 23.5% | 2,427 | 28.9% | 11,662 | 22.6% | <0.001 | 4,487 | 28.2% | 2,243 | 28.2% | 2,244 | 28.2% | 0.99 |
| Yes | 45,954 | 76.5% | 5,965 | 71.1% | 39,989 | 77.4% |  | 11,411 | 71.8% | 5,706 | 71.8% | 5,705 | 71.8% |  |
| Cancer Dx |  |  |  |  |  |  |  |  |  |  |  |  |  |  |
| No | 52,301 | 87.1% | 6,765 | 80.6% | 45,536 | 88.2% | <0.001 | 13,116 | 82.5% | 6,548 | 82.4% | 6,568 | 82.6% | 0.68 |
| Yes | 7,742 | 12.9% | 1,627 | 19.4% | 6,115 | 11.8% |  | 2,782 | 17.5% | 1,401 | 17.6% | 1,381 | 17.4% |  |
| COPD Dx |  |  |  |  |  |  |  |  |  |  |  |  |  |  |
| No | 53,446 | 89.0% | 7,035 | 83.8% | 46,411 | 89.9% | <0.001 | 13,705 | 86.2% | 6,834 | 86.0% | 6,871 | 86.4% | 0.39 |
| Yes | 6,597 | 11.0% | 1,357 | 16.2% | 5,240 | 10.1% |  | 2,193 | 13.8% | 1,115 | 14.0% | 1,078 | 13.6% |  |
| Heart Failure Dx |  |  |  |  |  |  |  |  |  |  |  |  |  |  |
| No | 56,005 | 93.3% | 7,492 | 89.3% | 48,513 | 93.9% | <0.001 | 14,475 | 91.0% | 7,218 | 90.8% | 7,257 | 91.3% | 0.28 |
| Yes | 4,038 | 6.7% | 900 | 10.7% | 3,138 | 6.1% |  | 1,423 | 9.0% | 731 | 9.2% | 692 | 8.7% |  |
| Hypertension Dx |  |  |  |  |  |  |  |  |  |  |  |  |  |  |
| No | 24,966 | 41.6% | 3,281 | 39.1% | 21,685 | 42.0% | <0.001 | 6,268 | 39.4% | 3,137 | 39.5% | 3,131 | 39.4% | 0.92 |
| Yes | 35,077 | 58.4% | 5,111 | 60.9% | 29,966 | 58.0% |  | 9,630 | 60.6% | 4,812 | 60.5% | 4,818 | 60.6% |  |
| Dyslipidemia Dx |  |  |  |  |  |  |  |  |  |  |  |  |  |  |
| No | 24,095 | 40.1% | 3,295 | 39.3% | 20,800 | 40.3% | 0.08 | 6,187 | 38.9% | 3,101 | 39.0% | 3,086 | 38.8% | 0.81 |
| Yes | 35,948 | 59.9% | 5,097 | 60.7% | 30,851 | 59.7% |  | 9,711 | 61.1% | 4,848 | 61.0% | 4,863 | 61.2% |  |
| Obesity Dx |  |  |  |  |  |  |  |  |  |  |  |  |  |  |
| No | 53,453 | 89.0% | 7,583 | 90.4% | 45,870 | 88.8% | <0.001 | 14,468 | 91.0% | 7,217 | 90.8% | 7,251 | 91.2% | 0.35 |
| Yes | 6,590 | 11.0% | 809 | 9.6% | 5,781 | 11.2% |  | 1,430 | 9.0% | 732 | 9.2% | 698 | 8.8% |  |
| Type 2 Diabetes Dx |  |  |  |  |  |  |  |  |  |  |  |  |  |  |
| No | 44,565 | 74.2% | 6,132 | 73.1% | 38,433 | 74.4% | 0.009 | 11,759 | 74.0% | 5,859 | 73.7% | 5,900 | 74.2% | 0.46 |
| Yes | 15,478 | 25.8% | 2,260 | 26.9% | 13,218 | 25.6% |  | 4,139 | 26.0% | 2,090 | 26.3% | 2,049 | 25.8% |  |
| Depression Dx |  |  |  |  |  |  |  |  |  |  |  |  |  |  |
| No | 51,609 | 86.0% | 7,114 | 84.8% | 44,495 | 86.1% | 0.001 | 13,697 | 86.2% | 6,844 | 86.1% | 6,853 | 86.2% | 0.84 |
| Yes | 8,434 | 14.0% | 1,278 | 15.2% | 7,156 | 13.9% |  | 2,201 | 13.8% | 1,105 | 13.9% | 1,096 | 13.8% |  |

BP: bisphosphonate; CCI: Charlson Comorbidity Index; CA: California; Dx: diagnosis; FL: Florida; IL: Illinois; NY: New York; PCP: primary care physician

**Table S8a:**
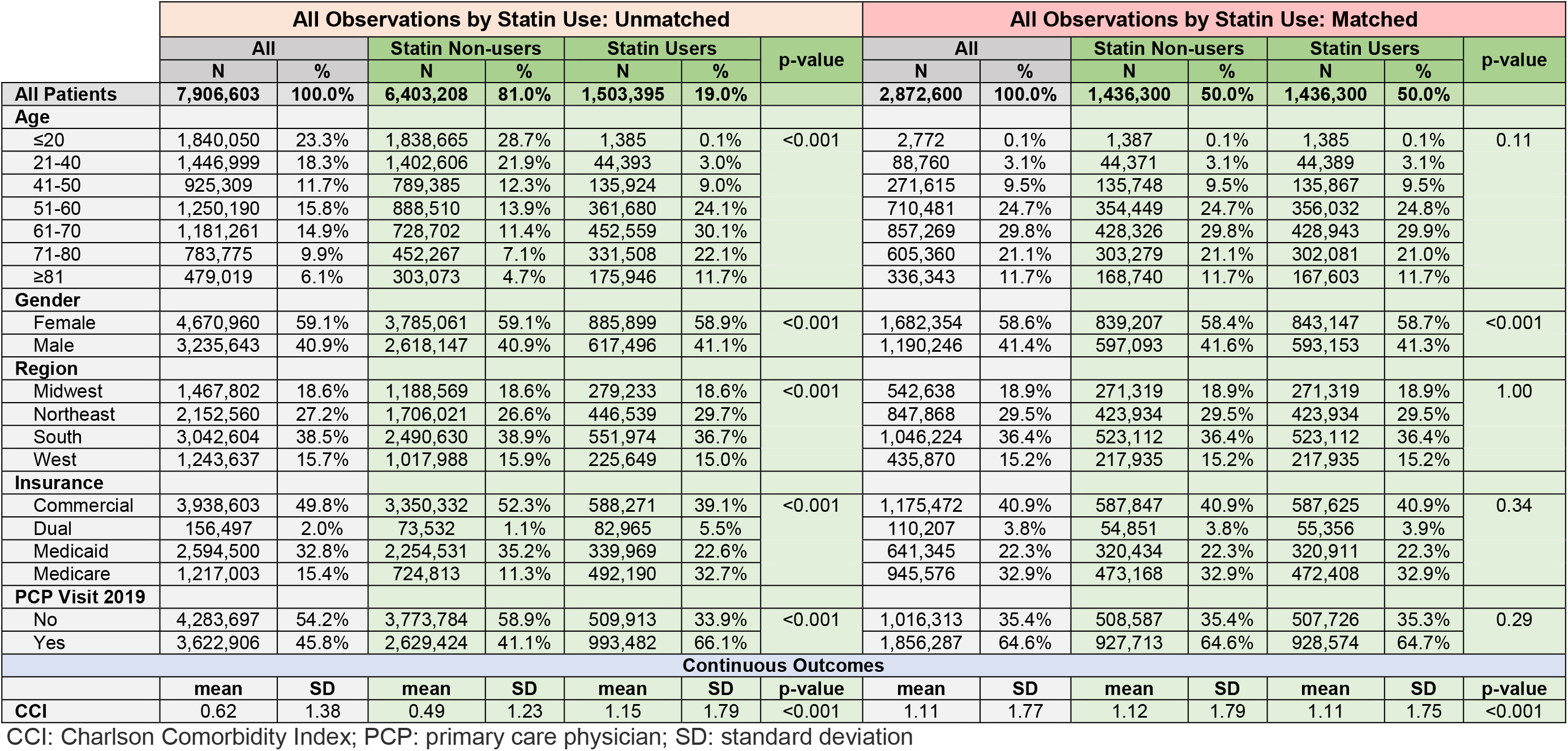
Statin Cohort (All Regions), Patient Characteristics Pre/Post Match

**Table S8b:**
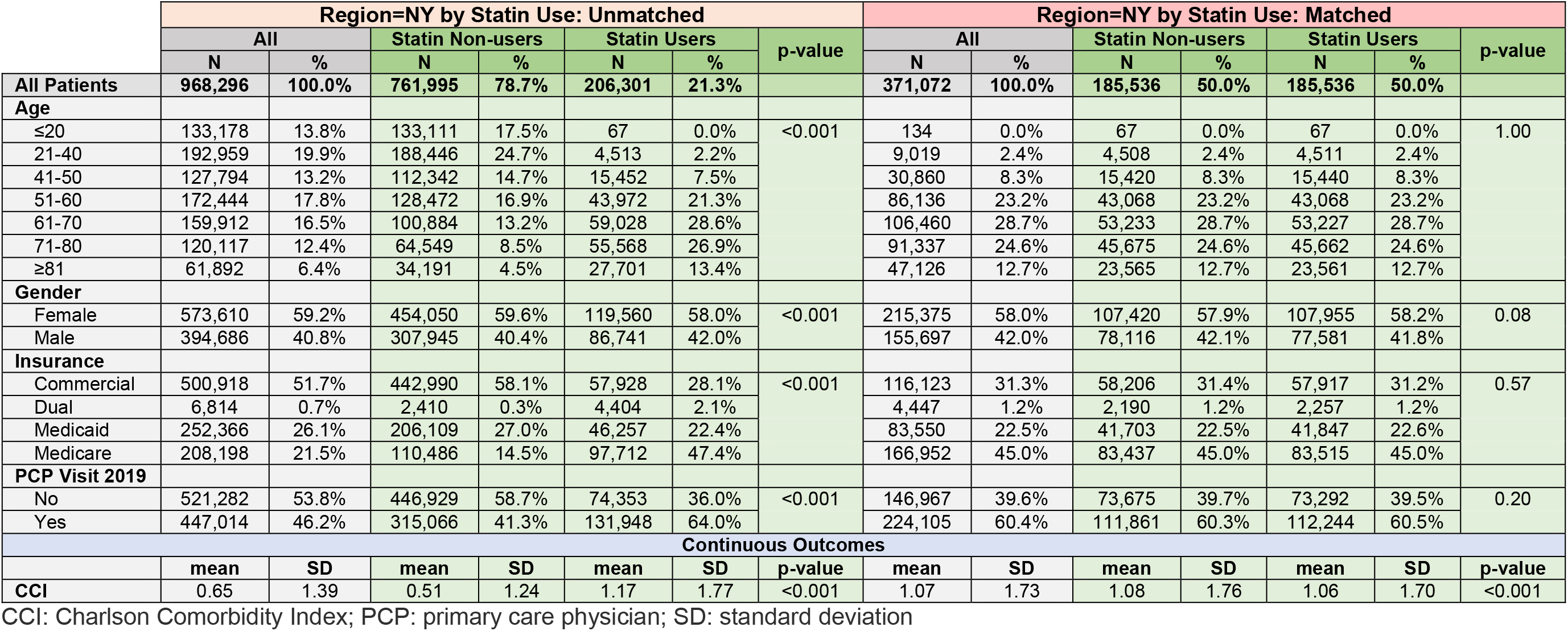
Statin Cohort (Region=New York State), Patient Characteristics Pre/Post Match

|  | Region=NY by Statin Use: Unmatched |  |  |  |  |  |  | Region=NY by Statin Use: Matched |  |  |  |  |  |  |
| --- | --- | --- | --- | --- | --- | --- | --- | --- | --- | --- | --- | --- | --- | --- |
|  | All |  | Statin Non-users |  | Statin Users |  | p-value | All |  | Statin Non-users |  | Statin Users |  | p-value |
|  | N | % | N | % | N | % |  | N | % | N | % | N | % |  |
| All Patients | 968,296 | 100.0% | 761,995 | 78.7% | 206,301 | 21.3% |  | 371,072 | 100.0% | 185,536 | 50.0% | 185,536 | 50.0% |  |
| Age |  |  |  |  |  |  |  |  |  |  |  |  |  |  |
| ≤20 | 133,178 | 13.8% | 133,111 | 17.5% | 67 | 0.0% | <0.001 | 134 | 0.0% | 67 | 0.0% | 67 | 0.0% | 1.00 |
| 21-40 | 192,959 | 19.9% | 188,446 | 24.7% | 4,513 | 2.2% |  | 9,019 | 2.4% | 4,508 | 2.4% | 4,511 | 2.4% |  |
| 41-50 | 127,794 | 13.2% | 112,342 | 14.7% | 15,452 | 7.5% |  | 30,860 | 8.3% | 15,420 | 8.3% | 15,440 | 8.3% |  |
| 51-60 | 172,444 | 17.8% | 128,472 | 16.9% | 43,972 | 21.3% |  | 86,136 | 23.2% | 43,068 | 23.2% | 43,068 | 23.2% |  |
| 61-70 | 159,912 | 16.5% | 100,884 | 13.2% | 59,028 | 28.6% |  | 106,460 | 28.7% | 53,233 | 28.7% | 53,227 | 28.7% |  |
| 71-80 | 120,117 | 12.4% | 64,549 | 8.5% | 55,568 | 26.9% |  | 91,337 | 24.6% | 45,675 | 24.6% | 45,662 | 24.6% |  |
| ≥81 | 61,892 | 6.4% | 34,191 | 4.5% | 27,701 | 13.4% |  | 47,126 | 12.7% | 23,565 | 12.7% | 23,561 | 12.7% |  |
| Gender |  |  |  |  |  |  |  |  |  |  |  |  |  |  |
| Female | 573,610 | 59.2% | 454,050 | 59.6% | 119,560 | 58.0% | <0.001 | 215,375 | 58.0% | 107,420 | 57.9% | 107,955 | 58.2% | 0.08 |
| Male | 394,686 | 40.8% | 307,945 | 40.4% | 86,741 | 42.0% |  | 155,697 | 42.0% | 78,116 | 42.1% | 77,581 | 41.8% |  |
| Insurance |  |  |  |  |  |  |  |  |  |  |  |  |  |  |
| Commercial | 500,918 | 51.7% | 442,990 | 58.1% | 57,928 | 28.1% | <0.001 | 116,123 | 31.3% | 58,206 | 31.4% | 57,917 | 31.2% | 0.57 |
| Dual | 6,814 | 0.7% | 2,410 | 0.3% | 4,404 | 2.1% |  | 4,447 | 1.2% | 2,190 | 1.2% | 2,257 | 1.2% |  |
| Medicaid | 252,366 | 26.1% | 206,109 | 27.0% | 46,257 | 22.4% |  | 83,550 | 22.5% | 41,703 | 22.5% | 41,847 | 22.6% |  |
| Medicare | 208,198 | 21.5% | 110,486 | 14.5% | 97,712 | 47.4% |  | 166,952 | 45.0% | 83,437 | 45.0% | 83,515 | 45.0% |  |
| PCP Visit 2019 |  |  |  |  |  |  |  |  |  |  |  |  |  |  |
| No | 521,282 | 53.8% | 446,929 | 58.7% | 74,353 | 36.0% | <0.001 | 146,967 | 39.6% | 73,675 | 39.7% | 73,292 | 39.5% | 0.20 |
| Yes | 447,014 | 46.2% | 315,066 | 41.3% | 131,948 | 64.0% |  | 224,105 | 60.4% | 111,861 | 60.3% | 112,244 | 60.5% |  |
| Continuous Outcomes |  |  |  |  |  |  |  |  |  |  |  |  |  |  |
|  | mean | SD | mean | SD | mean | SD | p-value | mean | SD | mean | SD | mean | SD | p-value |
| CCI | 0.65 | 1.39 | 0.51 | 1.24 | 1.17 | 1.77 | <0.001 | 1.07 | 1.73 | 1.08 | 1.76 | 1.06 | 1.70 | <0.001 |

CCI: Charlson Comorbidity Index; PCP: primary care physician; SD: standard deviation

**Table S8c:** Statin User Cohort (All Regions) by BP Use, Patient Characteristics Pre/Post Match of BP Users/Non-users

|  | All Statin Users by BP: Unmatched |  |  |  |  |  |  | All Statin Users by BP: Matched |  |  |  |  |  |  |
| --- | --- | --- | --- | --- | --- | --- | --- | --- | --- | --- | --- | --- | --- | --- |
|  | All |  | BP Non-user |  | BP User |  | p-value | All |  | BP Non-user |  | BP User |  | p-value |
|  | N | % | N | % | N | % |  | N | % | N | % | N | % |  |
| All Patients | 1,436,300 | 100.0% | 1,218,319 | 84.8% | 217,981 | 15.2% |  | 426,960 | 100.0% | 213,480 | 50.0% | 213,480 | 50.0% |  |
| Age |  |  |  |  |  |  |  |  |  |  |  |  |  |  |
| ≤20 | 1,385 | 0.1% | 1,365 | 0.1% | 20 | 0.0% | <0.001 | 42 | 0.0% | 22 | 0.0% | 20 | 0.0% | 1.00 |
| 21-40 | 44,389 | 3.1% | 44,042 | 3.6% | 347 | 0.2% |  | 704 | 0.2% | 357 | 0.2% | 347 | 0.2% |  |
| 41-50 | 135,867 | 9.5% | 133,850 | 11.0% | 2,017 | 0.9% |  | 4,033 | 0.9% | 2,016 | 0.9% | 2,017 | 0.9% |  |
| 51-60 | 356,032 | 24.8% | 333,325 | 27.4% | 22,707 | 10.4% |  | 45,439 | 10.6% | 22,732 | 10.6% | 22,707 | 10.6% |  |
| 61-70 | 428,943 | 29.9% | 356,208 | 29.2% | 72,735 | 33.4% |  | 144,861 | 33.9% | 72,341 | 33.9% | 72,520 | 34.0% |  |
| 71-80 | 302,081 | 21.0% | 223,651 | 18.4% | 78,430 | 36.0% |  | 150,527 | 35.3% | 75,316 | 35.3% | 75,211 | 35.2% |  |
| ≥81 | 167,603 | 11.7% | 125,878 | 10.3% | 41,725 | 19.1% |  | 81,354 | 19.1% | 40,696 | 19.1% | 40,658 | 19.0% |  |
| Gender |  |  |  |  |  |  |  |  |  |  |  |  |  |  |
| Female | 843,147 | 58.7% | 646,846 | 53.1% | 196,301 | 90.1% | <0.001 | 383,586 | 89.8% | 191,786 | 89.8% | 191,800 | 89.8% | 0.94 |
| Male | 593,153 | 41.3% | 571,473 | 46.9% | 21,680 | 9.9% |  | 43,374 | 10.2% | 21,694 | 10.2% | 21,680 | 10.2% |  |
| Region |  |  |  |  |  |  |  |  |  |  |  |  |  |  |
| Midwest | 271,319 | 18.9% | 237,718 | 19.5% | 33,601 | 15.4% | <0.001 | 67,050 | 15.7% | 33,525 | 15.7% | 33,525 | 15.7% | 1.00 |
| Northeast | 423,934 | 29.5% | 366,936 | 30.1% | 56,998 | 26.1% |  | 113,308 | 26.5% | 56,654 | 26.5% | 56,654 | 26.5% |  |
| South | 523,112 | 36.4% | 442,996 | 36.4% | 80,116 | 36.8% |  | 157,838 | 37.0% | 78,919 | 37.0% | 78,919 | 37.0% |  |
| West | 217,935 | 15.2% | 170,669 | 14.0% | 47,266 | 21.7% |  | 88,764 | 20.8% | 44,382 | 20.8% | 44,382 | 20.8% |  |
| Insurance |  |  |  |  |  |  |  |  |  |  |  |  |  |  |
| Commercial | 587,625 | 40.9% | 533,843 | 43.8% | 53,782 | 24.7% | <0.001 | 107,552 | 25.2% | 53,774 | 25.2% | 53,778 | 25.2% | 1.00 |
| Dual | 55,356 | 3.9% | 42,041 | 3.5% | 13,315 | 6.1% |  | 24,380 | 5.7% | 12,183 | 5.7% | 12,197 | 5.7% |  |
| Medicaid | 320,911 | 22.3% | 280,799 | 23.0% | 40,112 | 18.4% |  | 76,121 | 17.8% | 38,050 | 17.8% | 38,071 | 17.8% |  |
| Medicare | 472,408 | 32.9% | 361,636 | 29.7% | 110,772 | 50.8% |  | 218,907 | 51.3% | 109,473 | 51.3% | 109,434 | 51.3% |  |
| PCP Visit 2019 |  |  |  |  |  |  |  |  |  |  |  |  |  |  |
| No | 507,726 | 35.3% | 430,446 | 35.3% | 77,280 | 35.5% | 0.27 | 151,395 | 35.5% | 75,614 | 35.4% | 75,781 | 35.5% | 0.59 |
| Yes | 928,574 | 64.7% | 787,873 | 64.7% | 140,701 | 64.5% |  | 275,565 | 64.5% | 137,866 | 64.6% | 137,699 | 64.5% |  |
| Continuous Outcomes |  |  |  |  |  |  |  |  |  |  |  |  |  |  |
|  | mean | SD | mean | SD | mean | SD | p-value | mean | SD | mean | SD | mean | SD | p-value |
| CCI | 1.11 | 1.75 | 1.13 | 1.77 | 0.95 | 1.66 | <0.001 | 0.97 | 1.66 | 0.97 | 1.66 | 0.97 | 1.67 | 0.79 |

BP: bisphosphonate; CCI: Charlson Comorbidity Index; PCP: primary care physician; SD: standard deviation

**Table S8d:**
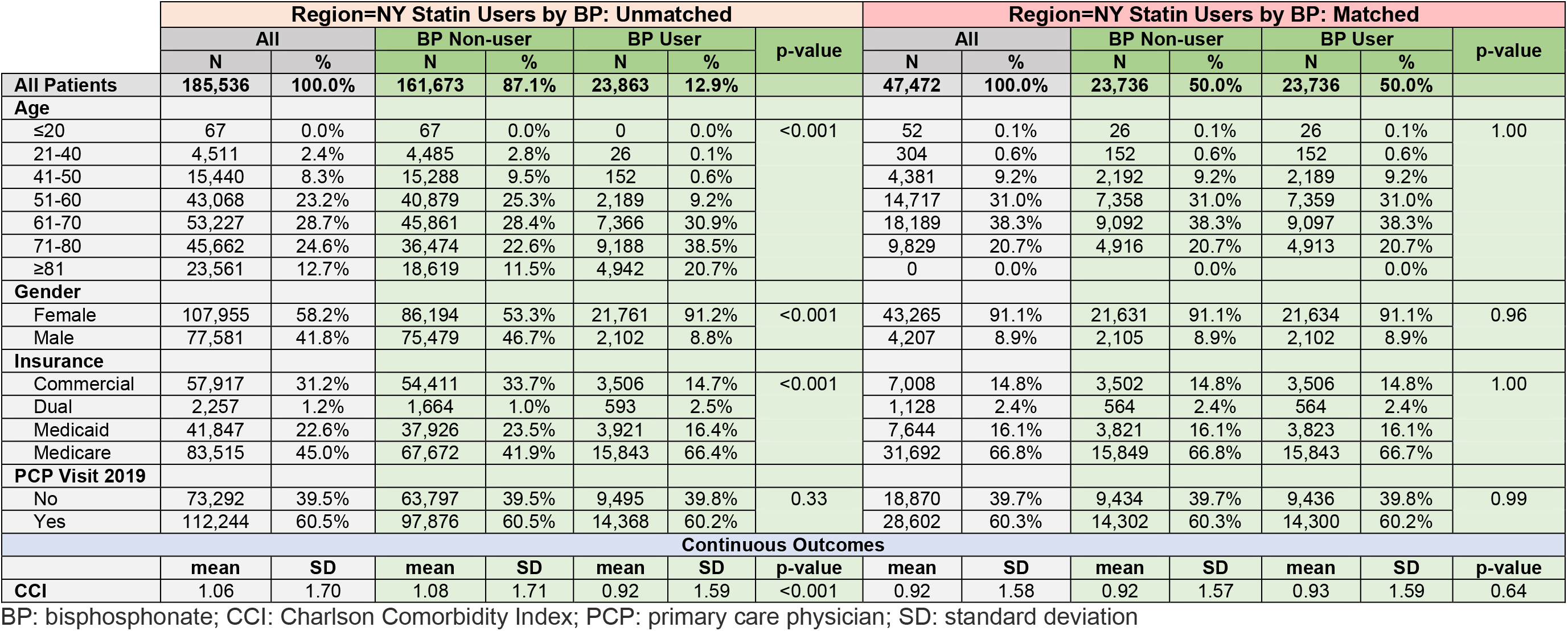
Statin User Cohort (Region=New York State) by BP Use, Patient Characteristics Pre/Post Match of BP Users/Non-users

**Table S8e:**
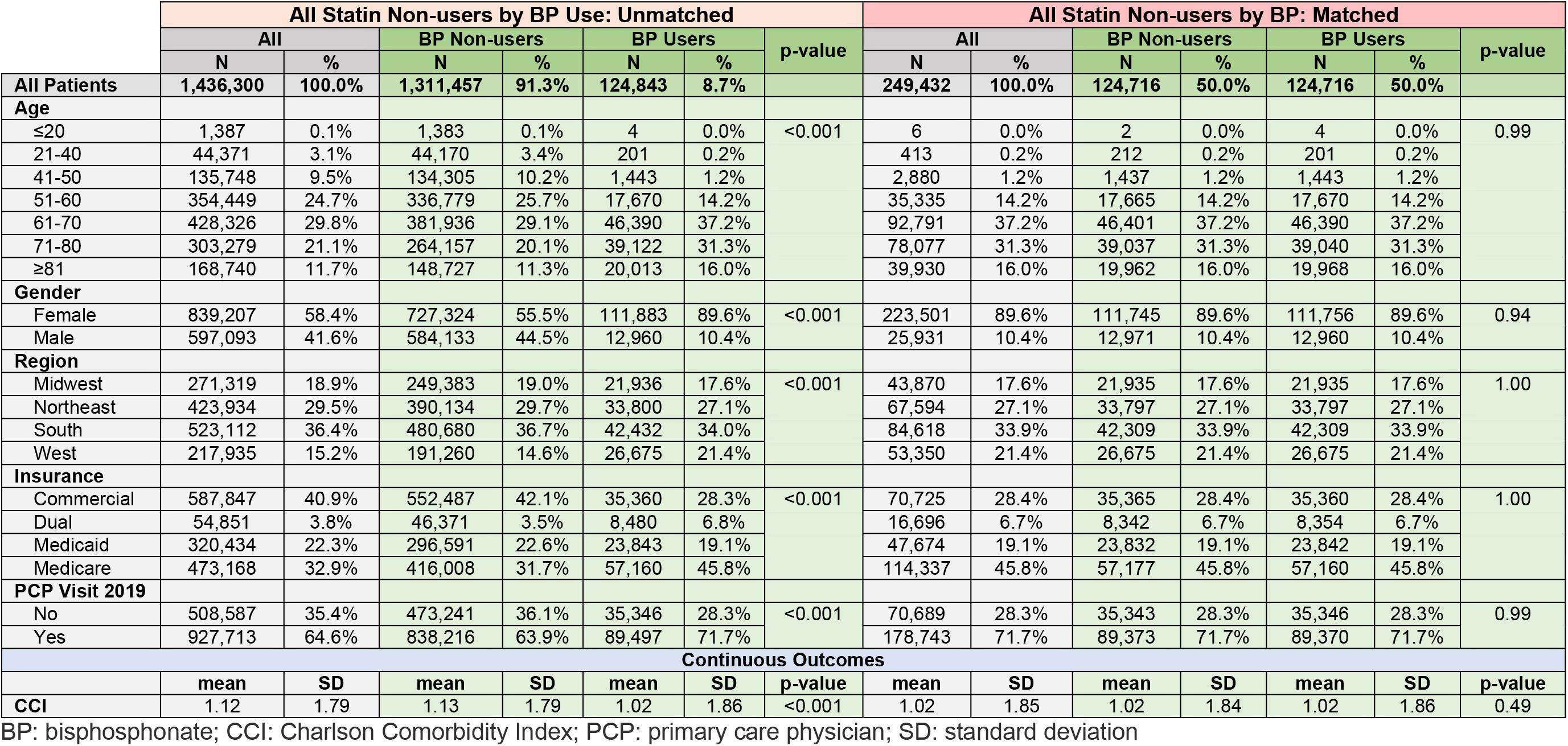
Statin Non-user Cohort (All Regions) by BP Use, Patient Characteristics Pre/Post Match of BP Users/Non-users

**Table S8f:** Statin Non-user Cohort (Region=New York State) by BP Use, Patient Characteristics Pre/Post Match of BP Users/Non-users

|  | Region=NY Statin Non-users by BP: Unmatched |  |  |  |  |  |  | Region=NY Statin Non-users by BP: Matched |  |  |  |  |  |  |
| --- | --- | --- | --- | --- | --- | --- | --- | --- | --- | --- | --- | --- | --- | --- |
|  | All |  | BP Non-users |  | BP Users |  | p-value | All |  | BP Non-users |  | BP Users |  | p-value |
|  | N | % | N | % | N | % |  | N | % | N | % | N | % |  |
| All Patients | 185,536 | 100.0% | 170,990 | 92.2% | 14,546 | 7.8% |  | 29,042 | 100.0% | 14,521 | 50.0% | 14,521 | 50.0% |  |
| Age |  |  |  |  |  |  |  |  |  |  |  |  |  |  |
| ≤20 | 67 | 0.0% | 67 | 0.0% | 0 | 0.0% | <0.001 | 0 | 0.0% | 0 | 0.0% | 0 | 0.0% | 1.00 |
| 21-40 | 4,508 | 2.4% | 4,498 | 2.6% | 10 | 0.1% |  | 23 | 0.1% | 13 | 0.1% | 10 | 0.1% |  |
| 41-50 | 15,420 | 8.3% | 15,314 | 9.0% | 106 | 0.7% |  | 211 | 0.7% | 105 | 0.7% | 106 | 0.7% |  |
| 51-60 | 43,068 | 23.2% | 41,317 | 24.2% | 1,751 | 12.0% |  | 3,502 | 12.1% | 1,751 | 12.1% | 1,751 | 12.1% |  |
| 61-70 | 53,233 | 28.7% | 48,148 | 28.2% | 5,085 | 35.0% |  | 10,174 | 35.0% | 5,089 | 35.0% | 5,085 | 35.0% |  |
| 71-80 | 45,675 | 24.6% | 40,731 | 23.8% | 4,944 | 34.0% |  | 9,877 | 34.0% | 4,937 | 34.0% | 4,940 | 34.0% |  |
| ≥81 | 23,565 | 12.7% | 20,915 | 12.2% | 2,650 | 18.2% |  | 5,255 | 18.1% | 2,626 | 18.1% | 2,629 | 18.1% |  |
| Gender |  |  |  |  |  |  |  |  |  |  |  |  |  |  |
| Female | 107,420 | 57.9% | 94,242 | 55.1% | 13,178 | 90.6% | <0.001 | 26,304 | 90.6% | 13,151 | 90.6% | 13,153 | 90.6% | 0.97 |
| Male | 78,116 | 42.1% | 76,748 | 44.9% | 1,368 | 9.4% |  | 2,738 | 9.4% | 1,370 | 9.4% | 1,368 | 9.4% |  |
| Insurance |  |  |  |  |  |  |  |  |  |  |  |  |  |  |
| Commercial | 58,206 | 31.4% | 56,313 | 32.9% | 1,893 | 13.0% | <0.001 | 3,785 | 13.0% | 1,892 | 13.0% | 1,893 | 13.0% | 0.96 |
| Dual | 2,190 | 1.2% | 1,754 | 1.0% | 436 | 3.0% |  | 883 | 3.0% | 449 | 3.1% | 434 | 3.0% |  |
| Medicaid | 41,703 | 22.5% | 38,177 | 22.3% | 3,526 | 24.2% |  | 6,994 | 24.1% | 3,491 | 24.0% | 3,503 | 24.1% |  |
| Medicare | 83,437 | 45.0% | 74,746 | 43.7% | 8,691 | 59.7% |  | 17,380 | 59.8% | 8,689 | 59.8% | 8,691 | 59.9% |  |
| PCP Visit 2019 |  |  |  |  |  |  |  |  |  |  |  |  |  |  |
| No | 73,675 | 39.7% | 69,382 | 40.6% | 4,293 | 29.5% | <0.001 | 8,564 | 29.5% | 4,280 | 29.5% | 4,284 | 29.5% | 0.96 |
| Yes | 111,861 | 60.3% | 101,608 | 59.4% | 10,253 | 70.5% |  | 20,478 | 70.5% | 10,241 | 70.5% | 10,237 | 70.5% |  |
| Continuous Outcomes |  |  |  |  |  |  |  |  |  |  |  |  |  |  |
|  | mean | SD | mean | SD | mean | SD | p-value | mean | SD | mean | SD | mean | SD | p-value |
| CCI | 1.08 | 1.76 | 1.09 | 1.76 | 0.95 | 1.75 | <0.001 | 0.95 | 1.74 | 0.95 | 1.73 | 0.95 | 1.75 | 0.82 |

BP: bisphosphonate; CCI: Charlson Comorbidity Index; PCP: primary care physician; SD: standard deviation

**Table S9a:**
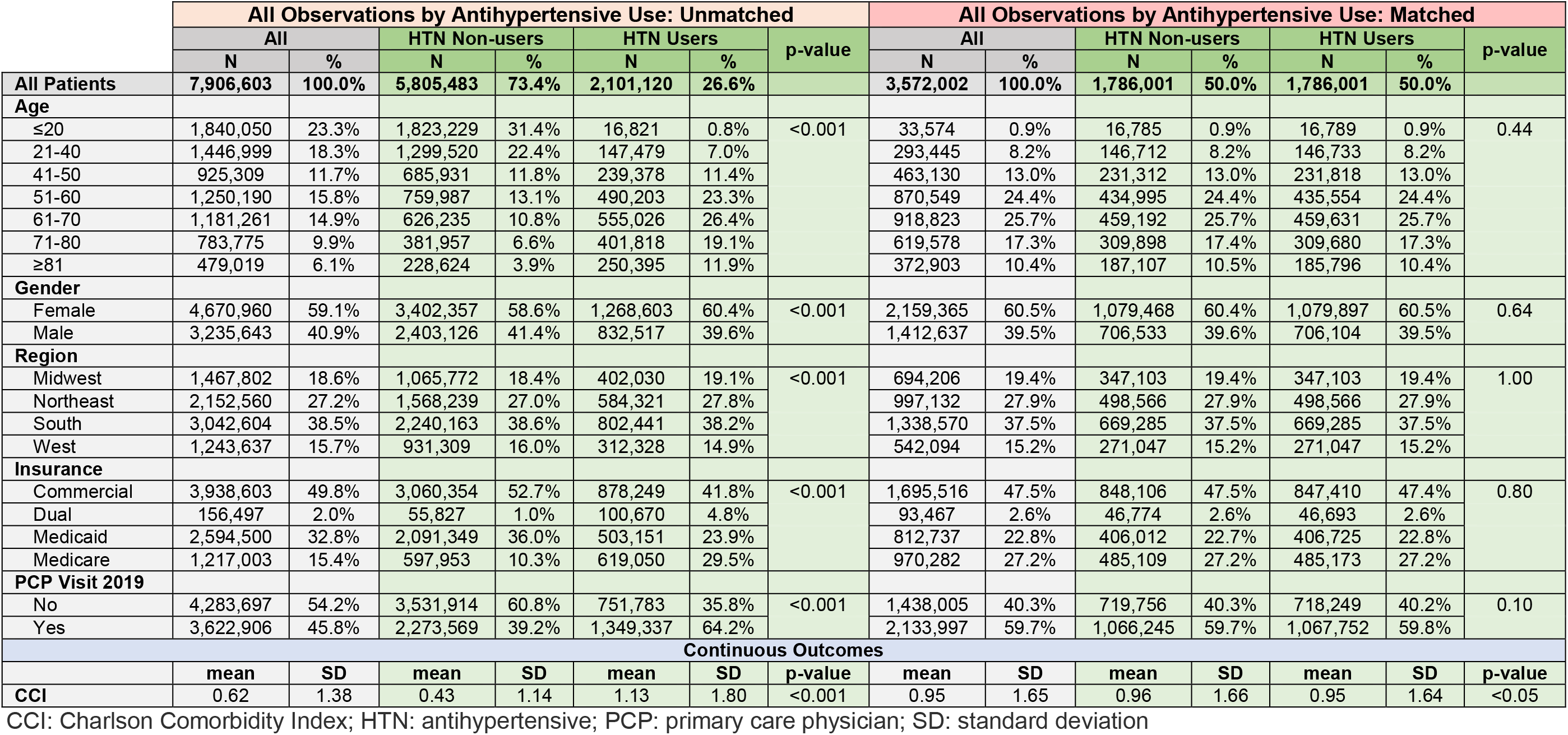
Antihypertensive Cohort (All Regions), Patient Characteristics Pre/Post Match

|  | All Observations by Antihypertensive Use: Unmatched |  |  |  |  |  |  | All Observations by Antihypertensive Use: Matched |  |  |  |  |  |  |
| --- | --- | --- | --- | --- | --- | --- | --- | --- | --- | --- | --- | --- | --- | --- |
|  | All |  | HTN Non-users |  | HTN Users |  | p-value | All |  | HTN Non-users |  | HTN Users |  | p-value |
|  | N | % | N | % | N | % |  | N | % | N | % | N | % |  |
| All Patients | 7,906,603 | 100.0% | 5,805,483 | 73.4% | 2,101,120 | 26.6% |  | 3,572,002 | 100.0% | 1,786,001 | 50.0% | 1,786,001 | 50.0% |  |
| Age |  |  |  |  |  |  |  |  |  |  |  |  |  |  |
| ≤20 | 1,840,050 | 23.3% | 1,823,229 | 31.4% | 16,821 | 0.8% | <0.001 | 33,574 | 0.9% | 16,785 | 0.9% | 16,789 | 0.9% | 0.44 |
| 21-40 | 1,446,999 | 18.3% | 1,299,520 | 22.4% | 147,479 | 7.0% |  | 293,445 | 8.2% | 146,712 | 8.2% | 146,733 | 8.2% |  |
| 41-50 | 925,309 | 11.7% | 685,931 | 11.8% | 239,378 | 11.4% |  | 463,130 | 13.0% | 231,312 | 13.0% | 231,818 | 13.0% |  |
| 51-60 | 1,250,190 | 15.8% | 759,987 | 13.1% | 490,203 | 23.3% |  | 870,549 | 24.4% | 434,995 | 24.4% | 435,554 | 24.4% |  |
| 61-70 | 1,181,261 | 14.9% | 626,235 | 10.8% | 555,026 | 26.4% |  | 918,823 | 25.7% | 459,192 | 25.7% | 459,631 | 25.7% |  |
| 71-80 | 783,775 | 9.9% | 381,957 | 6.6% | 401,818 | 19.1% |  | 619,578 | 17.3% | 309,898 | 17.4% | 309,680 | 17.3% |  |
| ≥81 | 479,019 | 6.1% | 228,624 | 3.9% | 250,395 | 11.9% |  | 372,903 | 10.4% | 187,107 | 10.5% | 185,796 | 10.4% |  |
| Gender |  |  |  |  |  |  |  |  |  |  |  |  |  |  |
| Female | 4,670,960 | 59.1% | 3,402,357 | 58.6% | 1,268,603 | 60.4% | <0.001 | 2,159,365 | 60.5% | 1,079,468 | 60.4% | 1,079,897 | 60.5% | 0.64 |
| Male | 3,235,643 | 40.9% | 2,403,126 | 41.4% | 832,517 | 39.6% |  | 1,412,637 | 39.5% | 706,533 | 39.6% | 706,104 | 39.5% |  |
| Region |  |  |  |  |  |  |  |  |  |  |  |  |  |  |
| Midwest | 1,467,802 | 18.6% | 1,065,772 | 18.4% | 402,030 | 19.1% | <0.001 | 694,206 | 19.4% | 347,103 | 19.4% | 347,103 | 19.4% | 1.00 |
| Northeast | 2,152,560 | 27.2% | 1,568,239 | 27.0% | 584,321 | 27.8% |  | 997,132 | 27.9% | 498,566 | 27.9% | 498,566 | 27.9% |  |
| South | 3,042,604 | 38.5% | 2,240,163 | 38.6% | 802,441 | 38.2% |  | 1,338,570 | 37.5% | 669,285 | 37.5% | 669,285 | 37.5% |  |
| West | 1,243,637 | 15.7% | 931,309 | 16.0% | 312,328 | 14.9% |  | 542,094 | 15.2% | 271,047 | 15.2% | 271,047 | 15.2% |  |
| Insurance |  |  |  |  |  |  |  |  |  |  |  |  |  |  |
| Commercial | 3,938,603 | 49.8% | 3,060,354 | 52.7% | 878,249 | 41.8% | <0.001 | 1,695,516 | 47.5% | 848,106 | 47.5% | 847,410 | 47.4% | 0.80 |
| Dual | 156,497 | 2.0% | 55,827 | 1.0% | 100,670 | 4.8% |  | 93,467 | 2.6% | 46,774 | 2.6% | 46,693 | 2.6% |  |
| Medicaid | 2,594,500 | 32.8% | 2,091,349 | 36.0% | 503,151 | 23.9% |  | 812,737 | 22.8% | 406,012 | 22.7% | 406,725 | 22.8% |  |
| Medicare | 1,217,003 | 15.4% | 597,953 | 10.3% | 619,050 | 29.5% |  | 970,282 | 27.2% | 485,109 | 27.2% | 485,173 | 27.2% |  |
| PCP Visit 2019 |  |  |  |  |  |  |  |  |  |  |  |  |  |  |
| No | 4,283,697 | 54.2% | 3,531,914 | 60.8% | 751,783 | 35.8% | <0.001 | 1,438,005 | 40.3% | 719,756 | 40.3% | 718,249 | 40.2% | 0.10 |
| Yes | 3,622,906 | 45.8% | 2,273,569 | 39.2% | 1,349,337 | 64.2% |  | 2,133,997 | 59.7% | 1,066,245 | 59.7% | 1,067,752 | 59.8% |  |
| Continuous Outcomes |  |  |  |  |  |  |  |  |  |  |  |  |  |  |
|  | mean | SD | mean | SD | mean | SD | p-value | mean | SD | mean | SD | mean | SD | p-value |
| CCI | 0.62 | 1.38 | 0.43 | 1.14 | 1.13 | 1.80 | <0.001 | 0.95 | 1.65 | 0.96 | 1.66 | 0.95 | 1.64 | <0.05 |

CCI: Charlson Comorbidity Index; HTN: antihypertensive; PCP: primary care physician; SD: standard deviation

**Table S9b:** Antihypertensive Cohort (Region=New York State), Patient Characteristics Pre/Post Match

|  | Region=NY by Antihypertensive Use: Unmatched |  |  |  |  |  |  | Region=NY by Antihypertensive Use: Matched |  |  |  |  |  |  |
| --- | --- | --- | --- | --- | --- | --- | --- | --- | --- | --- | --- | --- | --- | --- |
|  | All |  | HTN Non-users |  | HTN Users |  | p-value | All |  | HTN Non-users |  | HTN Users |  | p-value |
|  | N | % | N | % | N | % |  | N | % | N | % | N | % |  |
| All Patients | 968,296 | 100.0% | 709,644 | 73.3% | 258,652 | 26.7% |  | 407,248 | 100.0% | 203,624 | 50.0% | 203,624 | 50.0% |  |
| Age |  |  |  |  |  |  |  |  |  |  |  |  |  |  |
| ≤20 | 133,178 | 13.8% | 132,352 | 18.7% | 826 | 0.3% | <0.001 | 1,622 | 0.4% | 811 | 0.4% | 811 | 0.4% | 1.00 |
| 21-40 | 192,959 | 19.9% | 181,447 | 25.6% | 11,512 | 4.5% |  | 22,930 | 5.6% | 11,465 | 5.6% | 11,465 | 5.6% |  |
| 41-50 | 127,794 | 13.2% | 105,490 | 14.9% | 22,304 | 8.6% |  | 43,846 | 10.8% | 21,923 | 10.8% | 21,923 | 10.8% |  |
| 51-60 | 172,444 | 17.8% | 119,643 | 16.9% | 52,801 | 20.4% |  | 96,318 | 23.7% | 48,159 | 23.7% | 48,159 | 23.7% |  |
| 61-70 | 159,912 | 16.5% | 92,103 | 13.0% | 67,809 | 26.2% |  | 109,858 | 27.0% | 54,929 | 27.0% | 54,929 | 27.0% |  |
| 71-80 | 120,117 | 12.4% | 54,076 | 7.6% | 66,041 | 25.5% |  | 88,734 | 21.8% | 44,367 | 21.8% | 44,367 | 21.8% |  |
| ≥81 | 61,892 | 6.4% | 24,533 | 3.5% | 37,359 | 14.4% |  | 43,940 | 10.8% | 21,970 | 10.8% | 21,970 | 10.8% |  |
| Gender |  |  |  |  |  |  |  |  |  |  |  |  |  |  |
| Female | 573,610 | 59.2% | 419,901 | 59.2% | 153,709 | 59.4% | 0.02 | 240,930 | 59.2% | 120,465 | 59.2% | 120,465 | 59.2% | 1.00 |
| Male | 394,686 | 40.8% | 289,743 | 40.8% | 104,943 | 40.6% |  | 166,318 | 40.8% | 83,159 | 40.8% | 83,159 | 40.8% |  |
| Insurance |  |  |  |  |  |  |  |  |  |  |  |  |  |  |
| Commercial | 500,918 | 51.7% | 425,181 | 59.9% | 75,737 | 29.3% | <0.001 | 150,918 | 37.1% | 75,459 | 37.1% | 75,459 | 37.1% | 1.00 |
| Dual | 6,814 | 0.7% | 1,659 | 0.2% | 5,155 | 2.0% |  | 2,986 | 0.7% | 1,493 | 0.7% | 1,493 | 0.7% |  |
| Medicaid | 252,366 | 26.1% | 193,207 | 27.2% | 59,159 | 22.9% |  | 95,032 | 23.3% | 47,516 | 23.3% | 47,516 | 23.3% |  |
| Medicare | 208,198 | 21.5% | 89,597 | 12.6% | 118,601 | 45.9% |  | 158,312 | 38.9% | 79,156 | 38.9% | 79,156 | 38.9% |  |
| PCP Visit 2019 |  |  |  |  |  |  |  |  |  |  |  |  |  |  |
| No | 521,282 | 53.8% | 423,952 | 59.7% | 97,330 | 37.6% | <0.001 | 181,234 | 44.5% | 90,617 | 44.5% | 90,617 | 44.5% | 1.00 |
| Yes | 447,014 | 46.2% | 285,692 | 40.3% | 161,322 | 62.4% |  | 226,014 | 55.5% | 113,007 | 55.5% | 113,007 | 55.5% |  |
| Continuous Outcomes |  |  |  |  |  |  |  |  |  |  |  |  |  |  |
|  | mean | SD | mean | SD | mean | SD | p-value | mean | SD | mean | SD | mean | SD | p-value |
| CCI | 0.65 | 1.39 | 0.46 | 1.16 | 1.17 | 1.80 | <0.001 | 0.95 | 1.60 | 0.95 | 1.60 | 0.95 | 1.60 | 1.00 |

CCI: Charlson Comorbidity Index; HTN: antihypertensive; PCP: primary care physician; SD: standard deviation

**Table S9c:** Antihypertensive User Cohort (All Regions) by BP Use, Patient Characteristics Pre/Post Match of BP Users/Non-users

|  | All Antihypertensive Users by BP: Unmatched |  |  |  |  |  |  | All Antihypertensive Users by BP: Matched |  |  |  |  |  |  |
| --- | --- | --- | --- | --- | --- | --- | --- | --- | --- | --- | --- | --- | --- | --- |
|  | All |  | BP Non-user |  | BP User |  | p-value | All |  | BP Non-user |  | BP User |  | p-value |
|  | N | % | N | % | N | % |  | N | % | N | % | N | % |  |
| All Patients | 1,786,001 | 100.0% | 1,579,388 | 88.4% | 206,613 | 11.6% |  | 408,792 | 100.0% | 204,396 | 50.0% | 204,396 | 50.0% |  |
| Age |  |  |  |  |  |  |  |  |  |  |  |  |  |  |
| ≤20 | 16,789 | 0.9% | 16,586 | 1.1% | 203 | 0.1% | <0.001 | 411 | 0.1% | 208 | 0.1% | 203 | 0.1% | 1.00 |
| 21-40 | 146,733 | 8.2% | 145,872 | 9.2% | 861 | 0.4% |  | 1,728 | 0.4% | 868 | 0.4% | 860 | 0.4% |  |
| 41-50 | 231,818 | 13.0% | 229,150 | 14.5% | 2,668 | 1.3% |  | 5,333 | 1.3% | 2,667 | 1.3% | 2,666 | 1.3% |  |
| 51-60 | 435,554 | 24.4% | 413,155 | 26.2% | 22,399 | 10.8% |  | 44,796 | 11.0% | 22,399 | 11.0% | 22,397 | 11.0% |  |
| 61-70 | 459,631 | 25.7% | 390,664 | 24.7% | 68,967 | 33.4% |  | 137,730 | 33.7% | 68,862 | 33.7% | 68,868 | 33.7% |  |
| 71-80 | 309,680 | 17.3% | 237,749 | 15.1% | 71,931 | 34.8% |  | 140,882 | 34.5% | 70,439 | 34.5% | 70,443 | 34.5% |  |
| ≥81 | 185,796 | 10.4% | 146,212 | 9.3% | 39,584 | 19.2% |  | 77,912 | 19.1% | 38,953 | 19.1% | 38,959 | 19.1% |  |
| Gender |  |  |  |  |  |  |  |  |  |  |  |  |  |  |
| Female | 1,079,897 | 60.5% | 894,472 | 56.6% | 185,425 | 89.7% | <0.001 | 366,424 | 89.6% | 183,212 | 89.6% | 183,212 | 89.6% | 1.00 |
| Male | 706,104 | 39.5% | 684,916 | 43.4% | 21,188 | 10.3% |  | 42,368 | 10.4% | 21,184 | 10.4% | 21,184 | 10.4% |  |
| Region |  |  |  |  |  |  |  |  |  |  |  |  |  |  |
| Midwest | 347,103 | 19.4% | 313,523 | 19.9% | 33,580 | 16.3% | <0.001 | 67,058 | 16.4% | 33,529 | 16.4% | 33,529 | 16.4% | 1.00 |
| Northeast | 498,566 | 27.9% | 444,828 | 28.2% | 53,738 | 26.0% |  | 107,150 | 26.2% | 53,575 | 26.2% | 53,575 | 26.2% |  |
| South | 669,285 | 37.5% | 595,410 | 37.7% | 73,875 | 35.8% |  | 146,890 | 35.9% | 73,445 | 35.9% | 73,445 | 35.9% |  |
| West | 271,047 | 15.2% | 225,627 | 14.3% | 45,420 | 22.0% |  | 87,694 | 21.5% | 43,847 | 21.5% | 43,847 | 21.5% |  |
| Insurance |  |  |  |  |  |  |  |  |  |  |  |  |  |  |
| Commercial | 847,410 | 47.4% | 787,519 | 49.9% | 59,891 | 29.0% | <0.001 | 119,737 | 29.3% | 59,863 | 29.3% | 59,874 | 29.3% | 1.00 |
| Dual | 46,693 | 2.6% | 37,153 | 2.4% | 9,540 | 4.6% |  | 17,884 | 4.4% | 8,945 | 4.4% | 8,939 | 4.4% |  |
| Medicaid | 406,725 | 22.8% | 369,893 | 23.4% | 36,832 | 17.8% |  | 70,769 | 17.3% | 35,387 | 17.3% | 35,382 | 17.3% |  |
| Medicare | 485,173 | 27.2% | 384,823 | 24.4% | 100,350 | 48.6% |  | 200,402 | 49.0% | 100,201 | 49.0% | 100,201 | 49.0% |  |
| PCP Visit 2019 |  |  |  |  |  |  |  |  |  |  |  |  |  |  |
| No | 718,249 | 40.2% | 633,042 | 40.1% | 85,207 | 41.2% | <0.001 | 168,255 | 41.2% | 84,128 | 41.2% | 84,127 | 41.2% | 1.00 |
| Yes | 1,067,752 | 59.8% | 946,346 | 59.9% | 121,406 | 58.8% |  | 240,537 | 58.8% | 120,268 | 58.8% | 120,269 | 58.8% |  |
| Continuous Outcomes |  |  |  |  |  |  |  |  |  |  |  |  |  |  |
|  | mean | SD | mean | SD | mean | SD | p-value | mean | SD | mean | SD | mean | SD | p-value |
| CCI | 0.95 | 1.64 | 0.95 | 1.64 | 0.94 | 1.68 | 0.02 | 0.95 | 1.67 | 0.95 | 1.67 | 0.95 | 1.68 | 0.68 |

BP: bisphosphonate; CCI: Charlson Comorbidity Index; PCP: primary care physician; SD: standard deviation

**Table S9d:** Antihypertensive User Cohort (Region=New York State) by BP Use, Patient Characteristics Pre/Post Match of BP Users/Non-users

|  | Region=NY Antihypertensive Users by BP: Unmatched |  |  |  |  |  |  | Region=NY Antihypertensive Users by BP: Matched |  |  |  |  |  |  |
| --- | --- | --- | --- | --- | --- | --- | --- | --- | --- | --- | --- | --- | --- | --- |
|  | All |  | BP Non-user |  | BP User |  | p-value | All |  | BP Non-user |  | BP User |  | p-value |
|  | N | % | N | % | N | % |  | N | % | N | % | N | % |  |
| All Patients | 203,624 | 100.0% | 182,411 | 89.6% | 21,213 | 10.4% |  | 42,252 | 100.0% | 21,126 | 50.0% | 21,126 | 50.0% |  |
| Age |  |  |  |  |  |  |  |  |  |  |  |  |  |  |
| ≤20 | 811 | 0.4% | 798 | 0.4% | 13 | 0.1% | <0.001 | 27 | 0.1% | 14 | 0.1% | 13 | 0.1% | 1.00 |
| 21-40 | 11,465 | 5.6% | 11,396 | 6.2% | 69 | 0.3% |  | 137 | 0.3% | 68 | 0.3% | 69 | 0.3% |  |
| 41-50 | 21,923 | 10.8% | 21,747 | 11.9% | 176 | 0.8% |  | 354 | 0.8% | 178 | 0.8% | 176 | 0.8% |  |
| 51-60 | 48,159 | 23.7% | 46,047 | 25.2% | 2,112 | 10.0% |  | 4,218 | 10.0% | 2,108 | 10.0% | 2,110 | 10.0% |  |
| 61-70 | 54,929 | 27.0% | 48,022 | 26.3% | 6,907 | 32.6% |  | 13,804 | 32.7% | 6,902 | 32.7% | 6,902 | 32.7% |  |
| 71-80 | 44,367 | 21.8% | 36,409 | 20.0% | 7,958 | 37.5% |  | 15,777 | 37.3% | 7,886 | 37.3% | 7,891 | 37.4% |  |
| ≥81 | 21,970 | 10.8% | 17,992 | 9.9% | 3,978 | 18.8% |  | 7,935 | 18.8% | 3,970 | 18.8% | 3,965 | 18.8% |  |
| Gender |  |  |  |  |  |  |  |  |  |  |  |  |  |  |
| Female | 120,465 | 59.2% | 101,190 | 55.5% | 19,275 | 90.9% | <0.001 | 38,380 | 90.8% | 19,190 | 90.8% | 19,190 | 90.8% | 1.00 |
| Male | 83,159 | 40.8% | 81,221 | 44.5% | 1,938 | 9.1% |  | 3,872 | 9.2% | 1,936 | 9.2% | 1,936 | 9.2% |  |
| Insurance |  |  |  |  |  |  |  |  |  |  |  |  |  |  |
| Commercial | 75,459 | 37.1% | 71,460 | 39.2% | 3,999 | 18.9% | <0.001 | 7,993 | 18.9% | 3,997 | 18.9% | 3,996 | 18.9% | 1.00 |
| Dual | 1,493 | 0.7% | 1,151 | 0.6% | 342 | 1.6% |  | 643 | 1.5% | 322 | 1.5% | 321 | 1.5% |  |
| Medicaid | 47,516 | 23.3% | 44,248 | 24.3% | 3,268 | 15.4% |  | 6,414 | 15.2% | 3,207 | 15.2% | 3,207 | 15.2% |  |
| Medicare | 79,156 | 38.9% | 65,552 | 35.9% | 13,604 | 64.1% |  | 27,202 | 64.4% | 13,600 | 64.4% | 13,602 | 64.4% |  |
| PCP Visit 2019 |  |  |  |  |  |  |  |  |  |  |  |  |  |  |
| No | 90,617 | 44.5% | 80,739 | 44.3% | 9,878 | 46.6% | <0.001 | 19,672 | 46.6% | 9,837 | 46.6% | 9,835 | 46.6% | 0.98 |
| Yes | 113,007 | 55.5% | 101,672 | 55.7% | 11,335 | 53.4% |  | 22,580 | 53.4% | 11,289 | 53.4% | 11,291 | 53.4% |  |
| Continuous Outcomes |  |  |  |  |  |  |  |  |  |  |  |  |  |  |
|  | mean | SD | mean | SD | mean | SD | p-value | mean | SD | mean | SD | mean | SD | p-value |
| CCI | 0.95 | 1.60 | 0.95 | 1.61 | 0.88 | 1.54 | <0.001 | 0.87 | 1.53 | 0.87 | 1.52 | 0.87 | 1.53 | 0.87 |

BP: bisphosphonate; CCI: Charlson Comorbidity Index; PCP: primary care physician; SD: standard deviation

**Table S9e:** Antihypertensive Non-user Cohort (All Regions) by BP Use, Patient Characteristics Pre/Post Match of BP Users/Non-users

|  | All Antihypertensive Non-users by BP: Unmatched |  |  |  |  |  |  | All Antihypertensive Non-users by BP: Matched |  |  |  |  |  |  |
| --- | --- | --- | --- | --- | --- | --- | --- | --- | --- | --- | --- | --- | --- | --- |
|  | All |  | BP Non-user |  | BP User |  | p-value | All |  | BP Non-user |  | BP User |  | p-value |
|  | N | % | N | % | N | % |  | N | % | N | % | N | % |  |
| All Patients | 1,786,001 | 100.0% | 1,649,985 | 92.4% | 136,016 | 7.6% |  | 271,448 | 100.0% | 135,724 | 50.0% | 135,724 | 50.0% |  |
| Age |  |  |  |  |  |  |  |  |  |  |  |  |  |  |
| ≤20 | 16,785 | 0.9% | 16,767 | 1.0% | 18 | 0.0% | <0.001 | 34 | 0.0% | 16 | 0.0% | 18 | 0.0% | 1.00 |
| 21-40 | 146,712 | 8.2% | 146,210 | 8.9% | 502 | 0.4% |  | 1,009 | 0.4% | 507 | 0.4% | 502 | 0.4% |  |
| 41-50 | 231,312 | 13.0% | 228,725 | 13.9% | 2,587 | 1.9% |  | 5,163 | 1.9% | 2,577 | 1.9% | 2,586 | 1.9% |  |
| 51-60 | 434,995 | 24.4% | 410,636 | 24.9% | 24,359 | 17.9% |  | 48,700 | 17.9% | 24,349 | 17.9% | 24,351 | 17.9% |  |
| 61-70 | 459,192 | 25.7% | 404,445 | 24.5% | 54,747 | 40.3% |  | 109,415 | 40.3% | 54,711 | 40.3% | 54,704 | 40.3% |  |
| 71-80 | 309,898 | 17.4% | 271,617 | 16.5% | 38,281 | 28.1% |  | 76,139 | 28.0% | 38,070 | 28.0% | 38,069 | 28.0% |  |
| ≥81 | 187,107 | 10.5% | 171,585 | 10.4% | 15,522 | 11.4% |  | 30,988 | 11.4% | 15,494 | 11.4% | 15,494 | 11.4% |  |
| Gender |  |  |  |  |  |  |  |  |  |  |  |  |  |  |
| Female | 1,079,468 | 60.4% | 956,403 | 58.0% | 123,065 | 90.5% | <0.001 | 245,537 | 90.5% | 122,762 | 90.4% | 122,775 | 90.5% | 0.93 |
| Male | 706,533 | 39.6% | 693,582 | 42.0% | 12,951 | 9.5% |  | 25,911 | 9.5% | 12,962 | 9.6% | 12,949 | 9.5% |  |
| Region |  |  |  |  |  |  |  |  |  |  |  |  |  |  |
| Midwest | 347,103 | 19.4% | 321,267 | 19.5% | 25,836 | 19.0% | <0.001 | 51,638 | 19.0% | 25,819 | 19.0% | 25,819 | 19.0% | 1.00 |
| Northeast | 498,566 | 27.9% | 463,273 | 28.1% | 35,293 | 25.9% |  | 70,544 | 26.0% | 35,272 | 26.0% | 35,272 | 26.0% |  |
| South | 669,285 | 37.5% | 622,064 | 37.7% | 47,221 | 34.7% |  | 93,980 | 34.6% | 46,990 | 34.6% | 46,990 | 34.6% |  |
| West | 271,047 | 15.2% | 243,381 | 14.8% | 27,666 | 20.3% |  | 55,286 | 20.4% | 27,643 | 20.4% | 27,643 | 20.4% |  |
| Insurance |  |  |  |  |  |  |  |  |  |  |  |  |  |  |
| Commercial | 848,106 | 47.5% | 798,579 | 48.4% | 49,527 | 36.4% | <0.001 | 99,039 | 36.5% | 49,523 | 36.5% | 49,516 | 36.5% | 1.00 |
| Dual | 46,774 | 2.6% | 40,212 | 2.4% | 6,562 | 4.8% |  | 12,645 | 4.7% | 6,319 | 4.7% | 6,326 | 4.7% |  |
| Medicaid | 406,012 | 22.7% | 381,472 | 23.1% | 24,540 | 18.0% |  | 49,025 | 18.1% | 24,516 | 18.1% | 24,509 | 18.1% |  |
| Medicare | 485,109 | 27.2% | 429,722 | 26.0% | 55,387 | 40.7% |  | 110,739 | 40.8% | 55,366 | 40.8% | 55,373 | 40.8% |  |
| PCP Visit 2019 |  |  |  |  |  |  |  |  |  |  |  |  |  |  |
| No | 719,756 | 40.3% | 676,255 | 41.0% | 43,501 | 32.0% | <0.001 | 86,956 | 32.0% | 43,478 | 32.0% | 43,478 | 32.0% | 1.00 |
| Yes | 1,066,245 | 59.7% | 973,730 | 59.0% | 92,515 | 68.0% |  | 184,492 | 68.0% | 92,246 | 68.0% | 92,246 | 68.0% |  |
| Continuous Outcomes |  |  |  |  |  |  |  |  |  |  |  |  |  |  |
|  | mean | SD | mean | SD | mean | SD | p-value | mean | SD | mean | SD | mean | SD | p-value |
| CCI | 0.96 | 1.66 | 0.96 | 1.65 | 0.88 | 1.76 | <0.001 | 0.88 | 1.75 | 0.88 | 1.74 | 0.88 | 1.75 | 0.76 |

BP: bisphosphonate; CCI: Charlson Comorbidity Index; PCP: primary care physician; SD: standard deviation

**Table S9f:** Antihypertensive Non-user Cohort (Region=New York State) by BP Use, Patient Characteristics Pre/Post Match of BP Users/Non-users

|  | Region=NY Antihypertensive Non-Users by BP: Unmatched |  |  |  |  |  |  | Region=NY Antihypertensive Non-users by BP: Matched |  |  |  |  |  |  |
| --- | --- | --- | --- | --- | --- | --- | --- | --- | --- | --- | --- | --- | --- | --- |
|  | All |  | BP Non-user |  | BP User |  | p-value | All |  | BP Non-user |  | BP User |  | p-value |
|  | N | % | N | % | N | % |  | N | % | N | % | N | % |  |
| All Patients | 203,624 | 100.0% | 189,573 | 93.1% | 14,051 | 6.9% |  | 27,966 | 100.0% | 13,983 | 50.0% | 13,983 | 50.0% |  |
| Age |  |  |  |  |  |  |  |  |  |  |  |  |  |  |
| ≤20 | 811 | 0.4% | 810 | 0.4% | 1 | 0.0% | <0.001 | 2 | 0.0% | 1 | 0.0% | 1 | 0.0% | 1.00 |
| 21-40 | 11,465 | 5.6% | 11,451 | 6.0% | 14 | 0.1% |  | 28 | 0.1% | 14 | 0.1% | 14 | 0.1% |  |
| 41-50 | 21,923 | 10.8% | 21,762 | 11.5% | 161 | 1.1% |  | 324 | 1.2% | 163 | 1.2% | 161 | 1.2% |  |
| 51-60 | 48,159 | 23.7% | 46,035 | 24.3% | 2,124 | 15.1% |  | 4,245 | 15.2% | 2,121 | 15.2% | 2,124 | 15.2% |  |
| 61-70 | 54,929 | 27.0% | 49,409 | 26.1% | 5,520 | 39.3% |  | 11,027 | 39.4% | 5,512 | 39.4% | 5,515 | 39.4% |  |
| 71-80 | 44,367 | 21.8% | 39,789 | 21.0% | 4,578 | 32.6% |  | 9,054 | 32.4% | 4,528 | 32.4% | 4,526 | 32.4% |  |
| ≥81 | 21,970 | 10.8% | 20,317 | 10.7% | 1,653 | 11.8% |  | 3,286 | 11.7% | 1,644 | 11.8% | 1,642 | 11.7% |  |
| Gender |  |  |  |  |  |  |  |  |  |  |  |  |  |  |
| Female | 120,465 | 59.2% | 107,632 | 56.8% | 12,833 | 91.3% | <0.001 | 25,530 | 91.3% | 12,764 | 91.3% | 12,766 | 91.3% | 0.97 |
| Male | 83,159 | 40.8% | 81,941 | 43.2% | 1,218 | 8.7% |  | 2,436 | 8.7% | 1,219 | 8.7% | 1,217 | 8.7% |  |
| Insurance |  |  |  |  |  |  |  |  |  |  |  |  |  |  |
| Commercial | 75,459 | 37.1% | 73,115 | 38.6% | 2,344 | 16.7% | <0.001 | 4,683 | 16.7% | 2,342 | 16.7% | 2,341 | 16.7% | 1.00 |
| Dual | 1,493 | 0.7% | 1,211 | 0.6% | 282 | 2.0% |  | 554 | 2.0% | 277 | 2.0% | 277 | 2.0% |  |
| Medicaid | 47,516 | 23.3% | 43,809 | 23.1% | 3,707 | 26.4% |  | 7,295 | 26.1% | 3,648 | 26.1% | 3,647 | 26.1% |  |
| Medicare | 79,156 | 38.9% | 71,438 | 37.7% | 7,718 | 54.9% |  | 15,434 | 55.2% | 7,716 | 55.2% | 7,718 | 55.2% |  |
| PCP Visit 2019 |  |  |  |  |  |  |  |  |  |  |  |  |  |  |
| No | 90,617 | 44.5% | 85,875 | 45.3% | 4,742 | 33.7% | <0.001 | 9,461 | 33.8% | 4,728 | 33.8% | 4,733 | 33.8% | 0.95 |
| Yes | 113,007 | 55.5% | 103,698 | 54.7% | 9,309 | 66.3% |  | 18,505 | 66.2% | 9,255 | 66.2% | 9,250 | 66.2% |  |
| Continuous Outcomes |  |  |  |  |  |  |  |  |  |  |  |  |  |  |
|  | mean | SD | mean | SD | mean | SD | p-value | mean | SD | mean | SD | mean | SD | p-value |
| CCI | 0.95 | 1.60 | 0.96 | 1.60 | 0.81 | 1.60 | <0.001 | 0.81 | 1.59 | 0.81 | 1.58 | 0.81 | 1.59 | 0.92 |

BP: bisphosphonate; CCI: Charlson Comorbidity Index; PCP: primary care physician; SD: standard deviation

**Table S10a:** Antidiabetic Cohort (All Regions), Patient Characteristics Pre/Post Match

|  | All Observations by Antidiabetic Use: Unmatched |  |  |  |  |  |  | All Observations by Antidiabetic Use: Matched |  |  |  |  |  |  |
| --- | --- | --- | --- | --- | --- | --- | --- | --- | --- | --- | --- | --- | --- | --- |
|  | All |  | DIAB Non-users |  | DIAB Users |  | p-value | All |  | DIAB Non-users |  | DIAB Users |  | p-value |
|  | N | % | N | % | N | % |  | N | % | N | % | N | % |  |
| All Patients | 7,906,603 | 100.0% | 7,151,351 | 90.4% | 755,252 | 9.6% |  | 1,509,106 | 100.0% | 754,553 | 50.0% | 754,553 | 50.0% |  |
| Age |  |  |  |  |  |  | <0.001 |  |  |  |  |  |  | 1.00 |
| ≤20 | 1,840,050 | 23.3% | 1,833,838 | 25.6% | 6,212 | 0.8% |  | 12,422 | 0.8% | 6,211 | 0.8% | 6,211 | 0.8% |  |
| 21-40 | 1,446,999 | 18.3% | 1,389,243 | 19.4% | 57,756 | 7.6% |  | 115,448 | 7.7% | 57,723 | 7.6% | 57,725 | 7.7% |  |
| 41-50 | 925,309 | 11.7% | 833,333 | 11.7% | 91,976 | 12.2% |  | 183,810 | 12.2% | 91,905 | 12.2% | 91,905 | 12.2% |  |
| 51-60 | 1,250,190 | 15.8% | 1,058,878 | 14.8% | 191,312 | 25.3% |  | 382,390 | 25.3% | 191,196 | 25.3% | 191,194 | 25.3% |  |
| 61-70 | 1,181,261 | 14.9% | 973,670 | 13.6% | 207,591 | 27.5% |  | 414,869 | 27.5% | 207,435 | 27.5% | 207,434 | 27.5% |  |
| 71-80 | 783,775 | 9.9% | 645,256 | 9.0% | 138,519 | 18.3% |  | 276,619 | 18.3% | 138,310 | 18.3% | 138,309 | 18.3% |  |
| ≥81 | 479,019 | 6.1% | 417,133 | 5.8% | 61,886 | 8.2% |  | 123,548 | 8.2% | 61,773 | 8.2% | 61,775 | 8.2% |  |
| Gender |  |  |  |  |  |  |  |  |  |  |  |  |  |  |
| Female | 4,670,960 | 59.1% | 4,212,086 | 58.9% | 458,874 | 60.8% | <0.001 | 916,914 | 60.8% | 458,455 | 60.8% | 458,459 | 60.8% | 0.99 |
| Male | 3,235,643 | 40.9% | 2,939,265 | 41.1% | 296,378 | 39.2% |  | 592,192 | 39.2% | 296,098 | 39.2% | 296,094 | 39.2% |  |
| Region |  |  |  |  |  |  |  |  |  |  |  |  |  |  |
| Midwest | 1,467,802 | 18.6% | 1,333,631 | 18.6% | 134,171 | 17.8% | <0.001 | 268,044 | 17.8% | 134,022 | 17.8% | 134,022 | 17.8% | 1.00 |
| Northeast | 2,152,560 | 27.2% | 1,935,311 | 27.1% | 217,249 | 28.8% |  | 434,080 | 28.8% | 217,040 | 28.8% | 217,040 | 28.8% |  |
| South | 3,042,604 | 38.5% | 2,752,618 | 38.5% | 289,986 | 38.4% |  | 579,562 | 38.4% | 289,781 | 38.4% | 289,781 | 38.4% |  |
| West | 1,243,637 | 15.7% | 1,129,791 | 15.8% | 113,846 | 15.1% |  | 227,420 | 15.1% | 113,710 | 15.1% | 113,710 | 15.1% |  |
| Insurance |  |  |  |  |  |  |  |  |  |  |  |  |  |  |
| Commercial | 3,938,603 | 49.8% | 3,631,514 | 50.8% | 307,089 | 40.7% | <0.001 | 614,045 | 40.7% | 307,022 | 40.7% | 307,023 | 40.7% | 1.00 |
| Dual | 156,497 | 2.0% | 113,496 | 1.6% | 43,001 | 5.7% |  | 85,209 | 5.6% | 42,603 | 5.6% | 42,606 | 5.6% |  |
| Medicaid | 2,594,500 | 32.8% | 2,387,519 | 33.4% | 206,981 | 27.4% |  | 413,743 | 27.4% | 206,875 | 27.4% | 206,868 | 27.4% |  |
| Medicare | 1,217,003 | 15.4% | 1,018,822 | 14.2% | 198,181 | 26.2% |  | 396,109 | 26.2% | 198,053 | 26.2% | 198,056 | 26.2% |  |
| PCP Visit 2019 |  |  |  |  |  |  |  |  |  |  |  |  |  |  |
| No | 4,283,697 | 54.2% | 4,030,804 | 56.4% | 252,893 | 33.5% | <0.001 | 505,500 | 33.5% | 252,752 | 33.5% | 252,748 | 33.5% | 0.99 |
| Yes | 3,622,906 | 45.8% | 3,120,547 | 43.6% | 502,359 | 66.5% |  | 1,003,606 | 66.5% | 501,801 | 66.5% | 501,805 | 66.5% |  |
| Continuous Outcomes |  |  |  |  |  |  |  |  |  |  |  |  |  |  |
|  | mean | SD | mean | SD | mean | SD | p-value | mean | SD | mean | SD | mean | SD | p-value |
| CCI | 0.62 | 1.38 | 0.55 | 1.30 | 1.25 | 1.84 | <0.001 | 1.24 | 1.82 | 1.24 | 1.82 | 1.24 | 1.82 | 0.99 |

CCI: Charlson Comorbidity Index; DIAB: antidiabetic; PCP: primary care physician; SD: standard deviation

**Table S10b:** Antidiabetic Cohort (Region=New York State), Patient Characteristics Pre/Post Match

|  | Region=NY by Antidiabetic Use: Unmatched |  |  |  |  |  |  | Region=NY by Antidiabetic Use: Matched |  |  |  |  |  |  |
| --- | --- | --- | --- | --- | --- | --- | --- | --- | --- | --- | --- | --- | --- | --- |
|  | All |  | DIAB Non-users |  | DIAB Users |  | p-value | All |  | DIAB Non-users |  | DIAB Users |  | p-value |
|  | N | % | N | % | N | % |  | N | % | N | % | N | % |  |
| All Patients | 968,296 | 100.0% | 863,179 | 89.1% | 105,117 | 10.9% |  | 209,382 | 100.0% | 104,691 | 50.0% | 104,691 | 50.0% |  |
| Age |  |  |  |  |  |  |  |  |  |  |  |  |  |  |
| ≤20 | 133,178 | 13.8% | 132,723 | 15.4% | 455 | 0.4% | <0.001 | 910 | 0.4% | 455 | 0.4% | 455 | 0.4% | 1.00 |
| 21-40 | 192,959 | 19.9% | 186,785 | 21.6% | 6,174 | 5.9% |  | 12,328 | 5.9% | 6,164 | 5.9% | 6,164 | 5.9% |  |
| 41-50 | 127,794 | 13.2% | 117,342 | 13.6% | 10,452 | 9.9% |  | 20,880 | 10.0% | 10,440 | 10.0% | 10,440 | 10.0% |  |
| 51-60 | 172,444 | 17.8% | 148,040 | 17.2% | 24,404 | 23.2% |  | 48,735 | 23.3% | 24,369 | 23.3% | 24,366 | 23.3% |  |
| 61-70 | 159,912 | 16.5% | 130,968 | 15.2% | 28,944 | 27.5% |  | 57,638 | 27.5% | 28,819 | 27.5% | 28,819 | 27.5% |  |
| 71-80 | 120,117 | 12.4% | 95,621 | 11.1% | 24,496 | 23.3% |  | 48,625 | 23.2% | 24,311 | 23.2% | 24,314 | 23.2% |  |
| ≥81 | 61,892 | 6.4% | 51,700 | 6.0% | 10,192 | 9.7% |  | 20,266 | 9.7% | 10,133 | 9.7% | 10,133 | 9.7% |  |
| Gender |  |  |  |  |  |  |  |  |  |  |  |  |  |  |
| Female | 573,610 | 59.2% | 512,889 | 59.4% | 60,721 | 57.8% | <0.001 | 120,937 | 57.8% | 60,467 | 57.8% | 60,470 | 57.8% | 0.99 |
| Male | 394,686 | 40.8% | 350,290 | 40.6% | 44,396 | 42.2% |  | 88,445 | 42.2% | 44,224 | 42.2% | 44,221 | 42.2% |  |
| Insurance |  |  |  |  |  |  |  |  |  |  |  |  |  |  |
| Commercial | 500,918 | 51.7% | 468,804 | 54.3% | 32,114 | 30.6% | <0.001 | 64,200 | 30.7% | 32,100 | 30.7% | 32,100 | 30.7% | 1.00 |
| Dual | 6,814 | 0.7% | 4,408 | 0.5% | 2,406 | 2.3% |  | 4,389 | 2.1% | 2,196 | 2.1% | 2,193 | 2.1% |  |
| Medicaid | 252,366 | 26.1% | 224,334 | 26.0% | 28,032 | 26.7% |  | 55,853 | 26.7% | 27,925 | 26.7% | 27,928 | 26.7% |  |
| Medicare | 208,198 | 21.5% | 165,633 | 19.2% | 42,565 | 40.5% |  | 84,940 | 40.6% | 42,470 | 40.6% | 42,470 | 40.6% |  |
| PCP Visit 2019 |  |  |  |  |  |  |  |  |  |  |  |  |  |  |
| No | 521,282 | 53.8% | 484,071 | 56.1% | 37,211 | 35.4% | <0.001 | 74,215 | 35.4% | 37,106 | 35.4% | 37,109 | 35.4% | 0.99 |
| Yes | 447,014 | 46.2% | 379,108 | 43.9% | 67,906 | 64.6% |  | 135,167 | 64.6% | 67,585 | 64.6% | 67,582 | 64.6% |  |
| Continuous Outcomes |  |  |  |  |  |  |  |  |  |  |  |  |  |  |
|  | mean | SD | mean | SD | mean | SD | p-value | mean | SD | mean | SD | mean | SD | p-value |
| CCI | 0.65 | 1.39 | 0.56 | 1.30 | 1.34 | 1.84 | <0.001 | 1.32 | 1.79 | 1.32 | 1.79 | 1.32 | 1.79 | 0.98 |

CCI: Charlson Comorbidity Index; DIAB: antidiabetic; PCP: primary care physician; SD: standard deviation

**Table S10c:** Antidiabetic User Cohort (All Regions) by BP Use, Patient Characteristics Pre/Post Match of BP Users/Non-users

|  | All Antidiabetic Users by BP: Unmatched |  |  |  |  |  |  | All Antidiabetic Users by BP: Matched |  |  |  |  |  |  |
| --- | --- | --- | --- | --- | --- | --- | --- | --- | --- | --- | --- | --- | --- | --- |
|  | All |  | BP Non-user |  | BP User |  | p-value | All |  | BP Non-user |  | BP User |  | p-value |
|  | N | % | N | % | N | % |  | N | % | N | % | N | % |  |
| All Patients | 754,553 | 100.0% | 674,024 | 89.3% | 80,529 | 10.7% |  | 159,000 | 100.0% | 79,500 | 50.0% | 79,500 | 50.0% |  |
| Age |  |  |  |  |  |  |  |  |  |  |  |  |  |  |
| ≤20 | 6,211 | 0.8% | 6,169 | 0.9% | 42 | 0.1% | <0.001 | 83 | 0.1% | 41 | 0.1% | 42 | 0.1% | 1.00 |
| 21-40 | 57,725 | 7.7% | 57,535 | 8.5% | 190 | 0.2% |  | 380 | 0.2% | 190 | 0.2% | 190 | 0.2% |  |
| 41-50 | 91,905 | 12.2% | 90,952 | 13.5% | 953 | 1.2% |  | 1,905 | 1.2% | 952 | 1.2% | 953 | 1.2% |  |
| 51-60 | 191,194 | 25.3% | 182,922 | 27.1% | 8,272 | 10.3% |  | 16,536 | 10.4% | 8,268 | 10.4% | 8,268 | 10.4% |  |
| 61-70 | 207,434 | 27.5% | 180,895 | 26.8% | 26,539 | 33.0% |  | 53,028 | 33.4% | 26,512 | 33.3% | 26,516 | 33.4% |  |
| 71-80 | 138,309 | 18.3% | 107,467 | 15.9% | 30,842 | 38.3% |  | 60,240 | 37.9% | 30,121 | 37.9% | 30,119 | 37.9% |  |
| ≥81 | 61,775 | 8.2% | 48,084 | 7.1% | 13,691 | 17.0% |  | 26,828 | 16.9% | 13,416 | 16.9% | 13,412 | 16.9% |  |
| Gender |  |  |  |  |  |  |  |  |  |  |  |  |  |  |
| Female | 458,459 | 60.8% | 386,400 | 57.3% | 72,059 | 89.5% | <0.001 | 142,068 | 89.4% | 71,027 | 89.3% | 71,041 | 89.4% | 0.91 |
| Male | 296,094 | 39.2% | 287,624 | 42.7% | 8,470 | 10.5% |  | 16,932 | 10.6% | 8,473 | 10.7% | 8,459 | 10.6% |  |
| Region |  |  |  |  |  |  |  |  |  |  |  |  |  |  |
| Midwest | 134,022 | 17.8% | 123,909 | 18.4% | 10,113 | 12.6% | <0.001 | 20,168 | 12.7% | 10,084 | 12.7% | 10,084 | 12.7% | 1.00 |
| Northeast | 217,040 | 28.8% | 196,723 | 29.2% | 20,317 | 25.2% |  | 40,446 | 25.4% | 20,223 | 25.4% | 20,223 | 25.4% |  |
| South | 289,781 | 38.4% | 257,599 | 38.2% | 32,182 | 40.0% |  | 63,740 | 40.1% | 31,870 | 40.1% | 31,870 | 40.1% |  |
| West | 113,710 | 15.1% | 95,793 | 14.2% | 17,917 | 22.2% |  | 34,646 | 21.8% | 17,323 | 21.8% | 17,323 | 21.8% |  |
| Insurance |  |  |  |  |  |  |  |  |  |  |  |  |  |  |
| Commercial | 307,023 | 40.7% | 290,957 | 43.2% | 16,066 | 20.0% | <0.001 | 32,086 | 20.2% | 16,043 | 20.2% | 16,043 | 20.2% | 1.00 |
| Dual | 42,606 | 5.6% | 32,797 | 4.9% | 9,809 | 12.2% |  | 18,653 | 11.7% | 9,321 | 11.7% | 9,332 | 11.7% |  |
| Medicaid | 206,868 | 27.4% | 188,638 | 28.0% | 18,230 | 22.6% |  | 35,513 | 22.3% | 17,759 | 22.3% | 17,754 | 22.3% |  |
| Medicare | 198,056 | 26.2% | 161,632 | 24.0% | 36,424 | 45.2% |  | 72,748 | 45.8% | 36,377 | 45.8% | 36,371 | 45.7% |  |
| PCP Visit 2019 |  |  |  |  |  |  |  |  |  |  |  |  |  |  |
| No | 252,748 | 33.5% | 228,203 | 33.9% | 24,545 | 30.5% | <0.001 | 48,374 | 30.4% | 24,184 | 30.4% | 24,190 | 30.4% | 0.97 |
| Yes | 501,805 | 66.5% | 445,821 | 66.1% | 55,984 | 69.5% |  | 110,626 | 69.6% | 55,316 | 69.6% | 55,310 | 69.6% |  |
| Continuous Outcomes |  |  |  |  |  |  |  |  |  |  |  |  |  |  |
|  | mean | SD | mean | SD | mean | SD | p-value | mean | SD | mean | SD | mean | SD | p-value |
| CCI | 1.24 | 1.82 | 1.23 | 1.81 | 1.32 | 1.90 | <0.001 | 1.31 | 1.88 | 1.31 | 1.87 | 1.32 | 1.88 | 0.75 |

BP: bisphosphonate; CCI: Charlson Comorbidity Index; PCP: primary care physician; SD: standard deviation

**Table S10d:** Antidiabetic User Cohort (Region=New York State) by BP Use, Patient Characteristics Pre/Post Match of BP Users/Non-users

|  | Region=NY Antidiabetic Users by BP: Unmatched |  |  |  |  |  |  | Region=NY Antidiabetic Users by BP: Matched |  |  |  |  |  |  |
| --- | --- | --- | --- | --- | --- | --- | --- | --- | --- | --- | --- | --- | --- | --- |
|  | All |  | BP Non-user |  | BP User |  | p-value | All |  | BP Non-user |  | BP User |  | p-value |
|  | N | % | N | % | N | % |  | N | % | N | % | N | % |  |
| All Patients | 104,691 | 100.0% | 95,162 | 90.9% | 9,529 | 9.1% |  | 18,912 | 100.0% | 9,456 | 50.0% | 9,456 | 50.0% |  |
| Age |  |  |  |  |  |  |  |  |  |  |  |  |  |  |
| ≤20 | 455 | 0.4% | 454 | 0.5% | 1 | 0.0% | <0.001 | 2 | 0.0% | 1 | 0.0% | 1 | 0.0% | 1.00 |
| 21-40 | 6,164 | 5.9% | 6,152 | 6.5% | 12 | 0.1% |  | 25 | 0.1% | 13 | 0.1% | 12 | 0.1% |  |
| 41-50 | 10,440 | 10.0% | 10,363 | 10.9% | 77 | 0.8% |  | 151 | 0.8% | 75 | 0.8% | 76 | 0.8% |  |
| 51-60 | 24,366 | 23.3% | 23,532 | 24.7% | 834 | 8.8% |  | 1,665 | 8.8% | 831 | 8.8% | 834 | 8.8% |  |
| 61-70 | 28,819 | 27.5% | 25,939 | 27.3% | 2,880 | 30.2% |  | 5,741 | 30.4% | 2,870 | 30.4% | 2,871 | 30.4% |  |
| 71-80 | 24,314 | 23.2% | 20,338 | 21.4% | 3,976 | 41.7% |  | 7,880 | 41.7% | 3,941 | 41.7% | 3,939 | 41.7% |  |
| ≥81 | 10,133 | 9.7% | 8,384 | 8.8% | 1,749 | 18.4% |  | 3,448 | 18.2% | 1,725 | 18.2% | 1,723 | 18.2% |  |
| Gender |  |  |  |  |  |  |  |  |  |  |  |  |  |  |
| Female | 60,470 | 57.8% | 51,884 | 54.5% | 8,586 | 90.1% | <0.001 | 17,022 | 90.0% | 8,509 | 90.0% | 8,513 | 90.0% | 0.92 |
| Male | 44,221 | 42.2% | 43,278 | 45.5% | 943 | 9.9% |  | 1,890 | 10.0% | 947 | 10.0% | 943 | 10.0% |  |
| Insurance |  |  |  |  |  |  |  |  |  |  |  |  |  |  |
| Commercial | 32,100 | 30.7% | 31,172 | 32.8% | 928 | 9.7% | <0.001 | 1,849 | 9.8% | 924 | 9.8% | 925 | 9.8% | 1.00 |
| Dual | 2,193 | 2.1% | 1,693 | 1.8% | 500 | 5.2% |  | 978 | 5.2% | 490 | 5.2% | 488 | 5.2% |  |
| Medicaid | 27,928 | 26.7% | 25,978 | 27.3% | 1,950 | 20.5% |  | 3,793 | 20.1% | 1,897 | 20.1% | 1,896 | 20.1% |  |
| Medicare | 42,470 | 40.6% | 36,319 | 38.2% | 6,151 | 64.6% |  | 12,292 | 65.0% | 6,145 | 65.0% | 6,147 | 65.0% |  |
| PCP Visit 2019 |  |  |  |  |  |  |  |  |  |  |  |  |  |  |
| No | 37,109 | 35.4% | 33,894 | 35.6% | 3,215 | 33.7% | <.001 | 6,363 | 33.6% | 3,182 | 33.7% | 3,181 | 33.6% | 0.99 |
| Yes | 67,582 | 64.6% | 61,268 | 64.4% | 6,314 | 66.3% |  | 12,549 | 66.4% | 6,274 | 66.3% | 6,275 | 66.4% |  |
| Continuous Outcomes |  |  |  |  |  |  |  |  |  |  |  |  |  |  |
|  | mean | SD | mean | SD | mean | SD | p-value | mean | SD | mean | SD | mean | SD | p-value |
| CCI | 1.32 | 1.79 | 1.31 | 1.79 | 1.46 | 1.87 | <0.001 | 1.44 | 1.83 | 1.44 | 1.82 | 1.45 | 1.84 | 0.75 |

BP: bisphosphonate; CCI: Charlson Comorbidity Index; PCP: primary care physician; SD: standard deviation

**Table S10e:** Antidiabetic Non-user Cohort (All Regions) by BP Use, Patient Characteristics Pre/Post Match of BP Users/Non-users

|  | All Antidiabetic Non-users by BP: Unmatched |  |  |  |  |  |  | All Antidiabetic Non-users by BP: Matched |  |  |  |  |  |  |
| --- | --- | --- | --- | --- | --- | --- | --- | --- | --- | --- | --- | --- | --- | --- |
|  | All |  | BP Non-user |  | BP User |  | p-value | All |  | BP Non-user |  | BP User |  | p-value |
|  | N | % | N | % | N | % |  | N | % | N | % | N | % |  |
| All Patients | 754,553 | 100.0% | 681,380 | 90.3% | 73,173 | 9.7% |  | 145,028 | 100.0% | 72,514 | 50.0% | 72,514 | 50.0% |  |
| Age |  |  |  |  |  |  |  |  |  |  |  |  |  |  |
| ≤20 | 6,211 | 0.8% | 6,199 | 0.9% | 12 | 0.0% | <0.001 | 24 | 0.0% | 12 | 0.0% | 12 | 0.0% | 1.00 |
| 21-40 | 57,723 | 7.6% | 57,497 | 8.4% | 226 | 0.3% |  | 455 | 0.3% | 229 | 0.3% | 226 | 0.3% |  |
| 41-50 | 91,905 | 12.2% | 90,693 | 13.3% | 1,212 | 1.7% |  | 2,421 | 1.7% | 1,209 | 1.7% | 1,212 | 1.7% |  |
| 51-60 | 191,196 | 25.3% | 180,332 | 26.5% | 10,864 | 14.8% |  | 21,721 | 15.0% | 10,860 | 15.0% | 10,861 | 15.0% |  |
| 61-70 | 207,435 | 27.5% | 180,825 | 26.5% | 26,610 | 36.4% |  | 53,115 | 36.6% | 26,558 | 36.6% | 26,557 | 36.6% |  |
| 71-80 | 138,310 | 18.3% | 114,018 | 16.7% | 24,292 | 33.2% |  | 47,723 | 32.9% | 23,861 | 32.9% | 23,862 | 32.9% |  |
| ≥81 | 61,773 | 8.2% | 51,816 | 7.6% | 9,957 | 13.6% |  | 19,569 | 13.5% | 9,785 | 13.5% | 9,784 | 13.5% |  |
| Gender |  |  |  |  |  |  |  |  |  |  |  |  |  |  |
| Female | 458,455 | 60.8% | 393,376 | 57.7% | 65,079 | 88.9% | <0.001 | 128,836 | 88.8% | 64,411 | 88.8% | 64,425 | 88.8% | 0.91 |
| Male | 296,098 | 39.2% | 288,004 | 42.3% | 8,094 | 11.1% |  | 16,192 | 11.2% | 8,103 | 11.2% | 8,089 | 11.2% |  |
| Region |  |  |  |  |  |  |  |  |  |  |  |  |  |  |
| Midwest | 134,022 | 17.8% | 123,283 | 18.1% | 10,739 | 14.7% | <0.001 | 21,390 | 14.7% | 10,695 | 14.7% | 10,695 | 14.7% | 1.00 |
| Northeast | 217,040 | 28.8% | 197,710 | 29.0% | 19,330 | 26.4% |  | 38,510 | 26.6% | 19,255 | 26.6% | 19,255 | 26.6% |  |
| South | 289,781 | 38.4% | 261,382 | 38.4% | 28,399 | 38.8% |  | 55,812 | 38.5% | 27,906 | 38.5% | 27,906 | 38.5% |  |
| West | 113,710 | 15.1% | 99,005 | 14.5% | 14,705 | 20.1% |  | 29,316 | 20.2% | 14,658 | 20.2% | 14,658 | 20.2% |  |
| Insurance |  |  |  |  |  |  |  |  |  |  |  |  |  |  |
| Commercial | 307,022 | 40.7% | 289,018 | 42.4% | 18,004 | 24.6% | <0.001 | 35,983 | 24.8% | 17,988 | 24.8% | 17,995 | 24.8% | 1.00 |
| Dual | 42,603 | 5.6% | 33,444 | 4.9% | 9,159 | 12.5% |  | 17,221 | 11.9% | 8,611 | 11.9% | 8,610 | 11.9% |  |
| Medicaid | 206,875 | 27.4% | 190,166 | 27.9% | 16,709 | 22.8% |  | 33,264 | 22.9% | 16,636 | 22.9% | 16,628 | 22.9% |  |
| Medicare | 198,053 | 26.2% | 168,752 | 24.8% | 29,301 | 40.0% |  | 58,560 | 40.4% | 29,279 | 40.4% | 29,281 | 40.4% |  |
| PCP Visit 2019 |  |  |  |  |  |  |  |  |  |  |  |  |  |  |
| No | 252,752 | 33.5% | 233,775 | 34.3% | 18,977 | 25.9% | <0.001 | 37,812 | 26.1% | 18,903 | 26.1% | 18,909 | 26.1% | 0.97 |
| Yes | 501,801 | 66.5% | 447,605 | 65.7% | 54,196 | 74.1% |  | 107,216 | 73.9% | 53,611 | 73.9% | 53,605 | 73.9% |  |
| Continuous Outcomes |  |  |  |  |  |  |  |  |  |  |  |  |  |  |
|  | mean | SD | mean | SD | mean | SD | p-value | mean | SD | mean | SD | mean | SD | p-value |
| CCI | 1.24 | 1.82 | 1.24 | 1.81 | 1.24 | 1.89 | 0.92 | 1.24 | 1.87 | 1.24 | 1.87 | 1.25 | 1.88 | 0.63 |

BP: bisphosphonate; CCI: Charlson Comorbidity Index; PCP: primary care physician; SD: standard deviation

**Table S10f:** Antidiabetic Non-user Cohort (Region=New York State) by BP Use, Patient Characteristics Pre/Post Match of BP Users/Non-users

|  | Region=NY Antidiabetic Non-users by BP: Unmatched |  |  |  |  |  |  | Region=NY Antidiabetic Non-users by BP: Matched |  |  |  |  |  |  |
| --- | --- | --- | --- | --- | --- | --- | --- | --- | --- | --- | --- | --- | --- | --- |
|  | All |  | BP Non-user |  | BP User |  | p-value | All |  | BP Non-user |  | BP User |  | p-value |
|  | N | % | N | % | N | % |  | N | % | N | % | N | % |  |
| All Patients | 104,691 | 100.0% | 95,416 | 91.1% | 9,275 | 8.9% |  | 18,288 | 100.0% | 9,144 | 50.0% | 9,144 | 50.0% |  |
| Age |  |  |  |  |  |  | <0.001 |  |  |  |  |  |  | 1.00 |
| ≤20 | 455 | 0.4% | 455 | 0.5% | 0 | 0.0% |  | 0 | 0.0% | 0 | 0.0% | 0 | 0.0% |  |
| 21-40 | 6,164 | 5.9% | 6,146 | 6.4% | 18 | 0.2% |  | 36 | 0.2% | 18 | 0.2% | 18 | 0.2% |  |
| 41-50 | 10,440 | 10.0% | 10,367 | 10.9% | 73 | 0.8% |  | 147 | 0.8% | 74 | 0.8% | 73 | 0.8% |  |
| 51-60 | 24,369 | 23.3% | 23,304 | 24.4% | 1,065 | 11.5% |  | 2,128 | 11.6% | 1,064 | 11.6% | 1,064 | 11.6% |  |
| 61-70 | 28,819 | 27.5% | 25,720 | 27.0% | 3,099 | 33.4% |  | 6,190 | 33.8% | 3,097 | 33.9% | 3,093 | 33.8% |  |
| 71-80 | 24,311 | 23.2% | 20,826 | 21.8% | 3,485 | 37.6% |  | 6,839 | 37.4% | 3,419 | 37.4% | 3,420 | 37.4% |  |
| ≥81 | 10,133 | 9.7% | 8,598 | 9.0% | 1,535 | 16.5% |  | 2,948 | 16.1% | 1,472 | 16.1% | 1,476 | 16.1% |  |
| Gender |  |  |  |  |  |  | <0.001 |  |  |  |  |  |  | 0.98 |
| Female | 60,467 | 57.8% | 52,194 | 54.7% | 8,273 | 89.2% |  | 16,291 | 89.1% | 8,146 | 89.1% | 8,145 | 89.1% |  |
| Male | 44,224 | 42.2% | 43,222 | 45.3% | 1,002 | 10.8% |  | 1,997 | 10.9% | 998 | 10.9% | 999 | 10.9% |  |
| Insurance |  |  |  |  |  |  | <0.001 |  |  |  |  |  |  | 1.00 |
| Commercial | 32,100 | 30.7% | 31,095 | 32.6% | 1,005 | 10.8% |  | 2,002 | 10.9% | 1,000 | 10.9% | 1,002 | 11.0% |  |
| Dual | 2,196 | 2.1% | 1,675 | 1.8% | 521 | 5.6% |  | 1,006 | 5.5% | 502 | 5.5% | 504 | 5.5% |  |
| Medicaid | 27,925 | 26.7% | 25,530 | 26.8% | 2,395 | 25.8% |  | 4,575 | 25.0% | 2,289 | 25.0% | 2,286 | 25.0% |  |
| Medicare | 42,470 | 40.6% | 37,116 | 38.9% | 5,354 | 57.7% |  | 10,705 | 58.5% | 5,353 | 58.5% | 5,352 | 58.5% |  |
| PCP Visit 2019 |  |  |  |  |  |  | <0.001 |  |  |  |  |  |  | 0.96 |
| No | 37,106 | 35.4% | 34,553 | 36.2% | 2,553 | 27.5% |  | 5,039 | 27.6% | 2,518 | 27.5% | 2,521 | 27.6% |  |
| Yes | 67,585 | 64.6% | 60,863 | 63.8% | 6,722 | 72.5% |  | 13,249 | 72.4% | 6,626 | 72.5% | 6,623 | 72.4% |  |
| Continuous Outcomes |  |  |  |  |  |  |  |  |  |  |  |  |  |  |
|  | mean | SD | mean | SD | mean | SD | p-value | mean | SD | mean | SD | mean | SD | p-value |
| CCI | 1.32 | 1.79 | 1.32 | 1.79 | 1.37 | 1.81 | 0.007 | 1.37 | 1.78 | 1.36 | 1.78 | 1.37 | 1.79 | 0.92 |

BP: bisphosphonate; CCI: Charlson Comorbidity Index; PCP: primary care physician; SD: standard deviation

**Table S11a:**
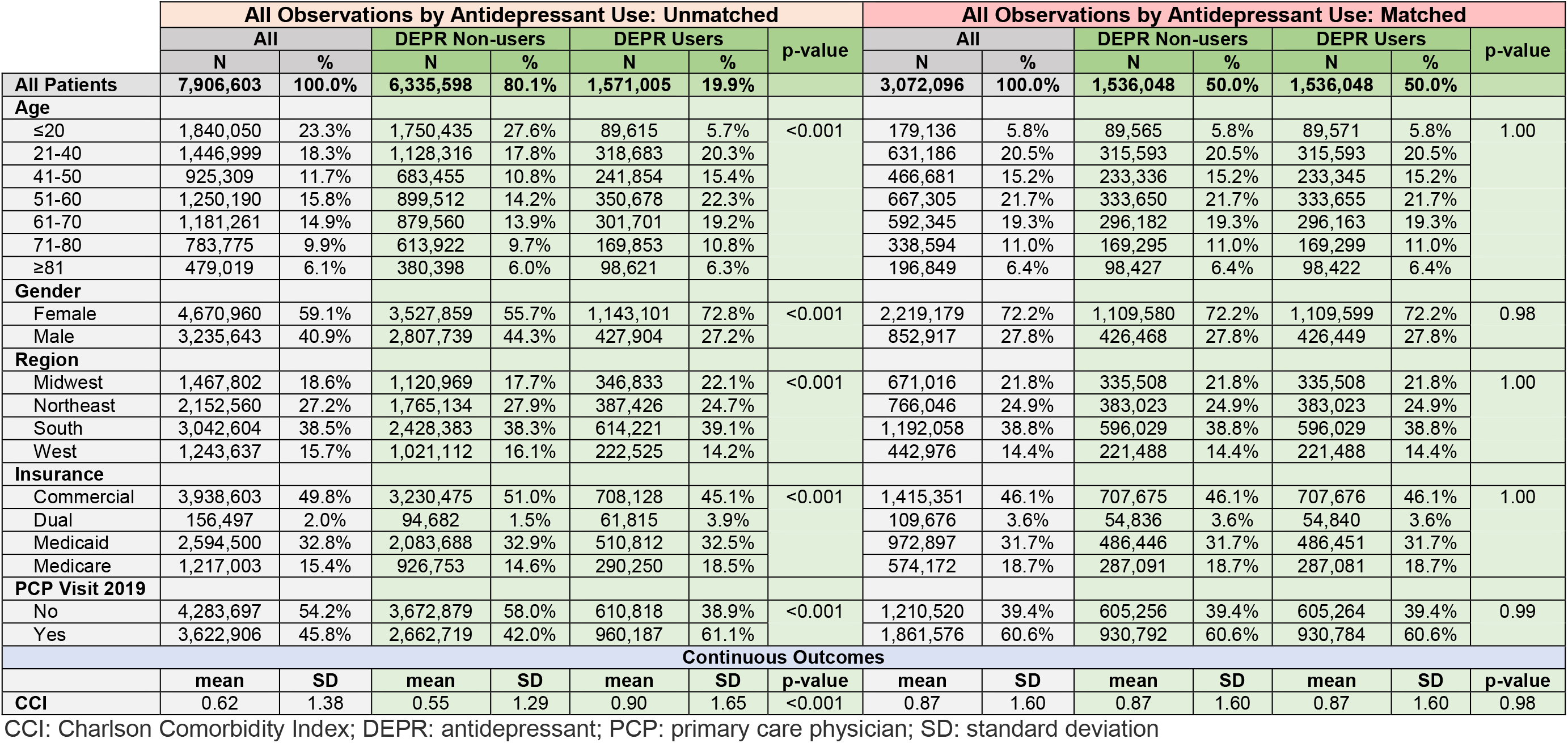
Antidepressant Cohort (All Regions), Patient Characteristics Pre/Post Match

|  | All Observations by Antidepressant Use: Unmatched |  |  |  |  |  |  | All Observations by Antidepressant Use: Matched |  |  |  |  |  |  |
| --- | --- | --- | --- | --- | --- | --- | --- | --- | --- | --- | --- | --- | --- | --- |
|  | All |  | DEPR Non-users |  | DEPR Users |  | p-value | All |  | DEPR Non-users |  | DEPR Users |  | p-value |
|  | N | % | N | % | N | % |  | N | % | N | % | N | % |  |
| All Patients | 7,906,603 | 100.0% | 6,335,598 | 80.1% | 1,571,005 | 19.9% |  | 3,072,096 | 100.0% | 1,536,048 | 50.0% | 1,536,048 | 50.0% |  |
| Age |  |  |  |  |  |  |  |  |  |  |  |  |  |  |
| ≤20 | 1,840,050 | 23.3% | 1,750,435 | 27.6% | 89,615 | 5.7% | <0.001 | 179,136 | 5.8% | 89,565 | 5.8% | 89,571 | 5.8% | 1.00 |
| 21-40 | 1,446,999 | 18.3% | 1,128,316 | 17.8% | 318,683 | 20.3% |  | 631,186 | 20.5% | 315,593 | 20.5% | 315,593 | 20.5% |  |
| 41-50 | 925,309 | 11.7% | 683,455 | 10.8% | 241,854 | 15.4% |  | 466,681 | 15.2% | 233,336 | 15.2% | 233,345 | 15.2% |  |
| 51-60 | 1,250,190 | 15.8% | 899,512 | 14.2% | 350,678 | 22.3% |  | 667,305 | 21.7% | 333,650 | 21.7% | 333,655 | 21.7% |  |
| 61-70 | 1,181,261 | 14.9% | 879,560 | 13.9% | 301,701 | 19.2% |  | 592,345 | 19.3% | 296,182 | 19.3% | 296,163 | 19.3% |  |
| 71-80 | 783,775 | 9.9% | 613,922 | 9.7% | 169,853 | 10.8% |  | 338,594 | 11.0% | 169,295 | 11.0% | 169,299 | 11.0% |  |
| ≥81 | 479,019 | 6.1% | 380,398 | 6.0% | 98,621 | 6.3% |  | 196,849 | 6.4% | 98,427 | 6.4% | 98,422 | 6.4% |  |
| Gender |  |  |  |  |  |  |  |  |  |  |  |  |  |  |
| Female | 4,670,960 | 59.1% | 3,527,859 | 55.7% | 1,143,101 | 72.8% | <0.001 | 2,219,179 | 72.2% | 1,109,580 | 72.2% | 1,109,599 | 72.2% | 0.98 |
| Male | 3,235,643 | 40.9% | 2,807,739 | 44.3% | 427,904 | 27.2% |  | 852,917 | 27.8% | 426,468 | 27.8% | 426,449 | 27.8% |  |
| Region |  |  |  |  |  |  |  |  |  |  |  |  |  |  |
| Midwest | 1,467,802 | 18.6% | 1,120,969 | 17.7% | 346,833 | 22.1% | <0.001 | 671,016 | 21.8% | 335,508 | 21.8% | 335,508 | 21.8% | 1.00 |
| Northeast | 2,152,560 | 27.2% | 1,765,134 | 27.9% | 387,426 | 24.7% |  | 766,046 | 24.9% | 383,023 | 24.9% | 383,023 | 24.9% |  |
| South | 3,042,604 | 38.5% | 2,428,383 | 38.3% | 614,221 | 39.1% |  | 1,192,058 | 38.8% | 596,029 | 38.8% | 596,029 | 38.8% |  |
| West | 1,243,637 | 15.7% | 1,021,112 | 16.1% | 222,525 | 14.2% |  | 442,976 | 14.4% | 221,488 | 14.4% | 221,488 | 14.4% |  |
| Insurance |  |  |  |  |  |  |  |  |  |  |  |  |  |  |
| Commercial | 3,938,603 | 49.8% | 3,230,475 | 51.0% | 708,128 | 45.1% | <0.001 | 1,415,351 | 46.1% | 707,675 | 46.1% | 707,676 | 46.1% | 1.00 |
| Dual | 156,497 | 2.0% | 94,682 | 1.5% | 61,815 | 3.9% |  | 109,676 | 3.6% | 54,836 | 3.6% | 54,840 | 3.6% |  |
| Medicaid | 2,594,500 | 32.8% | 2,083,688 | 32.9% | 510,812 | 32.5% |  | 972,897 | 31.7% | 486,446 | 31.7% | 486,451 | 31.7% |  |
| Medicare | 1,217,003 | 15.4% | 926,753 | 14.6% | 290,250 | 18.5% |  | 574,172 | 18.7% | 287,091 | 18.7% | 287,081 | 18.7% |  |
| PCP Visit 2019 |  |  |  |  |  |  |  |  |  |  |  |  |  |  |
| No | 4,283,697 | 54.2% | 3,672,879 | 58.0% | 610,818 | 38.9% | <0.001 | 1,210,520 | 39.4% | 605,256 | 39.4% | 605,264 | 39.4% | 0.99 |
| Yes | 3,622,906 | 45.8% | 2,662,719 | 42.0% | 960,187 | 61.1% |  | 1,861,576 | 60.6% | 930,792 | 60.6% | 930,784 | 60.6% |  |
| Continuous Outcomes |  |  |  |  |  |  |  |  |  |  |  |  |  |  |
|  | mean | SD | mean | SD | mean | SD | p-value | mean | SD | mean | SD | mean | SD | p-value |
| CCI | 0.62 | 1.38 | 0.55 | 1.29 | 0.90 | 1.65 | <0.001 | 0.87 | 1.60 | 0.87 | 1.60 | 0.87 | 1.60 | 0.98 |

CCI: Charlson Comorbidity Index; DEPR: antidepressant; PCP: primary care physician; SD: standard deviation

**Table S11b:** Antidepressant Cohort (Region=New York State), Patient Characteristics Pre/Post Match

|  | Region=NY by Antidepressant Use: Unmatched |  |  |  |  |  |  | Region=NY by Antidepressant Use: Matched |  |  |  |  |  |  |
| --- | --- | --- | --- | --- | --- | --- | --- | --- | --- | --- | --- | --- | --- | --- |
|  | All |  | DEPR Non-users |  | DEPR Users |  | p-value | All |  | DEPR Non-users |  | DEPR Users |  | p-value |
|  | N | % | N | % | N | % |  | N | % | N | % | N | % |  |
| All Patients | 968,296 | 100.0% | 832,215 | 85.9% | 136,081 | 14.1% |  | 271,032 | 100.0% | 135,516 | 50.0% | 135,516 | 50.0% |  |
| Age |  |  |  |  |  |  |  |  |  |  |  |  |  |  |
| ≤20 | 133,178 | 13.8% | 128,810 | 15.5% | 4,368 | 3.2% | <0.001 | 8,728 | 3.2% | 4,365 | 3.2% | 4,363 | 3.2% | 1.00 |
| 21-40 | 192,959 | 19.9% | 170,076 | 20.4% | 22,883 | 16.8% |  | 45,666 | 16.8% | 22,832 | 16.8% | 22,834 | 16.8% |  |
| 41-50 | 127,794 | 13.2% | 109,184 | 13.1% | 18,610 | 13.7% |  | 36,965 | 13.6% | 18,483 | 13.6% | 18,482 | 13.6% |  |
| 51-60 | 172,444 | 17.8% | 142,702 | 17.1% | 29,742 | 21.9% |  | 58,966 | 21.8% | 29,481 | 21.8% | 29,485 | 21.8% |  |
| 61-70 | 159,912 | 16.5% | 132,317 | 15.9% | 27,595 | 20.3% |  | 55,083 | 20.3% | 27,543 | 20.3% | 27,540 | 20.3% |  |
| 71-80 | 120,117 | 12.4% | 99,040 | 11.9% | 21,077 | 15.5% |  | 42,076 | 15.5% | 21,038 | 15.5% | 21,038 | 15.5% |  |
| ≥81 | 61,892 | 6.4% | 50,086 | 6.0% | 11,806 | 8.7% |  | 23,548 | 8.7% | 11,774 | 8.7% | 11,774 | 8.7% |  |
| Gender |  |  |  |  |  |  |  |  |  |  |  |  |  |  |
| Female | 573,610 | 59.2% | 476,684 | 57.3% | 96,926 | 71.2% | <0.001 | 192,930 | 71.2% | 96,468 | 71.2% | 96,462 | 71.2% | 0.98 |
| Male | 394,686 | 40.8% | 355,531 | 42.7% | 39,155 | 28.8% |  | 78,102 | 28.8% | 39,048 | 28.8% | 39,054 | 28.8% |  |
| Insurance |  |  |  |  |  |  |  |  |  |  |  |  |  |  |
| Commercial | 500,918 | 51.7% | 449,071 | 54.0% | 51,847 | 38.1% | <0.001 | 103,658 | 38.2% | 51,829 | 38.2% | 51,829 | 38.2% | 1.00 |
| Dual | 6,814 | 0.7% | 5,072 | 0.6% | 1,742 | 1.3% |  | 3,191 | 1.2% | 1,591 | 1.2% | 1,600 | 1.2% |  |
| Medicaid | 252,366 | 26.1% | 213,705 | 25.7% | 38,661 | 28.4% |  | 77,136 | 28.5% | 38,569 | 28.5% | 38,567 | 28.5% |  |
| Medicare | 208,198 | 21.5% | 164,367 | 19.8% | 43,831 | 32.2% |  | 87,047 | 32.1% | 43,527 | 32.1% | 43,520 | 32.1% |  |
| PCP Visit 2019 |  |  |  |  |  |  |  |  |  |  |  |  |  |  |
| No | 521,282 | 53.8% | 467,739 | 56.2% | 53,543 | 39.3% | <0.001 | 106,797 | 39.4% | 53,397 | 39.4% | 53,400 | 39.4% | 0.99 |
| Yes | 447,014 | 46.2% | 364,476 | 43.8% | 82,538 | 60.7% |  | 164,235 | 60.6% | 82,119 | 60.6% | 82,116 | 60.6% |  |
| Continuous Outcomes |  |  |  |  |  |  |  |  |  |  |  |  |  |  |
|  | mean | SD | mean | SD | mean | SD | p-value | mean | SD | mean | SD | mean | SD | p-value |
| CCI | 0.65 | 1.39 | 0.59 | 1.32 | 0.98 | 1.71 | <0.001 | 0.96 | 1.68 | 0.96 | 1.68 | 0.96 | 1.68 | 0.99 |

CCI: Charlson Comorbidity Index; DEPR: antidepressant; PCP: primary care physician; SD: standard deviation

**Table S11c:** Antidepressant User Cohort (All Regions) by BP Use, Patient Characteristics Pre/Post Match of BP Users/Non-users

|  | All Antidepressant Users by BP: Unmatched |  |  |  |  |  |  | All Antidepressant Users by BP: Matched |  |  |  |  |  |  |
| --- | --- | --- | --- | --- | --- | --- | --- | --- | --- | --- | --- | --- | --- | --- |
|  | All |  | BP Non-user |  | BP User |  | p-value | All |  | BP Non-user |  | BP User |  | p-value |
|  | N | % | N | % | N | % |  | N | % | N | % | N | % |  |
| All Patients | 1,536,048 | 100.0% | 1,390,939 | 90.6% | 145,109 | 9.4% |  | 288,564 | 100.0% | 144,282 | 50.0% | 144,282 | 50.0% |  |
| Age |  |  |  |  |  |  |  |  |  |  |  |  |  |  |
| ≤20 | 89,571 | 5.8% | 89,415 | 6.4% | 156 | 0.1% | <0.001 | 313 | 0.1% | 157 | 0.1% | 156 | 0.1% | 1.00 |
| 21-40 | 315,593 | 20.5% | 314,429 | 22.6% | 1,164 | 0.8% |  | 2,326 | 0.8% | 1,162 | 0.8% | 1,164 | 0.8% |  |
| 41-50 | 233,345 | 15.2% | 229,878 | 16.5% | 3,467 | 2.4% |  | 6,933 | 2.4% | 3,467 | 2.4% | 3,466 | 2.4% |  |
| 51-60 | 333,655 | 21.7% | 310,316 | 22.3% | 23,339 | 16.1% |  | 46,674 | 16.2% | 23,339 | 16.2% | 23,335 | 16.2% |  |
| 61-70 | 296,163 | 19.3% | 244,247 | 17.6% | 51,916 | 35.8% |  | 103,798 | 36.0% | 51,905 | 36.0% | 51,893 | 36.0% |  |
| 71-80 | 169,299 | 11.0% | 126,089 | 9.1% | 43,210 | 29.8% |  | 85,292 | 29.6% | 42,643 | 29.6% | 42,649 | 29.6% |  |
| ≥81 | 98,422 | 6.4% | 76,565 | 5.5% | 21,857 | 15.1% |  | 43,228 | 15.0% | 21,609 | 15.0% | 21,619 | 15.0% |  |
| Gender |  |  |  |  |  |  |  |  |  |  |  |  |  |  |
| Female | 1,109,599 | 72.2% | 976,214 | 70.2% | 133,385 | 91.9% | <0.001 | 265,123 | 91.9% | 132,553 | 91.9% | 132,570 | 91.9% | 0.91 |
| Male | 426,449 | 27.8% | 414,725 | 29.8% | 11,724 | 8.1% |  | 23,441 | 8.1% | 11,729 | 8.1% | 11,712 | 8.1% |  |
| Region |  |  |  |  |  |  |  |  |  |  |  |  |  |  |
| Midwest | 335,508 | 21.8% | 309,597 | 22.3% | 25,911 | 17.9% | <0.001 | 51,754 | 17.9% | 25,877 | 17.9% | 25,877 | 17.9% | 1.00 |
| Northeast | 383,023 | 24.9% | 347,944 | 25.0% | 35,079 | 24.2% |  | 70,010 | 24.3% | 35,005 | 24.3% | 35,005 | 24.3% |  |
| South | 596,029 | 38.8% | 540,382 | 38.9% | 55,647 | 38.3% |  | 110,518 | 38.3% | 55,259 | 38.3% | 55,259 | 38.3% |  |
| West | 221,488 | 14.4% | 193,016 | 13.9% | 28,472 | 19.6% |  | 56,282 | 19.5% | 28,141 | 19.5% | 28,141 | 19.5% |  |
| Insurance |  |  |  |  |  |  |  |  |  |  |  |  |  |  |
| Commercial | 707,676 | 46.1% | 664,625 | 47.8% | 43,051 | 29.7% | <0.001 | 86,053 | 29.8% | 43,023 | 29.8% | 43,030 | 29.8% | 1.00 |
| Dual | 54,840 | 3.6% | 43,171 | 3.1% | 11,669 | 8.0% |  | 22,384 | 7.8% | 11,193 | 7.8% | 11,191 | 7.8% |  |
| Medicaid | 486,451 | 31.7% | 457,656 | 32.9% | 28,795 | 19.8% |  | 56,959 | 19.7% | 28,479 | 19.7% | 28,480 | 19.7% |  |
| Medicare | 287,081 | 18.7% | 225,487 | 16.2% | 61,594 | 42.4% |  | 123,168 | 42.7% | 61,587 | 42.7% | 61,581 | 42.7% |  |
| PCP Visit 2019 |  |  |  |  |  |  |  |  |  |  |  |  |  |  |
| No | 605,264 | 39.4% | 553,886 | 39.8% | 51,378 | 35.4% | <0.001 | 102,148 | 35.4% | 51,064 | 35.4% | 51,084 | 35.4% | 0.94 |
| Yes | 930,784 | 60.6% | 837,053 | 60.2% | 93,731 | 64.6% |  | 186,416 | 64.6% | 93,218 | 64.6% | 93,198 | 64.6% |  |
| Continuous Outcomes |  |  |  |  |  |  |  |  |  |  |  |  |  |  |
|  | mean | SD | mean | SD | mean | SD | p-value | mean | SD | mean | SD | mean | SD | p-value |
| CCI | 0.87 | 1.60 | 0.84 | 1.58 | 1.09 | 1.81 | <0.001 | 1.09 | 1.79 | 1.08 | 1.78 | 1.09 | 1.79 | 0.56 |

BP: bisphosphonate; CCI: Charlson Comorbidity Index; PCP: primary care physician; SD: standard deviation

**Table S11d:**
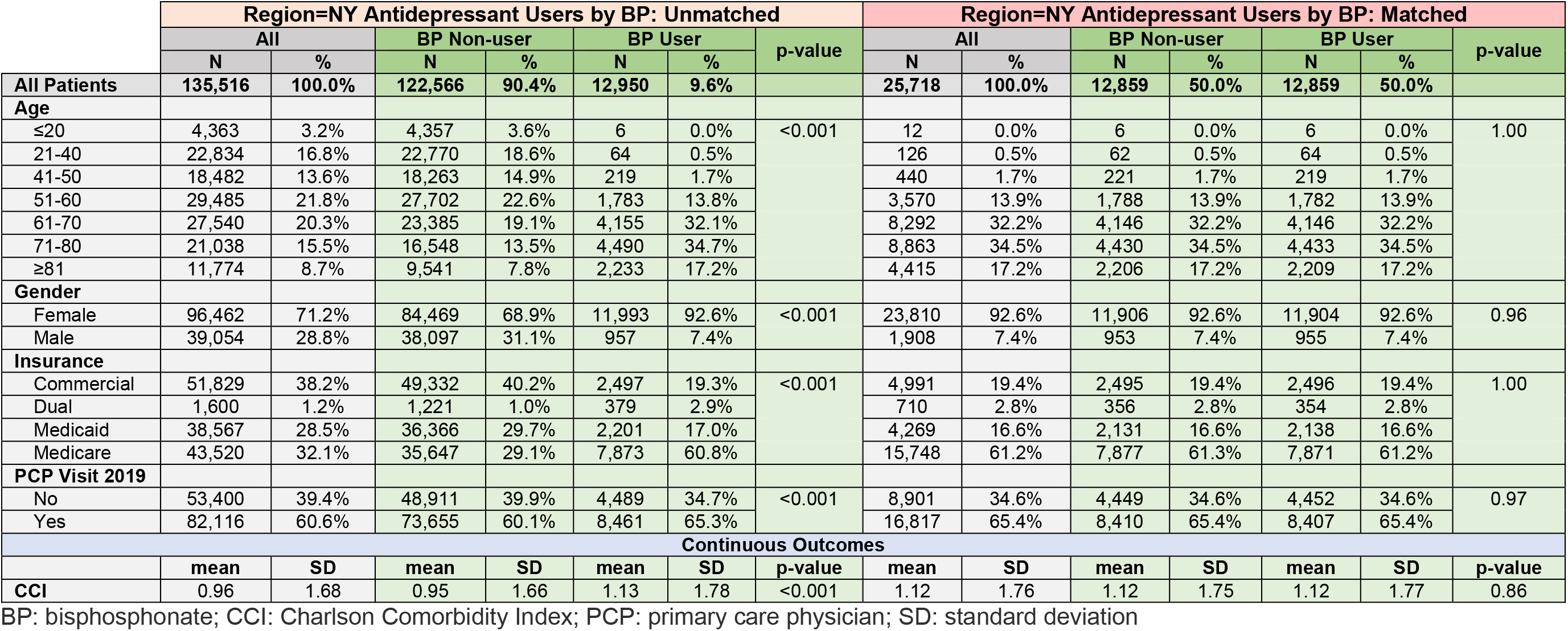
Antidepressant User Cohort (Region=New York State) by BP Use, Patient Characteristics Pre/Post Match of BP Users/Non-users

**Table S11e:** Antidepressant Non-user Cohort (All Regions) by BP Use, Patient Characteristics Pre/Post Match of BP Users/Non-users

|  | All Antidepressant Non-users by BP: Unmatched |  |  |  |  |  |  | All Antidepressant Non-users by BP: Matched |  |  |  |  |  |  |
| --- | --- | --- | --- | --- | --- | --- | --- | --- | --- | --- | --- | --- | --- | --- |
|  | All |  | BP Non-user |  | BP User |  | p-value | All |  | BP Non-user |  | BP User |  | p-value |
|  | N | % | N | % | N | % |  | N | % | N | % | N | % |  |
| All Patients | 1,536,048 | 100.0% | 1,422,938 | 92.6% | 113,110 | 7.4% |  | 224,804 | 100.0% | 112,402 | 50.0% | 112,402 | 50.0% |  |
| Age |  |  |  |  |  |  | <0.001 |  |  |  |  |  |  | 1.00 |
| ≤20 | 89,565 | 5.8% | 89,486 | 6.3% | 79 | 0.1% |  | 155 | 0.1% | 76 | 0.1% | 79 | 0.1% |  |
| 21-40 | 315,593 | 20.5% | 314,815 | 22.1% | 778 | 0.7% |  | 1,562 | 0.7% | 784 | 0.7% | 778 | 0.7% |  |
| 41-50 | 233,336 | 15.2% | 230,961 | 16.2% | 2,375 | 2.1% |  | 4,746 | 2.1% | 2,371 | 2.1% | 2,375 | 2.1% |  |
| 51-60 | 333,650 | 21.7% | 314,109 | 22.1% | 19,541 | 17.3% |  | 39,072 | 17.4% | 19,536 | 17.4% | 19,536 | 17.4% |  |
| 61-70 | 296,182 | 19.3% | 254,286 | 17.9% | 41,896 | 37.0% |  | 83,664 | 37.2% | 41,834 | 37.2% | 41,830 | 37.2% |  |
| 71-80 | 169,295 | 11.0% | 136,746 | 9.6% | 32,549 | 28.8% |  | 64,163 | 28.5% | 32,073 | 28.5% | 32,090 | 28.5% |  |
| ≥81 | 98,427 | 6.4% | 82,535 | 5.8% | 15,892 | 14.1% |  | 31,442 | 14.0% | 15,728 | 14.0% | 15,714 | 14.0% |  |
| Gender |  |  |  |  |  |  |  |  |  |  |  |  |  |  |
| Female | 1,109,580 | 72.2% | 1,004,112 | 70.6% | 105,468 | 93.2% | <0.001 | 209,510 | 93.2% | 104,743 | 93.2% | 104,767 | 93.2% | 0.84 |
| Male | 426,468 | 27.8% | 418,826 | 29.4% | 7,642 | 6.8% |  | 15,294 | 6.8% | 7,659 | 6.8% | 7,635 | 6.8% |  |
| Region |  |  |  |  |  |  |  |  |  |  |  |  |  |  |
| Midwest | 335,508 | 21.8% | 315,179 | 22.1% | 20,329 | 18.0% | <0.001 | 40,548 | 18.0% | 20,274 | 18.0% | 20,274 | 18.0% | 1.00 |
| Northeast | 383,023 | 24.9% | 356,184 | 25.0% | 26,839 | 23.7% |  | 53,590 | 23.8% | 26,795 | 23.8% | 26,795 | 23.8% |  |
| South | 596,029 | 38.8% | 552,754 | 38.8% | 43,275 | 38.3% |  | 85,440 | 38.0% | 42,720 | 38.0% | 42,720 | 38.0% |  |
| West | 221,488 | 14.4% | 198,821 | 14.0% | 22,667 | 20.0% |  | 45,226 | 20.1% | 22,613 | 20.1% | 22,613 | 20.1% |  |
| Insurance |  |  |  |  |  |  |  |  |  |  |  |  |  |  |
| Commercial | 707,675 | 46.1% | 672,990 | 47.3% | 34,685 | 30.7% | <0.001 | 69,354 | 30.9% | 34,675 | 30.8% | 34,679 | 30.9% | 1.00 |
| Dual | 54,836 | 3.6% | 44,281 | 3.1% | 10,555 | 9.3% |  | 19,871 | 8.8% | 9,927 | 8.8% | 9,944 | 8.8% |  |
| Medicaid | 486,446 | 31.7% | 463,857 | 32.6% | 22,589 | 20.0% |  | 45,057 | 20.0% | 22,537 | 20.1% | 22,520 | 20.0% |  |
| Medicare | 287,091 | 18.7% | 241,810 | 17.0% | 45,281 | 40.0% |  | 90,522 | 40.3% | 45,263 | 40.3% | 45,259 | 40.3% |  |
| PCP Visit 2019 |  |  |  |  |  |  |  |  |  |  |  |  |  |  |
| No | 605,256 | 39.4% | 572,701 | 40.2% | 32,555 | 28.8% | <0.001 | 64,959 | 28.9% | 32,483 | 28.9% | 32,476 | 28.9% | 0.97 |
| Yes | 930,792 | 60.6% | 850,237 | 59.8% | 80,555 | 71.2% |  | 159,845 | 71.1% | 79,919 | 71.1% | 79,926 | 71.1% |  |
| Continuous Outcomes |  |  |  |  |  |  |  |  |  |  |  |  |  |  |
|  | mean | SD | mean | SD | mean | SD | p-value | mean | SD | mean | SD | mean | SD | p-value |
| CCI | 0.87 | 1.60 | 0.85 | 1.58 | 1.06 | 1.84 | <0.001 | 1.06 | 1.82 | 1.05 | 1.81 | 1.06 | 1.83 | 0.57 |

BP: bisphosphonate; CCI: Charlson Comorbidity Index; PCP: primary care physician; SD: standard deviation

**Table S11f:**
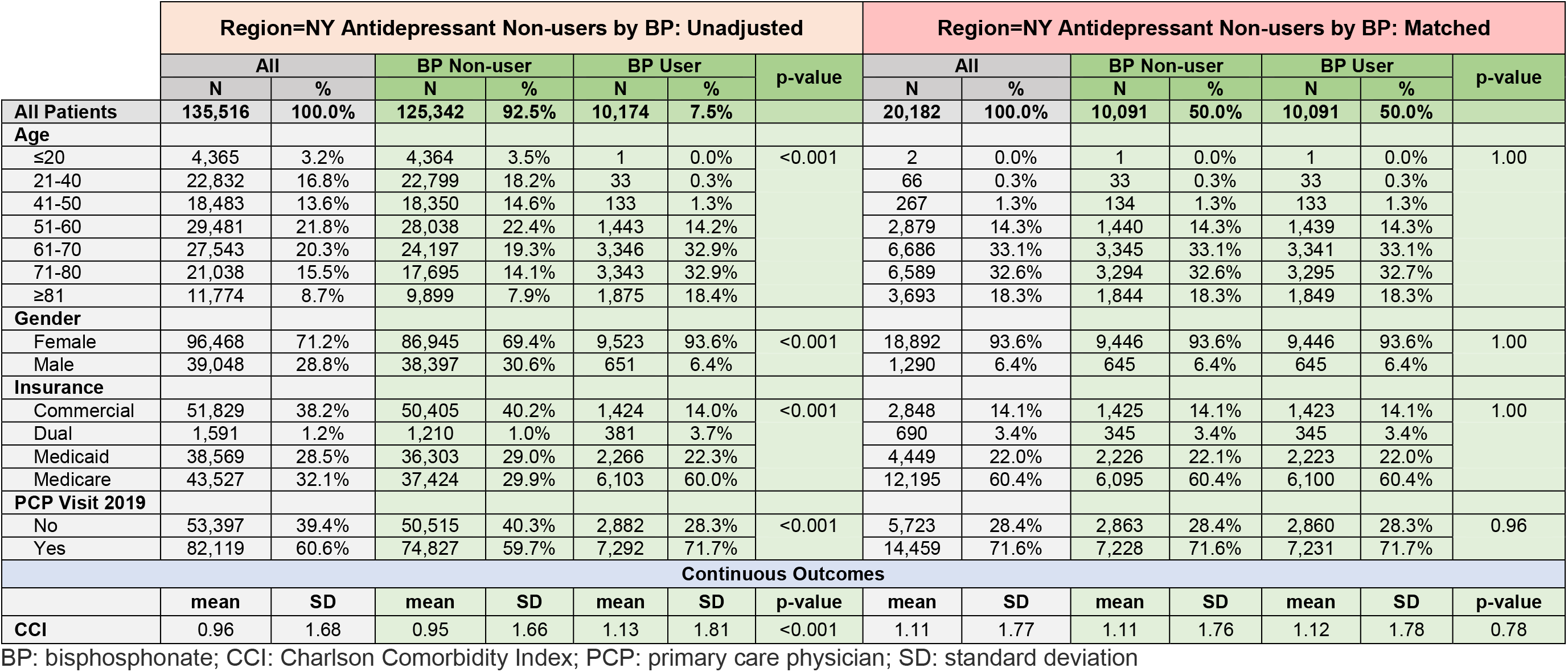
Antidepressant Non-user Cohort (Region=New York State) by BP Use, Patient Characteristics Pre/Post Match of BP Users/Non-users

**Table S12a:**
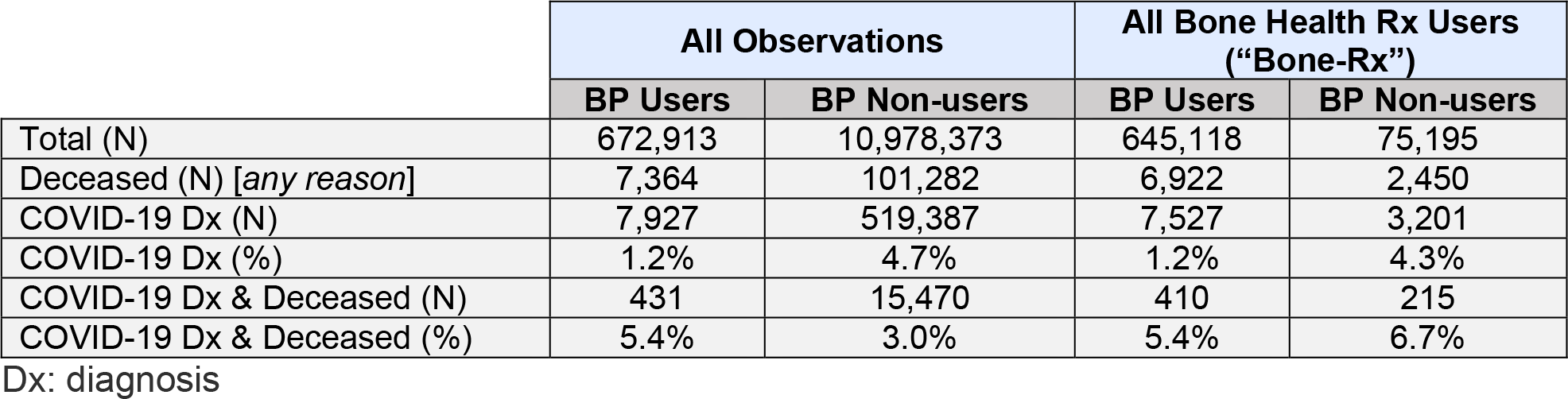
Patient Count Distribution Inclusive of Deceased Enrollees

|  | All Observations |  | All Bone Health Rx Users<br>("Bone-Rx") |  |
| --- | --- | --- | --- | --- |
|  | BP Users | BP Non-users | BP Users | BP Non-users |
| Total (N) | 672,913 | 10,978,373 | 645,118 | 75,195 |
| Deceased (N) [any reason] | 7,364 | 101,282 | 6,922 | 2,450 |
| COVID-19 Dx (N) | 7,927 | 519,387 | 7,527 | 3,201 |
| COVID-19 Dx (%) | 1.2% | 4.7% | 1.2% | 4.3% |
| COVID-19 Dx & Deceased (N) | 431 | 15,470 | 410 | 215 |
| COVID-19 Dx & Deceased (%) | 5.4% | 3.0% | 5.4% | 6.7% |

Dx: diagnosis

**Table S12b:**
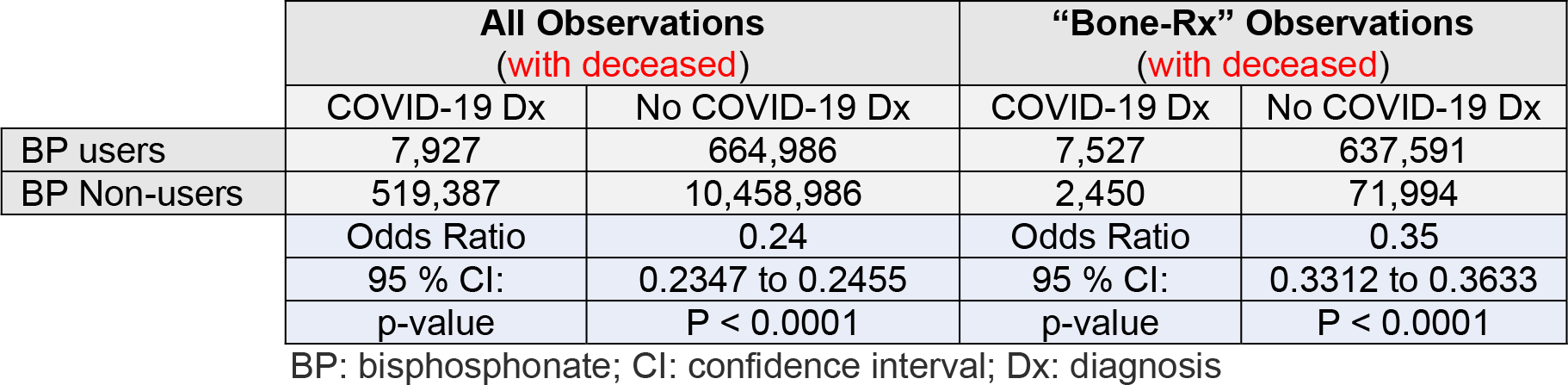
Unadjusted Chi-Square Comparison Inclusive of Deceased Patients

**Table S12c:**
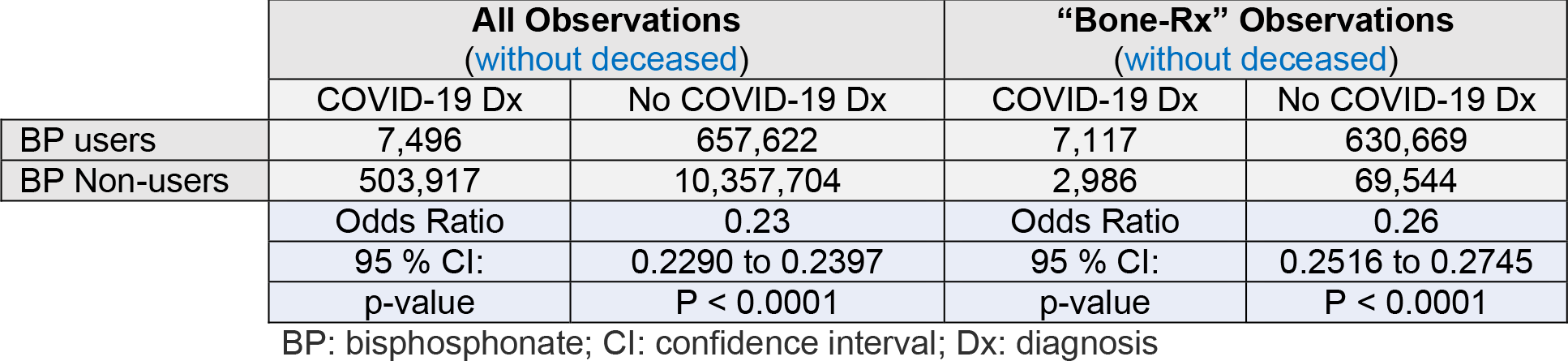
Unadjusted Chi-Square Comparison with Deceased Patients Removed

**Table S12d:**
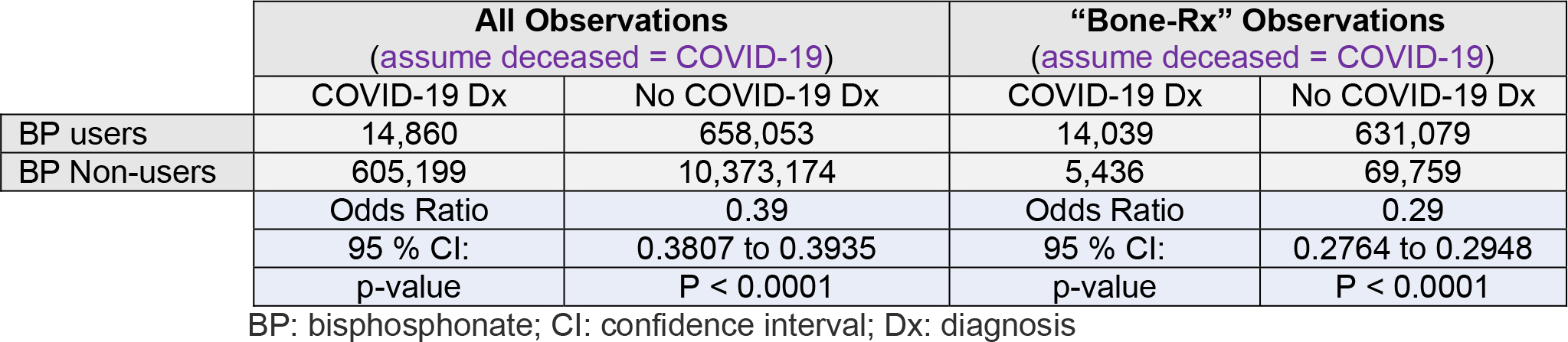
Unadjusted Chi-Square Comparison Assuming all Deceased Patients had COVID-19

**Table S12e:**
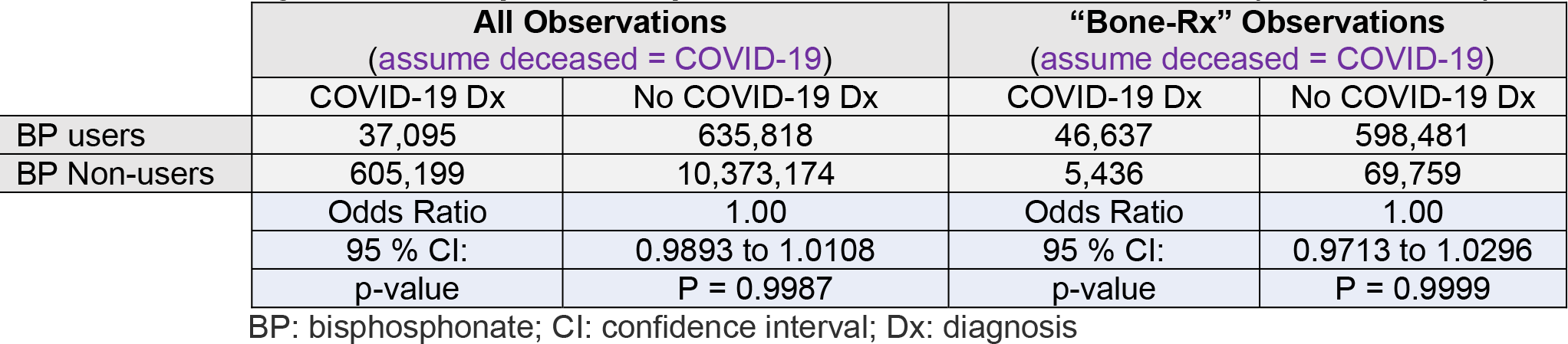
Unadjusted Chi-Square Comparison to Yield Odds Ratio = 1.00 (no difference)

